# Integrative Genetic Analyses of Lipid Metabolism and Multiple Sclerosis Severity Using Metabolome-Wide and Cis-Mendelian Randomization

**DOI:** 10.64898/2026.05.27.26354239

**Authors:** Rose Noroozi, Cesar Higgins Tejera, Mingjing Chen, Farren B.S. Briggs, Pavan Bhargava, Kathryn C. Fitzgerald

## Abstract

The course of multiple sclerosis (MS) is highly heterogeneous, yet the biological mechanisms underlying this variability remain incompletely understood. Although metabolic alterations have increasingly been associated with disease progression, existing observational evidence is limited by confounding, reverse causation, and an inability to establish causal mechanisms. To bridge this gap, we used a metabolome-wide Mendelian Randomization (MR) framework, including thorough sensitivity analyses, to identify metabolites genetically linked to MS severity that can causally affect it. Bidirectional MR analyses revealed a subset of amino acid and lipid pathways with strong, consistent effects across different MR approaches, confirmed by tests for heterogeneity, horizontal pleiotropy, and LD confounding. For metabolites prioritized by metabolome-wide MR with evidence of causal effects, we conducted genetic colocalization at loci encompassing proximal enzyme-encoding genes, leveraging the corresponding instrumental variants to assess shared underlying genetic signals. This process revealed shared genetic signals between metabolite levels and MS severity, mapped to the *FADS1/2* and *CYP4F2* loci. A subsequent pathway-resolved set of cis-MR analyses across *FADS1/2*-derived polyunsaturated fatty acid (PUFA) metabolites, using a functional variant that proxies reduced Δ5-desaturase activity, showed consistent effects indicating that *FADS1* perturbation is associated with MS severity. Collectively, these results highlight *FADS1* as a key driver of PUFA-related causal effects on MS severity in both systemic (circulating metabolites) and brain cell-specific contexts. Additional supportive cis-MR evidence implicates the disruption of *CYP4F2* as another PUFA-metabolizing enzyme.

**Highlight:**

- Metabolome-wide MR identifies lipid and amino acid metabolites causally linked to MS severity
- Genetic colocalization and cis-MR prioritize PUFA metabolism as the key causal pathway
- Brain cell-specific cis-MR links reduced *FADS1* activity to increased MS severity
- *CYP4F2* implicates additional PUFA metabolic regulation in MS severity

**Graphical abstract:** 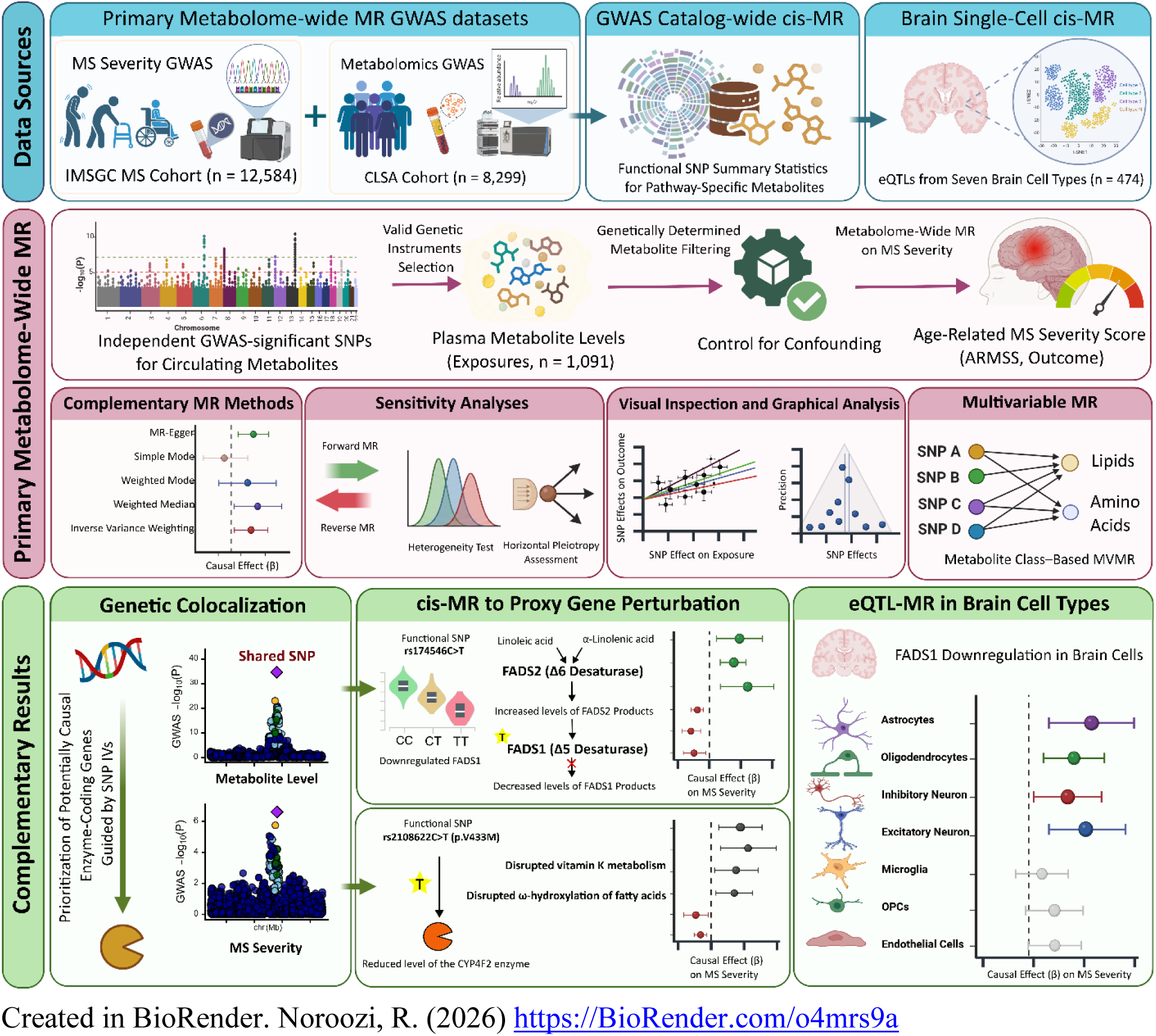

## Introduction

The determinants underlying the heterogeneous severity of multiple sclerosis (MS) limit accurate prediction of disease course and slow the development of effective interventions(1–7). This challenge has led to efforts to develop biomarkers that capture underlying biological differences and provide insights into disease progression. In this regard, metabolomics has emerged as a promising avenue for elucidating potential biological pathways. Metabolites are small molecules that reflect the integrated output of genetic, environmental, and immunologic processes and play essential roles in cellular energy, lipid metabolism, and inflammation. As such, they may provide a functional readout of pathways that may contribute to MS pathogenesis and progression. For example, in our prior work, we identified widespread alterations in lipid and aromatic amino acid pathways that were associated with downstream clinical outcomes in people with MS. However, these observational associations are vulnerable to confounding bias and reverse causation (8–11).

This limitation can be addressed using genomic-informed analytical approaches, including Mendelian randomization (MR). These methods allow systematic assessment of the impact of metabolite levels on MS severity. MR is a type of instrumental variable analysis that leverages genetic instruments for a given exposure (metabolite levels in our context) to assess its relationship to an outcome (here, MS severity). This approach minimizes confounding because genetic variants are randomly inherited and fixed at conception, making them largely independent of the environmental and clinical confounders that commonly affect observational studies. In addition, MR reduces the risk of reverse causation, thereby enabling a more robust assessment of whether metabolic alterations may causally contribute to MS severity(12).

MR analyses, particularly when combined with downstream approaches such as fine-mapping or genetic colocalization, enable identification of genetic determinants of MS severity (e.g., disability progression and disease worsening) and the biological pathways through which these causal effects operate. Together, these methods provide mechanistic insight that may ultimately inform the (1) development of new strategies for prevention of disease progression, or (2) identification of biomarkers more proximally linked to such underlying contributing processes.

In this study, we leveraged summary statistics from the largest available genome-wide association studies (GWAS) of plasma metabolites and age-adjusted measures of MS severity, available at the time of analysis, to systematically test the potential effects of circulating metabolites on disease progression. To address the substantial correlation structure among metabolites, we applied pathway-informed multivariable MR models to estimate the independent effects of individual metabolites on MS severity. To further prioritize candidate biological mechanisms, we integrated genetic colocalization analyses to identify loci where shared causal variants may jointly influence both metabolite levels and disease progression. At these loci, we performed cis-MR analyses using functional genetic variants as proxies for gene perturbation, enabling us to evaluate the potential causal effects of implicated genes on MS severity across peripheral tissues and cell type-specific contexts in the brain.

## Methods

### Data sources and MR design

To identify relevant metabolites and shared regulatory mechanisms influencing MS severity, we conducted a metabolome-wide two-sample MR study that integrates genetic colocalization and cis-MR analyses. **Figure 1** provides an overview of our approach; all analyses were performed in accordance with the STROBE-MR reporting guidelines(13). We used summary statistics from the most comprehensive available GWAS for both circulating metabolite levels (with respect to sample size and breadth of metabolites [exposures]) and MS severity (outcome). For each metabolite, we selected independent, significant single-nucleotide polymorphisms (SNPs) that satisfy the following three MR assumptions: (1) instrumental variables (IVs) with a strong association with metabolites; (2) IVs independent of any confounders; and (3) IVs linked to MS severity only via metabolite changes.

**Figure 1.**
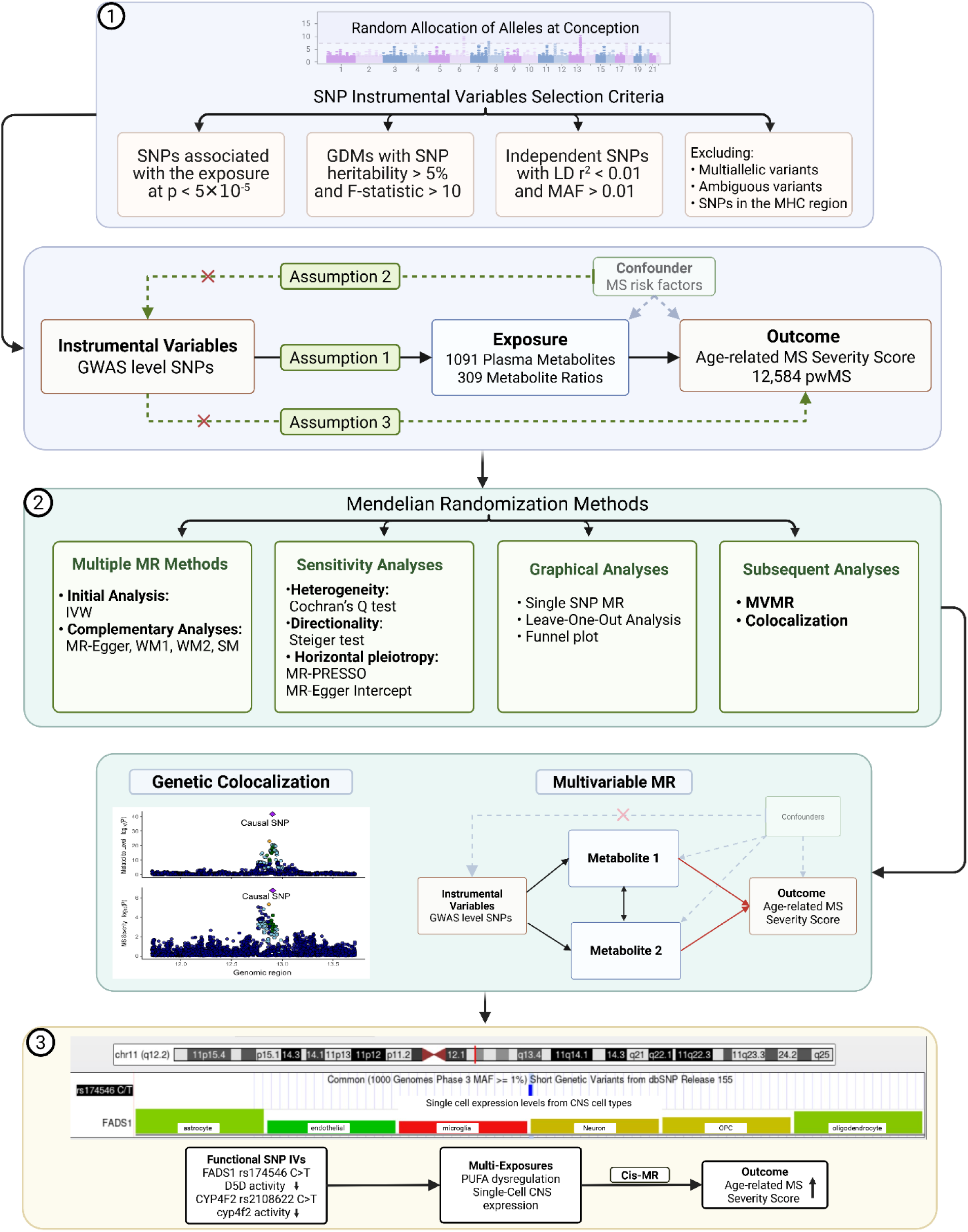
Flowchart of different phases of the study and the MR analysis pipeline GDM: Genetically Determined Metabolites, MHC: Major Histocompatibility Complex region, IVW: Inverse Variance Weighted, WM1: Weighted Median, WM2: Weighted Mode, and SM: Simple Mode methods, MVMR: Multivariable MR.

### Metabolite Instruments

Genome-wide association data for plasma metabolite levels were obtained from a comprehensive non-targeted metabolomics GWAS (mGWAS) that assayed 1,091 metabolites and 309 metabolite ratios, as previously described. Briefly, 8,299 participants aged 45 to 85 years from the Canadian Longitudinal Study on Aging (CLSA). Participants were genotyped using the Affymetrix Axiom genotyping platform, and imputation was performed using the Trans-Omics for Precision Medicine program(14). GWAS preprocessing and quality control, as outlined previously, retained samples of European ancestry with ∼15.4 million SNPs, a minor allele frequency (MAF) > 0.1%, an imputation quality score > 0.3, and a call rate > 0.99. Metabolomic profiles were obtained from the non-fasting blood samples using ultrahigh-performance liquid chromatography tandem mass spectrometry (UPLC–MS/MS) (Metabolon, Inc.). Details of the quality control parameters for metabolomics and genetic data used to identify mQTLs have been previously described (15, 16).

### Definition and Genetic Assessment of MS Severity

Genetic liability to multiple sclerosis (MS) severity was evaluated using summary statistics from the discovery genome-wide association study (GWAS) conducted by the International Multiple Sclerosis Genetics Consortium, comprising approximately 7.8 million autosomal SNPs with a minor allele frequency (MAF) ≥ 0.01 in 12,584 people with MS (pwMS) of European ancestry(17). The study comprised discovery and replication cohorts enriched for older individuals with longer disease duration to improve the stability of disability-related phenotypes. In the discovery cohort, 71.7% of participants were female, with a mean age of 51.7 years and a disease duration of 18.2 ± 10.6 years. Neurological disability was measured using the age-related MS severity score (ARMSS; mean 4.23 ± 2.77), which combines an individual’s disability level, as quantified by the Expanded Disability Status Scale (EDSS), and their age (18).

### Genetic instrumental variables selection

Across each metabolite considered in the original mGWAS, we calculated the variance in plasma levels explained by genetic variants using linkage disequilibrium (LD) score regression and estimated SNP-based heritability (h^2^_SNP_) (19). We retained only metabolites with an estimated h^2^_SNP_>5% for the MR analysis (i.e., genetically determined metabolites [GDMs])(20).

We selected genetic IVs in accordance with MR assumptions. For each metabolite, we selected SNPs with MAF > 0.01 and those that were significantly associated with circulating levels, using a P-value threshold of 1×10^−5^; we applied this relatively relaxed threshold because the original mGWAS sample size was modest. Multiallelic and strand-ambiguous variants were excluded, as were single-nucleotide polymorphisms located within the major histocompatibility complex (MHC) region (GRCh38/hg38: chr6:28,510,020-33,480,577) (21). We selected independent SNPs using LD clumping (r^2^=0.01 within a clump-kb10,000 window) using the 1000 Genomes European reference. We included metabolites with at least three SNP IVs in the final MR analysis. Finally, to fulfill the assumption that IVs are independent of confounders, we explored any reported association of IVs with MS severity risk factors, including BMI, smoking, and education-related CNS reserve in the European population (P < 1 × 10^−8^) using the latest version of the GWAS catalog data (*gwas_catalog_v1.0-associations_e115_r2025-09-15*)(16). Further, we queried SNPs within a ±250 kb window of the selected IVs to confirm the absence of high linkage disequilibrium(r² > 0.8) with variants associated with any of the mentioned MS risk factors using the LDtrait module of the LDlink web-based application(22).

### Mendelian Randomization Analysis

For the selected IVs, effect estimates for both metabolite levels and MS severity were extracted from the exposure and outcome GWAS summary statistics and harmonized to the same effect allele. When a selected IV was absent from the outcome GWAS, LD proxy SNPs were identified using a European LD reference panel (r^2^=0.8). For our primary analysis, the causal effect estimate of circulating metabolites on MS severity was estimated via inverse variance weighting (IVW)(23). Using a previously estimated effective number of 73 independent metabolites using the eigendecomposition(15), significance was defined at P < 0.05/73 (6.8 × 10⁻⁴).

### Sensitivity analysis

Metabolites with suggestive causal effect (IVW P < 0.05) were evaluated using complementary MR methods (MR-Egger, weighted median, simple mode, weighted mode). Those metabolites with consistent MR estimates across methods underwent sensitivity analyses to assess robustness and IV validity. Cochran’s Q test was used to assess potential heterogeneity, and complementary approaches, including MR-Egger intercept and MR-PRESSO, were used to test for horizontal pleiotropy (24). Additionally, we visually inspected MR results using scatter and funnel plots (to characterize symmetry and precision of causal estimates). We conducted single-SNP MR and a leave-one-out (LOO) analysis to assess whether any single SNP disproportionately influenced the overall MR estimate. To assess reverse causality, we applied Steiger filtering to all significant metabolites. This approach assesses bidirectional effects by retaining only IVs that explain more variance in the exposure than in the outcome, excluding variants that violate this assumption (25). In addition, we conducted reverse MR by exchanging the exposure and outcome variables, in which we designated ARMSS as the exposure and metabolite levels as the outcomes; here, we used SNP IVs associated with the ARMSS (P < 1 × 10–5) to assess if MS severity influences metabolite levels using the IVW method.

To prioritize overall metabolite pathways associated with MS severity and to try to disentangle correlated metabolic effects, we performed multivariable MR (MVMR) using inverse variance weighting to estimate the direct effects of specific lipid and amino acid (AA) subsets, as our prior study suggested these may be particularly relevant for MS(26). Pathway-based MVMR was performed by including all significant AAs and the lipids involved in the fatty acid (FA) metabolism sub-pathway in separate models. We excluded metabolites with evidence of heterogeneity, palindromic IVs, and those associated with MS severity risk factors at the GWAS level. All analyses were done using R (version 4.0.1). The R packages included: TwoSampleMR(27), MendelianRandomization(26), MRPRESSO [29], ggplot2, pheatmap, and forestplot.

### Genetic Colocalization Analysis

To evaluate the potential for false-positive MR estimates due to LD (i.e., distinct variants for the exposure and outcome are correlated), we performed genetic colocalization analyses on MR-significant GDMs. This approach identified whether specific loci in which GDM levels and MS severity are likely driven by a shared variant rather than by correlated signals, thereby serving as a sensitivity analysis for our overall analysis. Here, we filtered metabolome-wide MR results, identified the genomic coordinates of the IVs associated with significant GDMs, and mapped them to nearby enzyme-coding regions. For any of these genomic regions, we then applied the Bayesian colocalization method (coloc), implemented in the coloc R package (v5.2.3)(25). This approach specifies prior probabilities for association under the assumption of a single causal variant within a genomic region of interest for two traits. It then estimates posterior probabilities (PPs) for five competing hypotheses: no association with either trait (PP.H0); association with the metabolite level (trait 1) only (PP.H1); association with MS severity (trait 2) only (PP.H2); two distinct causal variants, one for each trait (PP.H3; no colocalization); or a shared causal variant influencing both traits (PP.H4; colocalization). We used default priors in our analysis (p1 = 1 × 10⁻⁴, p2 = 1 × 10⁻⁴, p12 = 1 × 10⁻⁵). We then applied single-SNP MR and leave-one-out (LOO) analyses to prioritize loci driving metabolite-MS severity associations by repeating colocalization analyses with a more stringent prior (p_12_ = 1 × 10⁻⁴).

### Cis-MR Analysis of Colocalized Loci Influencing Multiple Sclerosis Severity

To further evaluate the functional relevance of colocalized loci to MS severity, we applied cis-MR analysis to estimate the effects of genetically proxied perturbations in the implicated cis-genes. Consistent with cis-MR principles, we selected exposure biomarkers for gene perturbation, including circulating levels of downstream metabolite products of the cis-gene (reflecting perturbed enzyme activity) and tissue-specific single-cell expression of the cis-gene in the brain. For each colocalized locus, a functional cis-acting variant was selected as a single IV within the gene body or regulatory regions to minimize horizontal pleiotropy. These variants were chosen based on reported functional evaluations of their effects on gene expression or on the activity of the encoded enzyme.

Primary multi-exposure cis-MR analyses in peripheral tissues were implemented using functional SNP IVs associated with circulating metabolite levels as exposure biomarkers. Metabolites were selected a priori based on biological knowledge as downstream products of the colocalized gene, thereby capturing perturbed enzyme activity. Genetic associations of the selected IVs and metabolite levels in European populations were obtained from the GWAS Catalog (accessed November 17, 2025). Complementary tissue-specific cis-MR leveraged brain cell-type-specific gene expression as an exposure biomarker, with an expression quantitative trait locus (eQTL) IV. For both peripheral (plasma metabolite) and brain expression cis-MR analyses, ARMSS was used as the outcome(28).

## Results

### Selected Metabolites and IVs

Using summary statistics from the CLSA metabolite GWAS, comprising 1,091 metabolites and 309 metabolite ratios measured in individuals of European ancestry (mean age 62.4 ± 9.9 years; 50.9% female), we performed MR analyses on GDMs (h^2^_SNP_ > 0.05), along with all available metabolite ratios. For each GDM, the number of selected IVs ranged from 12 to 55 SNPs, demonstrating adequate instrumental strength, as indicated by F-statistics greater than 10 (**Supplementary Data S1**).

### Causal Effects of Metabolite Levels on MS Severity

In primary analyses incorporating the full set of selected IVs for each GDM, we identified 45 metabolites with nominally significant estimates (P < 0.05), of which 36 were annotated metabolites or metabolite ratios, and nine corresponded to uncharacterized compounds (**Figure 2**).

**Figure 2.**
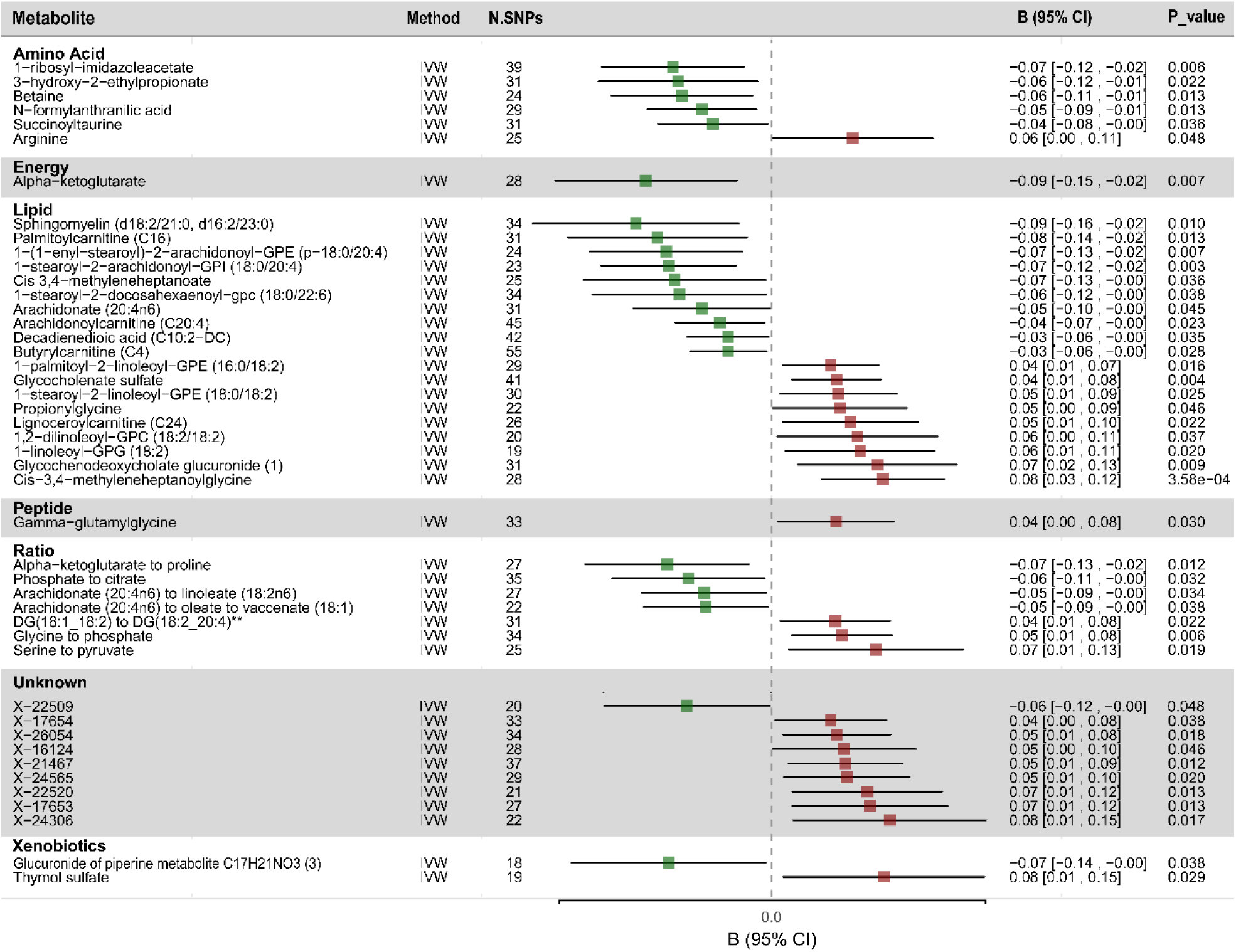
Forest plots presenting the results of MR analysis and effect estimates of plasma metabolites on MS severity. **Oleoyl-linoleoyl-glycerol (18:1 to 18:2) [2] to linoleoyl-arachidonoyl-glycerol (18:2 to 20:4) [2] ratio. IVW: Inverse variance weighted; SNP: single nucleotide polymorphism. CI: confidence interval

We next applied complementary MR methods to evaluate the robustness of the IVW findings under alternative assumptions regarding pleiotropy. Among the nominally significant metabolites identified in the primary IVW analysis, 40 demonstrated concordant effect directions using the MR-Egger method **(Figure 3).** Cochran’s Q statistic indicated no evidence of heterogeneity for most metabolites, except for glycochenodeoxycholate glucuronide and 1-stearoyl-2-docosahexaenoyl-GPC (18:0/22:6), which were excluded from the MVMR analysis. Notably, for these two metabolites, LOO analyses showed consistent directionality and distribution of causal effect estimates. A summary of sensitivity analyses and visual assessments is provided in the **Supplementary File.**

**Figure 3.**
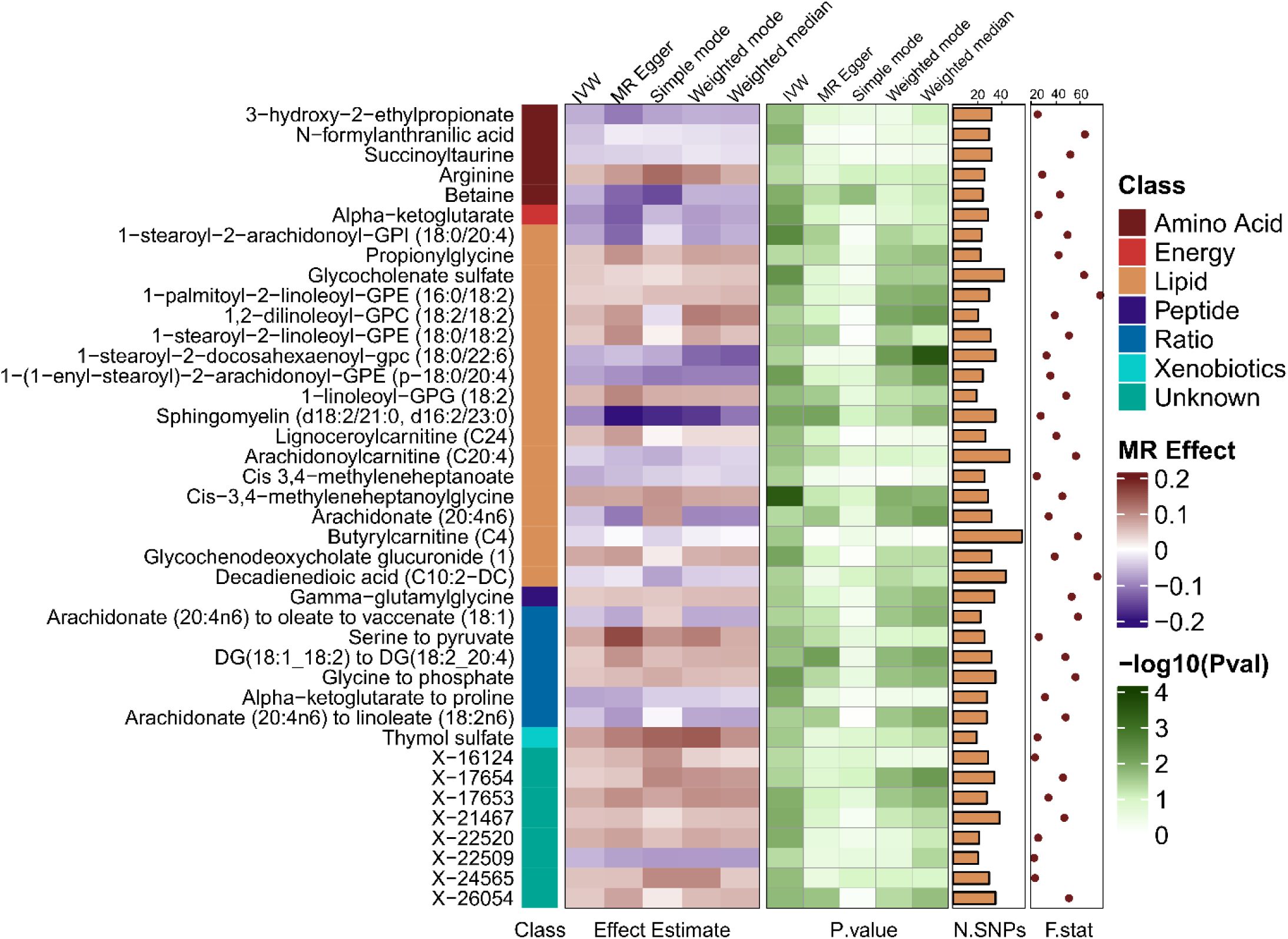
The causal effects of metabolites on MS severity across MR methods.

To further evaluate potential violations of MR assumptions, we examined whether instrumental variants were associated with established MS severity risk factors, including BMI, smoking, and education. Excluding variants with genome-wide significance did not materially alter the effect estimates. MR-PRESSO identified a single outlier for glycochenodeoxycholate glucuronide; removal of this variant did not change the significance of the association **(Supplementary Data S2)**. Steiger filtering confirmed the expected causal direction, with the selected instrumental variables explaining more variance in the exposure than in the outcome, supporting their suitability as valid instruments**(Supplementary Data S3)**(25). Finally, reverse IVW-MR analyses provided no evidence of reverse causation for the identified associations, except for decadienedioic acid (C10:2-DC)**(Supplementary Data S4).**

### Multivariable MR analysis

To account for correlations among instruments for related metabolites, we performed MVMR analyses that included IVW-significant GDMs within the same metabolite class. This approach enabled estimation of the direct effect of each metabolite on MS severity independent of related metabolites. Using the IVW framework for MVMR, we identified significant direct effects for three AA and two fatty acid-related lipids (**Figure 4**).

**Figure 4.**
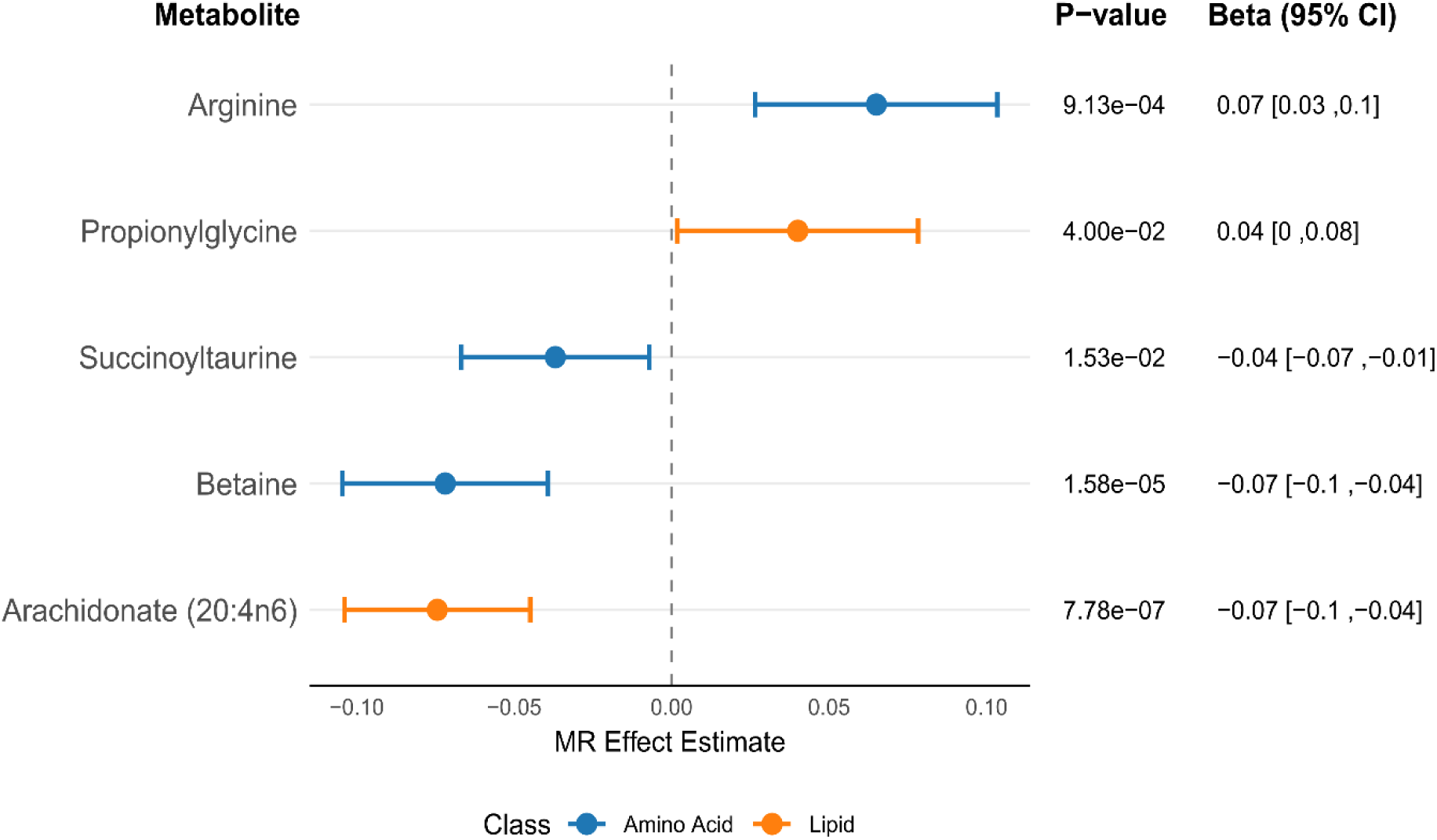
Significant results from pathway-specific Multivariable MR analysis.

### Genetic Colocalization and Cis-MR analysis

To further assess the robustness of our metabolome-wide MR, we performed genetic colocalization to determine whether the significant MR estimates could be explained by shared risk variants for both GDM and MS severity, rather than by LD-related confounding with nearby variants. To reduce the potential for horizontal pleiotropy, we limited the analysis to instrumental variables mapped to enzyme-coding gene loci implicated by genome-wide significant associations with MR-identified metabolites. Posterior probabilities were estimated for each model, with particular emphasis on PP.H4 (shared causal variant) relative to PP.H3 (distinct causal variants).

Our analyses provided evidence of genetic colocalization (PP.H4 > 0.70) within the well-established pleiotropic locus on chromosome 11, which encompasses multiple overlapping genes. Specifically, we observed shared colocalization signals for *FADS1*, *FADS2*, *MYRF*, and *TMEM258* across three fatty acid-related metabolite ratios: arachidonate (20:4n6)-to-linoleate (18:2n6), arachidonate (20:4n6)-to-oleate/vaccenate (18:1), and oleoyl-linoleoyl-glycerol (18:1/18:2)-to-linoleoyl-arachidonoyl-glycerol (18:2/20:4) (Figure 5.A). Colocalization analyses identified rs174564 as the putative shared causal variant underlying the associations at *FADS1*/2, whereas rs174537 and rs102274 were implicated for *MYRF* and *TMEM258*, respectively. However, both variants are in near-complete linkage disequilibrium with rs174564 (D′ = 1.0, R² = 0.99) (Figure 5.B). Considering that the colocalized metabolite ratios are associated with the FADS1/FADS2-mediated polyunsaturated fatty acid metabolic pathway, we prioritized the biological plausibility of a causal effect of *FADS1/2* genes, which encode rate-limiting enzymes for PUFA metabolism, and focused subsequent cis-MR analyses on this pathway.

**Figure 5.**
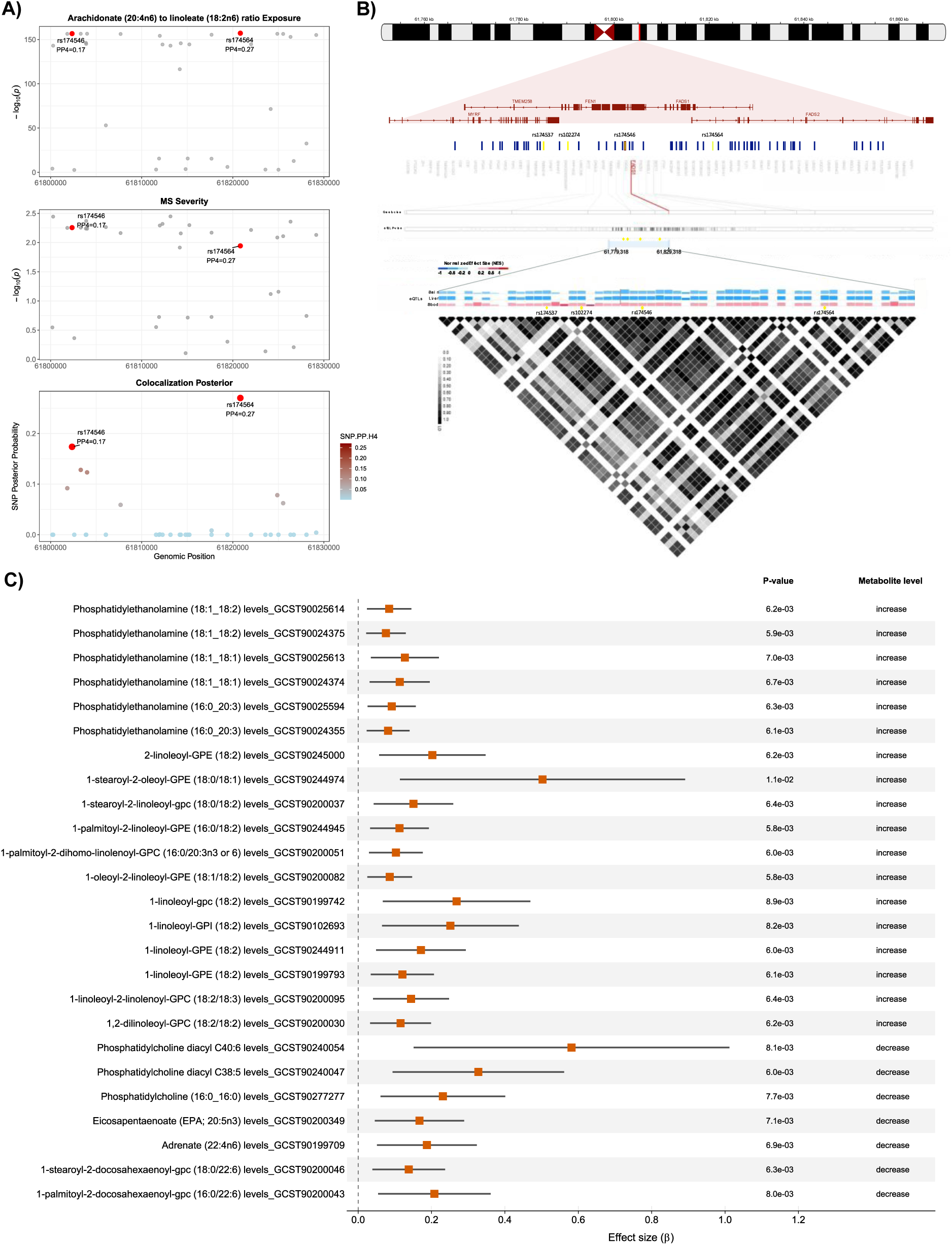
cis-MR of FADS1 Downregulation–Related Metabolite Changes on MS Severity **A)** Colocalization at the *FADS1* locus; **B)** LD Structure and Haplotype Architecture at the FADS Locus. The genomic coordinates on chromosome 11 (hg38) are shown at the top right, and the four linked causal SNPs identified for genes in the region are labeled. The haplotype block in the region is shown for the eQTLs across brain, liver, and blood tissue, illustrated using the GTEx Locus Browser; **C)** Forest plot showing the causal estimate for GWAS-wide Cis-MR of PUFA levels to proxy downregulated *FADS1* effects on MS severity.

To further interrogate genetic insights from colocalized loci while minimizing horizontal pleiotropy, we performed single-instrument cis-MR analyses using functional variants within the cis-gene regions of *FADS1/2*. We selected rs174546C>T, located within the 3′-untranslated region (3′-UTR) of *FADS1*, as a functional cis-instrument to genetically proxy the effect of *FADS1* perturbation on MS severity. The T allele has previously been shown to create a miR-149-5p binding site, resulting in allele-specific repression of *FADS1* expression. Genotype-Tissue expression data analysis demonstrated that the rs174546 TT genotype was consistently associated with significantly reduced *FADS1* expression across multiple brain regions, liver, and whole blood. In contrast, the TT genotype was not associated with significant reductions in the expression of *FADS2*, *MYRF*, or *TMEM258*, and generally showed modestly increased expression of these genes across brain tissues **(Supplementary File)**. Furthermore, carriers of the rs174546 T allele in Europeans have a consistent PUFA lipid profile characterized by increased levels of Δ6-desaturase (*FADS2*) products and decreased levels of Δ5-desaturase (*FADS1*) products(29, 30). Experimental evidence also suggests reduced *FADS1* enzymatic activity among T carriers(31).

We performed cis-MR analyses using circulating plasma metabolites within the FADS1/2-regulated PUFA pathway as downstream biomarkers of genetically proxied *FADS1* perturbation, leveraging the functional cis-acting variant rs174546C>T as a single instrumental variable. Wald ratio estimates indicated a consistent pattern across metabolites: genetically proxied reductions in Δ5-desaturase (*FADS1*) activity, as reflected by increased upstream FADS2-derived intermediates and decreased downstream FADS1-derived products, were associated with greater MS severity. This pathway-consistent directionality supports a role for *FADS1* in linking PUFA metabolism to disease severity (**Figure 5**).

Additionally, we identified evidence of moderate colocalization between circulating succinoyltaurine levels and MS severity at the *CYP4F2* locus (PP.H4 > 0.4). The identified shared causal variant was rs2108622, c.1297G>A (p.Val433Met), which is a missense mutation, as reported in PharmVar, and this polymorphism decreases CYP4F2 protein levels rather than mRNA transcription or enzymatic activity.

For *CYP4F2*, we selected the rs2108622C>T (p.V433M) variant in which the T allele is associated with reduced *CYP4F2* protein levels. In European mGWAS records, this variant is associated with higher circulating levels of phylloquinone and phosphatidylethanolamine (25:0), and lower metabolite levels downstream of CYP4F2 activity. We then used circulating plasma metabolite levels downstream of CYP4F2 activity as exposures in cis-MR analyses, with rs2108622 as the instrumental variable. Wald ratio estimates, scaled to a one standard deviation increase in metabolite levels, suggested that genetically proxied reductions in *CYP4F2* activity were associated with greater MS severity (**Figure 6**).

**Figure 6.**
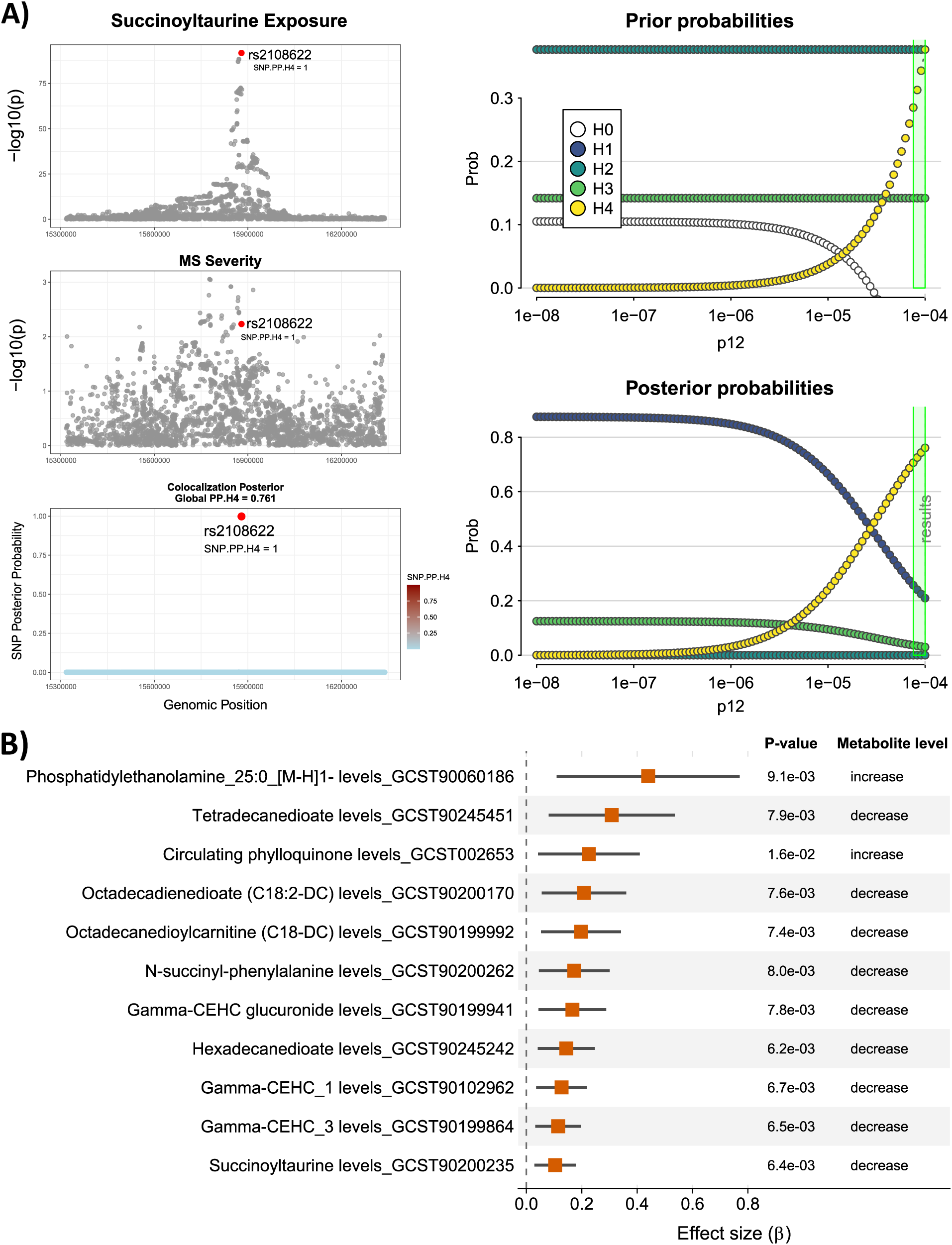
cis-MR of Metabolite Changes related to *CYP4F2* perturbation on MS Severity **A)** Regional colocalization plot showing SNP-level association signals for the metabolite trait and MS severity across the colocalized locus. The right panel presents the coloc sensitivity analysis, illustrating the impact of varying prior probabilities on posterior probabilities for each colocalization hypothesis (H0–H4). Robust support for H4 indicates a shared causal variant underlying both traits. **B)** Forest plot of GWAS-wide cis-MR estimates for CYP4F2-related metabolites, assessing genetically proxied CYP4F2 perturbation effects on MS severity.

### Brain-specific single-cell cis-MR analysis of proxied FADS1 perturbation on MS severity

To examine effects within the brain, we used rs174546 as an eQTL instrument for *FADS1*, across seven brain cell types (n = 474)(29, 32). This allowed us to assess the impact of genetically proxied FADS1 perturbation on MS severity at cell type–specific resolution. The T allele was significantly associated with reduced FADS1 expression in astrocytes, oligodendrocytes, and both inhibitory and excitatory neurons, but not in endothelial cells, microglia, or oligodendrocyte precursor cells. Using this variant as a single-instrument MR, Wald ratio estimates suggest that reduced *FADS1* expression in these cell types was associated with increased MS severity **(Figure 7).** Colocalization analyses supported the presence of a shared causal variant in each relevant cell type, further implicating reduced *FADS1* expression in specific neuronal and glial cell populations as a contributor to MS severity.

**Figure 7.**
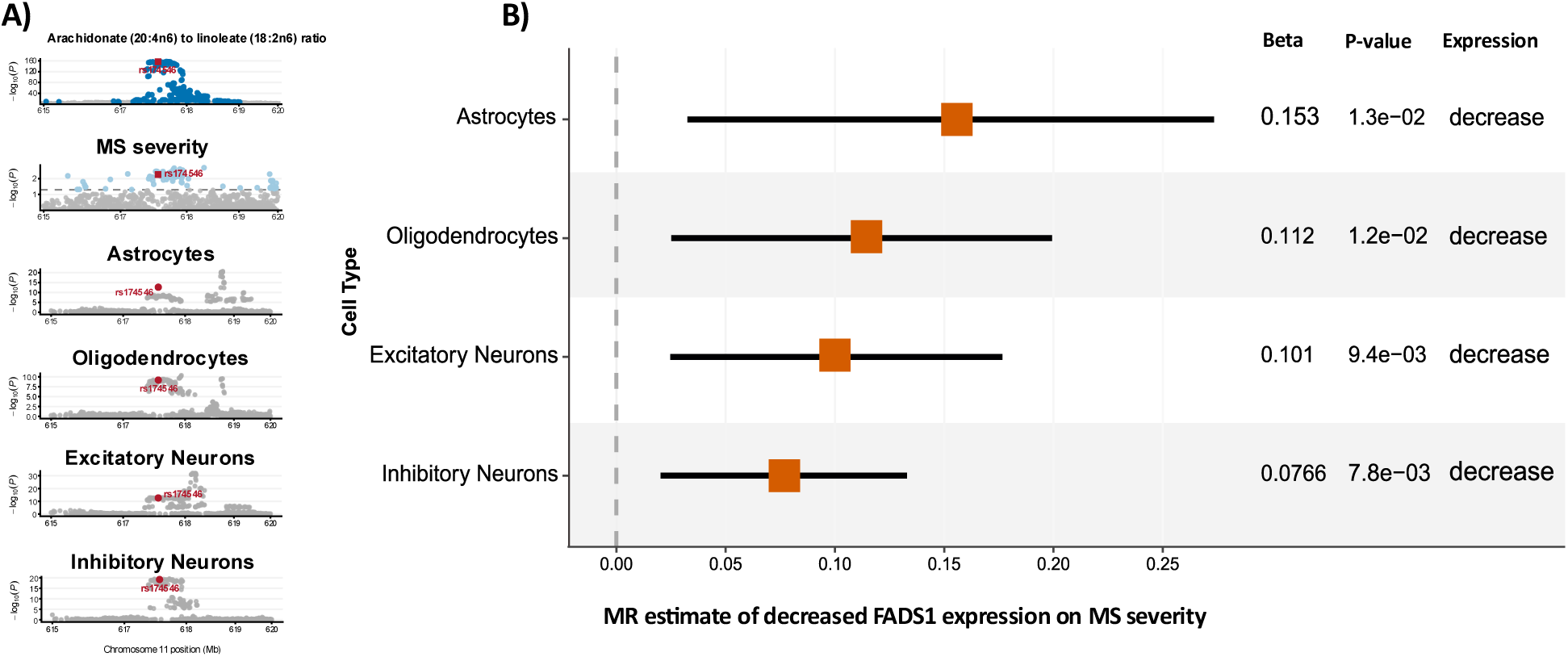
Single-cell Cis-MR effects of downregulated *FADS1* on MS severity across brain cells **A)** Multi-trait colocalization showing a shared causal variant across metabolite levels, MS severity, and brain cell–type expression at the FADS locus; **B)** Forest plot of cis-MR estimates for genetically proxied FADS1 downregulation in brain cells on MS severity using rs174546 as the cis-eQTL instrument.

## Discussion

The biological factors underlying variability in MS severity are not well understood. Although growing evidence links metabolic dysregulation to MS progression, it remains unclear whether these changes are causal or merely correlated with the disease outcomes. This uncertainty has hindered efforts to identify the specific mechanisms contributing to MS severity.

In this study, we implemented a multi-layered MR framework in which genetic variants serve as instrumental variables to systematically evaluate the impact of over 1,000 plasma metabolites and metabolite ratios on MS severity. First, a bidirectional, metabolome-wide MR analysis identified a subset of amino acid metabolites and lipid-related pathways consistently associated with MS severity. Next, we traced the genomic coordinates of IVs underlying significant causal metabolites and mapped them to nearby enzyme-coding regions. We then performed genetic colocalization at these loci, identifying shared variants that influence both metabolite levels and MS severity, including signals in the *FADS1/2* cluster and the *CYP4F2* genomic region.

Previous studies support the role of *FADS1/2* enzyme-coding genes in neurological conditions, consistent with our findings for MS severity. Previous MR results have shown that altered *FADS1* expression in multiple brain regions exerts a causal effect on cognitive function(32, 33)and reported a causal association between altered omega-3 and omega-6 fatty acid levels with schizophrenia(34). A large GWAS has identified the *FADS1/2* locus as a replicated risk region for bipolar disorder(35), and functional knockout mouse models linked reduced desaturase activity to neuropsychiatric phenotypes(36). In addition, cortical tissue studies have reported that *FADS1* haplotypes modulate estimated desaturase activity and fatty acid composition in the prefrontal cortex, further supporting an influence of FADS-driven PUFA remodeling within the brain (37).

The *FADS1/2* locus represents a central regulatory node in endogenous PUFA metabolism, and our findings provide convergent genetic evidence linking this pathway to MS severity. By leveraging a functional cis-variant to proxy reduced *FADS1* activity, we observed a coherent pattern across the PUFA biosynthetic cascade: accumulation of upstream (FADS2-derived) intermediates, and depletion of downstream (FADS1-derived) products were both associated with greater MS severity. This consistent directionality across metabolites strongly implicates *FADS1* as the key rate-limiting determinant linking PUFA metabolism to disease progression. This interpretation is supported by prior work demonstrating that variation at the FADS locus influences desaturase activity and fatty acid composition in both peripheral and brain tissues, with downstream effects on neuroinflammatory and neurobiological(32, 37). The locus is characterized by extensive linkage disequilibrium and functional haplotypes that modulate *FADS1/2* expression, resulting in substantial inter-individual variability in long-chain PUFA synthesis. Such variation is biologically meaningful given the role of PUFAs in regulating inflammatory signaling, membrane composition, and neuronal function(30). From a translational perspective, these findings suggest that genetic variation at the FADS locus may contribute to heterogeneity in MS severity and could inform risk stratification. In particular, individuals carrying low-activity FADS1/2 haplotypes (i.e., associated with reduced long-chain PUFA synthesis) may represent a subgroup in whom targeted dietary or supplementation strategies could mitigate inflammatory burden and influence disease progression.

We also identified a putative causal genetic signal at the *CYP4F2* locus, where metabolite-modulating variants colocalized with MS severity. *CYP4F2* encodes a cytochrome P450 ω-hydroxylase that metabolizes long- and very-long-chain PUFAs, including arachidonic acid, to generate 20-HETE and related ω-hydroxylated eicosanoids, thereby regulating PUFA turnover and bioactive lipid signaling. Several lines of evidence, including prior genetic studies, implicate *CYP4F2* in MS pathogenesis and disease severity, as well as in optic neuritis susceptibility and fingolimod metabolism (38, 39).

Our cis-MR analysis identifies *CYP4F2* perturbation as a potential factor in circulating metabolic dysregulation in MS. The exact molecular mechanisms by which *CYP4F2* dysfunction worsens disease severity require further detailed experimental investigation. These mechanisms may involve various pharmacogenetic variants that affect the enzyme’s efficiency, influencing PUFA-derived inflammatory signaling in the brain’s circulation. This supports the idea that individual differences in *CYP4F2* pharmacogenetics could modify neurodegenerative severity by altering PUFA metabolism.

In addition to the lipid metabolic pathways highlighted by our complementary analyses, our primary metabolome-wide MR analysis also implicated mitochondrial energy metabolism in MS severity. Specifically, we observed associations for the mitochondrial intermediate α-ketoglutarate (AKG), the α-ketoglutarate-to-proline ratio, and multiple succinyl-CoA–related metabolites, collectively suggesting a potential contribution of mitochondrial metabolic dysfunction to disease progression. These findings may reflect the role of mitochondrial dysfunction in MS severity and support the benefits of AKG supplementation, which has previously been shown to influence epigenetic reprogramming and ameliorate other neurodegenerative disorders, suggesting its potential neuroprotective role and suggesting that ketogenic interventions may hold therapeutic relevance for MS (32, 33).

Additionally, our metabolome-wide MR results identified specific AAs, such as arginine, and the serine-to-pyruvate ratio, as being associated with increased MS severity, whereas betaine was associated with reduced disease severity. Betaine is a neuroprotective methyl donor that has been reported to regulate the liver-brain axis by modulating lipid oxidation in the liver and phospholipid metabolism in the brain (40). A recent study on the cuprizone (CPZ)-induced multiple sclerosis mouse model suggested that oral administration of betaine could serve as a novel adjunct therapy against functional impairment in MS patients(41). Our findings are consistent with these results, as betaine is associated with lower MS severity.

Our study has several strengths. First, we used the largest available GWAS data for both metabolite levels and MS severity, enabling a comprehensive and systematic assessment of candidate metabolites. Second, we conducted extensive sensitivity analyses to assess the robustness and reliability of our findings across a range of MR methods. Third, to reduce potential horizontal pleiotropy, colocalization analyses were performed to assess whether the same causal variants underlie both metabolite levels and MS severity, thereby suggesting potential molecular pathways underlying the reported causal effects. Finally, functional cis-acting variants were used as instruments in a multi-exposure cis-MR framework to proxy the effect of perturbation of the target genes on MS severity, and the inferred causal relationships were corroborated at the levels of plasma metabolite concentrations and single-cell gene expression across CNS cell types.

We acknowledge the limitations of this study. First, our study was restricted to GWAS data from individuals of European ancestry, and replication across diverse populations is needed. Additionally, conducting sex- and age-stratified analyses is necessary to delineate the effects of metabolites within specific subgroups. Collectively, these results provide important insights that warrant further validation through clinical and experimental research.

In conclusion, our systematic MR framework provides evidence that dysregulated PUFA metabolism, driven in part by genetic variation at the FADS locus, contributes to MS severity. This relationship is supported by consistent pathway-level effects and cell type-specific analyses in the brain. We also identify a complementary metabolic axis involving *CYP4F2* activity and mitochondrial function, further implicating lipid metabolism in disease progression. By anchoring metabolite-severity relationships in genetic instruments, our findings extend observational associations and highlight specific metabolic pathways as candidates for mechanistic investigation and therapeutic targeting in MS.

## Data Availability

All datasets analyzed in the current study are available from the GWAS Catalog and from the International Multiple Sclerosis Genetics Consortium (IMSGC), subject to approval by the relevant Data Access Committee. Access to the single-cell brain eQTL data can be requested from the corresponding author and will be granted in accordance with institutional data governance and confidentiality policies.

https://www.ebi.ac.uk/gwas/publications/36635386

https://doi.org/10.1038/s41586-023-06250-x

## Notes

### Competing Interest Statement

The authors have declared no competing interest.

### Funding

- National Institutes of Health: R0NS190109328

