## Supplementary File for "Integrative Genetic Analyses of Lipid Metabolism and Multiple Sclerosis Severity Using Metabolome-Wide and Cis-Mendelian Randomization"

### Graphical Assessment of the Metabolome-wide MR Results

### Supplementary Figure 1: GETx: Tissue-Specific eQTL Effects of rs174546 Genotypes Across Colocalized Genes in the FADS-region

FADS1 (ENSG00000149485.19) and rs174546 (chr11_61802358_C_T_b38)

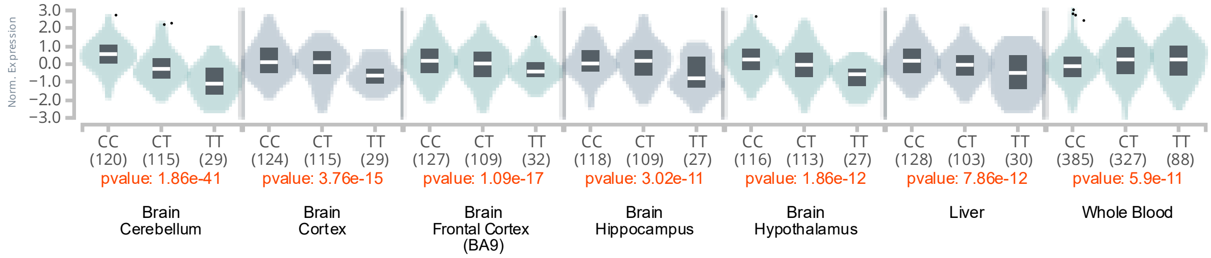

FADS2 (ENSG00000134824.14) and rs174546 (chr11_61802358_C_T_b38)

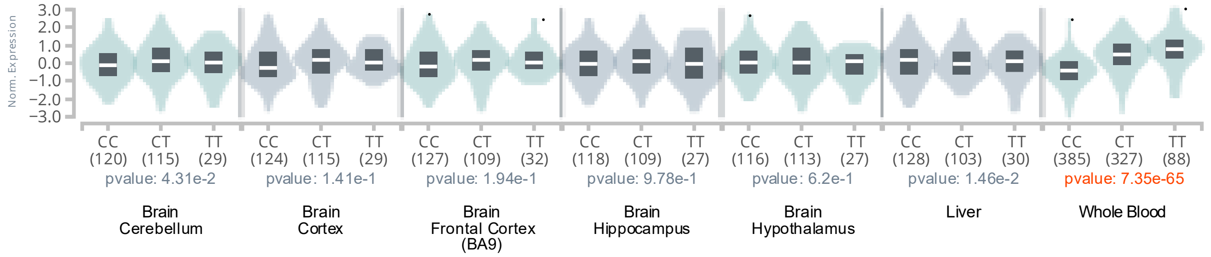

TMEM258 (ENSG00000134825 .16) and rs174546 (chr11_61802358_C_T_b38)

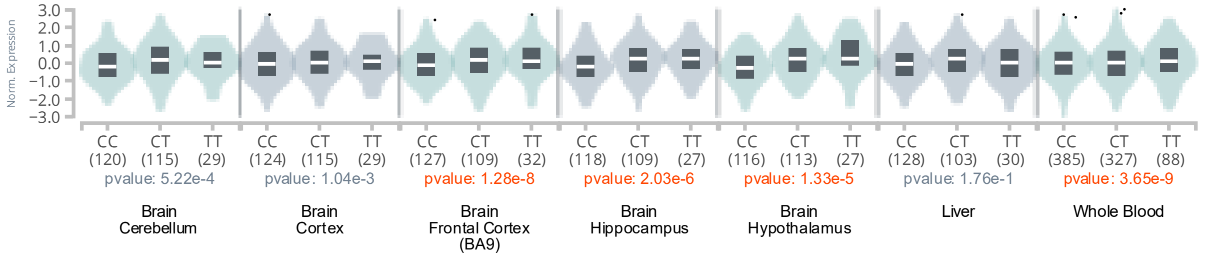

MYRF (ENSG00000124920.14) and rs174546 (chr11_61802358_C_T_b38)

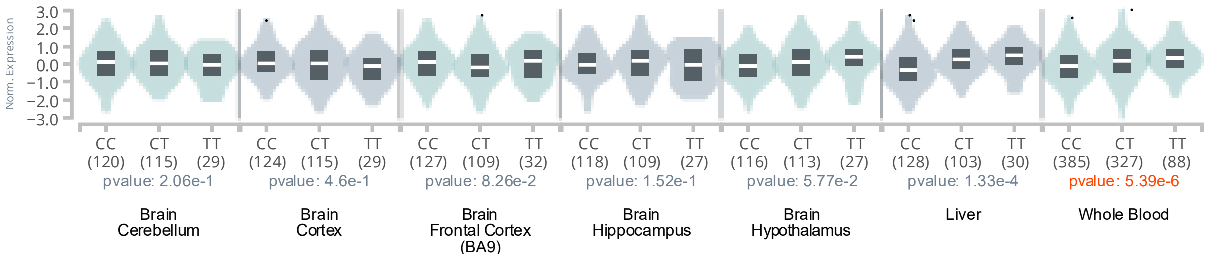

Genotype-Tissue Expression data used for visualizing this plot were obtained from the GTEx Portal on 05/20/2026.

#### 1-stearoyl-2-arachidonoyl-GPI (18:0/20:4)

| method | nsnp | b | se | pval |
|---|---|---|---|---|
| MR Egger | 23 | -0.1236922 | 0.0543216 | 0.0333660 |
| Weighted median | 23 | -0.0626104 | 0.0346685 | 0.0709223 |
| Inverse variance weighted | 23 | -0.0709686 | 0.0238718 | 0.0029500 |
| Simple mode | 23 | -0.0253687 | 0.0577212 | 0.6645828 |
| Weighted mode | 23 | -0.0777155 | 0.0400173 | 0.0650432 |

##### Heterogeneity tests

| method | Q | Q_df | Q_pval |
|---|---|---|---|
| MR Egger | 17.74043 | 21 | 0.6653663 |
| Inverse variance weighted | 18.90793 | 22 | 0.6510070 |

##### Test for directional horizontal pleiotropy

| egger_intercept | se | pval |
|---|---|---|
| 0.0088367 | 0.0081783 | 0.2921693 |

Forest plot of single SNP MR

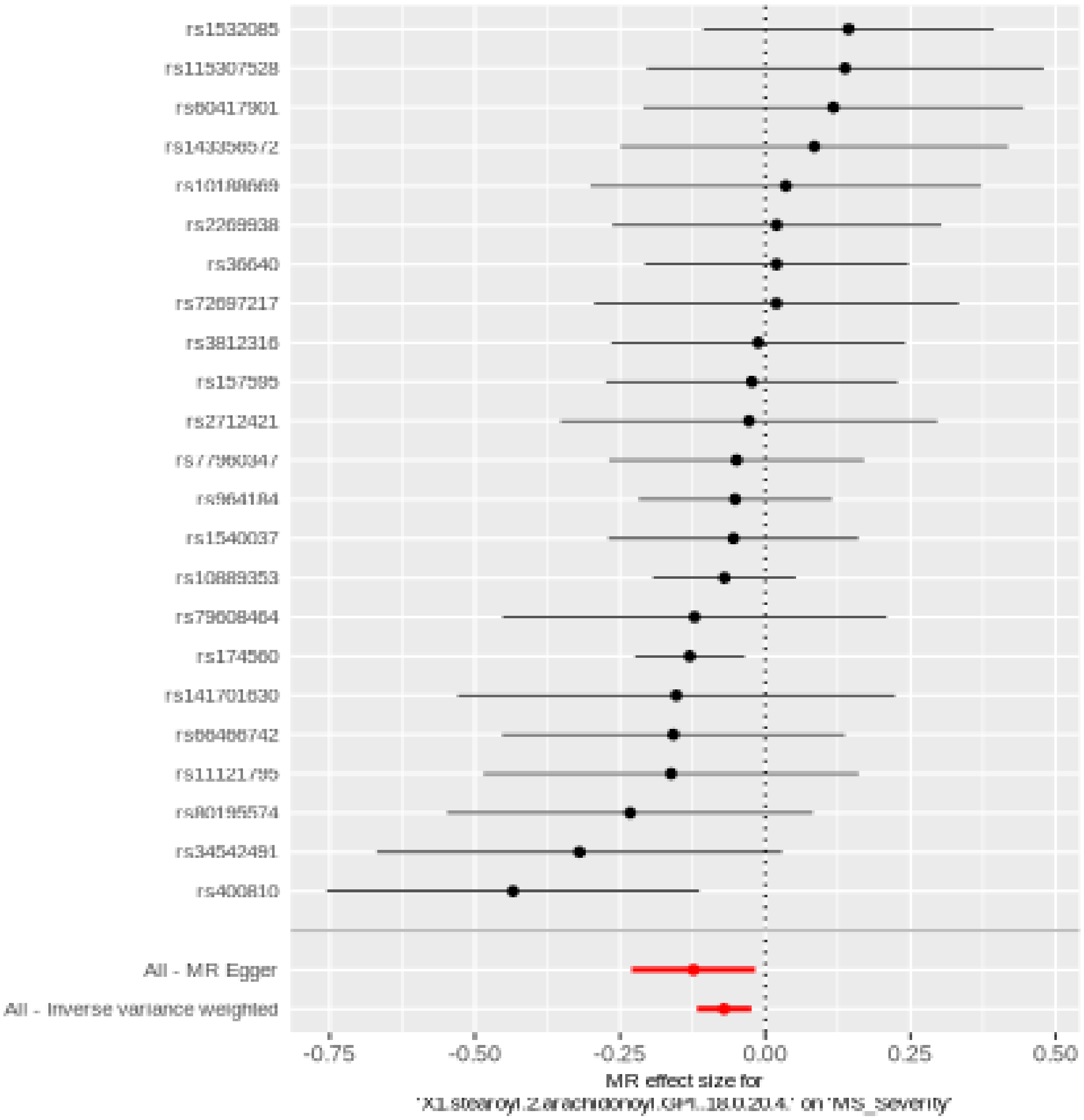

Comparison of results using different MR methods

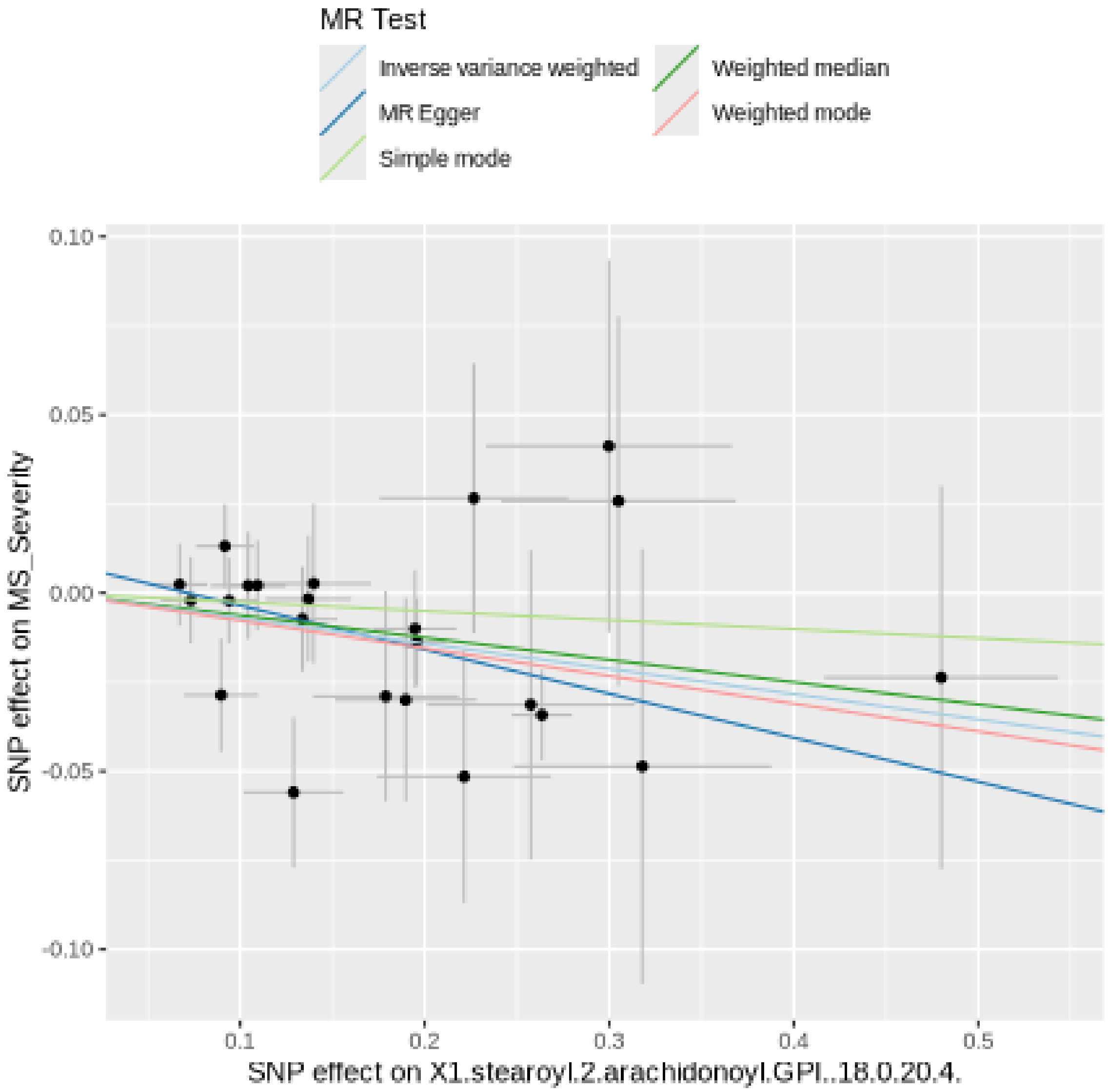

Funnel plot

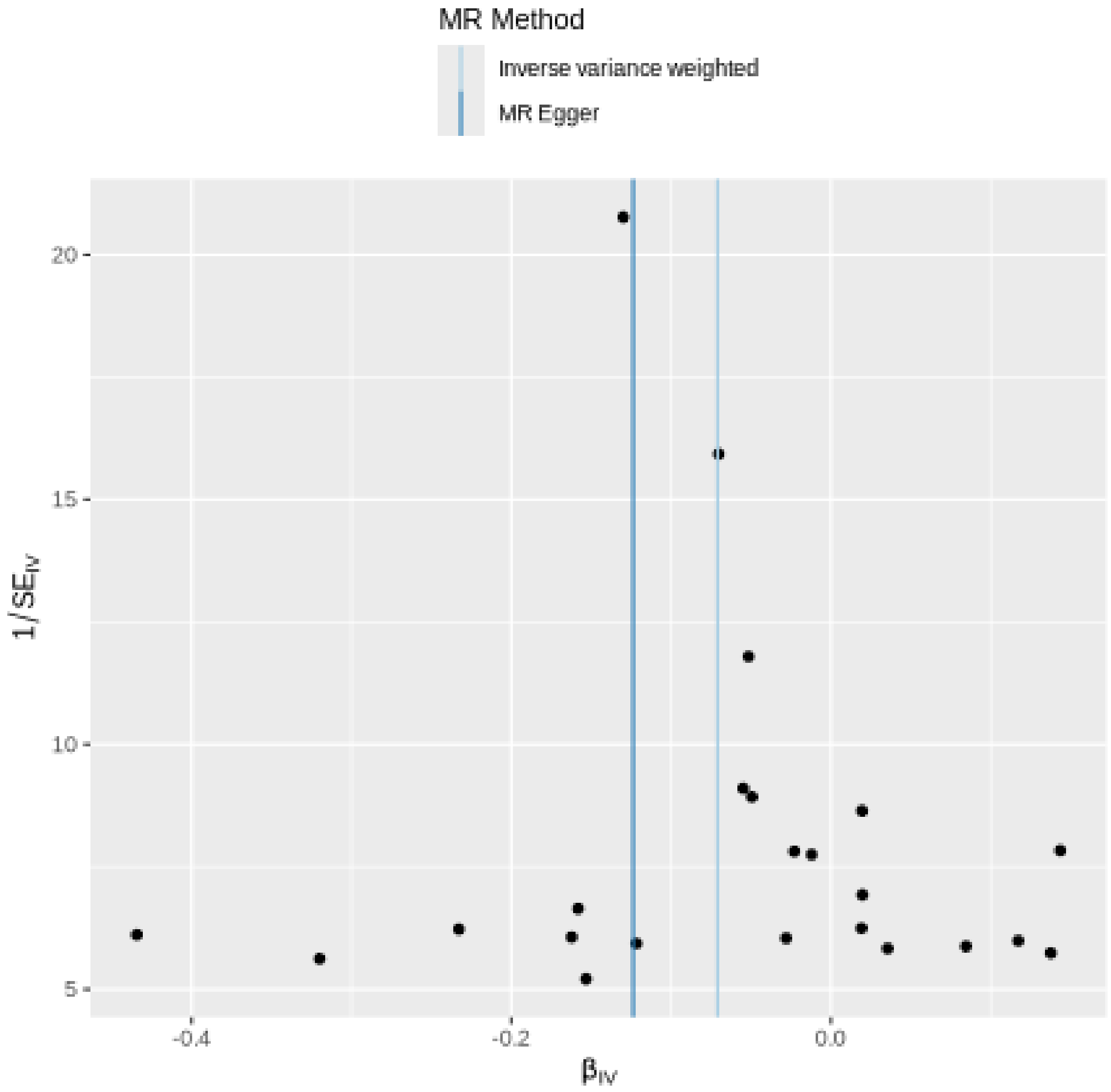
MR Method
Inverse variance weighted
MR Egger
20
15
10
5
1/SE_IV
-0.4
-0.2
0.0
β_IV

#### Leave-one-out sensitivity analysis

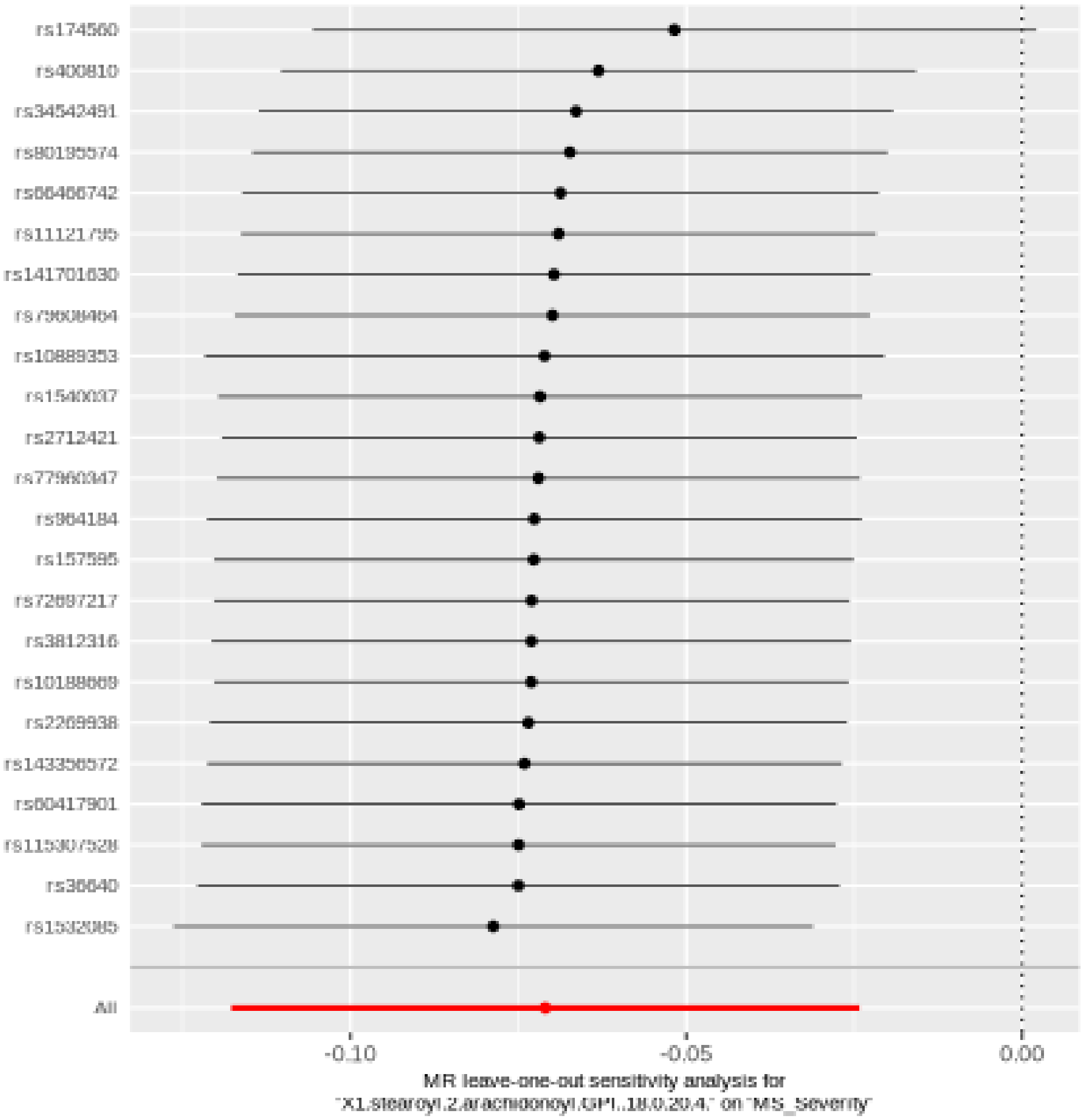

#### Palmitoylcarnitine (C16)

| method | nsnp | b | se | pval |
|---|---|---|---|---|
| MR Egger | 31 | 0.0025477 | 0.0794517 | 0.9746386 |
| Weighted median | 31 | -0.0765260 | 0.0450788 | 0.0895822 |
| Inverse variance weighted | 31 | -0.0788011 | 0.0315747 | 0.0125708 |
| Simple mode | 31 | -0.1103913 | 0.0913249 | 0.2361903 |
| Weighted mode | 31 | -0.0683170 | 0.0718040 | 0.3489853 |

##### Heterogeneity tests

| method | Q | Q_df | Q_pval |
|---|---|---|---|
| MR Egger | 35.88744 | 29 | 0.1768667 |
| Inverse variance weighted | 37.42573 | 30 | 0.1650570 |

##### Test for directional horizontal pleiotropy

| egger_intercept | se | pval |
|---|---|---|
| -0.0099606 | 0.0089338 | 0.2740349 |

Forest plot of single SNP MR

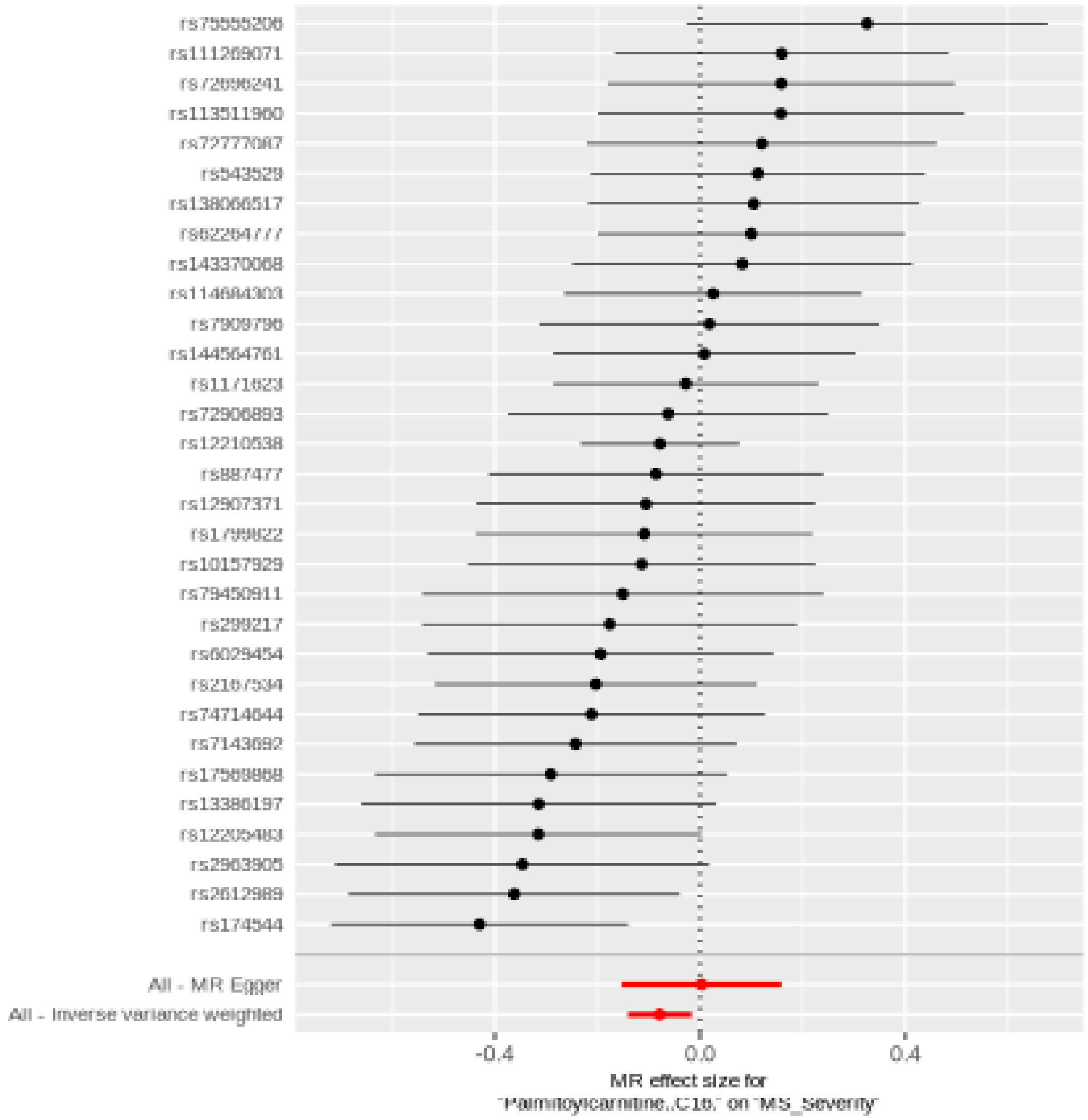

Comparison of results using different MR methods

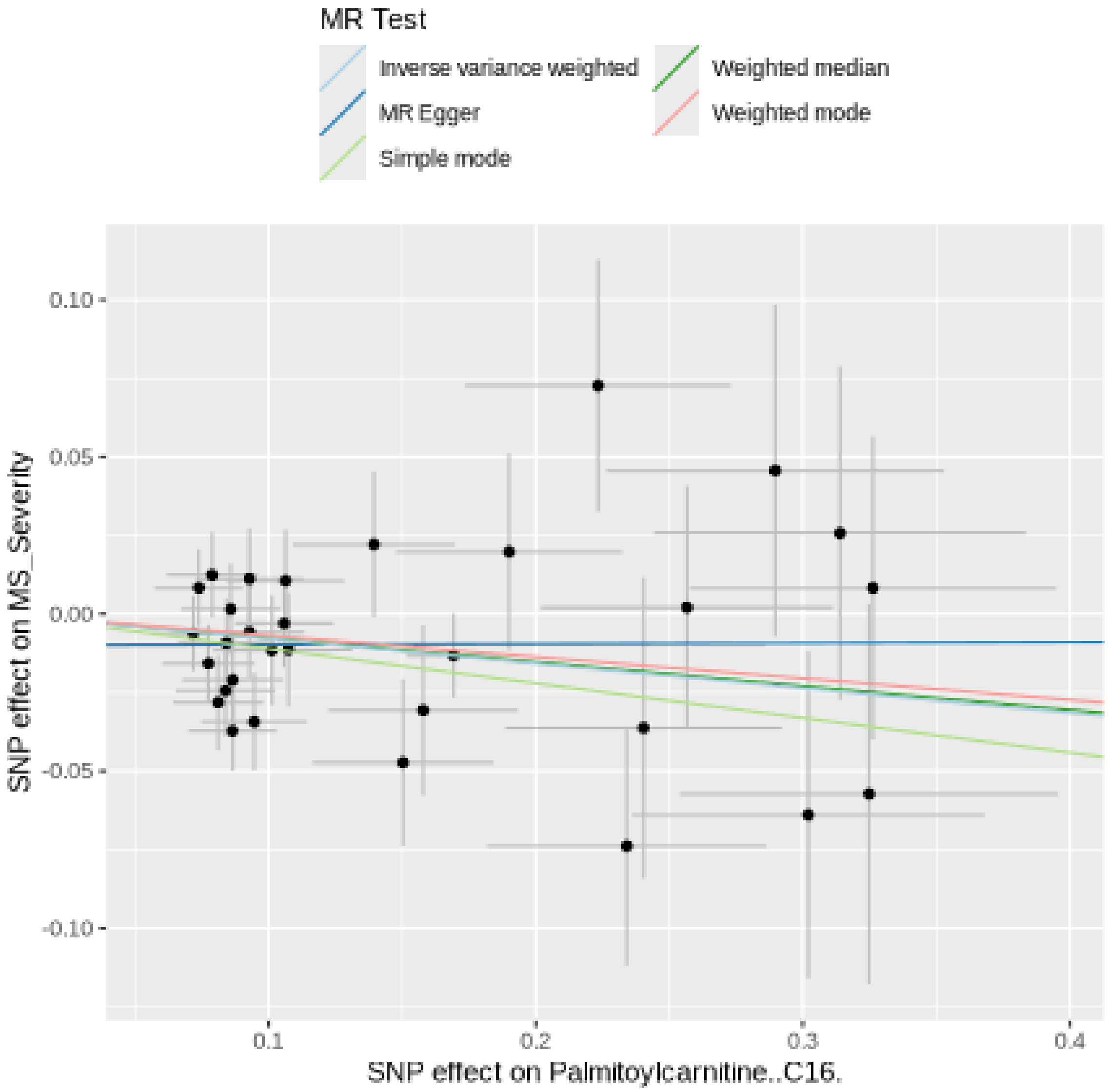

Funnel plot

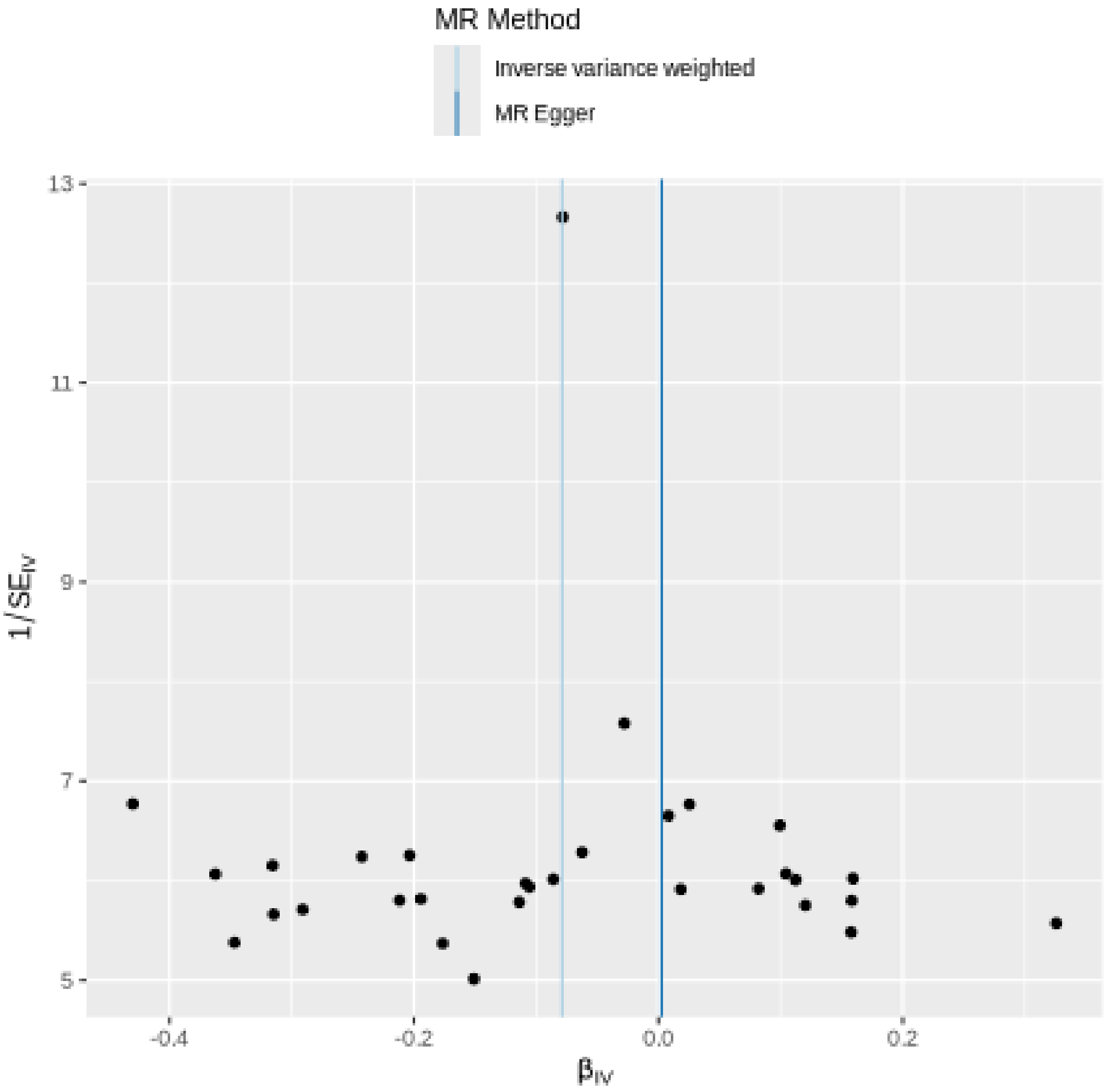
MR Method
Inverse variance weighted
MR Egger
13
11
9
7
5
1/SE$_{IV}$
-0.4
-0.2
0.0
0.2
β$_{IV}$

#### Leave-one-out sensitivity analysis

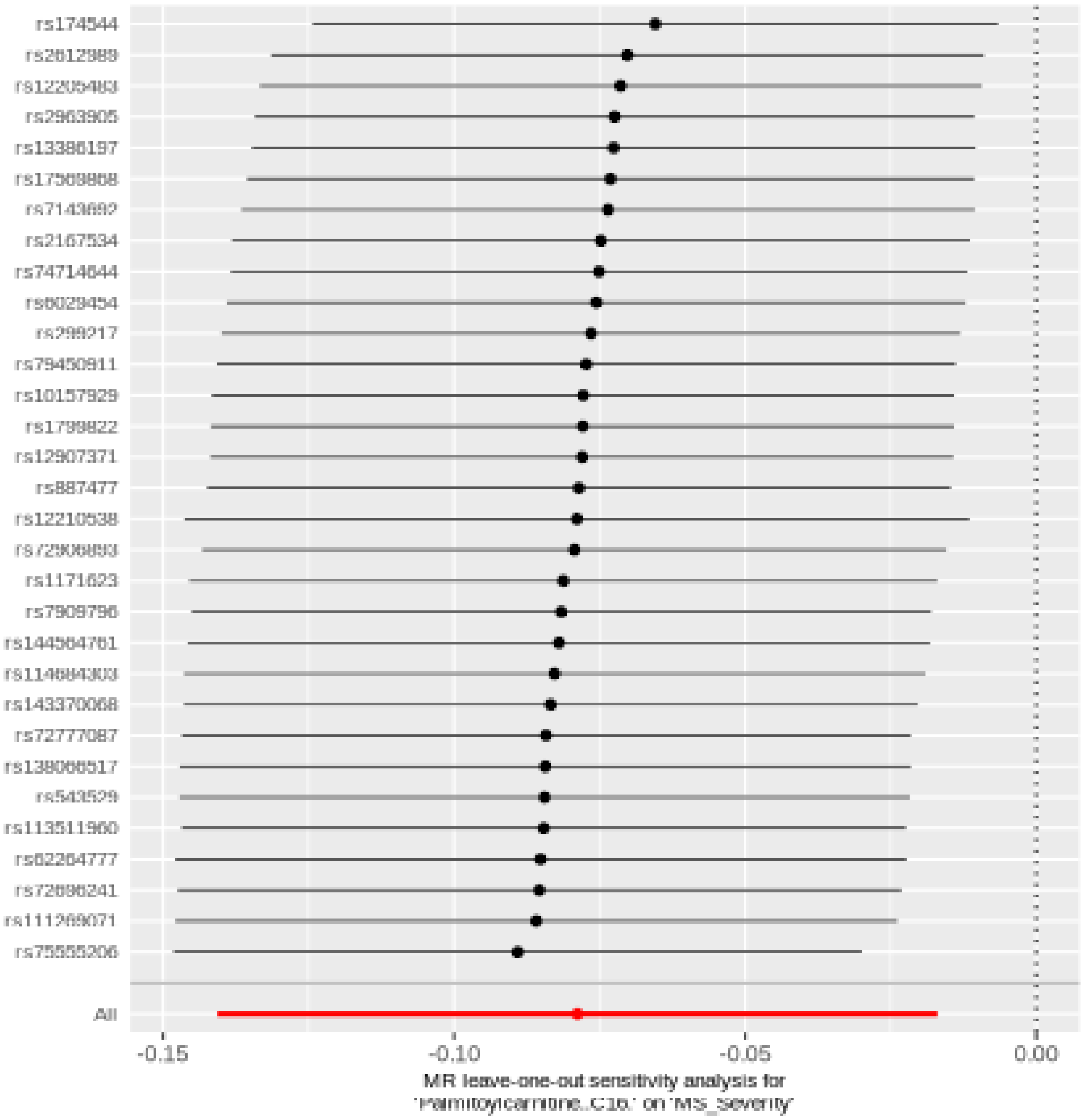

#### Propionylglycine

| method | nsnp | b | se | pval |
|---|---|---|---|---|
| MR Egger | 22 | 0.0935270 | 0.0450171 | 0.0508417 |
| Weighted median | 22 | 0.0747140 | 0.0317616 | 0.0186558 |
| Inverse variance weighted | 22 | 0.0472715 | 0.0237444 | 0.0464982 |
| Simple mode | 22 | 0.0536739 | 0.0737661 | 0.4748793 |
| Weighted mode | 22 | 0.0786827 | 0.0336939 | 0.0295344 |

##### Heterogeneity tests

| method | Q | Q_df | Q_pval |
|---|---|---|---|
| MR Egger | 16.43869 | 20 | 0.6890633 |
| Inverse variance weighted | 17.90140 | 21 | 0.6552351 |

##### Test for directional horizontal pleiotropy

| egger_intercept | se | pval |
|---|---|---|
| -0.0088807 | 0.0073429 | 0.2406016 |

Forest plot of single SNP MR

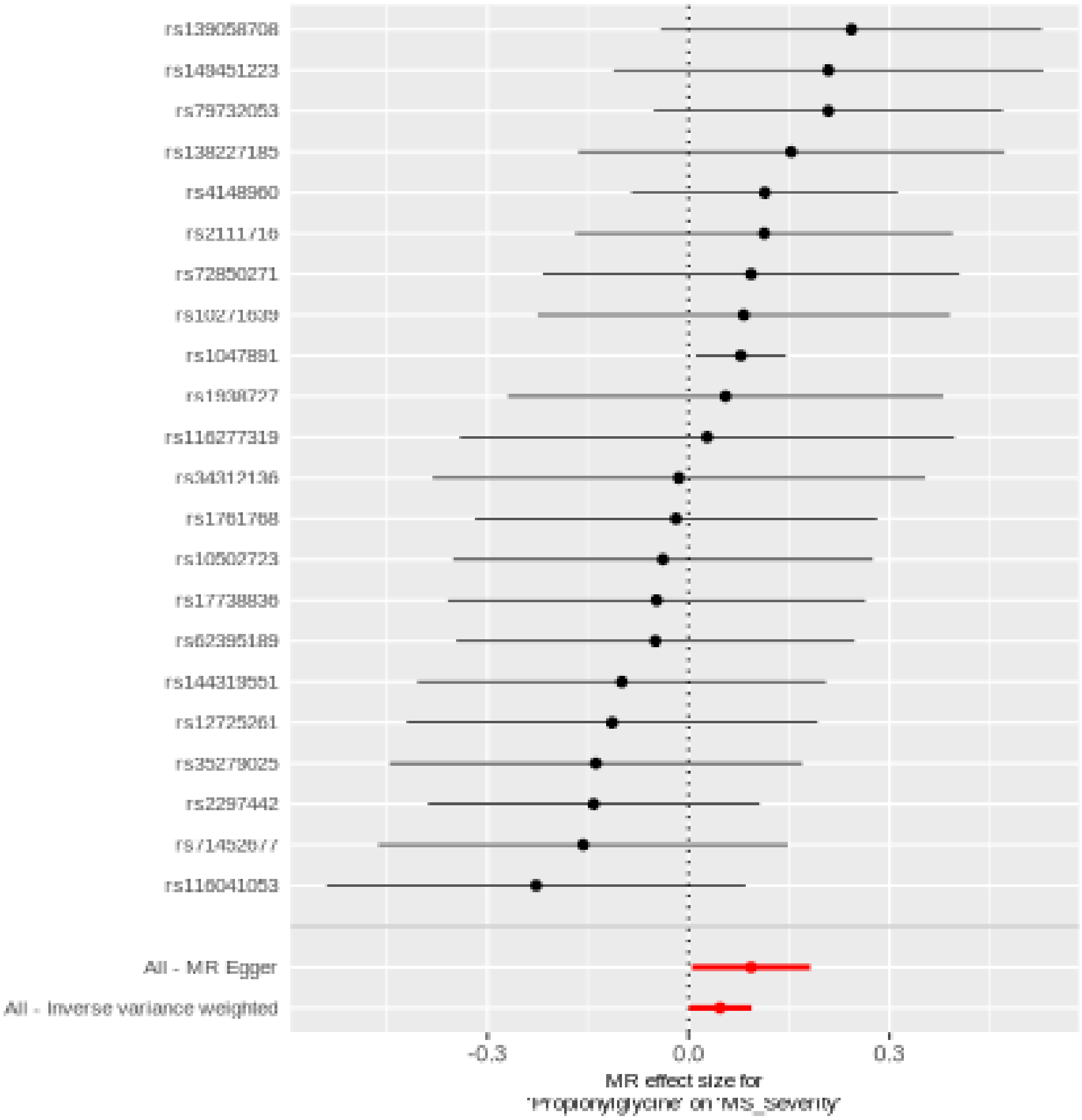

Comparison of results using different MR methods

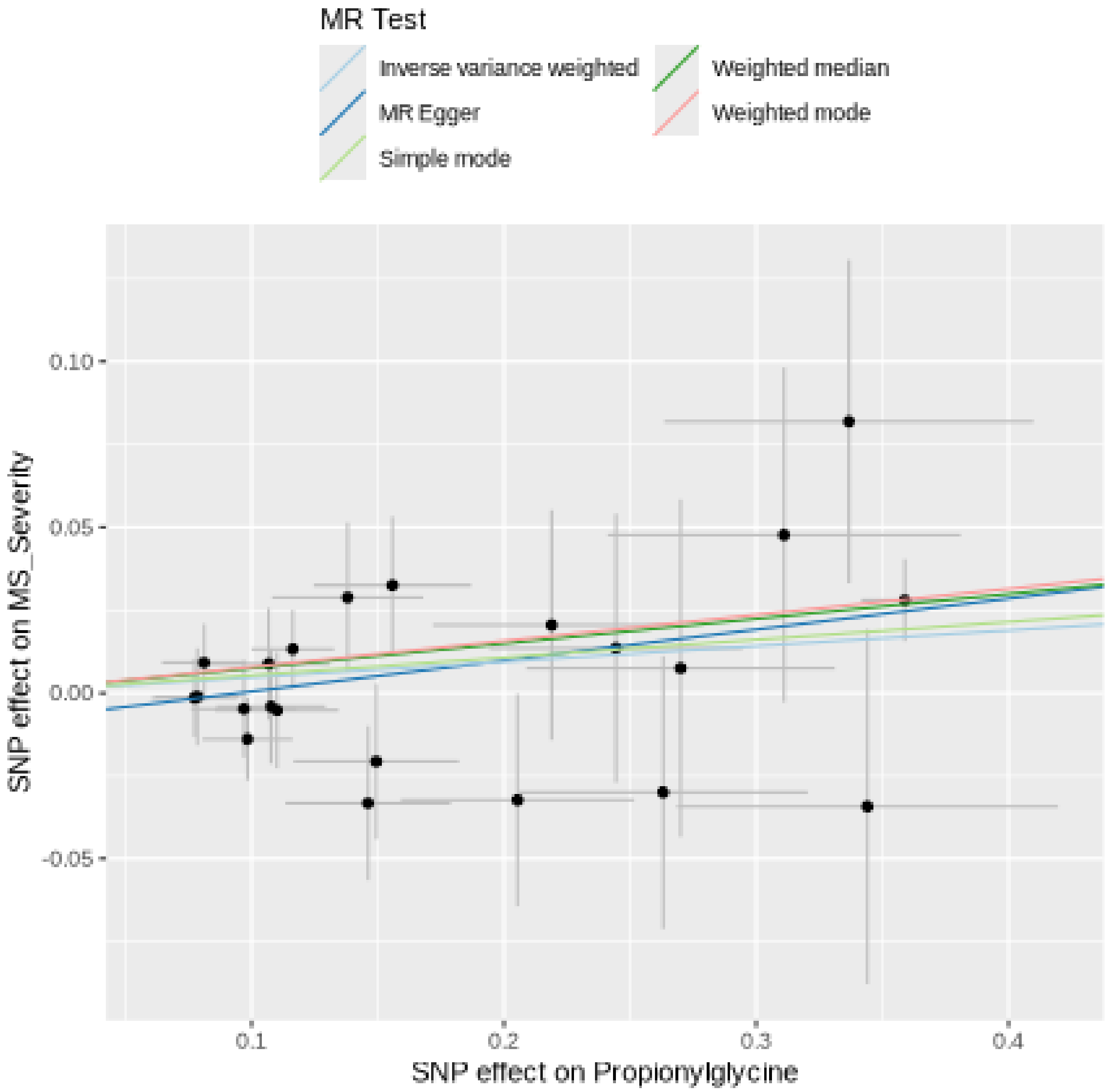

Funnel plot

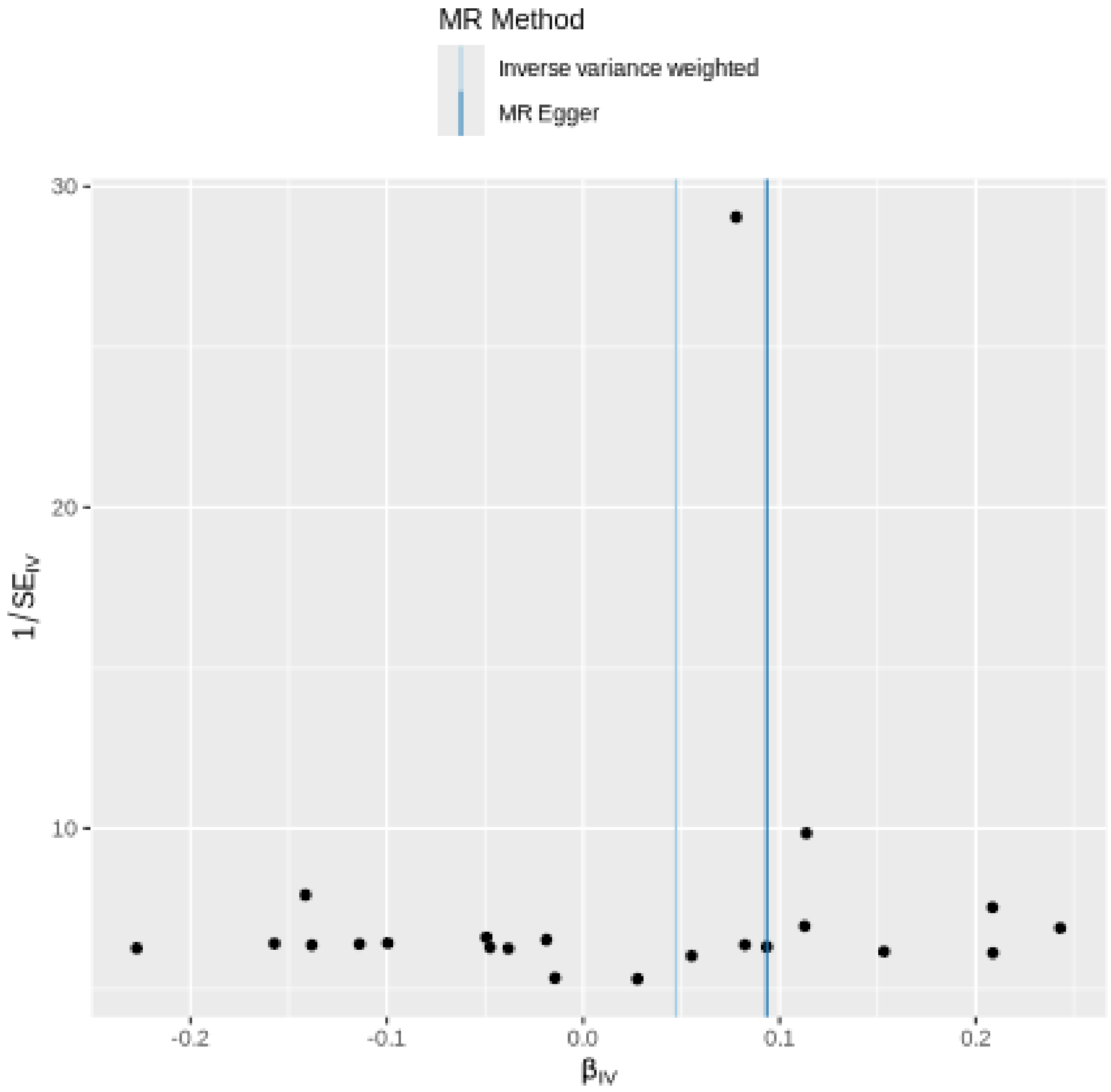

MR Method
Inverse variance weighted
MR Egger
30
20
10
-0.2
-0.1
0.0
0.1
0.2
1/SE_IV
β_IV

Leave-one-out sensitivity analysis

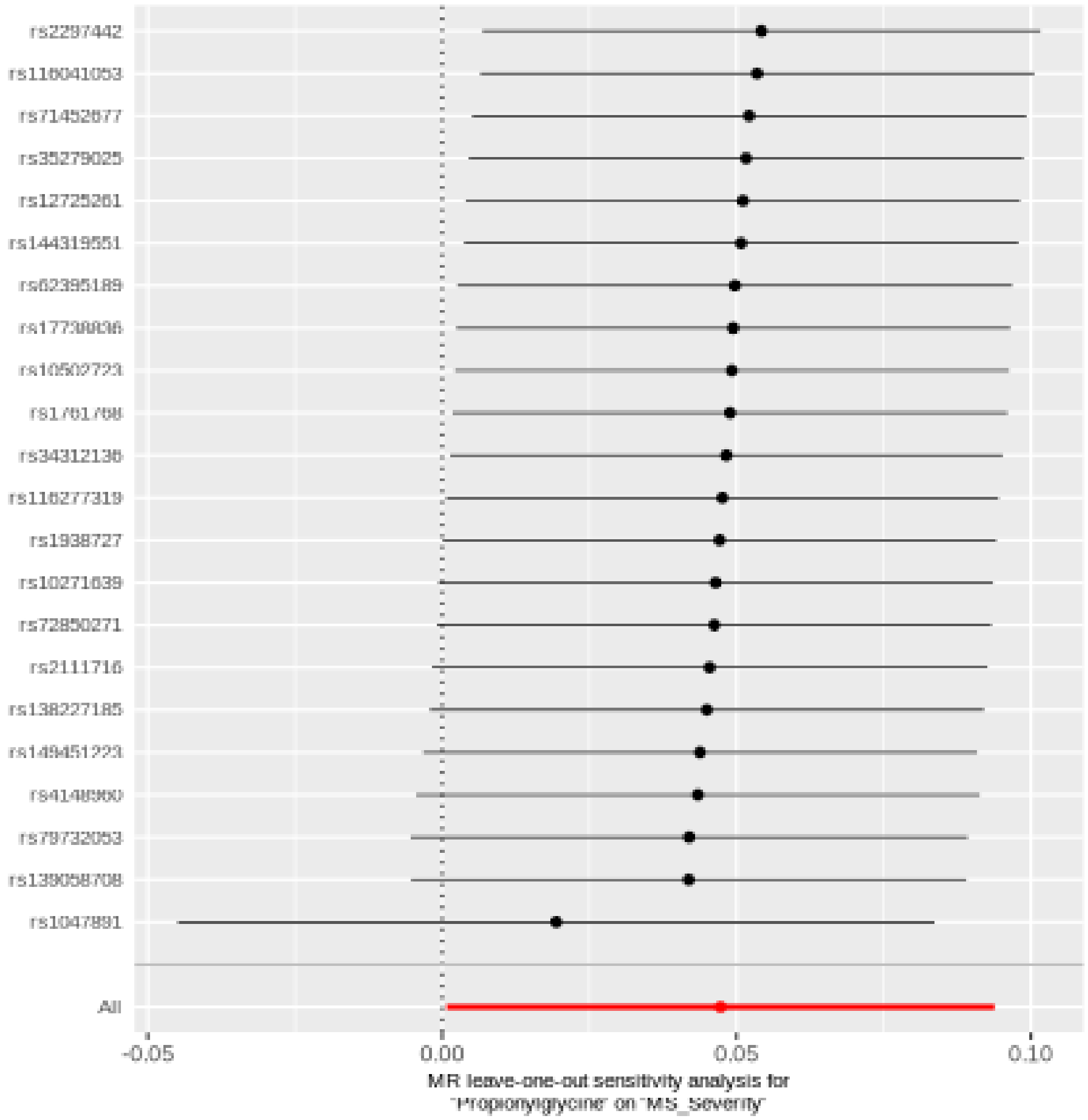

#### 3-hydroxy-2-ethylpropionate

| method | nsnp | b | se | pval |
|---|---|---|---|---|
| MR Egger | 32 | -0.0676920 | 0.0785331 | 0.3955501 |
| Weighted median | 32 | -0.0624493 | 0.0372561 | 0.0936962 |
| Inverse variance weighted | 32 | -0.0568178 | 0.0277013 | 0.0402582 |
| Simple mode | 32 | -0.0707340 | 0.0733840 | 0.3425608 |
| Weighted mode | 32 | -0.0585025 | 0.0638093 | 0.3663081 |

##### Heterogeneity tests

| method | Q | Q_df | Q_pval |
|---|---|---|---|
| MR Egger | 20.66387 | 30 | 0.8981034 |
| Inverse variance weighted | 20.68577 | 31 | 0.9199531 |

##### Test for directional horizontal pleiotropy

| egger_intercept | se | pval |
|---|---|---|
| 0.0012322 | 0.0083268 | 0.8833507 |

Forest plot of single SNP MR

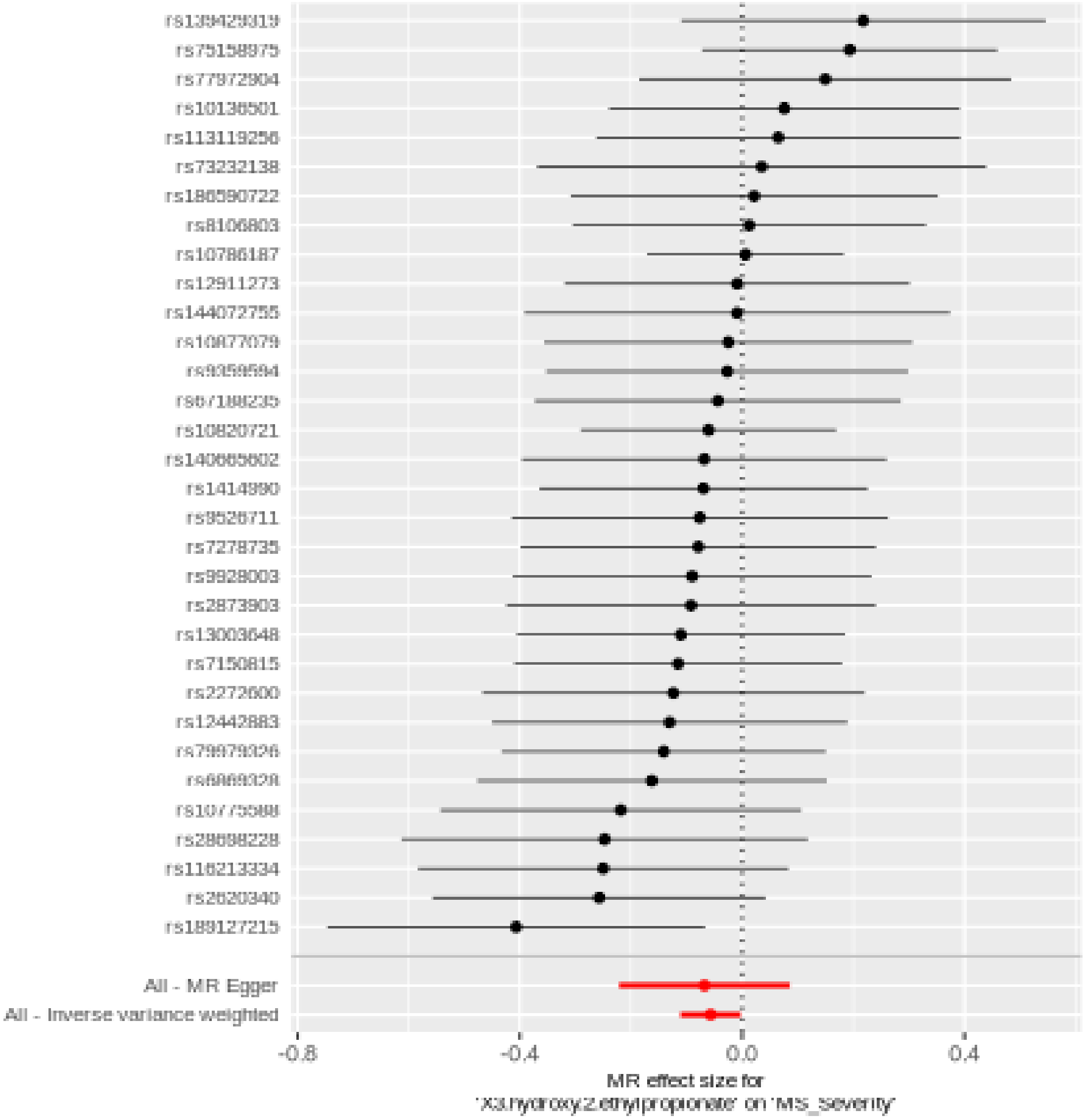

Comparison of results using different MR methods

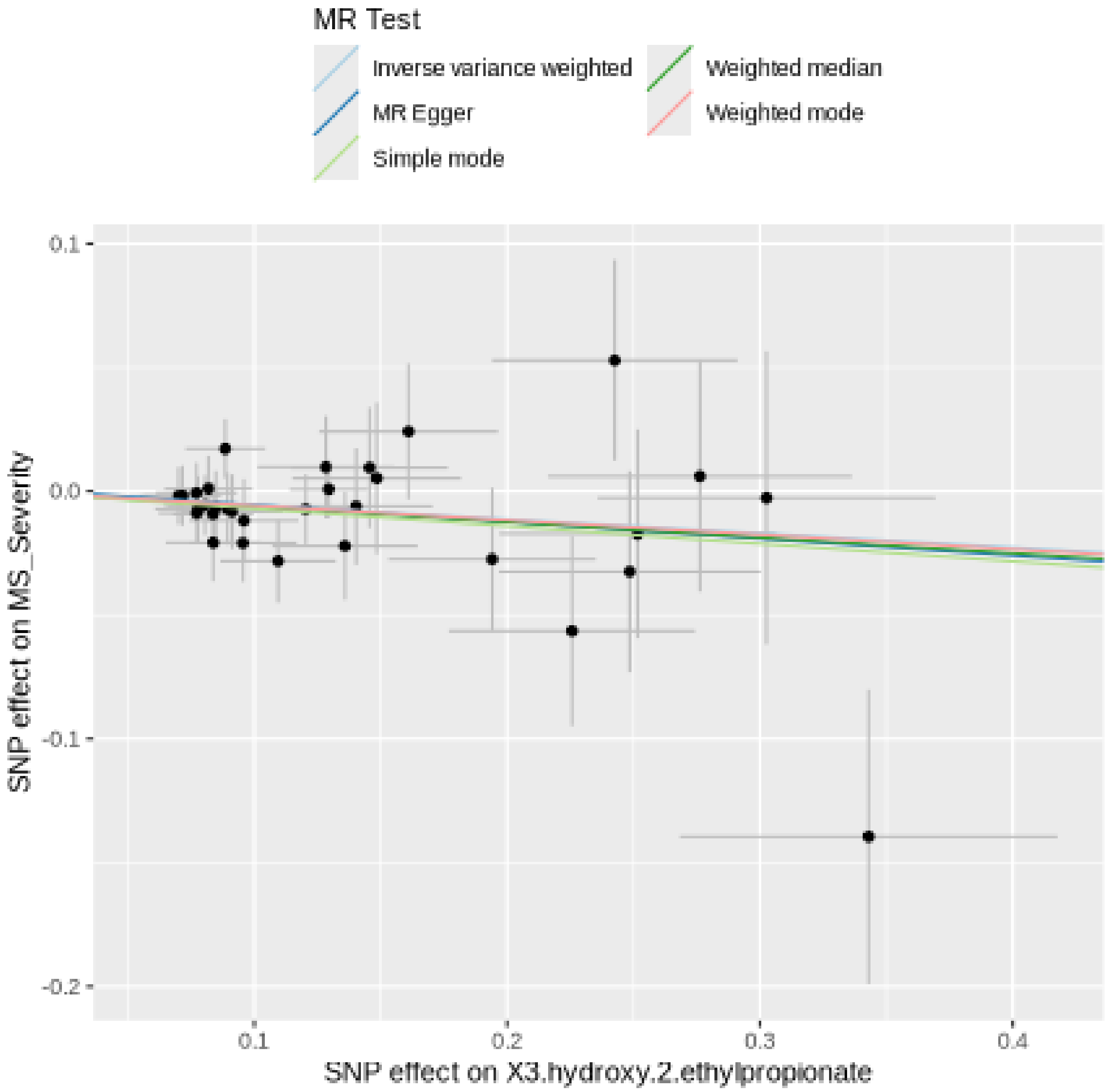

Funnel plot

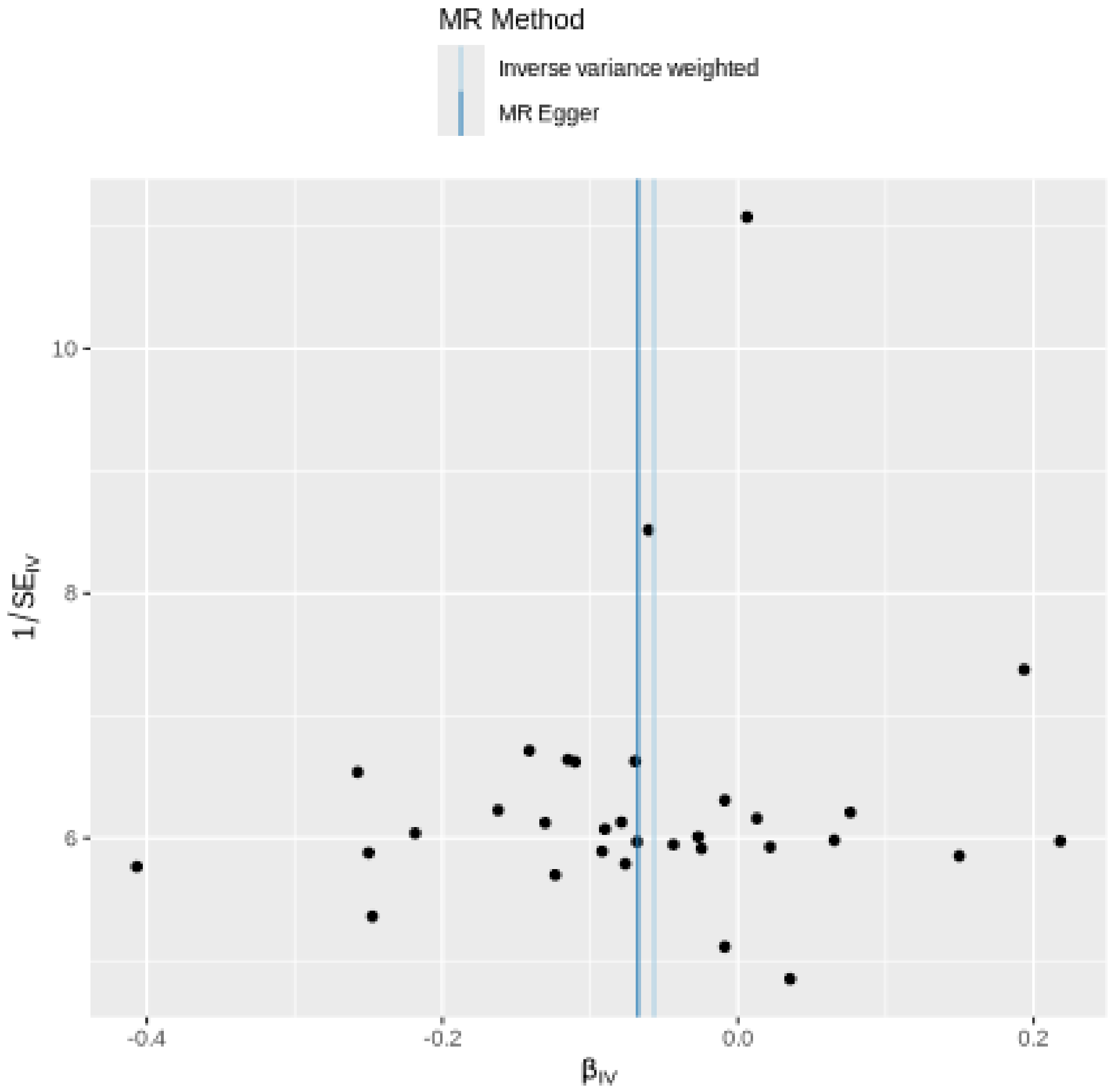
MR Method
Inverse variance weighted
MR Egger
10
8
6
1/SE$_{IV}$
-0.4
-0.2
0.0
0.2
β$_{IV}$

Leave-one-out sensitivity analysis

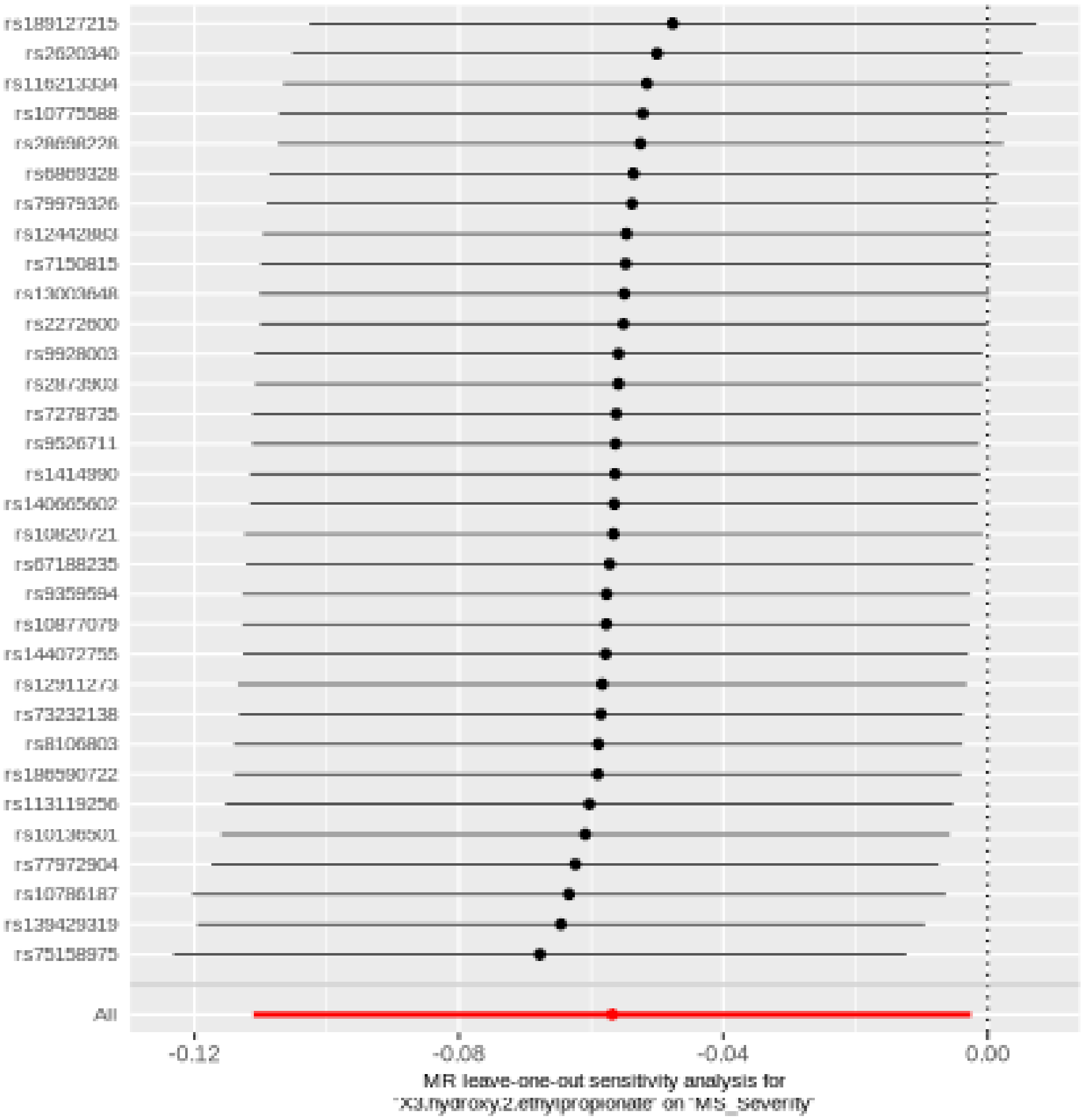

#### Gamma-glutamylglycine

| method | nsnp | b | se | pval |
|---|---|---|---|---|
| MR Egger | 33 | 0.0506957 | 0.0334741 | 0.1400357 |
| Weighted median | 33 | 0.0575120 | 0.0243865 | 0.0183561 |
| Inverse variance weighted | 33 | 0.0445275 | 0.0205526 | 0.0302725 |
| Simple mode | 33 | 0.0464920 | 0.0622990 | 0.4609503 |
| Weighted mode | 33 | 0.0562418 | 0.0245419 | 0.0286523 |

##### Heterogeneity tests

| method | Q | Q_df | Q_pval |
|---|---|---|---|
| MR Egger | 37.95984 | 31 | 0.1817861 |
| Inverse variance weighted | 38.02783 | 32 | 0.2138901 |

##### Test for directional horizontal pleiotropy

| egger_intercept | se | pval |
|---|---|---|
| -0.0011302 | 0.0047963 | 0.8152673 |

Forest plot of single SNP MR

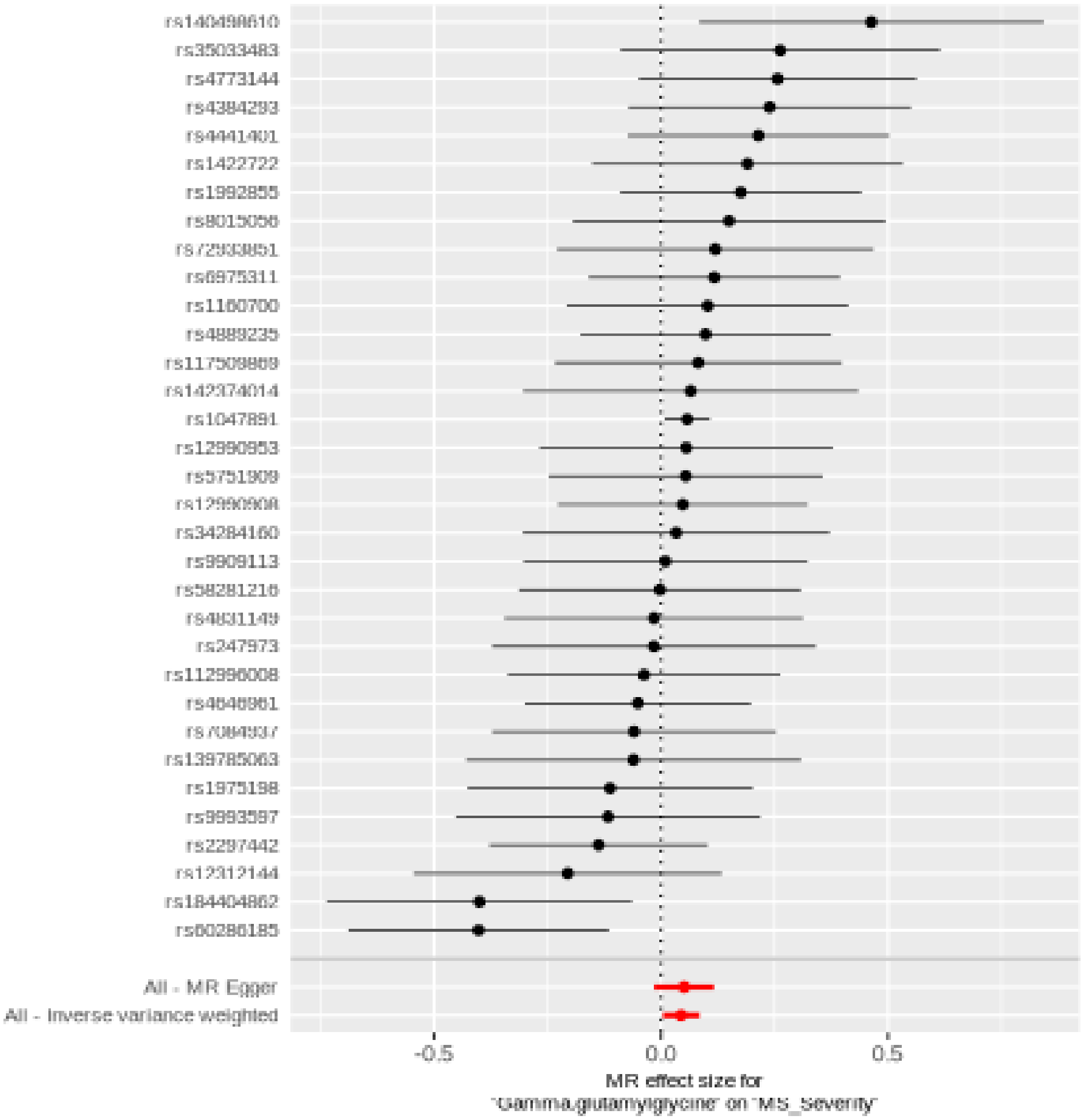

Comparison of results using different MR methods

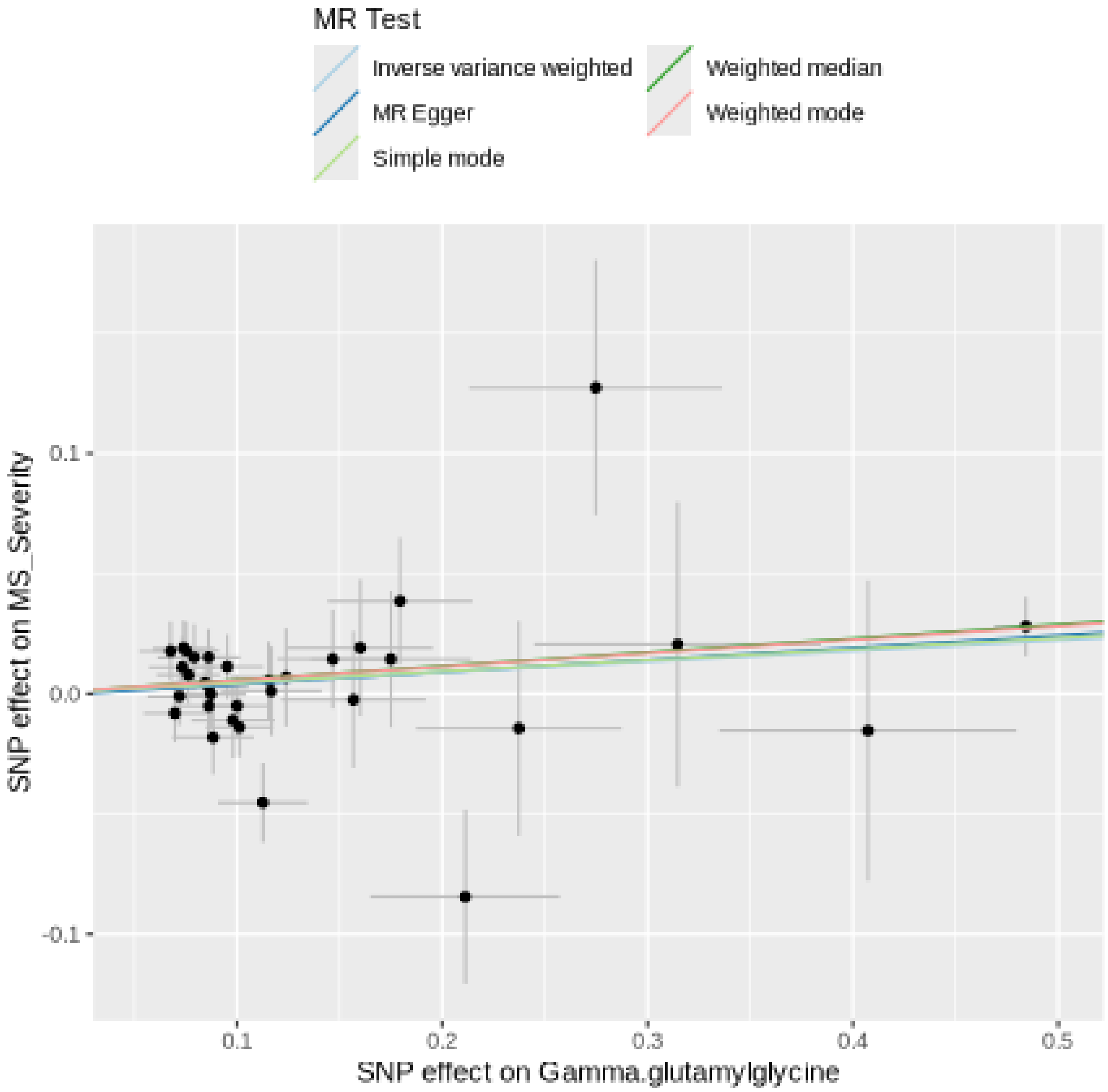

Funnel plot

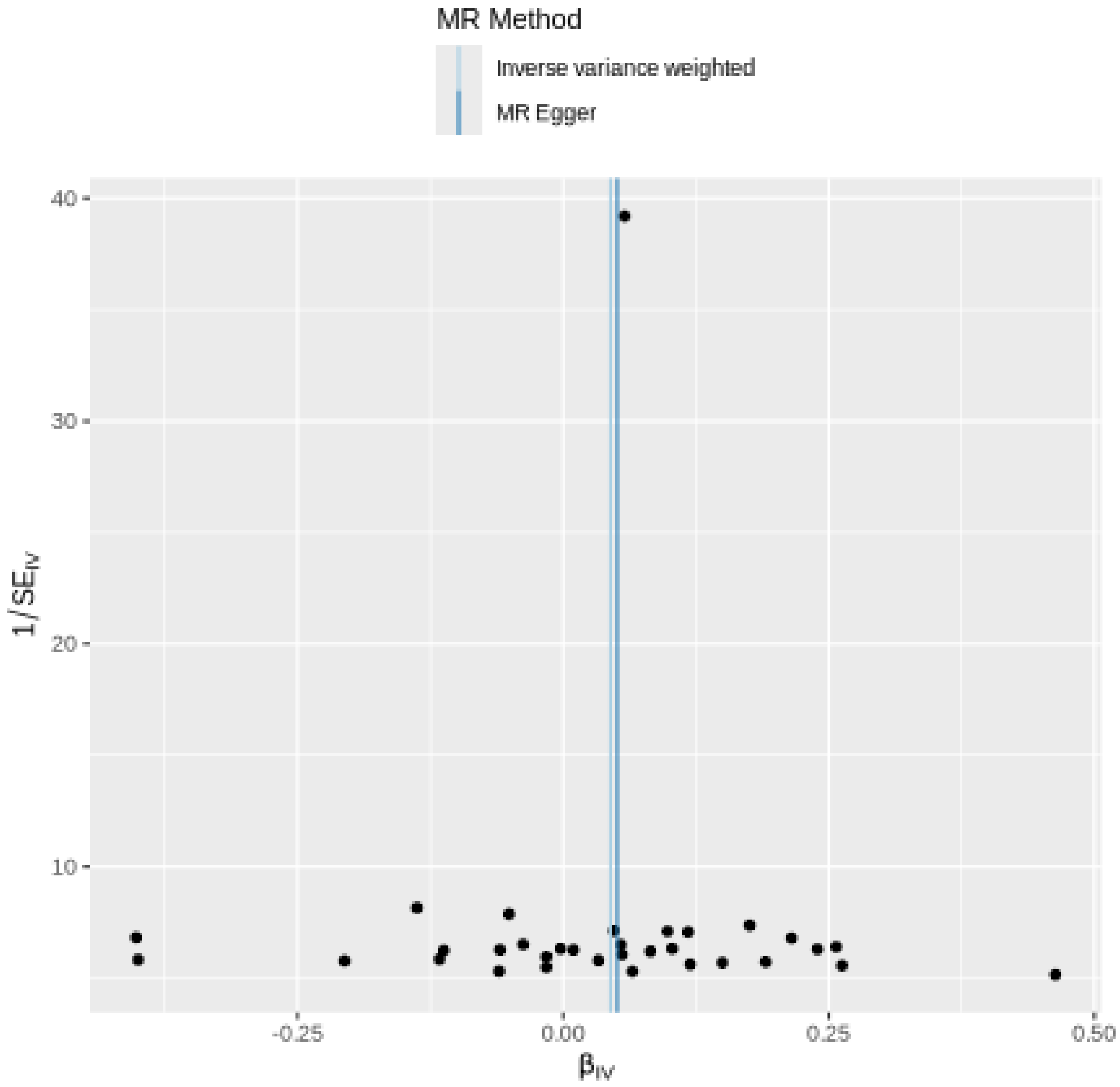

MR Method
Inverse variance weighted
MR Egger
40
30
20
10
1/SE$_{IV}$
-0.25
0.00
0.25
0.50
β$_{IV}$

Leave-one-out sensitivity analysis

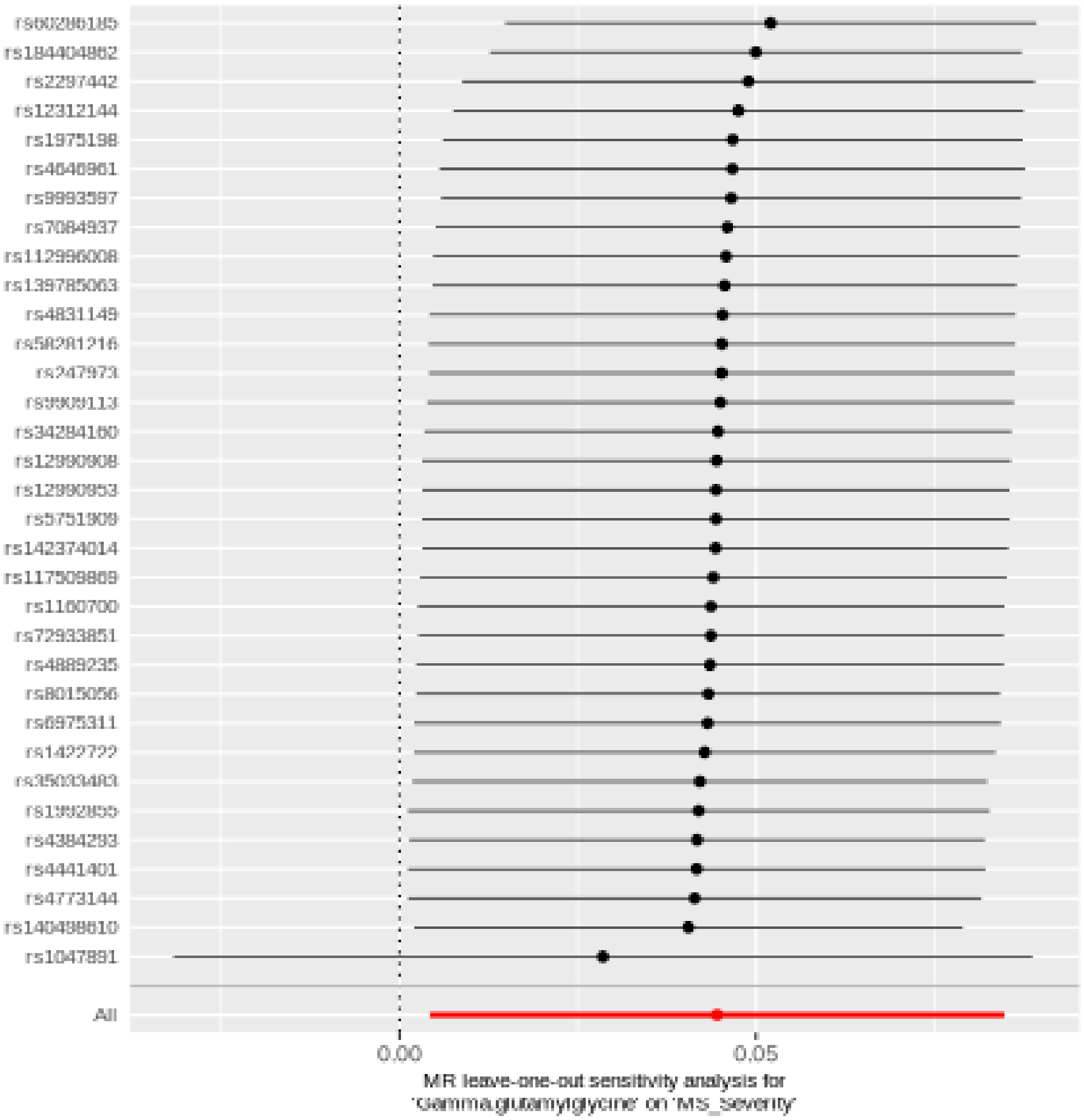

#### 1-ribosyl-imidazoleacetate

| method | nsnp | b | se | pval |
|---|---|---|---|---|
| MR Egger | 39 | 0.0433685 | 0.0662236 | 0.5165957 |
| Weighted median | 39 | -0.0643705 | 0.0396101 | 0.1041409 |
| Inverse variance weighted | 39 | -0.0681306 | 0.0250265 | 0.0064821 |
| Simple mode | 39 | -0.0523056 | 0.0834100 | 0.5343500 |
| Weighted mode | 39 | -0.0480538 | 0.0783730 | 0.5434348 |

Heterogeneity tests

| method | Q | Q_df | Q_pval |
|---|---|---|---|
| MR Egger | 33.16778 | 37 | 0.6493568 |
| Inverse variance weighted | 36.47484 | 38 | 0.5400344 |

Test for directional horizontal pleiotropy

| egger_intercept | se | pval |
|---|---|---|
| -0.0147327 | 0.0081014 | 0.0770848 |

Forest plot of single SNP MR

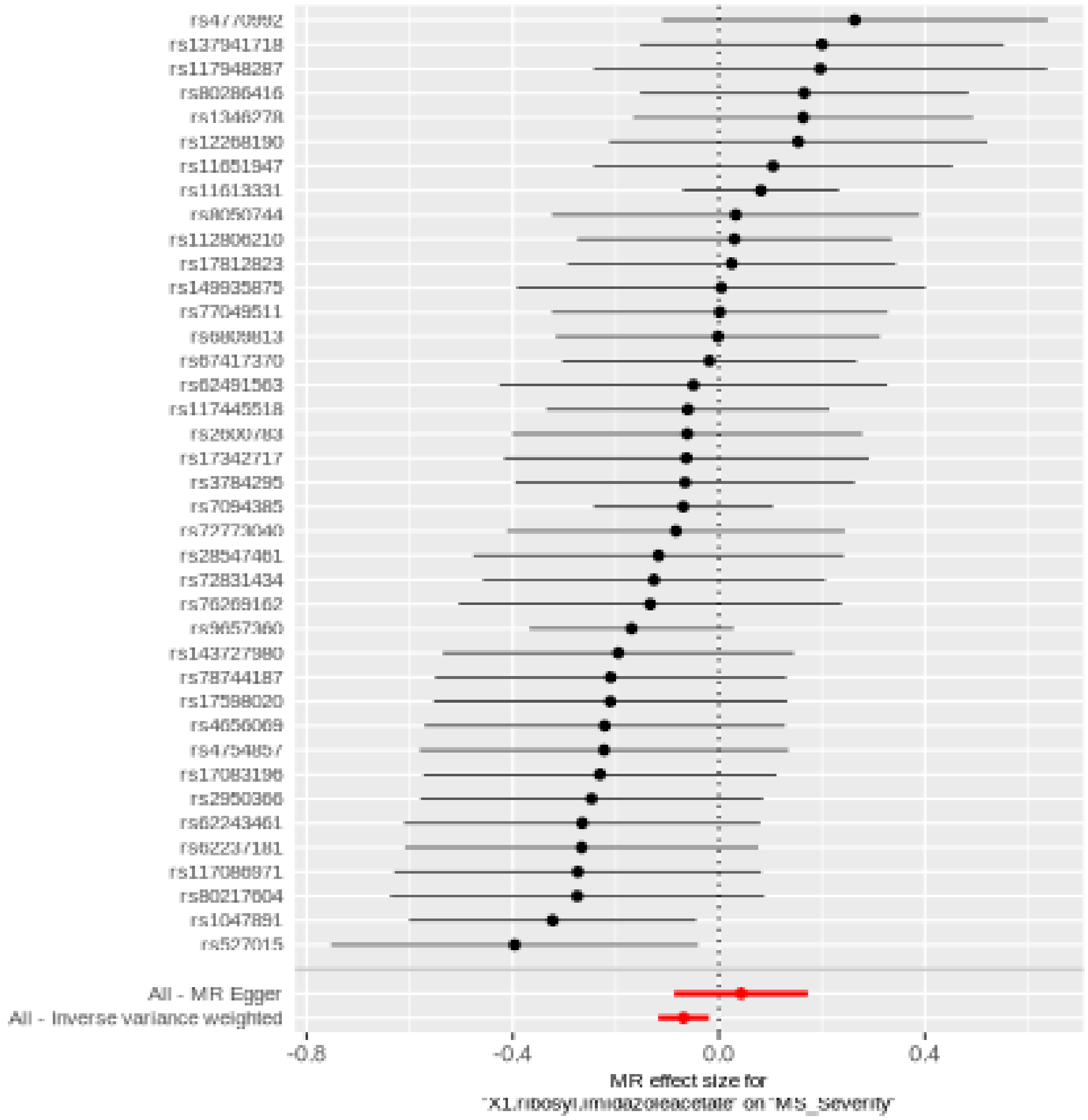

Comparison of results using different MR methods

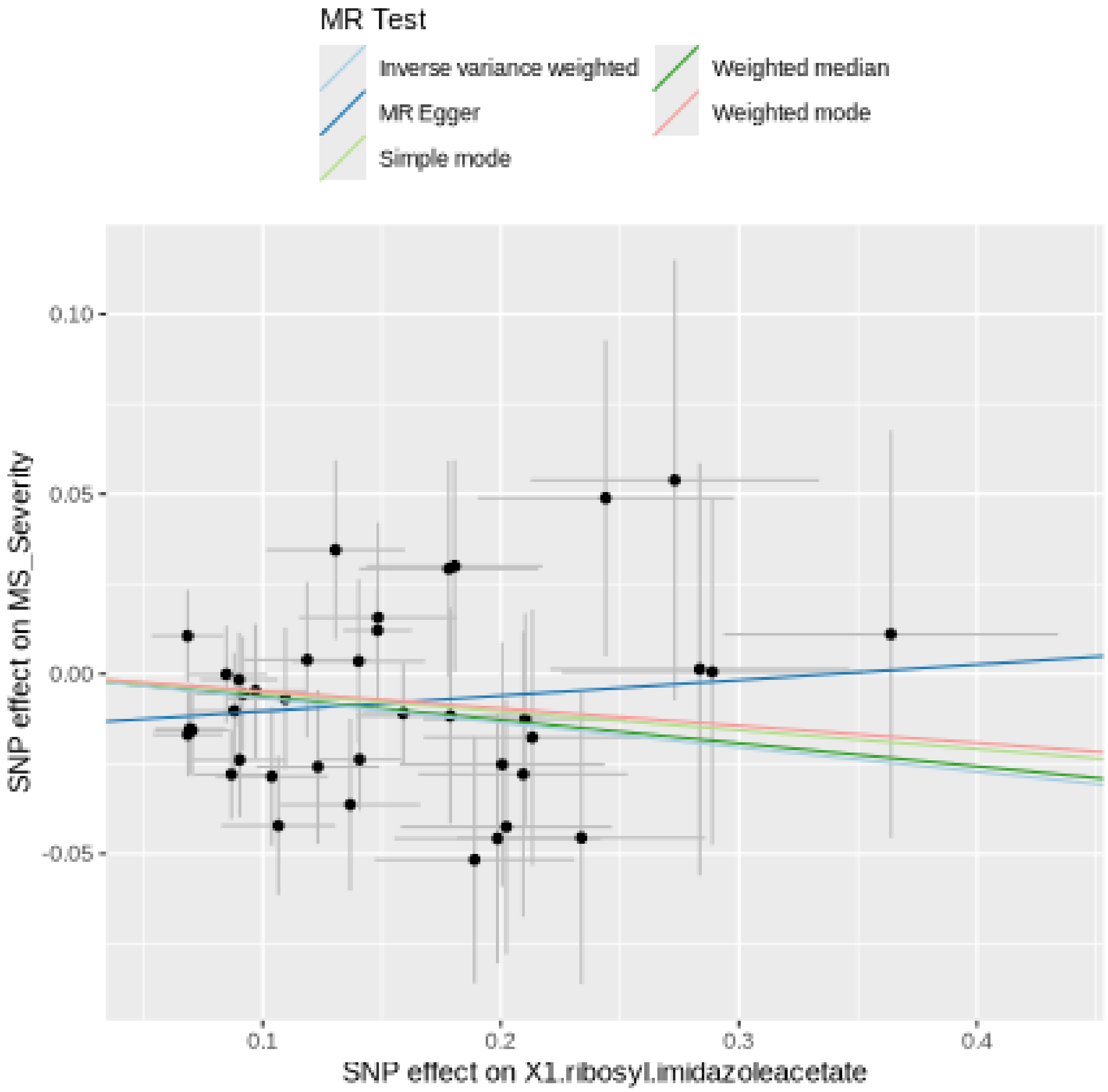

Funnel plot

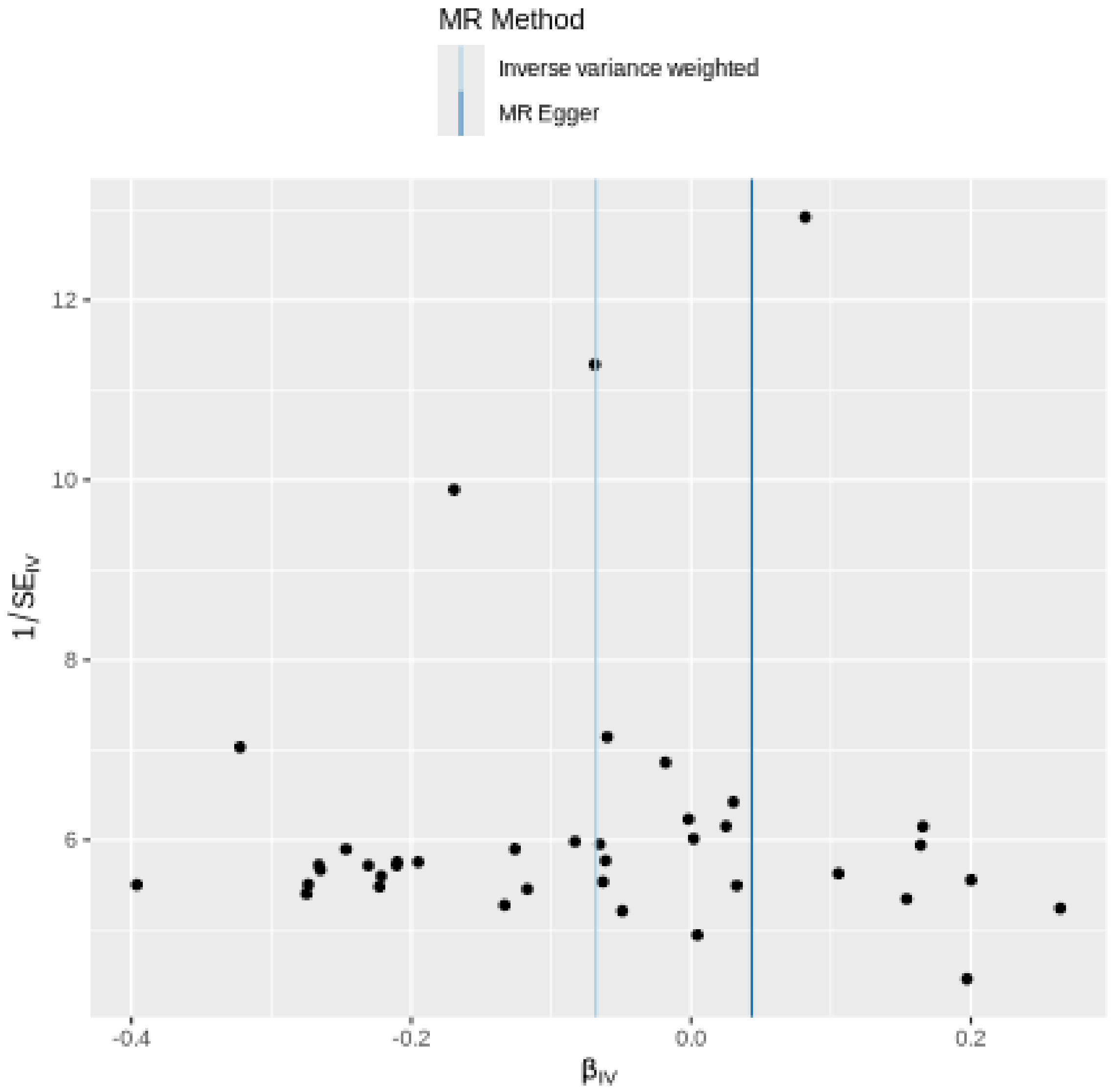
MR Method
Inverse variance weighted
MR Egger
12
10
8
6
1/SE$_{IV}$
-0.4
-0.2
0.0
0.2
β$_{IV}$

Leave-one-out sensitivity analysis

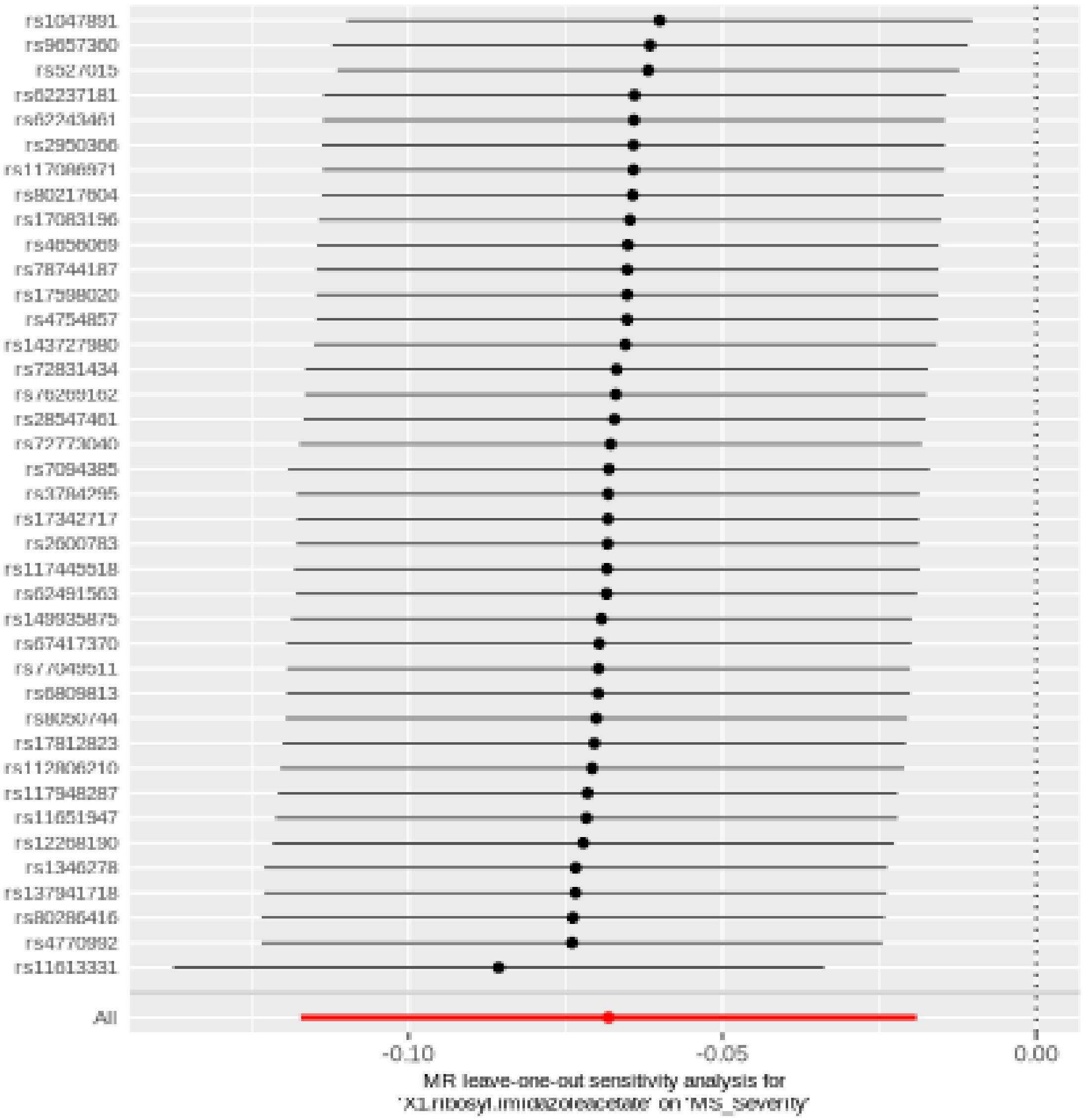

#### Thymol sulfate

| method | nsnp | b | se | pval |
|---|---|---|---|---|
| MR Egger | 19 | 0.1102418 | 0.0923631 | 0.2490371 |
| Weighted median | 19 | 0.0940129 | 0.0515810 | 0.0683598 |
| Inverse variance weighted | 19 | 0.0775236 | 0.0354636 | 0.0288150 |
| Simple mode | 19 | 0.1354168 | 0.0964456 | 0.1773158 |
| Weighted mode | 19 | 0.1435068 | 0.0782468 | 0.0832373 |

Heterogeneity tests

| method | Q | Q_df | Q_pval |
|---|---|---|---|
| MR Egger | 15.59934 | 17 | 0.5523944 |
| Inverse variance weighted | 15.74652 | 18 | 0.6102328 |

Test for directional horizontal pleiotropy

| egger_intercept | se | pval |
|---|---|---|
| -0.0041745 | 0.0108813 | 0.7060008 |

Forest plot of single SNP MR

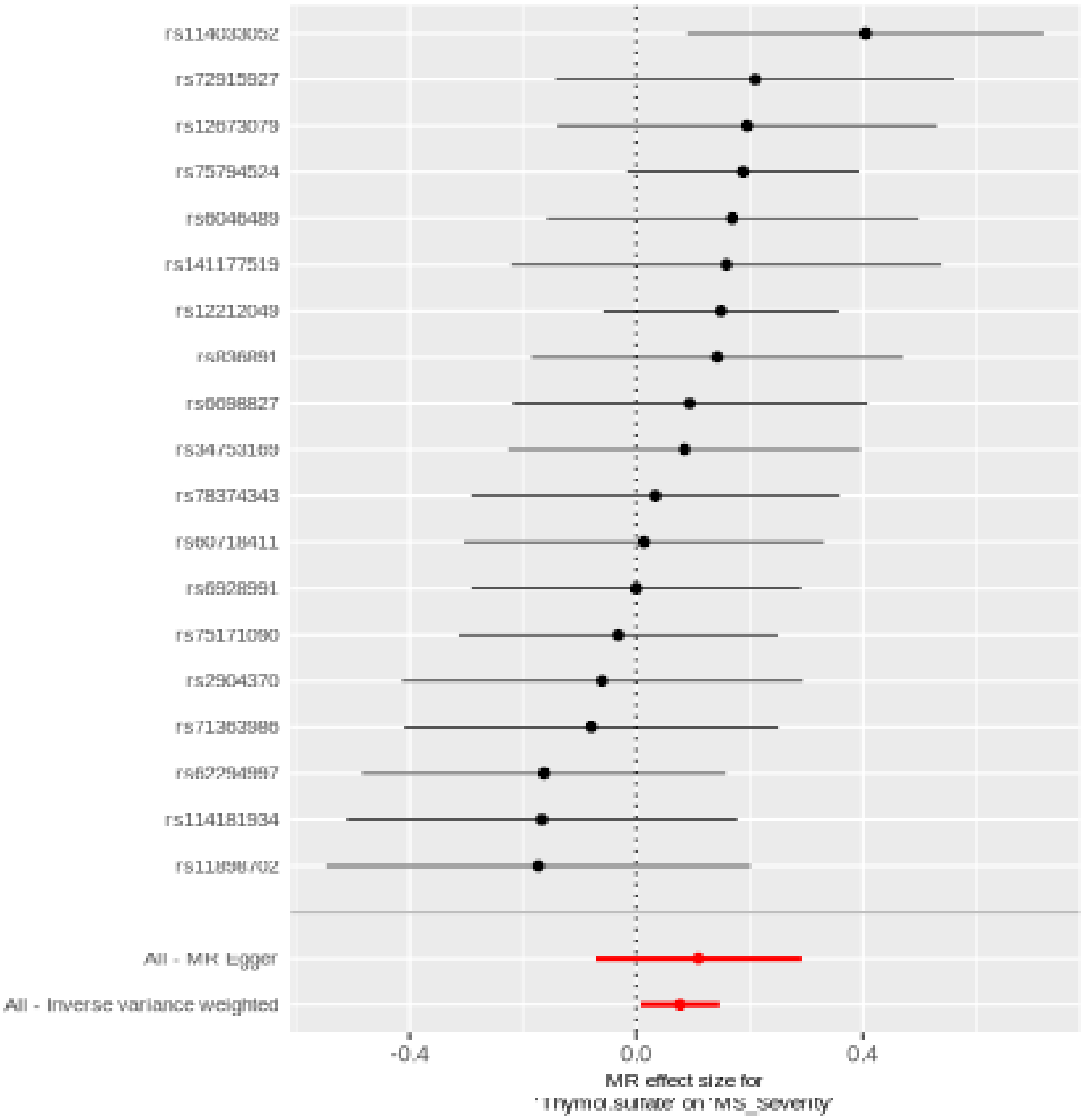

Comparison of results using different MR methods

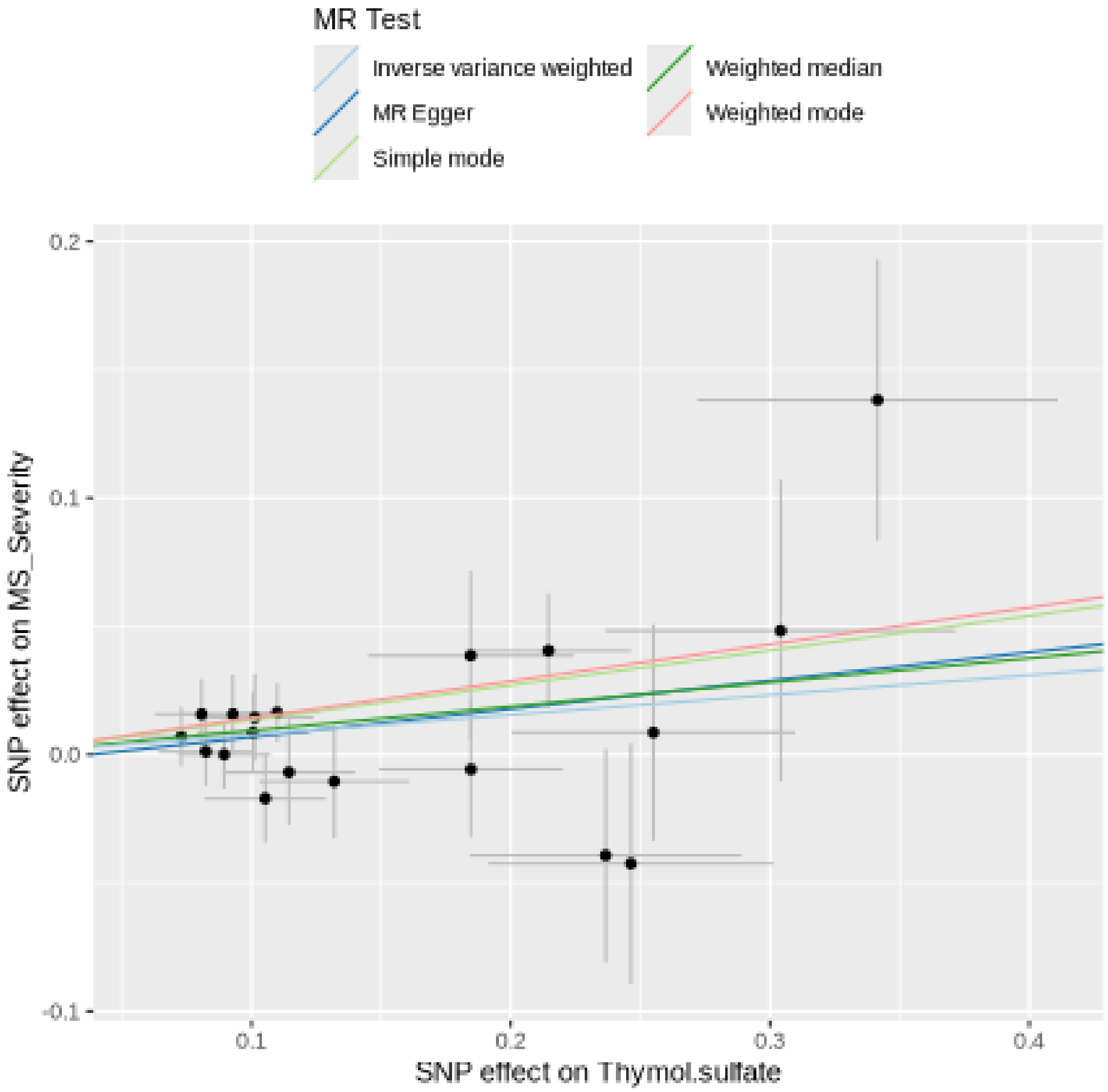

Funnel plot

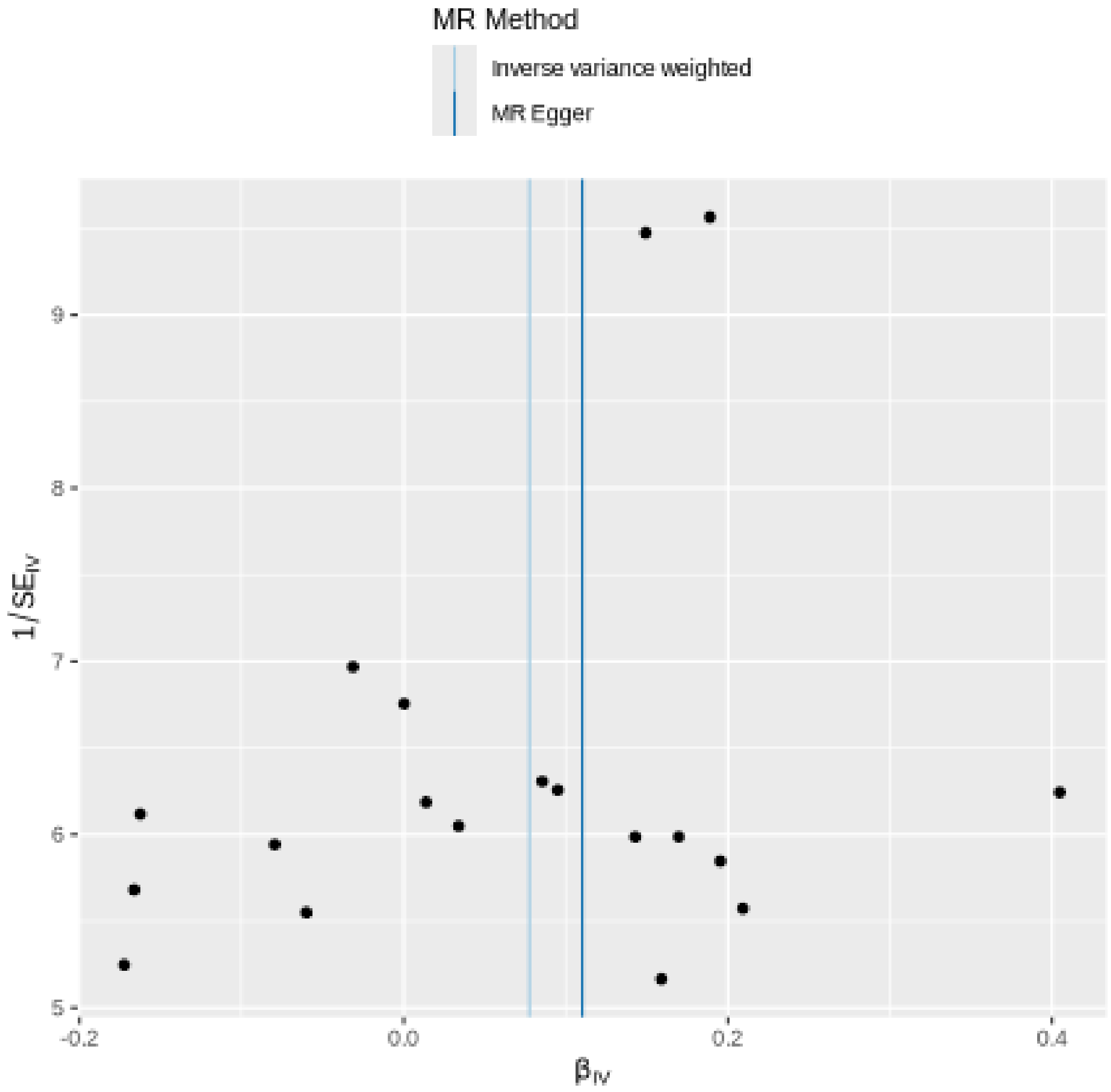

MR Method
Inverse variance weighted
MR Egger
9
8
7
6
5
-0.2
0.0
0.2
0.4
1/SE_IV
β_IV

Leave-one-out sensitivity analysis

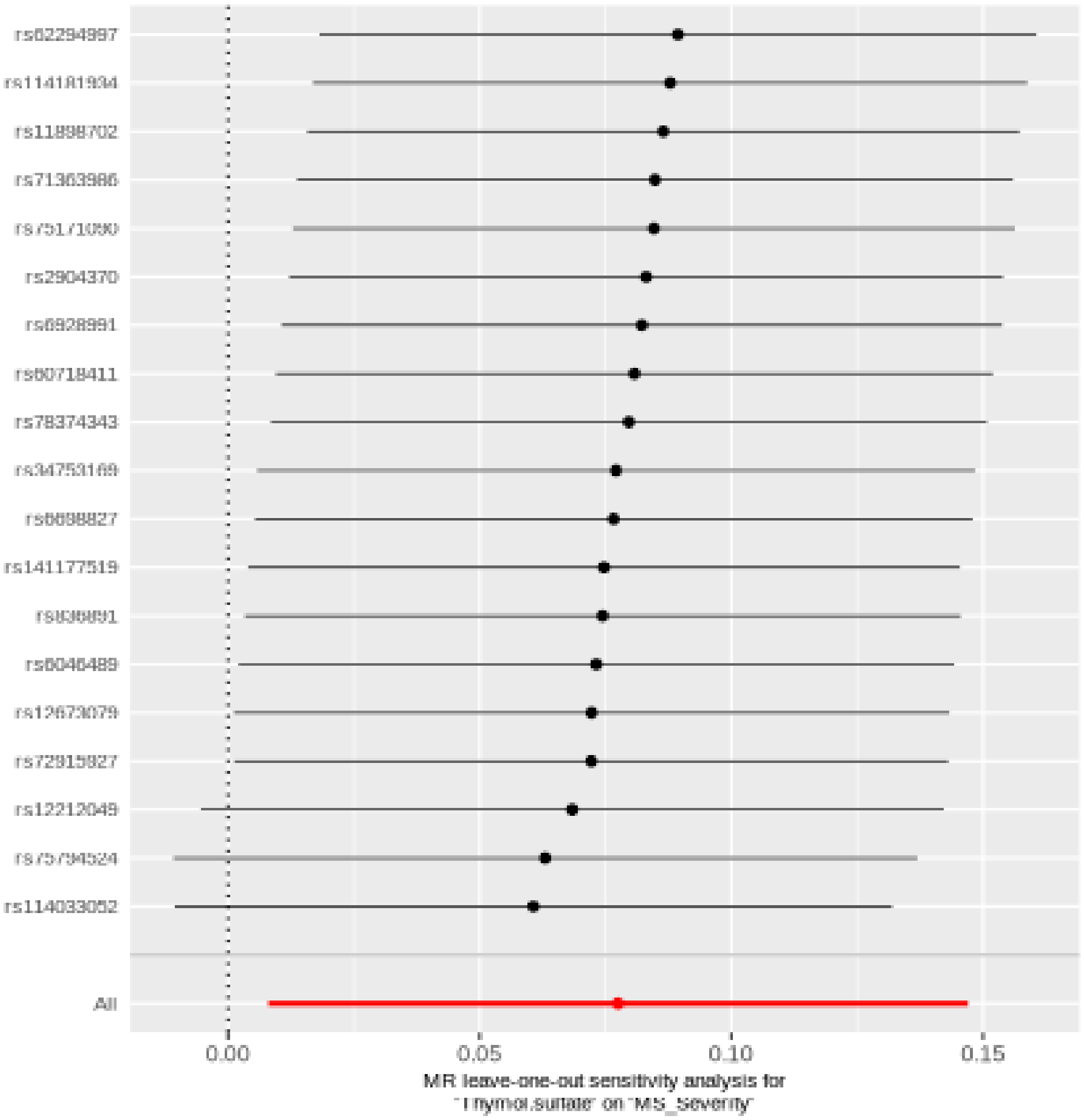

#### Glycocholenate sulfate

| method | nsnp | b | se | pval |
|---|---|---|---|---|
| MR Egger | 41 | 0.0348031 | 0.0249540 | 0.1710040 |
| Weighted median | 41 | 0.0499576 | 0.0219636 | 0.0229319 |
| Inverse variance weighted | 41 | 0.0449603 | 0.0157900 | 0.0044080 |
| Simple mode | 41 | 0.0271456 | 0.0472718 | 0.5690168 |
| Weighted mode | 41 | 0.0467802 | 0.0210311 | 0.0318347 |

##### Heterogeneity tests

| method | Q | Q_df | Q_pval |
|---|---|---|---|
| MR Egger | 38.54084 | 39 | 0.4906437 |
| Inverse variance weighted | 38.81716 | 40 | 0.5234304 |

##### Test for directional horizontal pleiotropy

| egger_intercept | se | pval |
|---|---|---|
| 0.0025169 | 0.0047882 | 0.6021032 |

Forest plot of single SNP MR

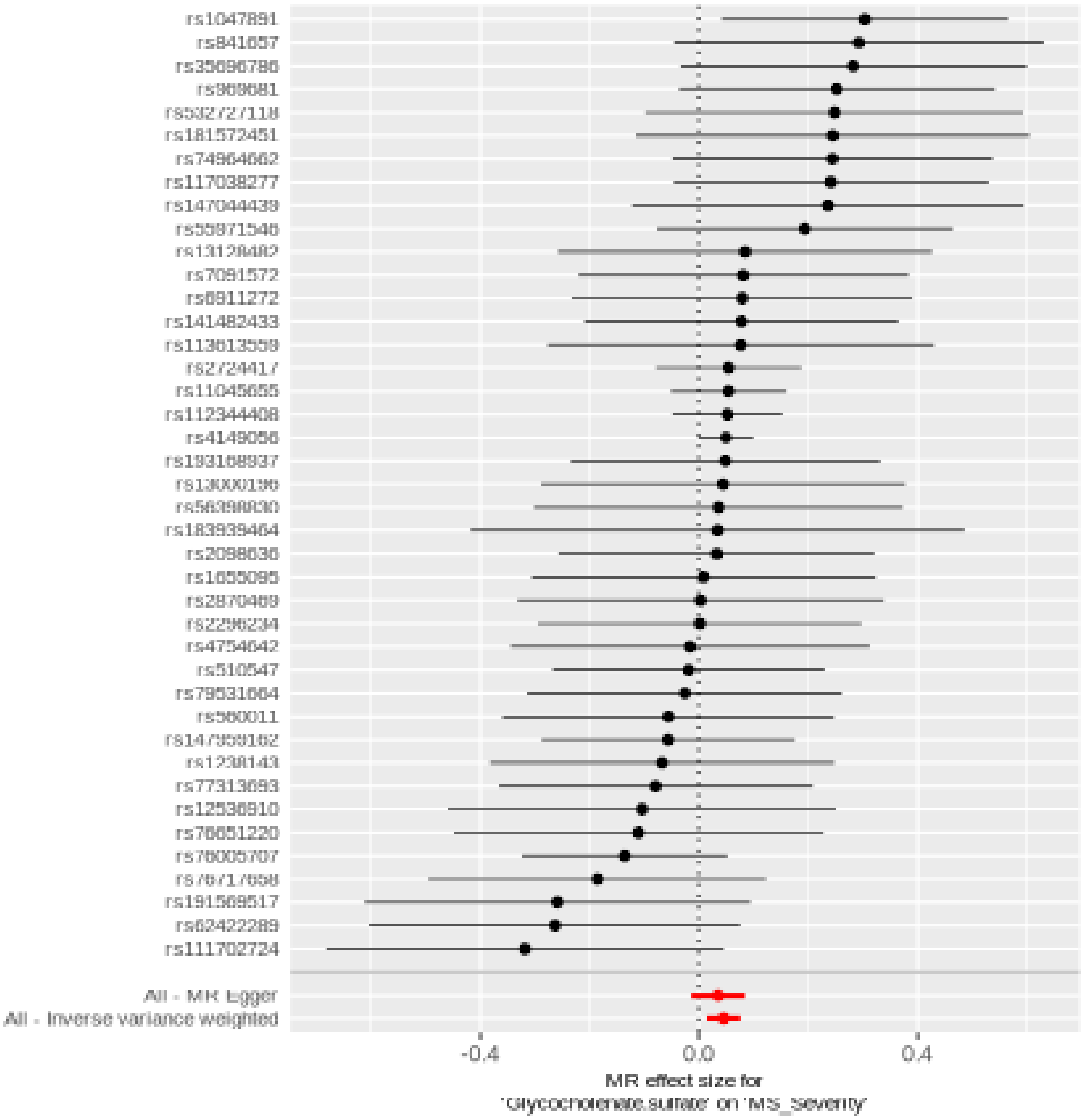

Comparison of results using different MR methods

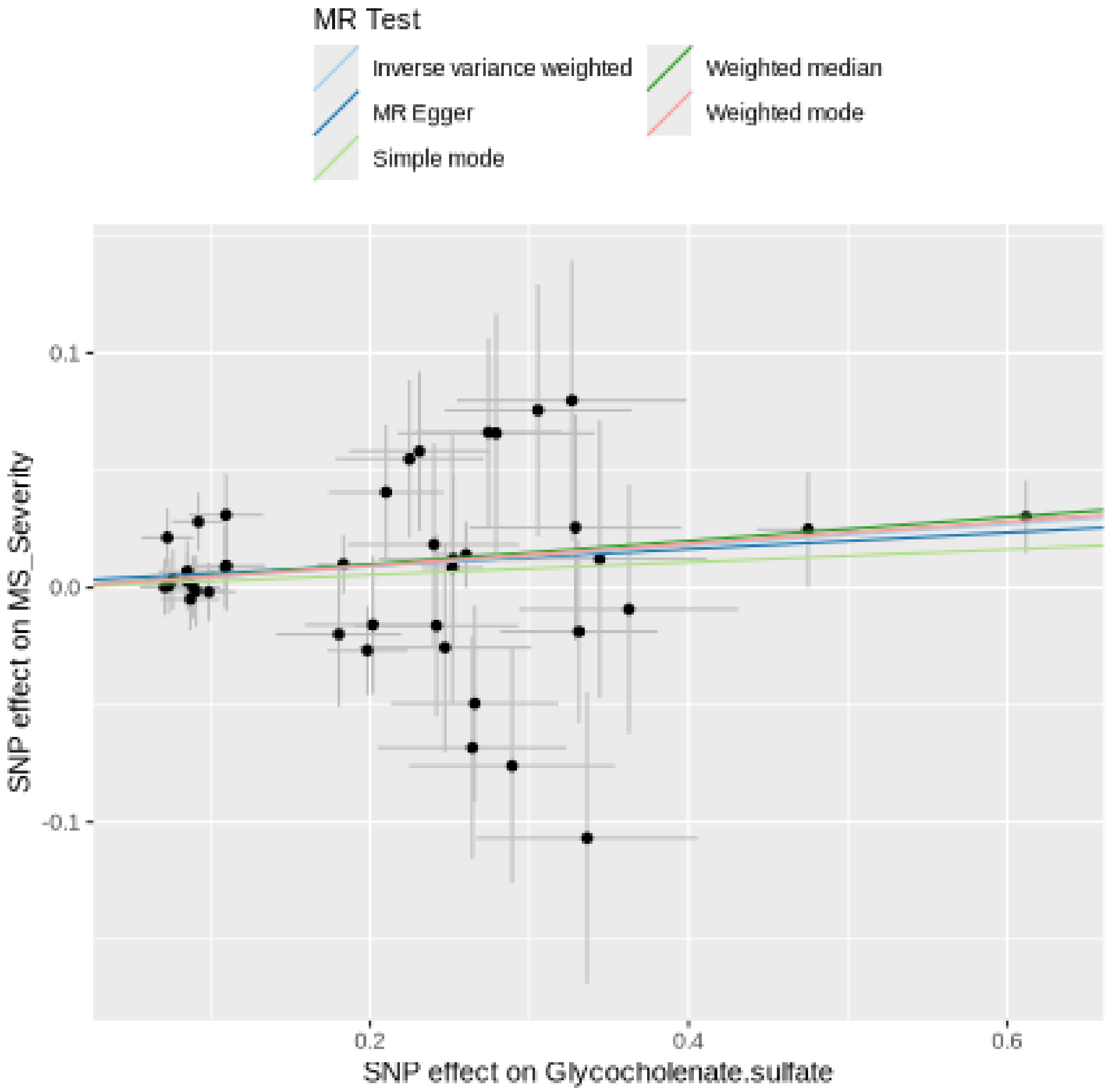

Funnel plot

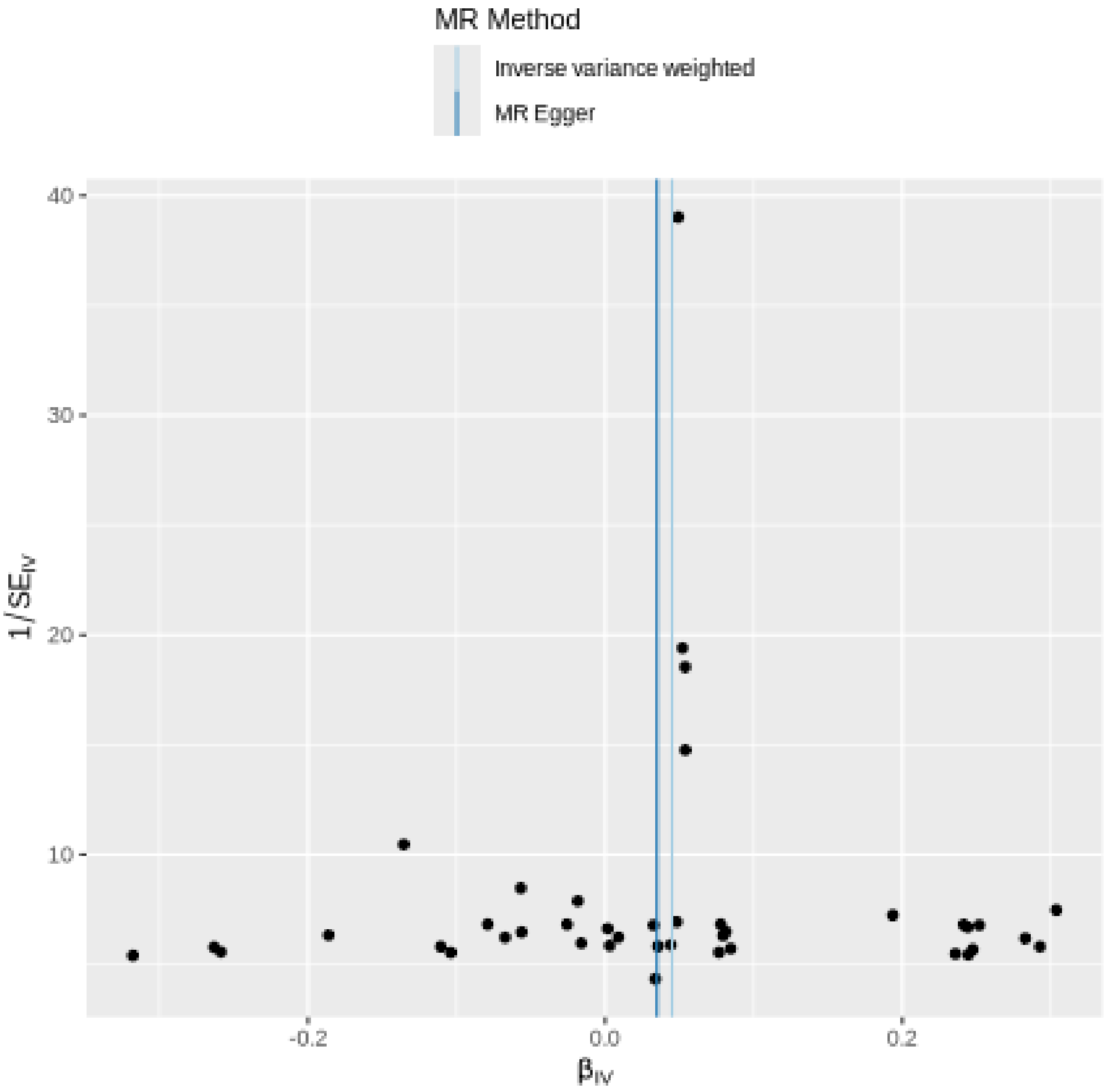

MR Method
Inverse variance weighted
MR Egger
40
30
20
10
1/SE_IV
-0.2
0.0
0.2
β_IV

#### Leave-one-out sensitivity analysis

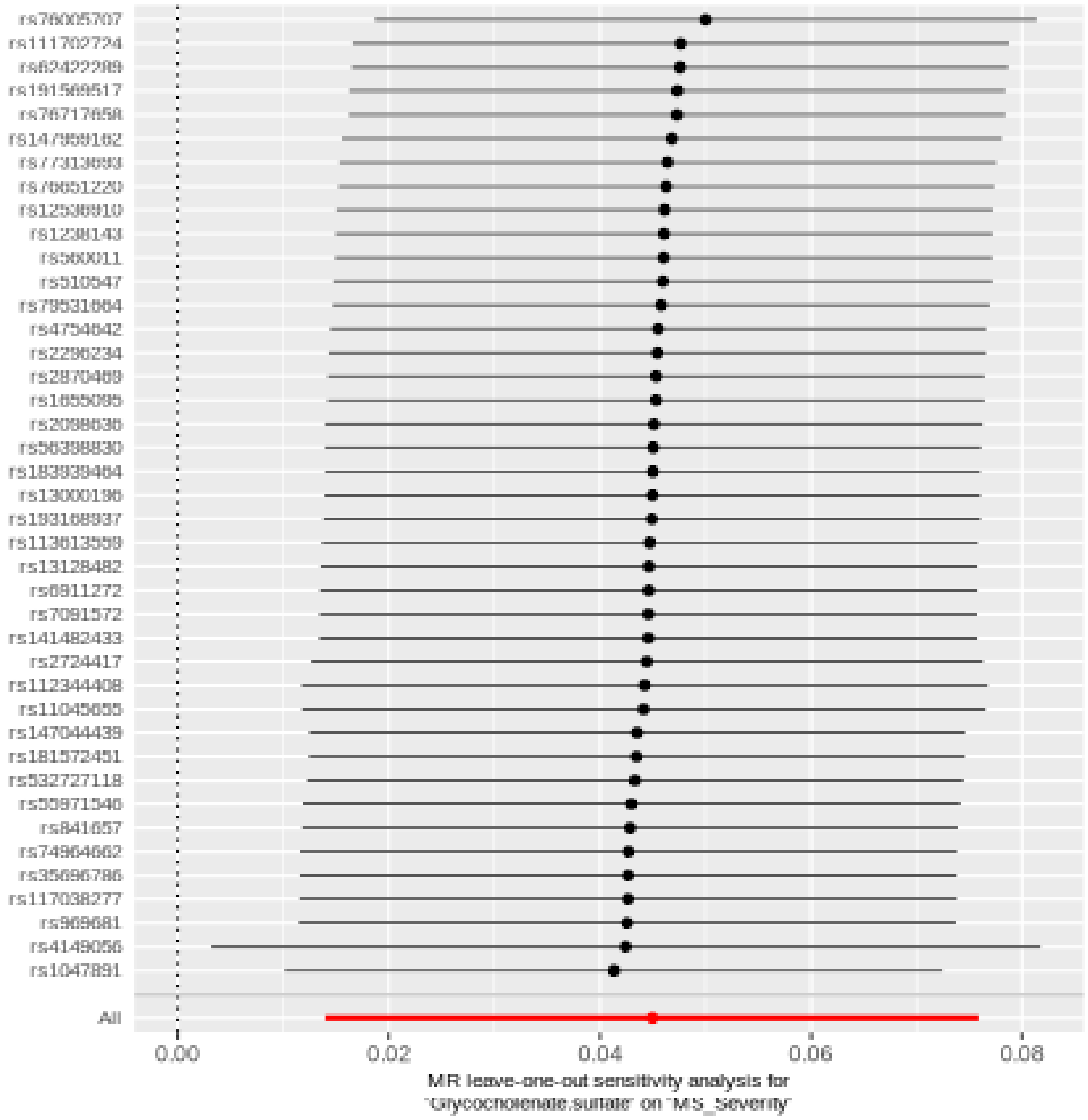

#### 1-palmitoyl-2-linoleoyl-GPE (16:0/18:2)

| method | nsnp | b | se | pval |
|---|---|---|---|---|
| MR Egger | 29 | 0.0395585 | 0.0293574 | 0.1890260 |
| Weighted median | 29 | 0.0612149 | 0.0234489 | 0.0090394 |
| Inverse variance weighted | 29 | 0.0408343 | 0.0169460 | 0.0159668 |
| Simple mode | 29 | 0.0554214 | 0.0525108 | 0.3002517 |
| Weighted mode | 29 | 0.0554214 | 0.0219576 | 0.0175538 |

##### Heterogeneity tests

| method | Q | Q_df | Q_pval |
|---|---|---|---|
| MR Egger | 30.27271 | 27 | 0.3020405 |
| Inverse variance weighted | 30.27595 | 28 | 0.3501497 |

##### Test for directional horizontal pleiotropy

| egger_intercept | se | pval |
|---|---|---|
| 0.0002935 | 0.0054639 | 0.9575549 |

Forest plot of single SNP MR

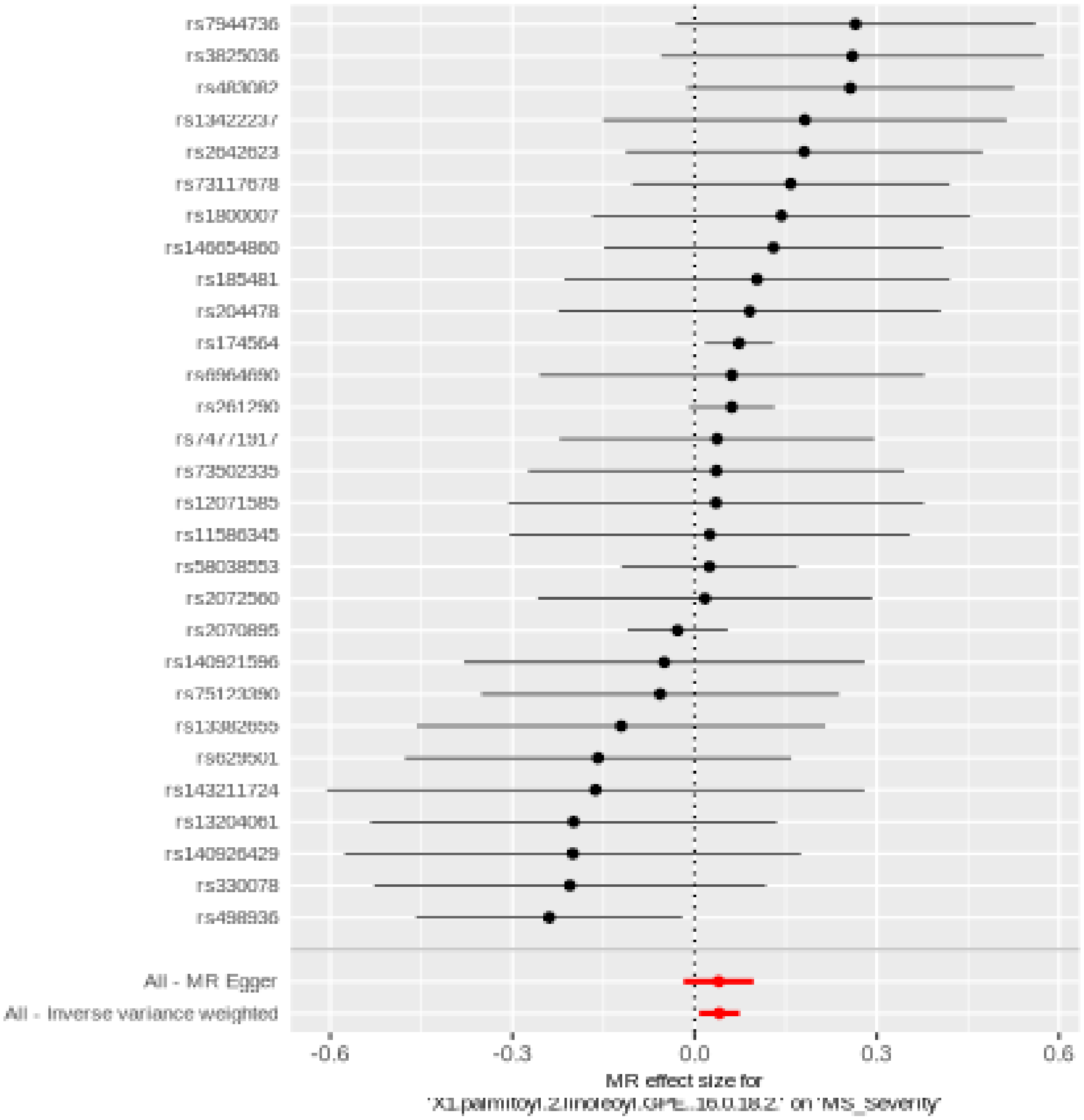

Comparison of results using different MR methods

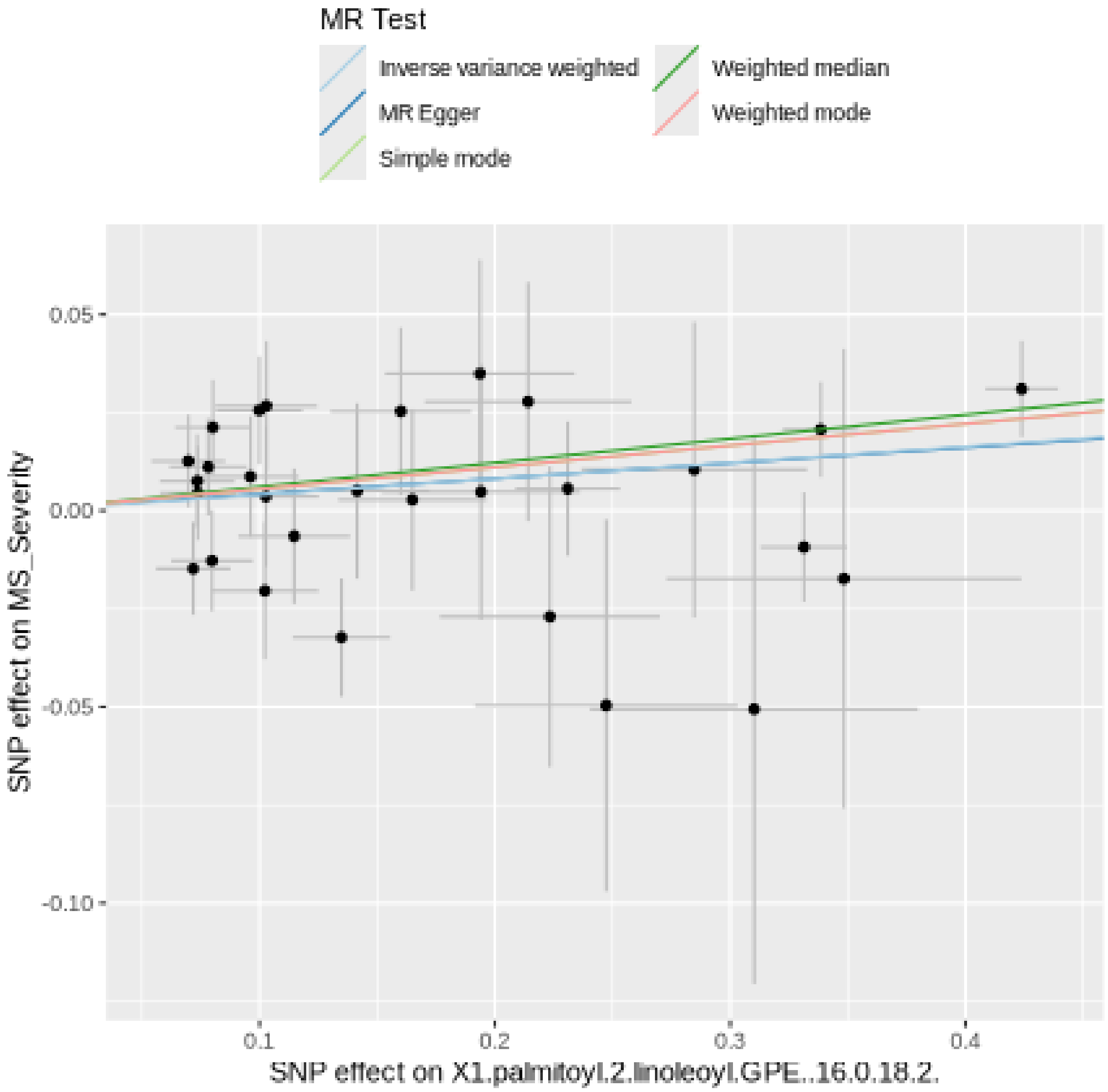

Funnel plot

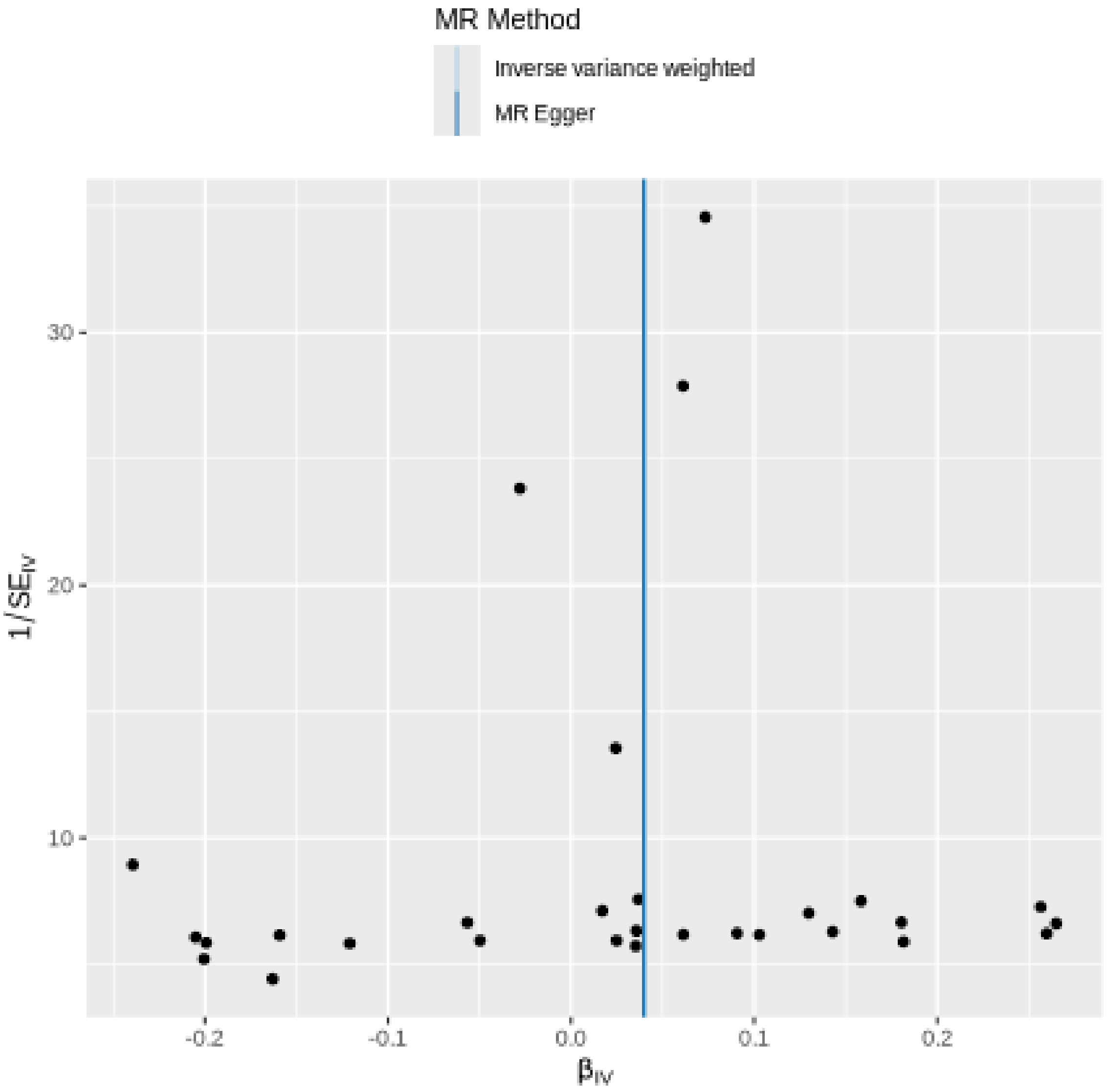
MR Method
Inverse variance weighted
MR Egger
30
20
10
1/SE_IV
-0.2
-0.1
0.0
0.1
0.2
β_IV

### Leave-one-out sensitivity analysis

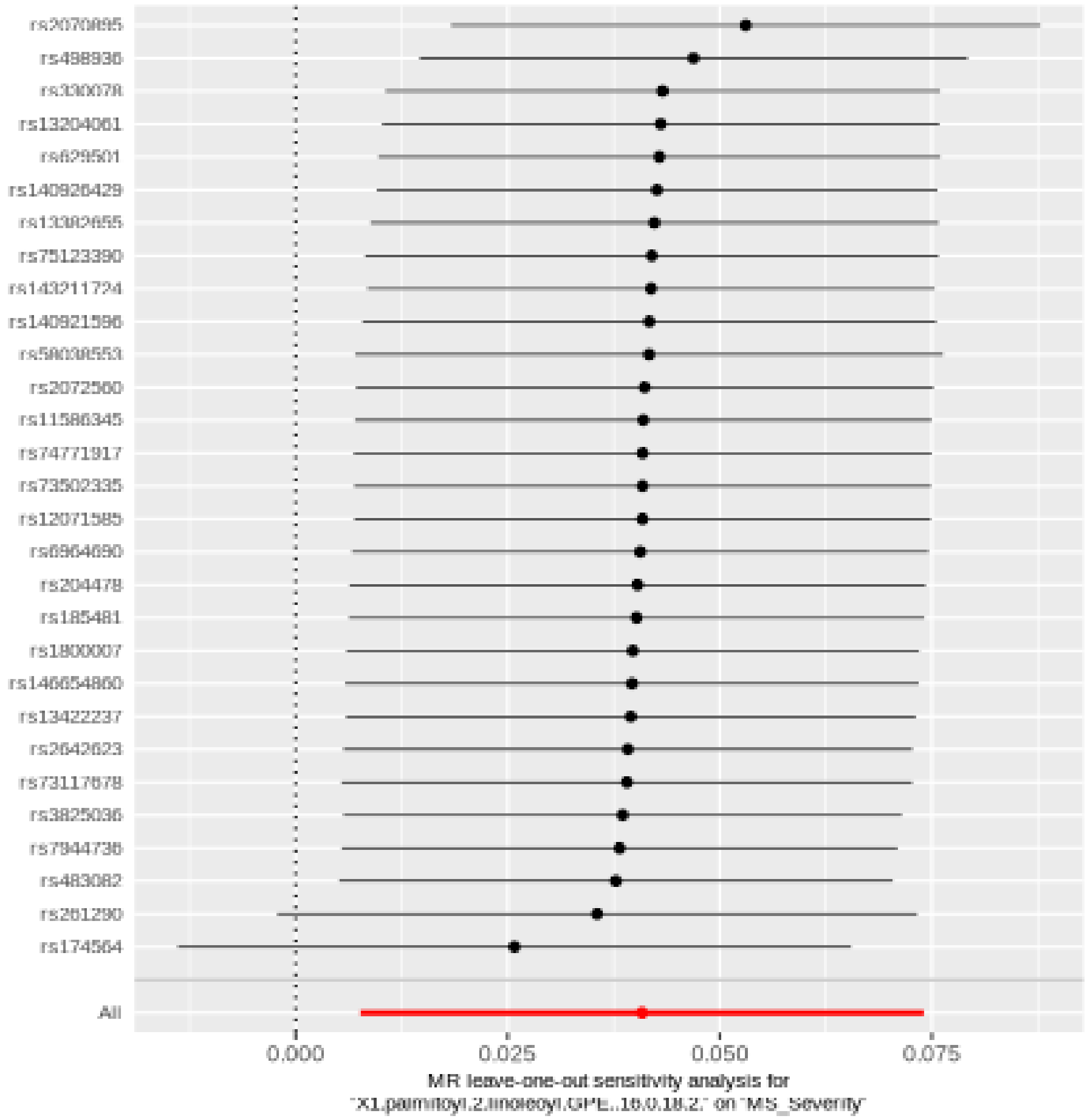

#### N-formylanthranilic acid

| method | nsnp | b | se | pval |
|---|---|---|---|---|
| MR Egger | 29 | -0.0162362 | 0.0286349 | 0.5753931 |
| Weighted median | 29 | -0.0278795 | 0.0253964 | 0.2723035 |
| Inverse variance weighted | 29 | -0.0481326 | 0.0192977 | 0.0126234 |
| Simple mode | 29 | -0.0204041 | 0.0591810 | 0.7328405 |
| Weighted mode | 29 | -0.0252399 | 0.0251787 | 0.3247189 |

##### Heterogeneity tests

| method | Q | Q_df | Q_pval |
|---|---|---|---|
| MR Egger | 20.22516 | 27 | 0.8211754 |
| Inverse variance weighted | 22.49834 | 28 | 0.7577150 |

##### Test for directional horizontal pleiotropy

| egger_intercept | se | pval |
|---|---|---|
| -0.008102 | 0.0053737 | 0.1432425 |

### Forest plot of single SNP MR

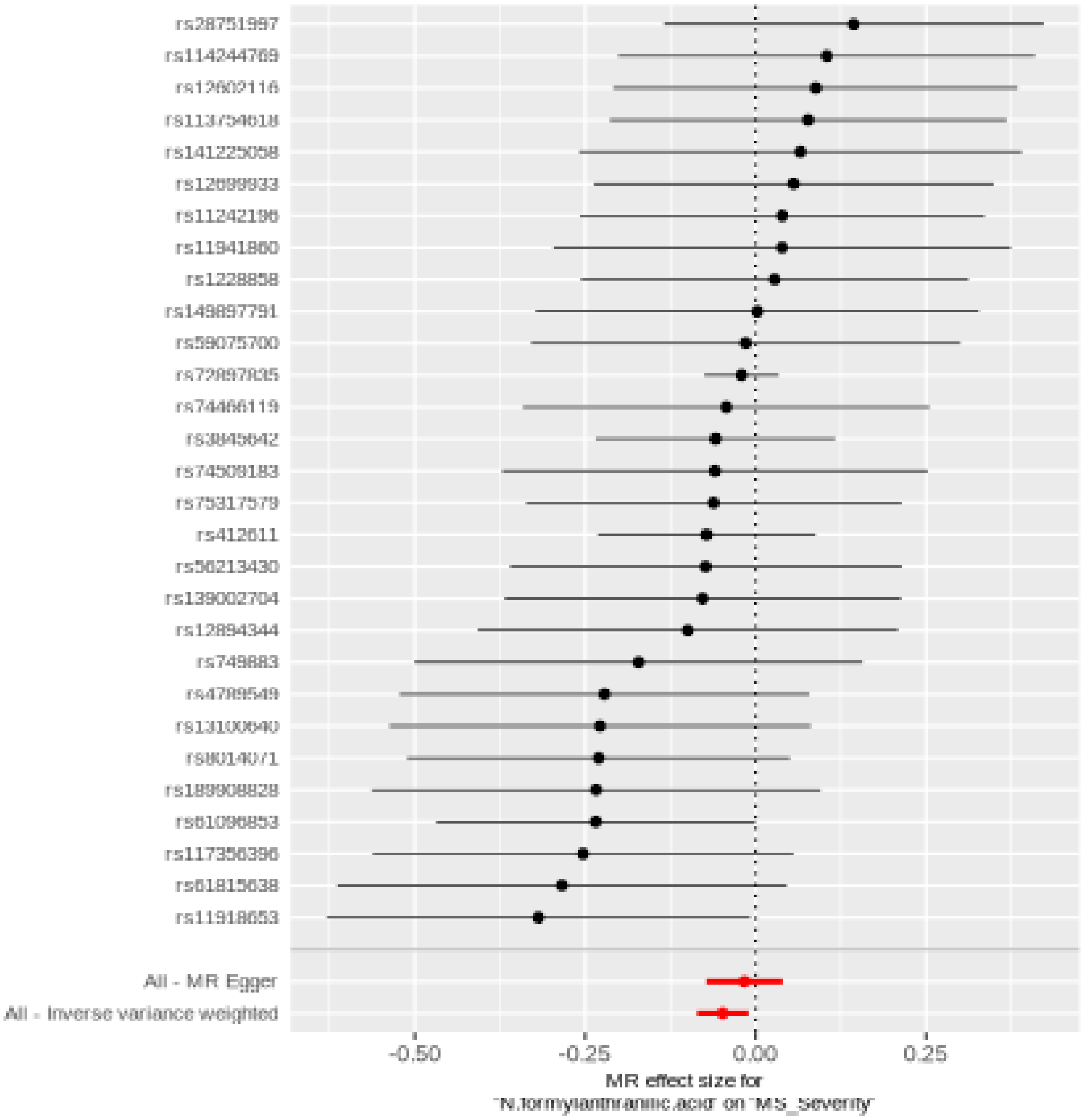

Comparison of results using different MR methods

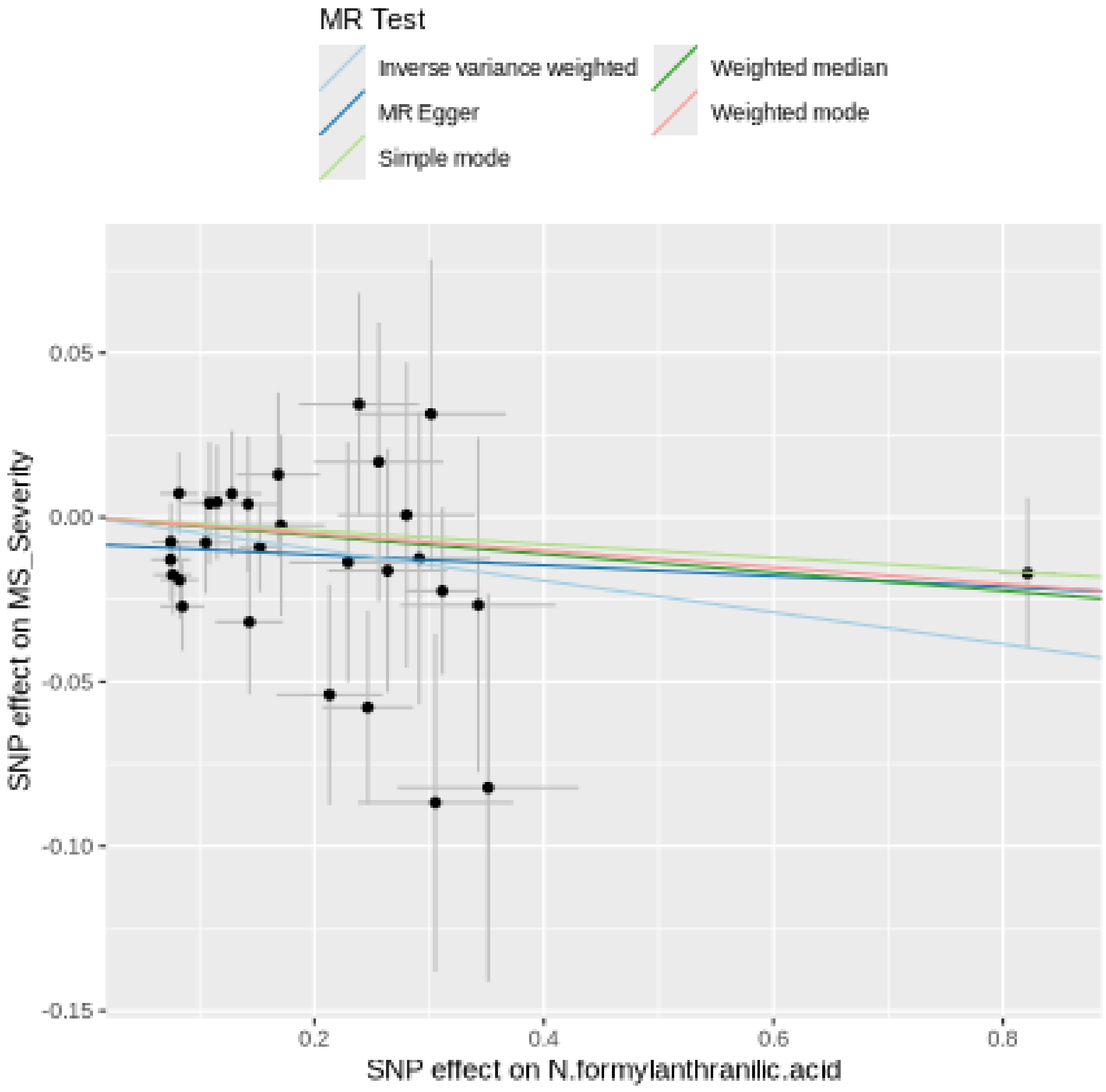

Funnel plot

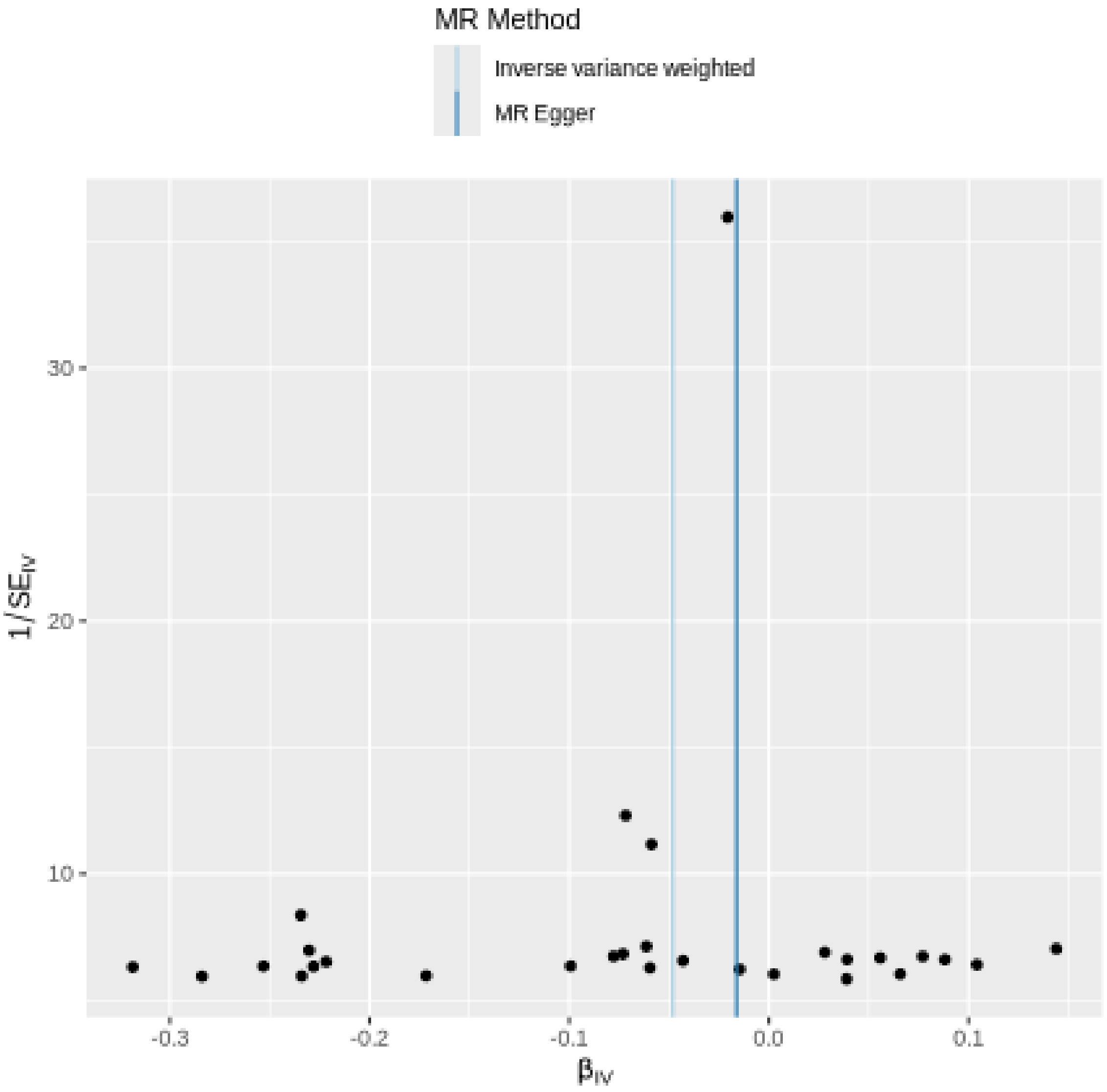

MR Method
Inverse variance weighted
MR Egger
30
20
10
1/SE$_{IV}$
-0.3
-0.2
-0.1
0.0
0.1
β$_{IV}$

### Leave-one-out sensitivity analysis

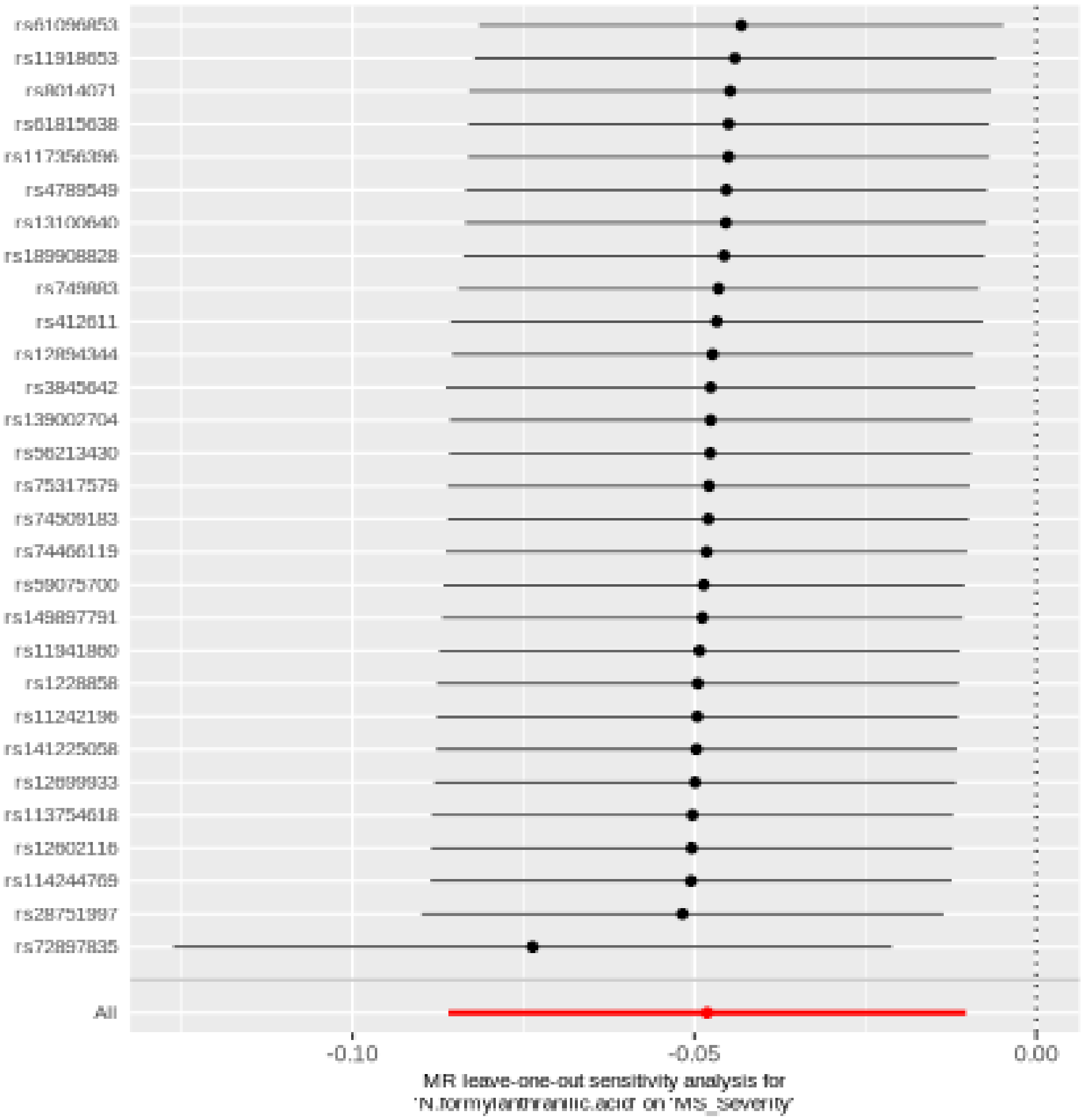

#### 1,2-dilinoleoyl-GPC (18:2/18:2)

| method | nsnp | b | se | pval |
|---|---|---|---|---|
| MR Egger | 20 | 0.0875569 | 0.0512083 | 0.1044818 |
| Weighted median | 20 | 0.1021079 | 0.0394295 | 0.0096078 |
| Inverse variance weighted | 20 | 0.0592622 | 0.0283517 | 0.0365950 |
| Simple mode | 20 | -0.0265724 | 0.0829520 | 0.7522113 |
| Weighted mode | 20 | 0.1130386 | 0.0423128 | 0.0150877 |

##### Heterogeneity tests

| method | Q | Q_df | Q_pval |
|---|---|---|---|
| MR Egger | 14.30934 | 18 | 0.7087147 |
| Inverse variance weighted | 14.74959 | 19 | 0.7383745 |

##### Test for directional horizontal pleiotropy

| egger_intercept | se | pval |
|---|---|---|
| -0.0042459 | 0.0063991 | 0.5154098 |

Forest plot of single SNP MR

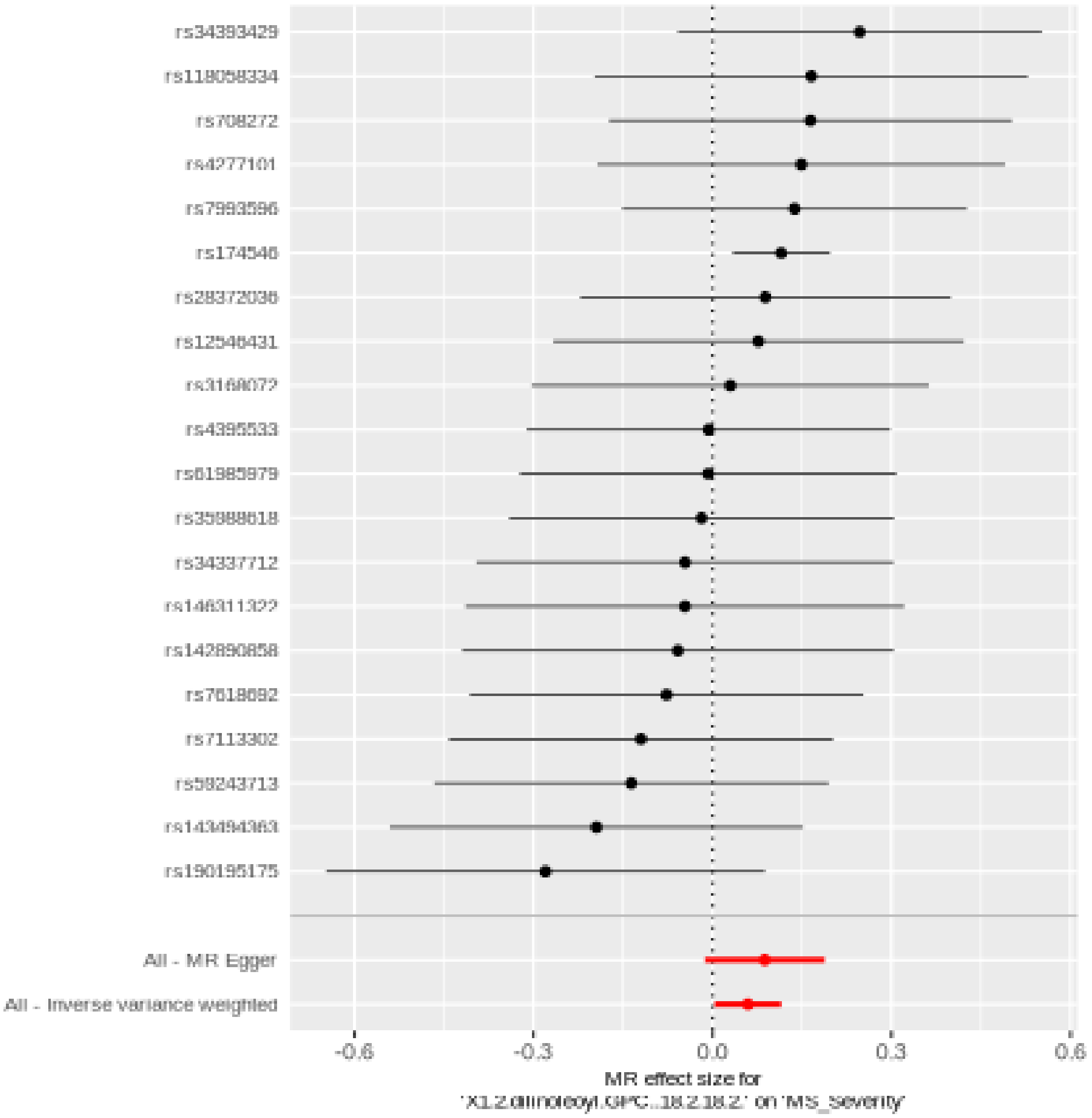

Comparison of results using different MR methods

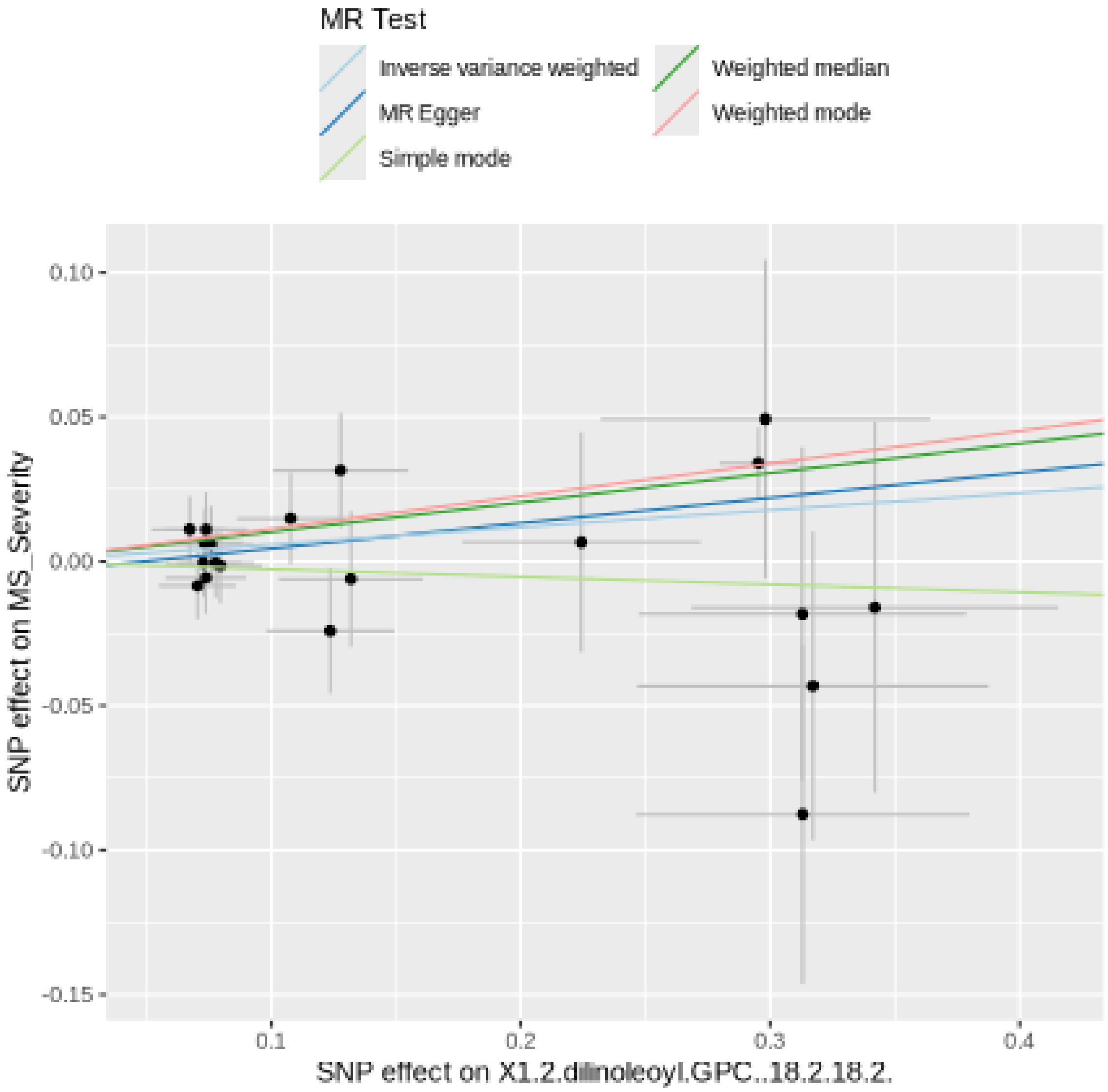

Funnel plot

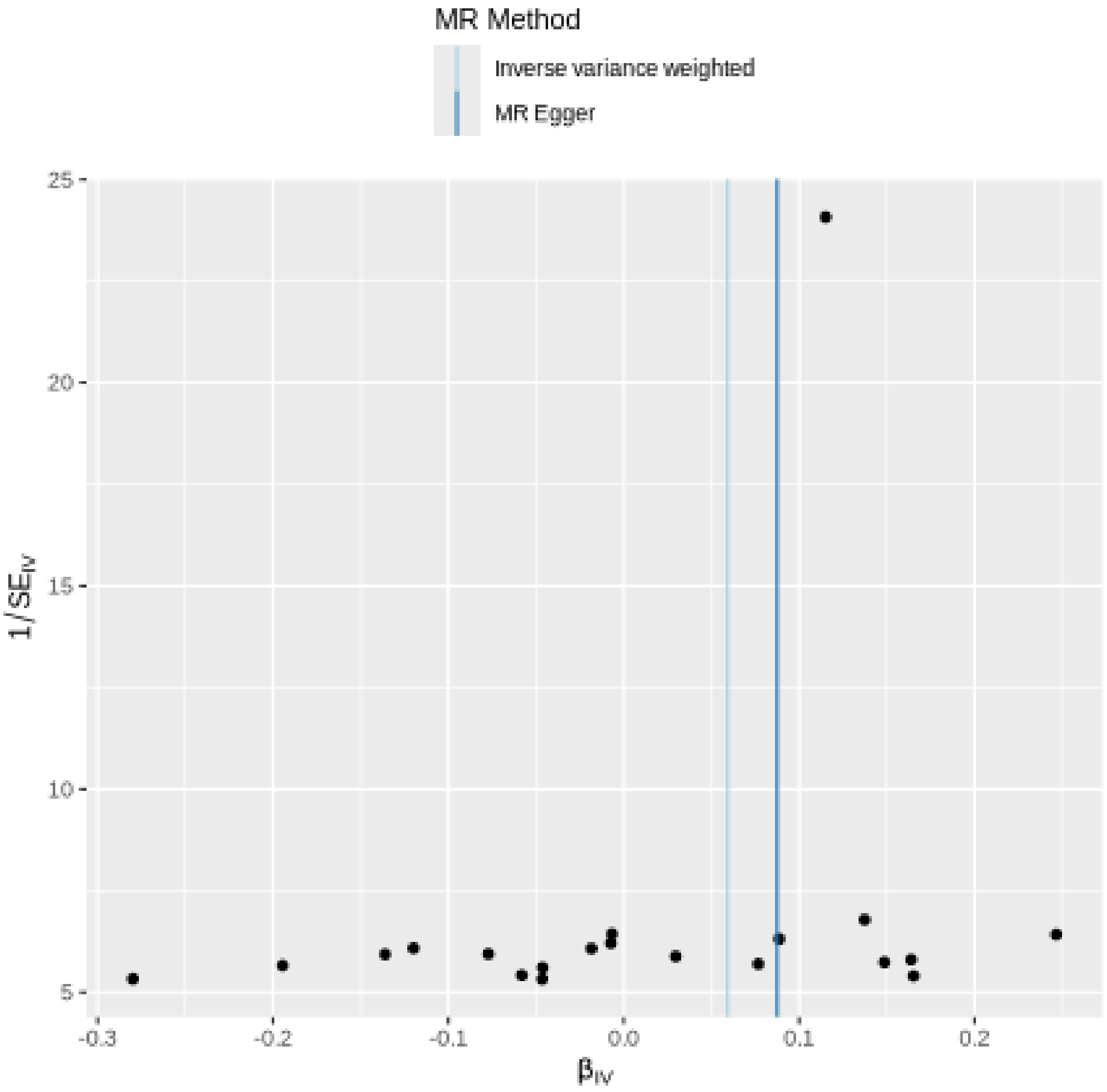

MR Method
Inverse variance weighted
MR Egger
25
20
15
10
5
$1/SE_{IV}$
-0.3
-0.2
-0.1
0.0
0.1
0.2
$\beta_{IV}$

#### Leave-one-out sensitivity analysis

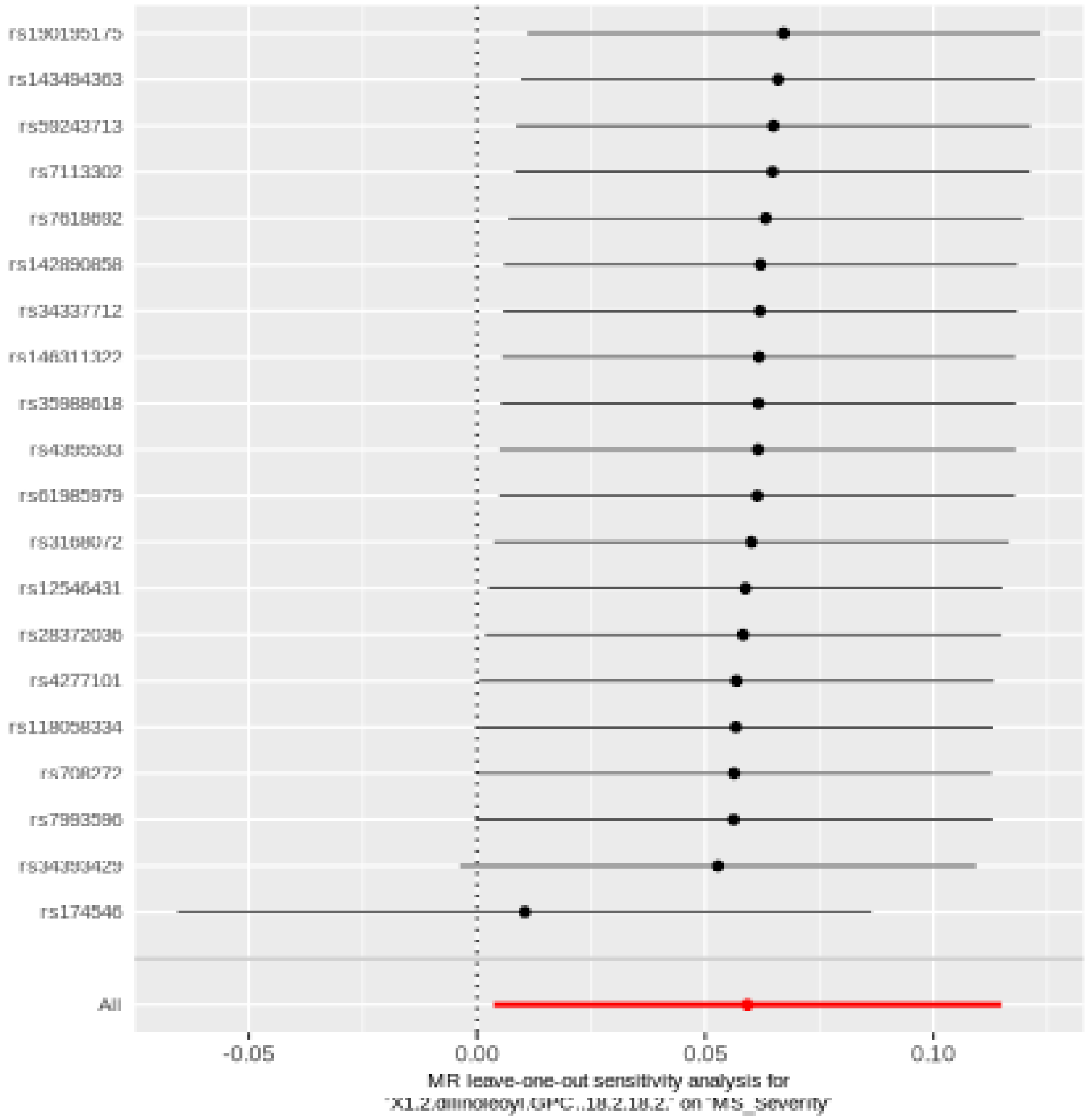

#### 1-stearoyl-2-linoleoyl-GPE (18:0/18:2)

| method | nsnp | b | se | pval |
|---|---|---|---|---|
| MR Egger | 30 | 0.0976370 | 0.0428959 | 0.0306839 |
| Weighted median | 30 | 0.0519955 | 0.0312193 | 0.0958142 |
| Inverse variance weighted | 30 | 0.0460862 | 0.0205671 | 0.0250407 |
| Simple mode | 30 | 0.0115106 | 0.0581926 | 0.8445799 |
| Weighted mode | 30 | 0.0742703 | 0.0316121 | 0.0258237 |

##### Heterogeneity tests

| method | Q | Q_df | Q_pval |
|---|---|---|---|
| MR Egger | 29.99023 | 28 | 0.3636850 |
| Inverse variance weighted | 31.98178 | 29 | 0.3206736 |

##### Test for directional horizontal pleiotropy

| egger_intercept | se | pval |
|---|---|---|
| -0.0100788 | 0.0073914 | 0.1835594 |

### Forest plot of single SNP MR

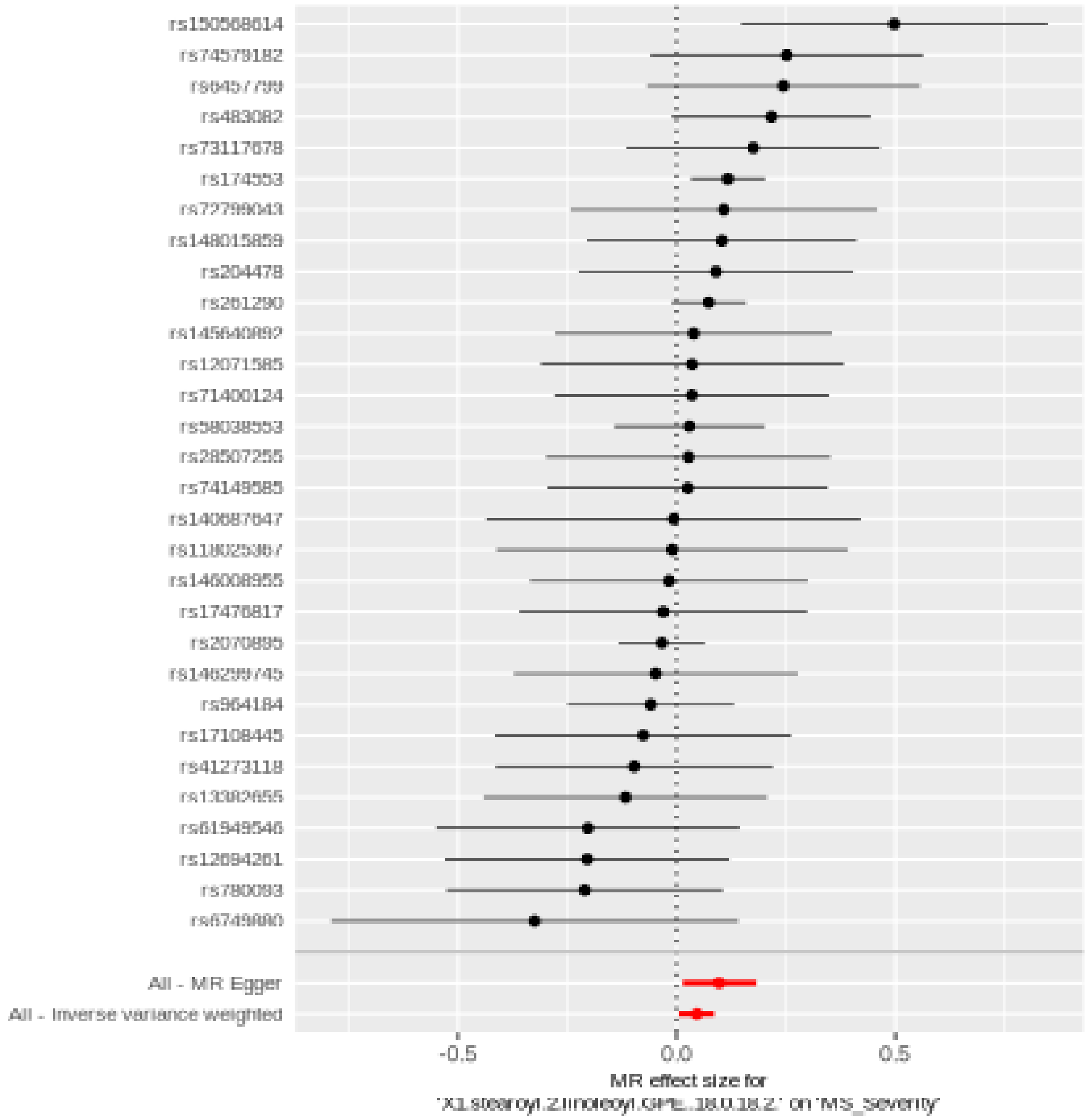

Comparison of results using different MR methods

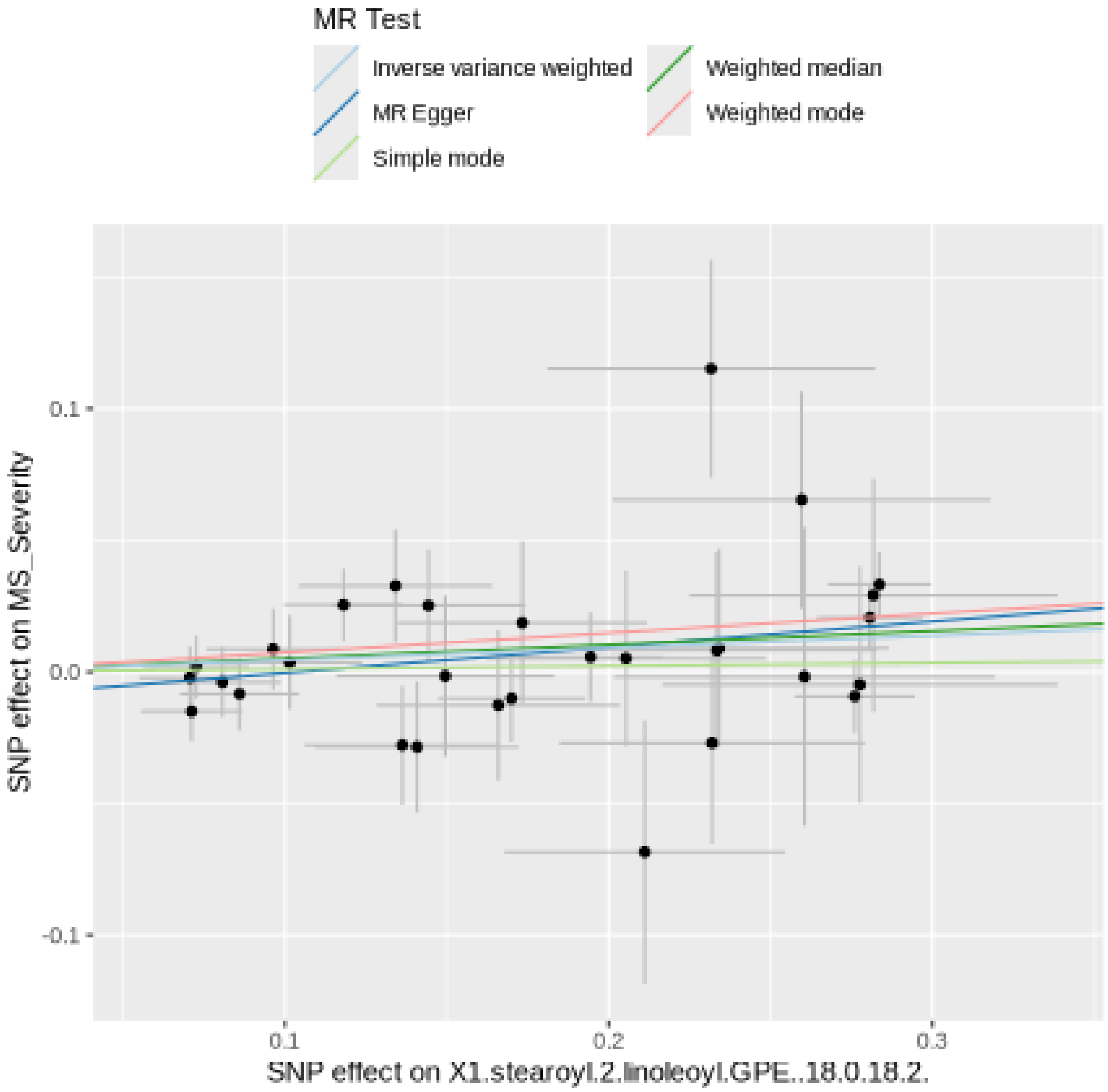

Funnel plot

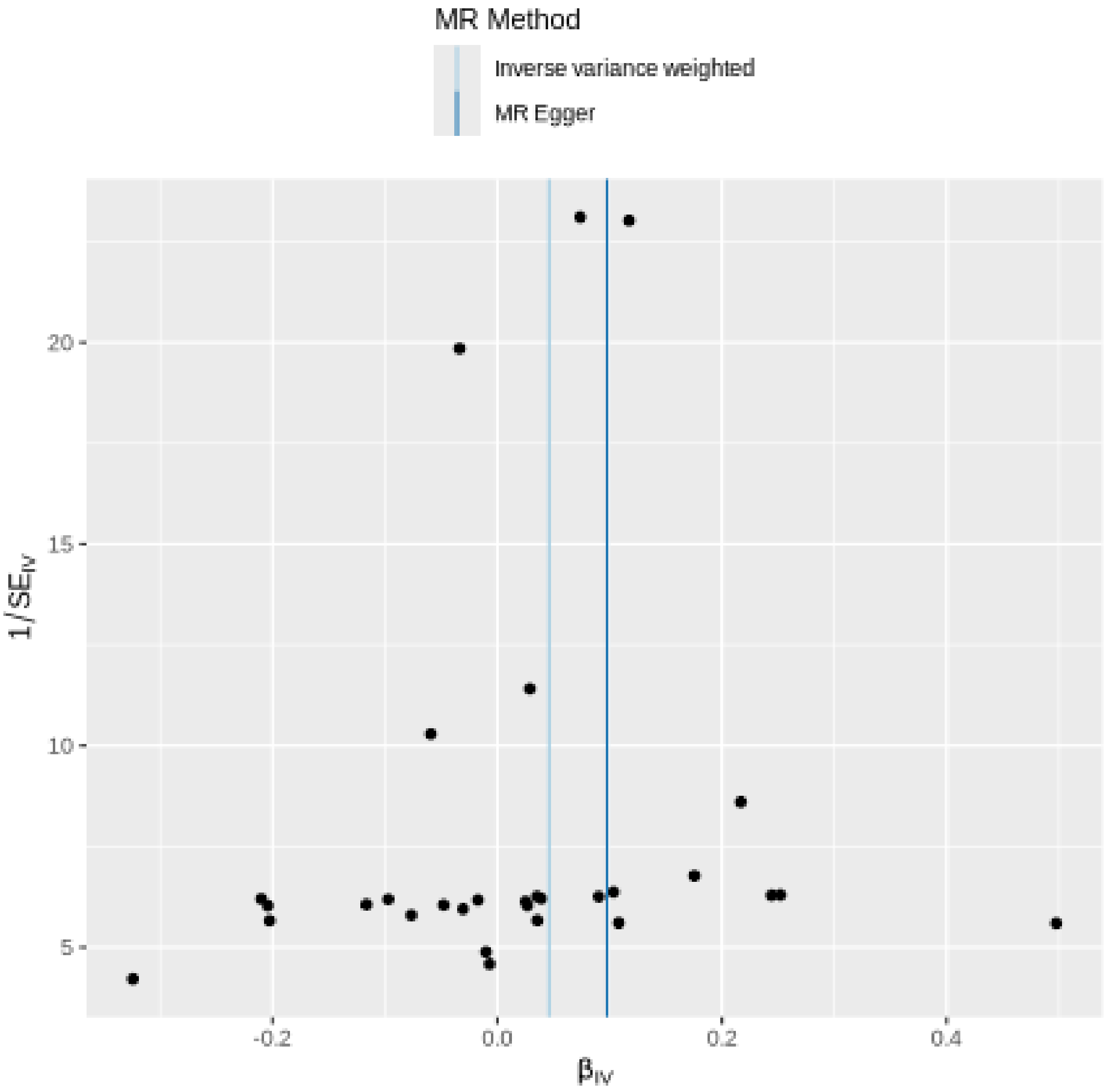

MR Method
Inverse variance weighted
MR Egger
20
15
10
5
$1/SE_{IV}$
-0.2
0.0
0.2
0.4
$\beta_{IV}$

Leave-one-out sensitivity analysis

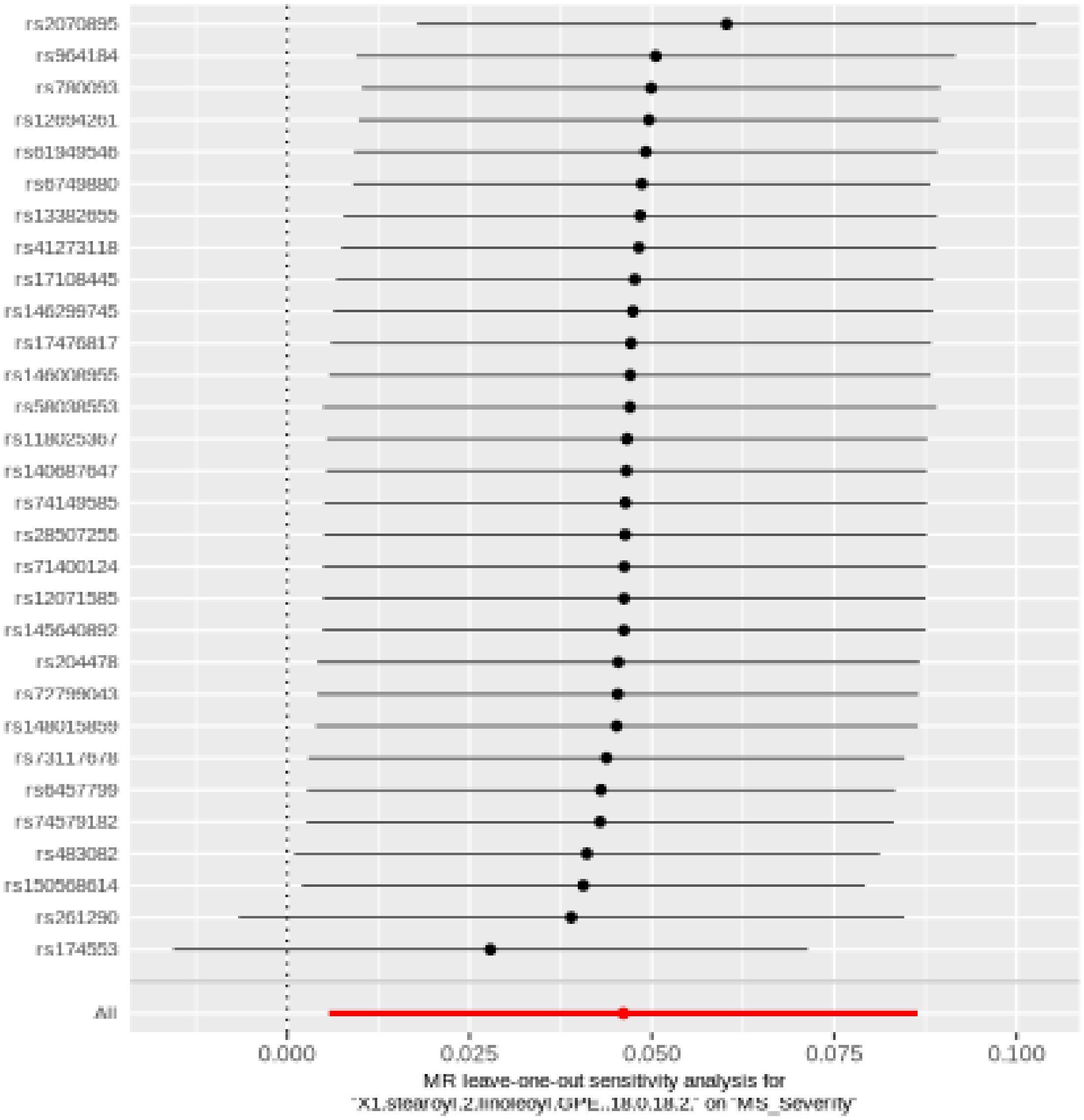

#### 1-stearoyl-2-docosahexaenoyl-gpc (18:0/22:6)

| method | nsnp | b | se | pval |
|---|---|---|---|---|
| MR Egger | 34 | -0.0509193 | 0.0671001 | 0.4534922 |
| Weighted median | 34 | -0.1350475 | 0.0372759 | 0.0002913 |
| Inverse variance weighted | 34 | -0.0634682 | 0.0306311 | 0.0382636 |
| Simple mode | 34 | -0.0676622 | 0.0912365 | 0.4635672 |
| Weighted mode | 34 | -0.1238664 | 0.0407932 | 0.0046493 |

##### Heterogeneity tests

| method | Q | Q_df | Q_pval |
|---|---|---|---|
| MR Egger | 53.72234 | 32 | 0.0094517 |
| Inverse variance weighted | 53.79710 | 33 | 0.0125624 |

##### Test for directional horizontal pleiotropy

| egger_intercept | se | pval |
|---|---|---|
| -0.0018677 | 0.0088507 | 0.834205 |

### Forest plot of single SNP MR

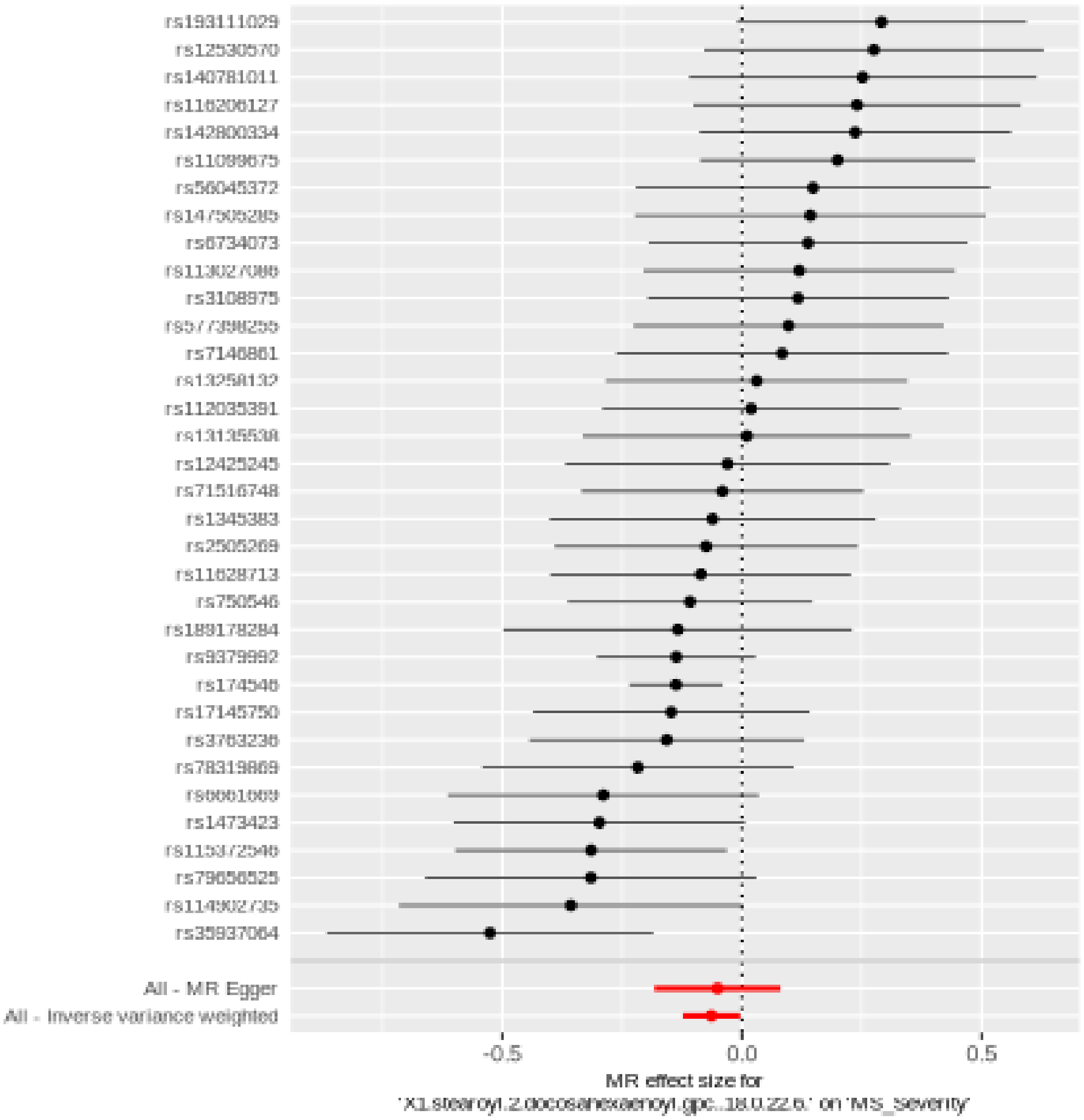

Comparison of results using different MR methods

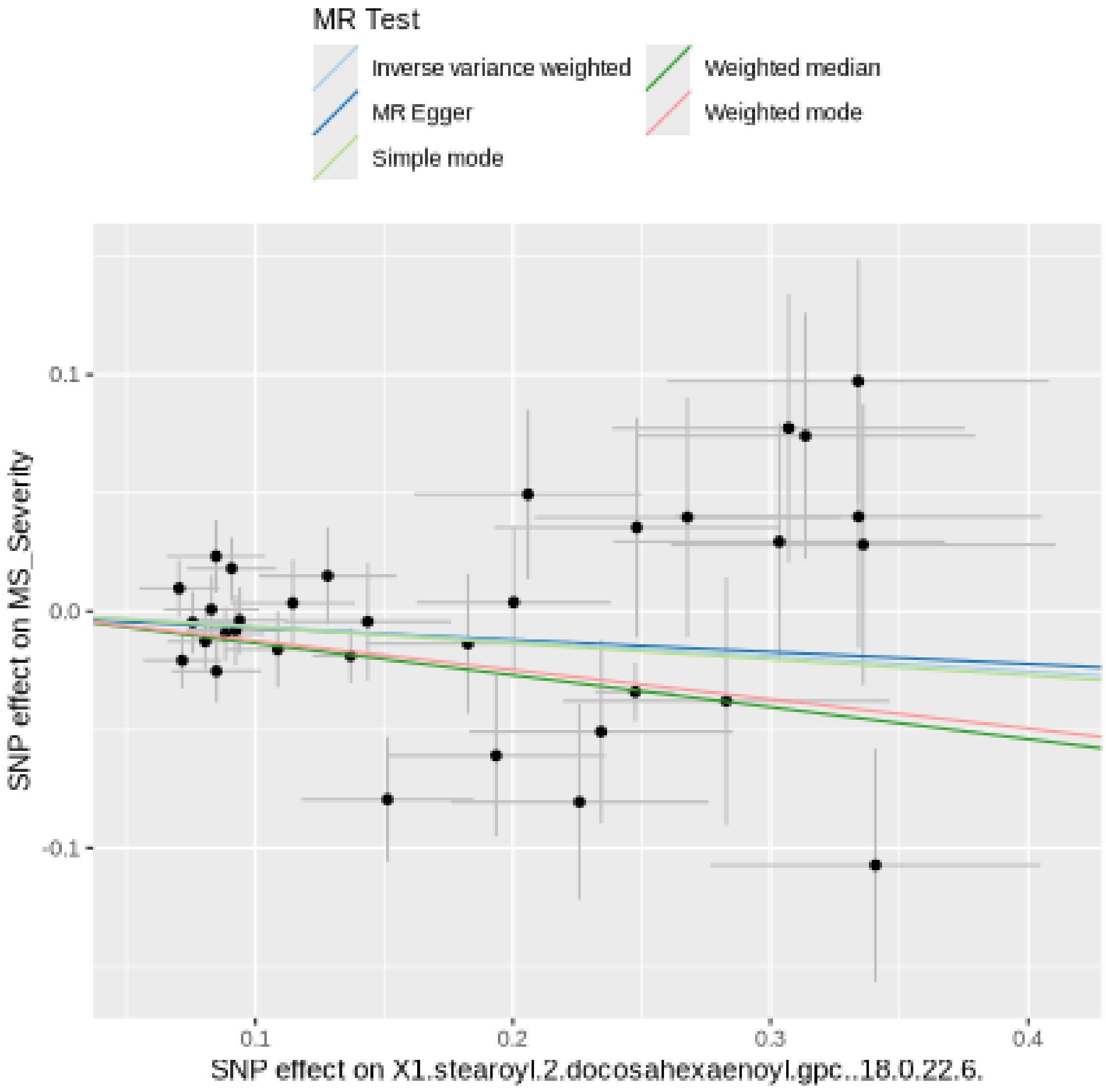

Funnel plot

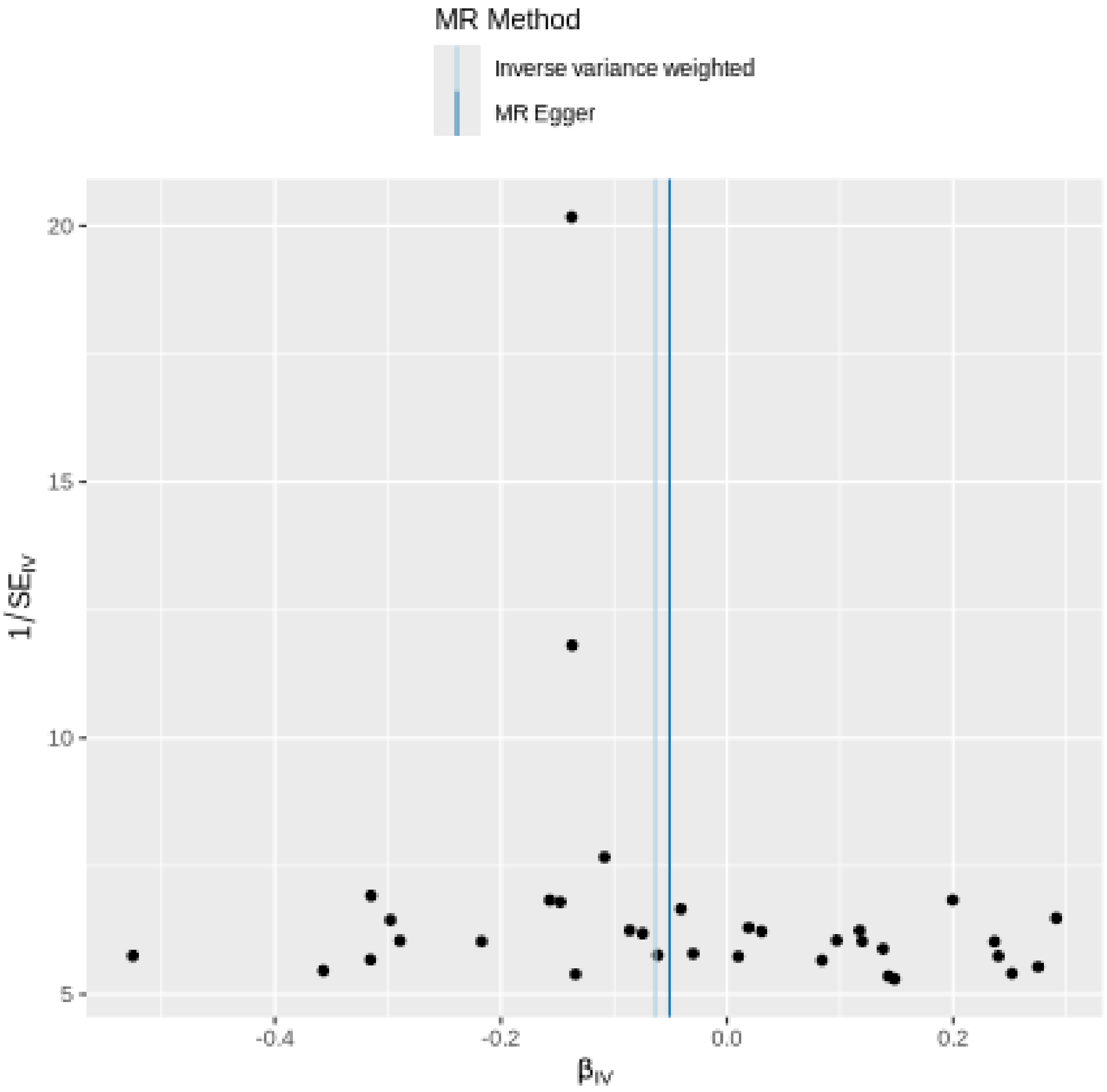

MR Method
Inverse variance weighted
MR Egger
20
15
10
5
1/SE$_{IV}$
-0.4
-0.2
0.0
0.2
β$_{IV}$

#### Leave-one-out sensitivity analysis

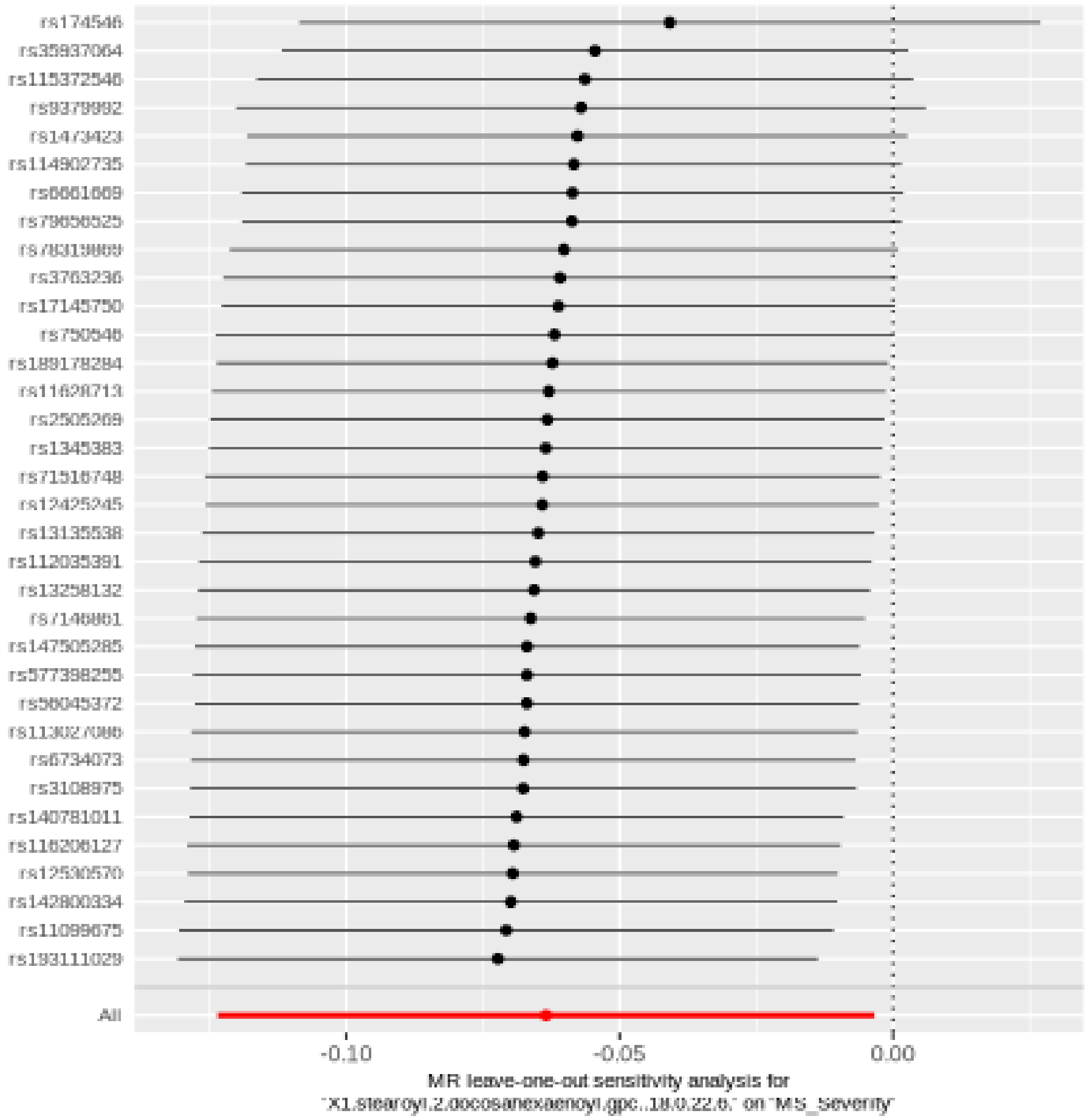

#### 1-(1-enyl-stearoyl)-2-arachidonoyl-GPE (p-18:0/20:4)

| method | nsnp | b | se | pval |
|---|---|---|---|---|
| MR Egger | 24 | -0.0889856 | 0.0561341 | 0.1271837 |
| Weighted median | 24 | -0.1025662 | 0.0389689 | 0.0084884 |
| Inverse variance weighted | 24 | -0.0727563 | 0.0268966 | 0.0068297 |
| Simple mode | 24 | -0.1088692 | 0.0804710 | 0.1892381 |
| Weighted mode | 24 | -0.1025497 | 0.0403316 | 0.0181928 |

##### Heterogeneity tests

| method | Q | Q_df | Q_pval |
|---|---|---|---|
| MR Egger | 23.23148 | 22 | 0.3887587 |
| Inverse variance weighted | 23.34744 | 23 | 0.4406418 |

##### Test for directional horizontal pleiotropy

| egger_intercept | se | pval |
|---|---|---|
| 0.0027655 | 0.0083452 | 0.7434906 |

Forest plot of single SNP MR

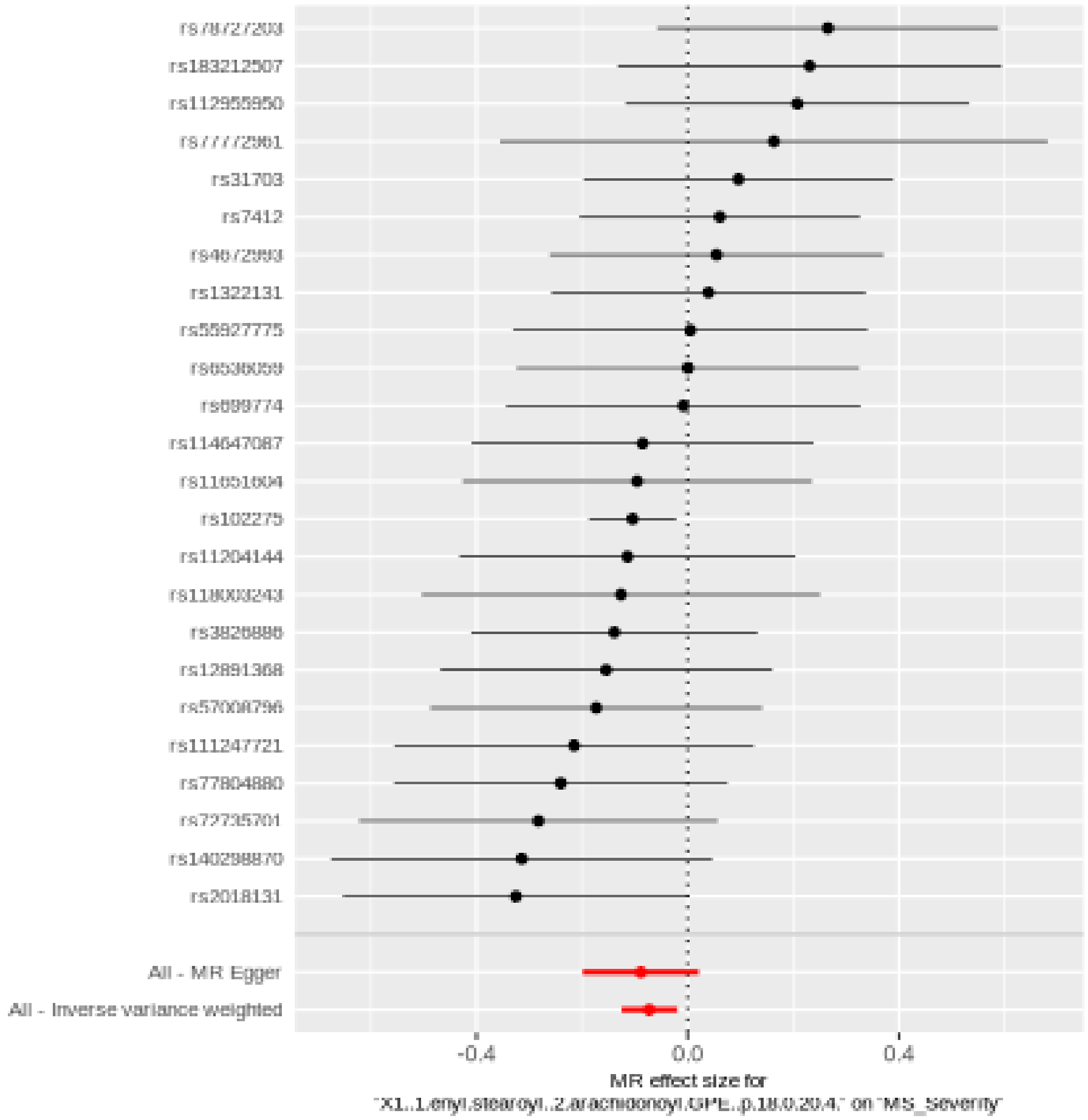

Comparison of results using different MR methods

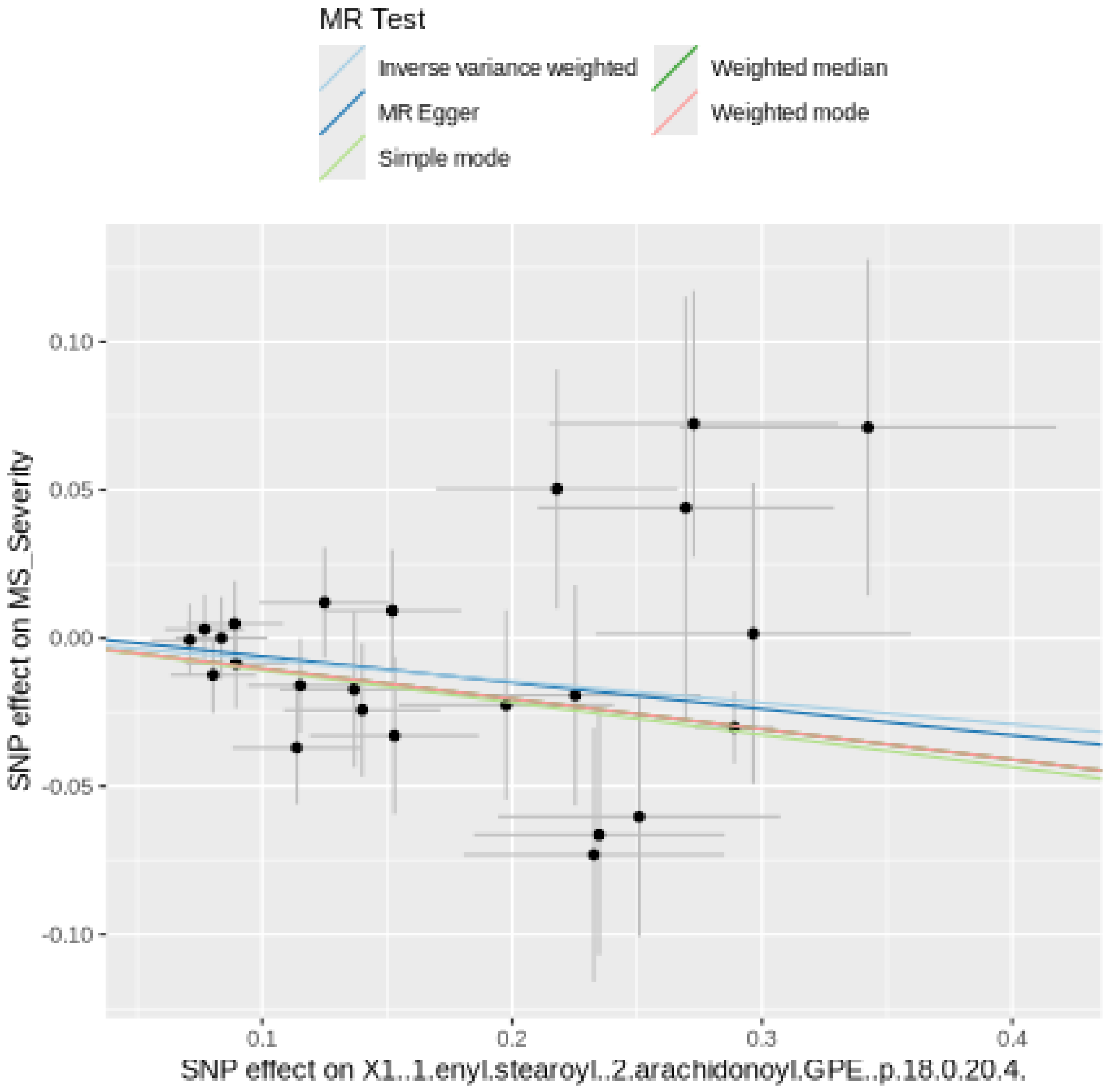

Funnel plot

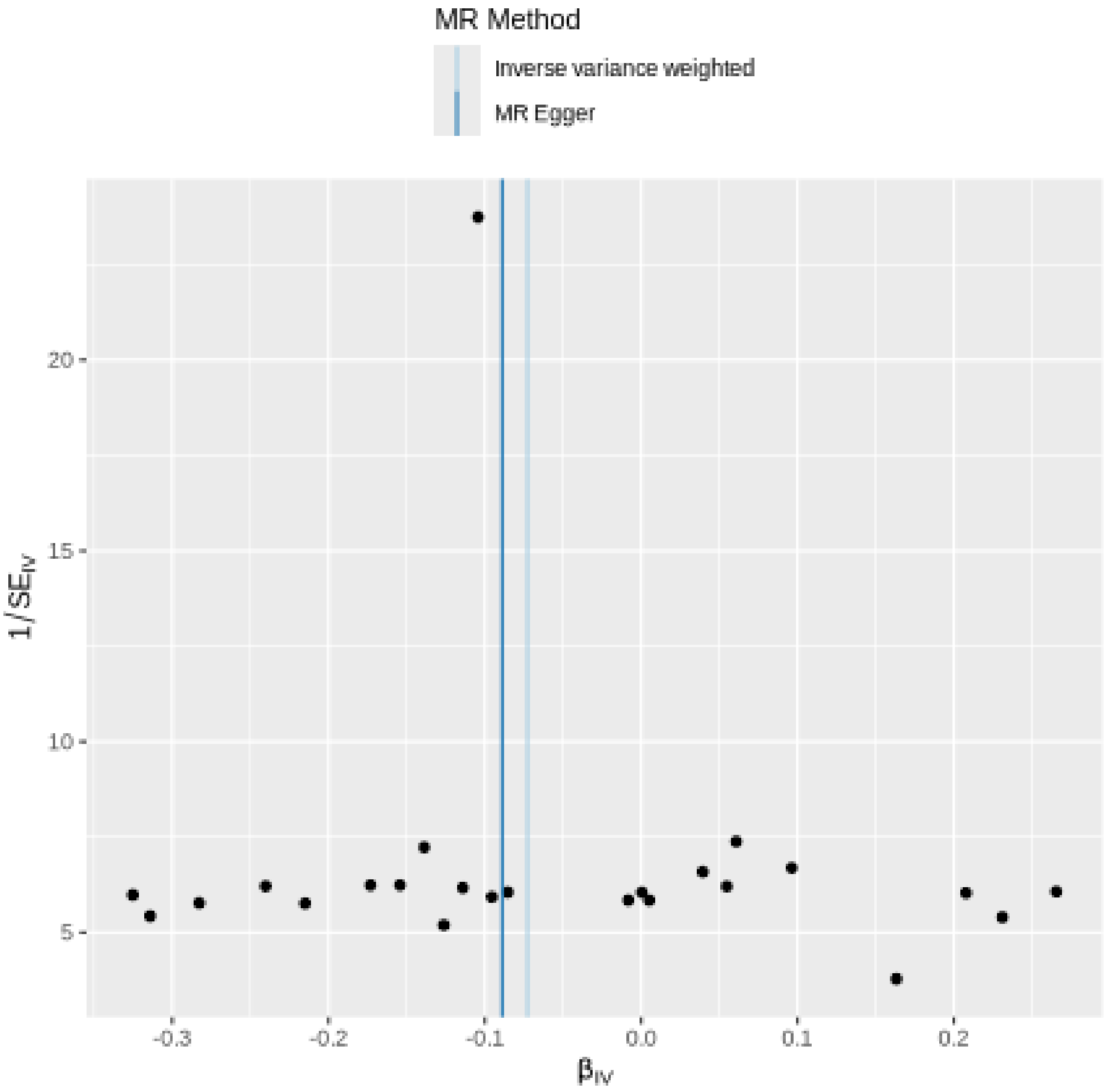

MR Method
Inverse variance weighted
MR Egger
20
15
10
5
1/SE$_{IV}$
-0.3
-0.2
-0.1
0.0
0.1
0.2
β$_{IV}$

Leave-one-out sensitivity analysis

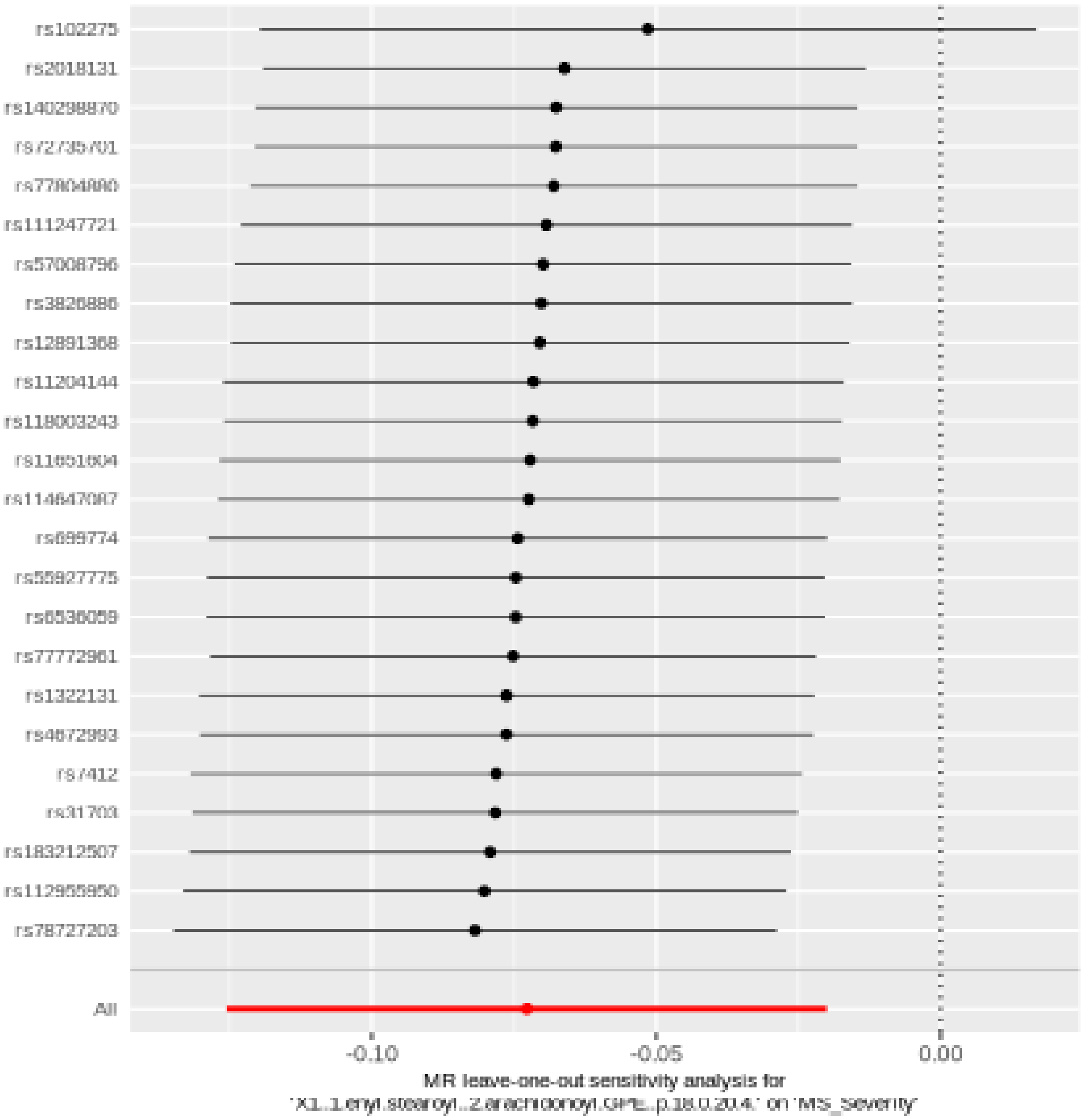

#### 1-linoleoyl-GPG (18:2)

| method | nsnp | b | se | pval |
|---|---|---|---|---|
| MR Egger | 19 | 0.1041508 | 0.0440103 | 0.0300934 |
| Weighted median | 19 | 0.0662854 | 0.0325598 | 0.0417701 |
| Inverse variance weighted | 19 | 0.0610690 | 0.0261613 | 0.0195786 |
| Simple mode | 19 | 0.0688525 | 0.0624730 | 0.2849302 |
| Weighted mode | 19 | 0.0668130 | 0.0344668 | 0.0684103 |

Heterogeneity tests

| method | Q | Q_df | Q_pval |
|---|---|---|---|
| MR Egger | 17.95382 | 17 | 0.3917602 |
| Inverse variance weighted | 19.49774 | 18 | 0.3617941 |

Test for directional horizontal pleiotropy

| egger_intercept | se | pval |
|---|---|---|
| -0.0085964 | 0.0071098 | 0.2431764 |

Forest plot of single SNP MR

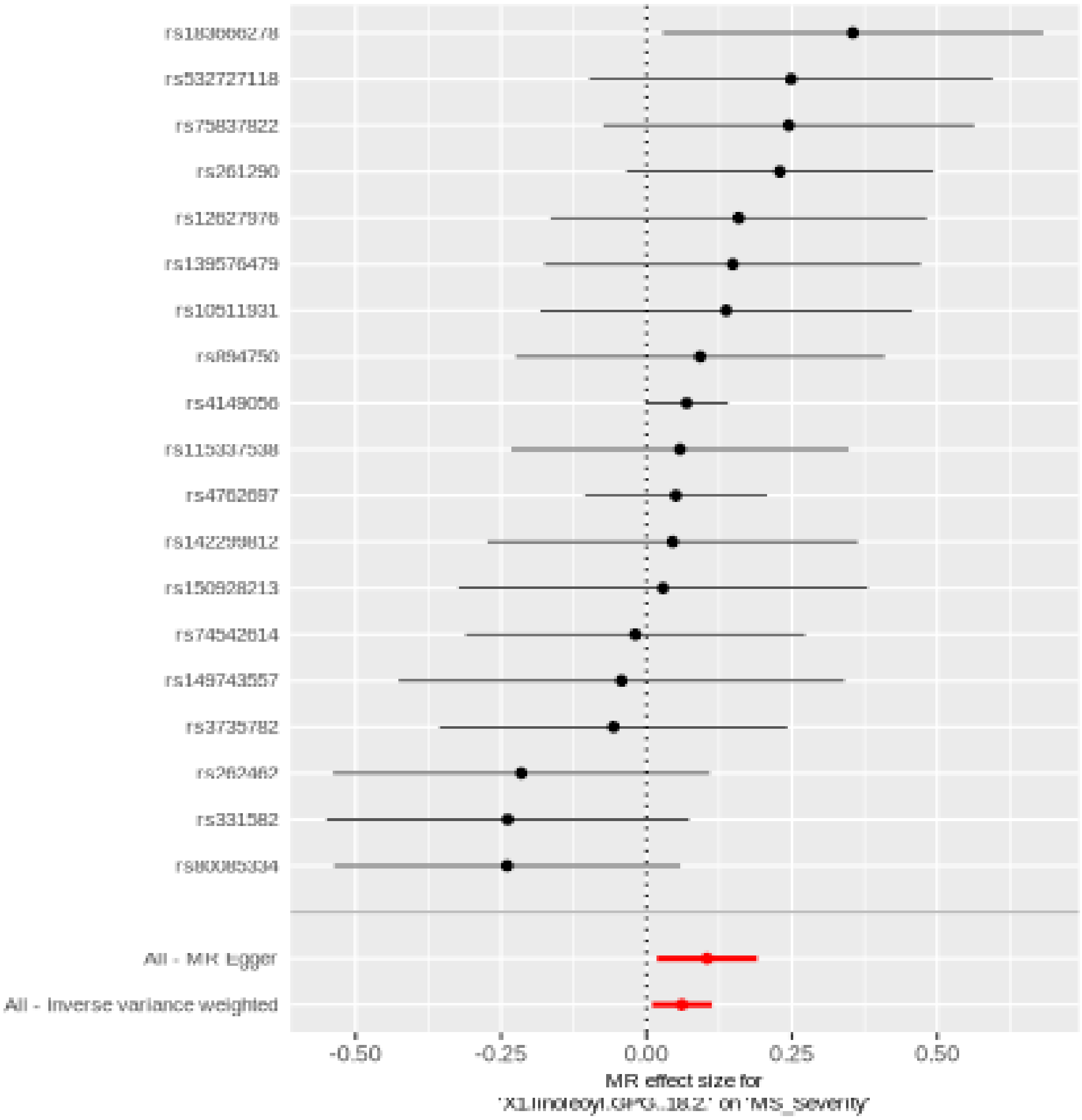

Comparison of results using different MR methods

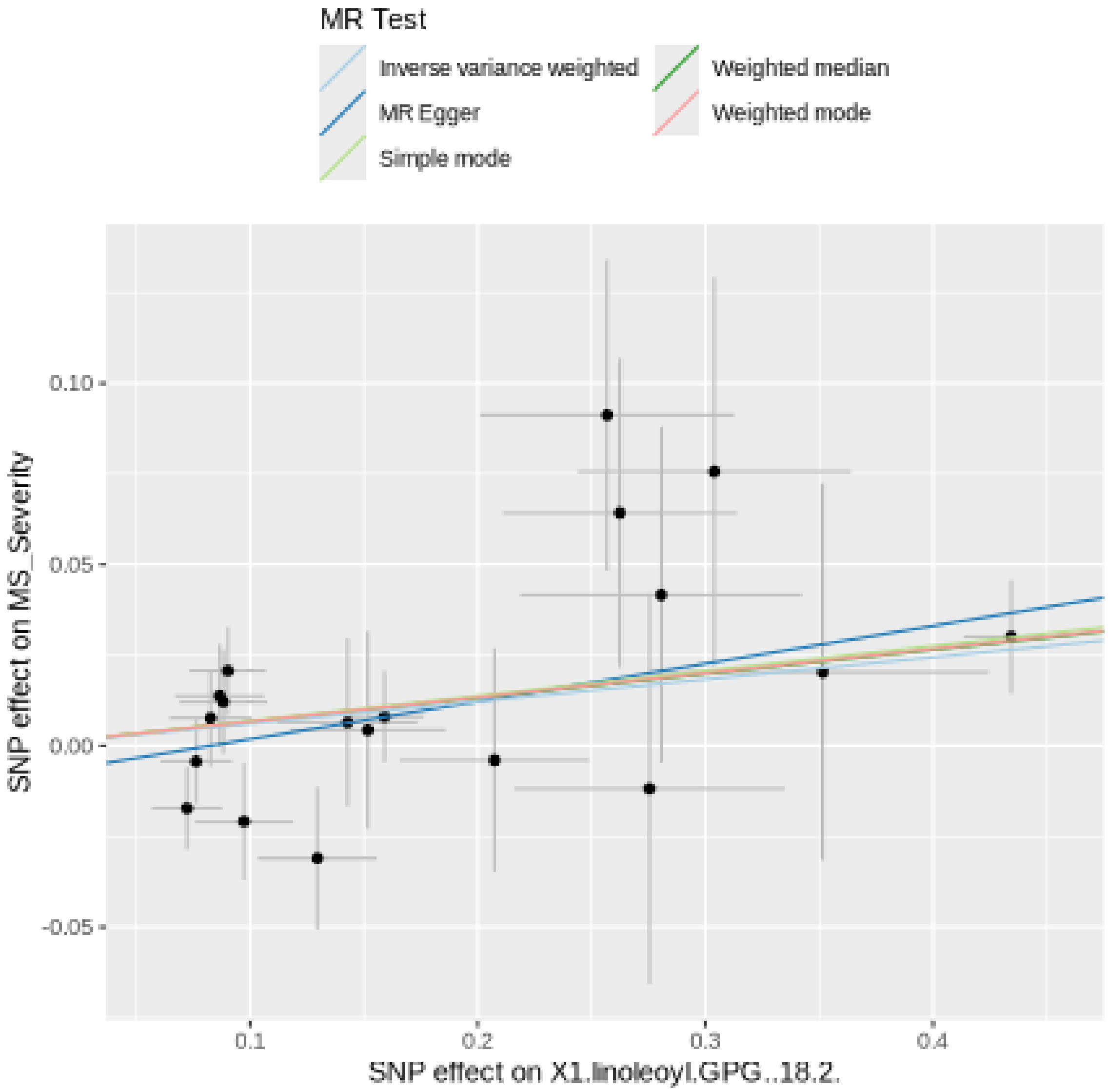

Funnel plot

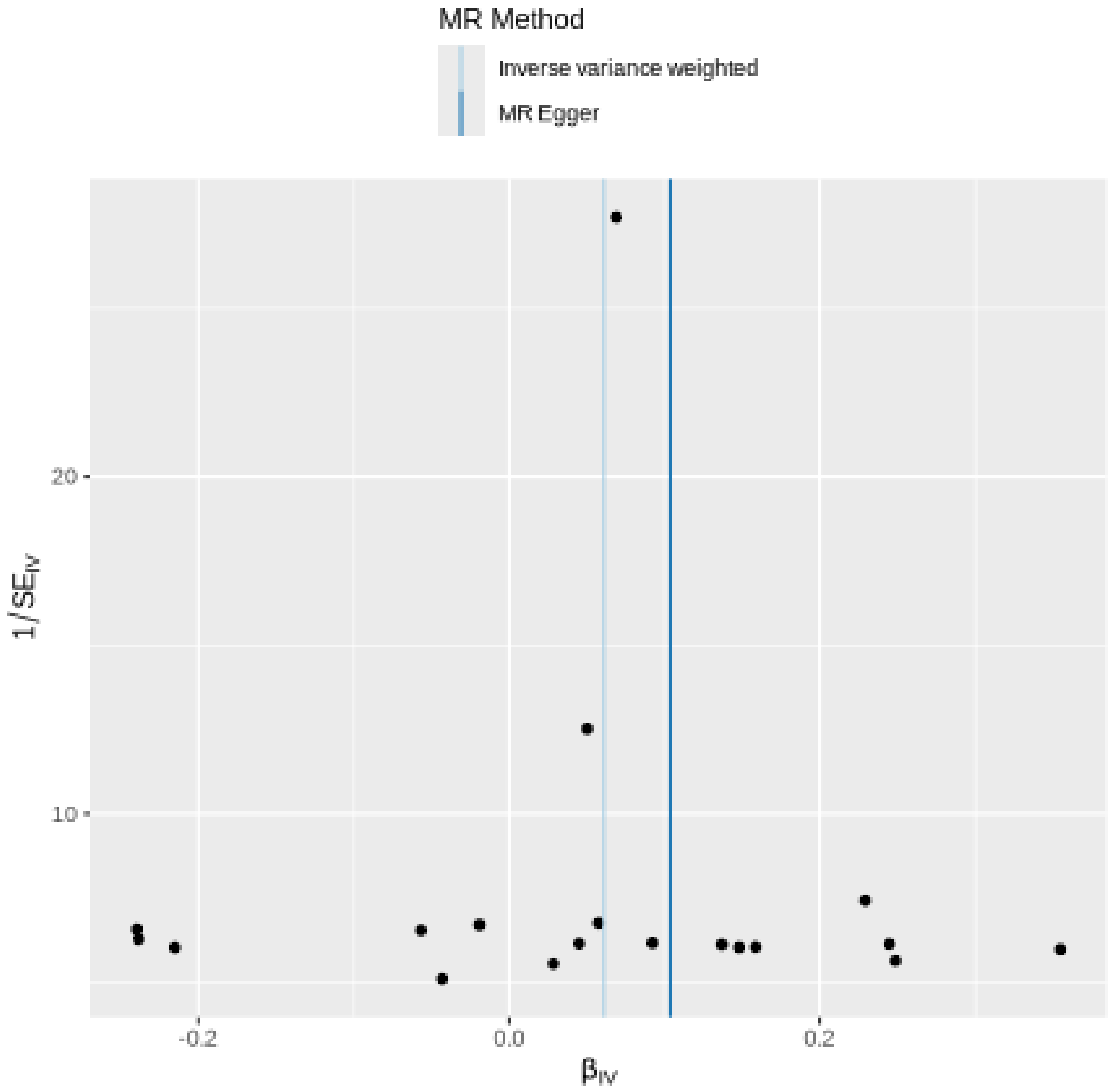

MR Method
Inverse variance weighted
MR Egger
20
10
1/SE$_{IV}$
-0.2
0.0
0.2
β$_{IV}$

Leave-one-out sensitivity analysis

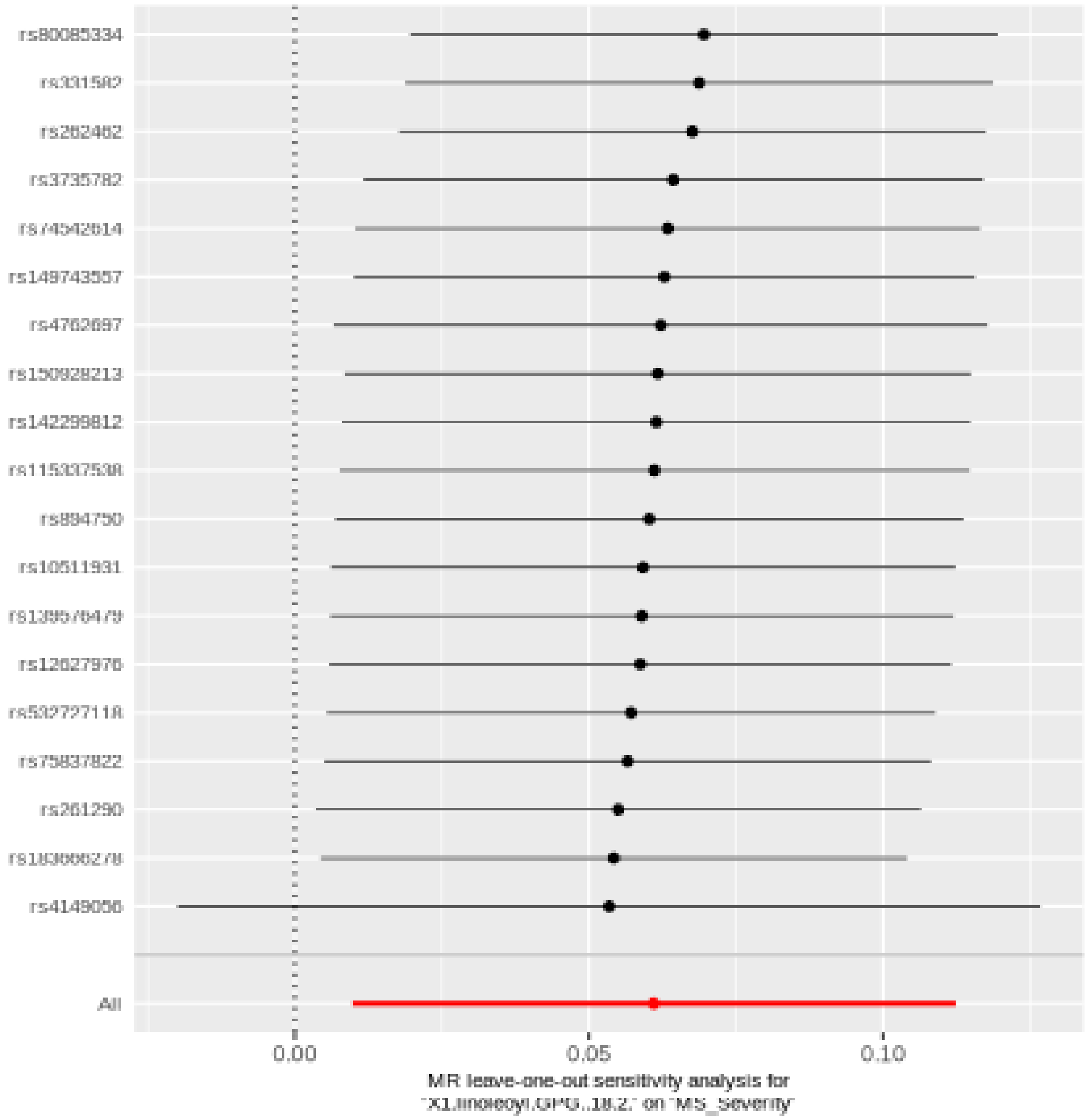

#### Sphingomyelin (d18:2/21:0, d16:2/23:0)

| method | nsnp | b | se | pval |
|---|---|---|---|---|
| MR Egger | 37 | -0.2157359 | 0.0758270 | 0.0073708 |
| Weighted median | 37 | -0.0958854 | 0.0444776 | 0.0310983 |
| Inverse variance weighted | 37 | -0.0914968 | 0.0336532 | 0.0065516 |
| Simple mode | 37 | -0.1677948 | 0.0999012 | 0.1016935 |
| Weighted mode | 37 | -0.1622838 | 0.0771030 | 0.0423567 |

##### Heterogeneity tests

| method | Q | Q_df | Q_pval |
|---|---|---|---|
| MR Egger | 44.85360 | 35 | 0.1229224 |
| Inverse variance weighted | 49.07573 | 36 | 0.0717831 |

##### Test for directional horizontal pleiotropy

| egger_intercept | se | pval |
|---|---|---|
| 0.0134161 | 0.0073914 | 0.0780854 |

### Forest plot of single SNP MR

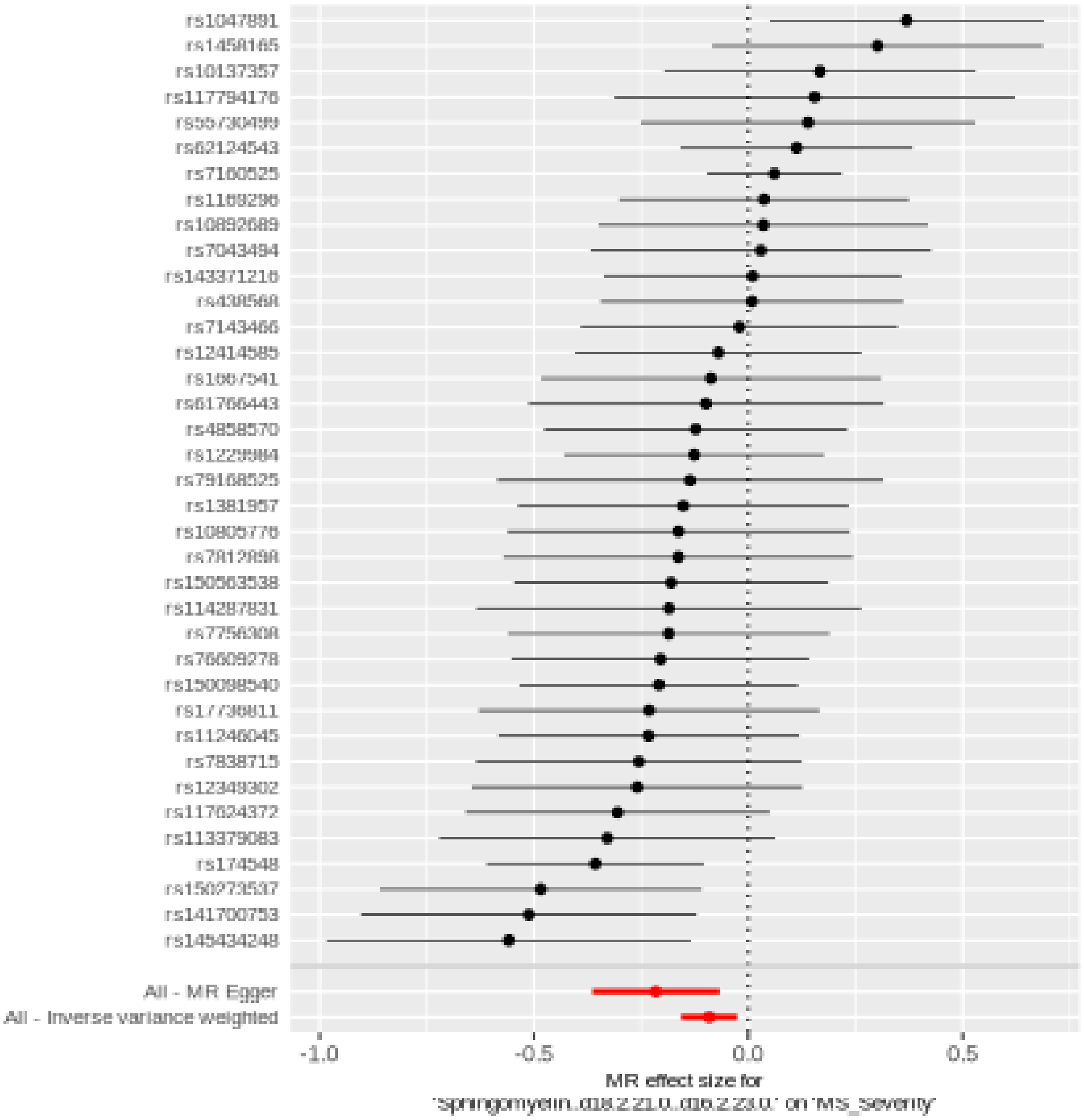

Comparison of results using different MR methods

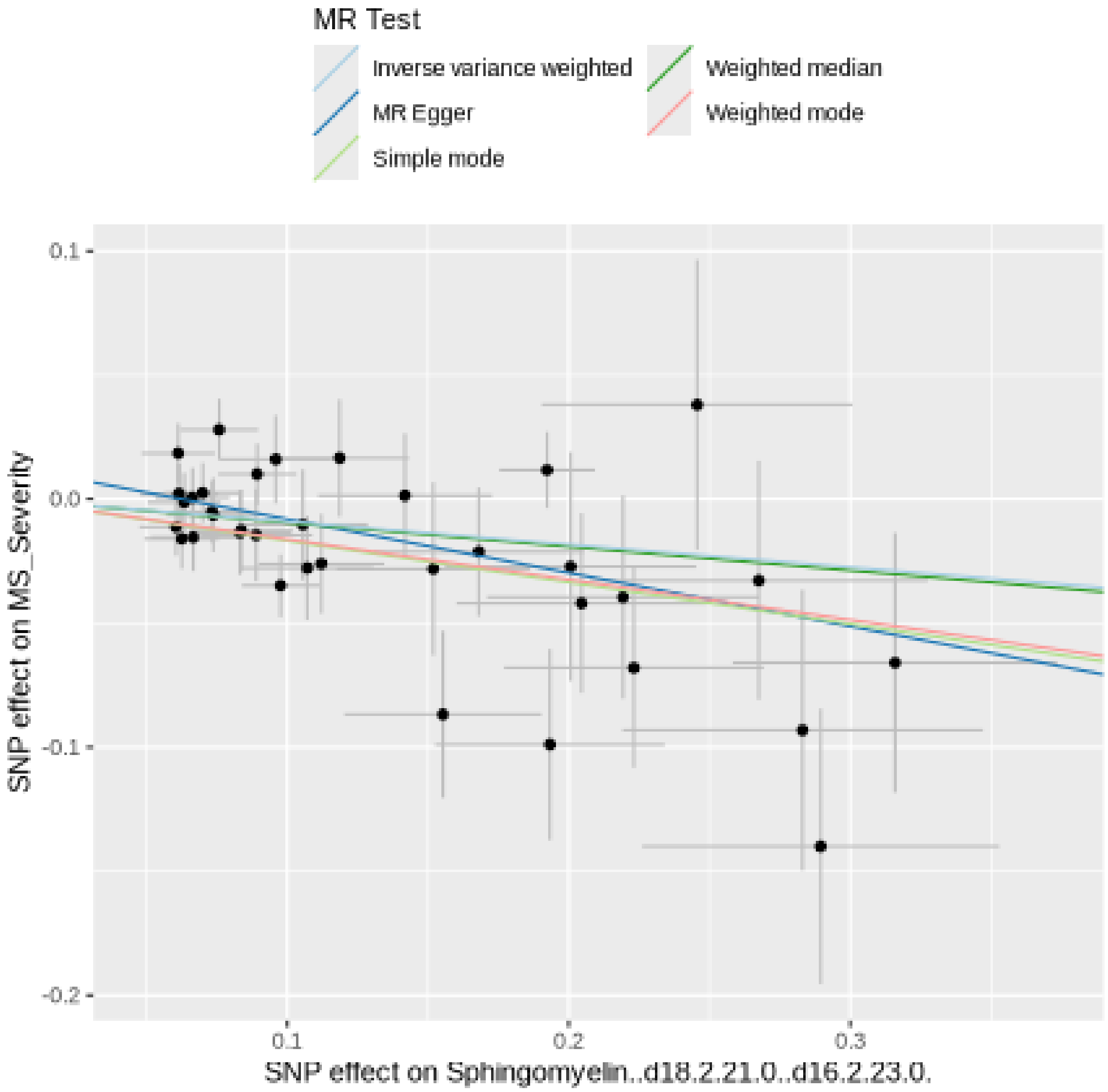

Funnel plot

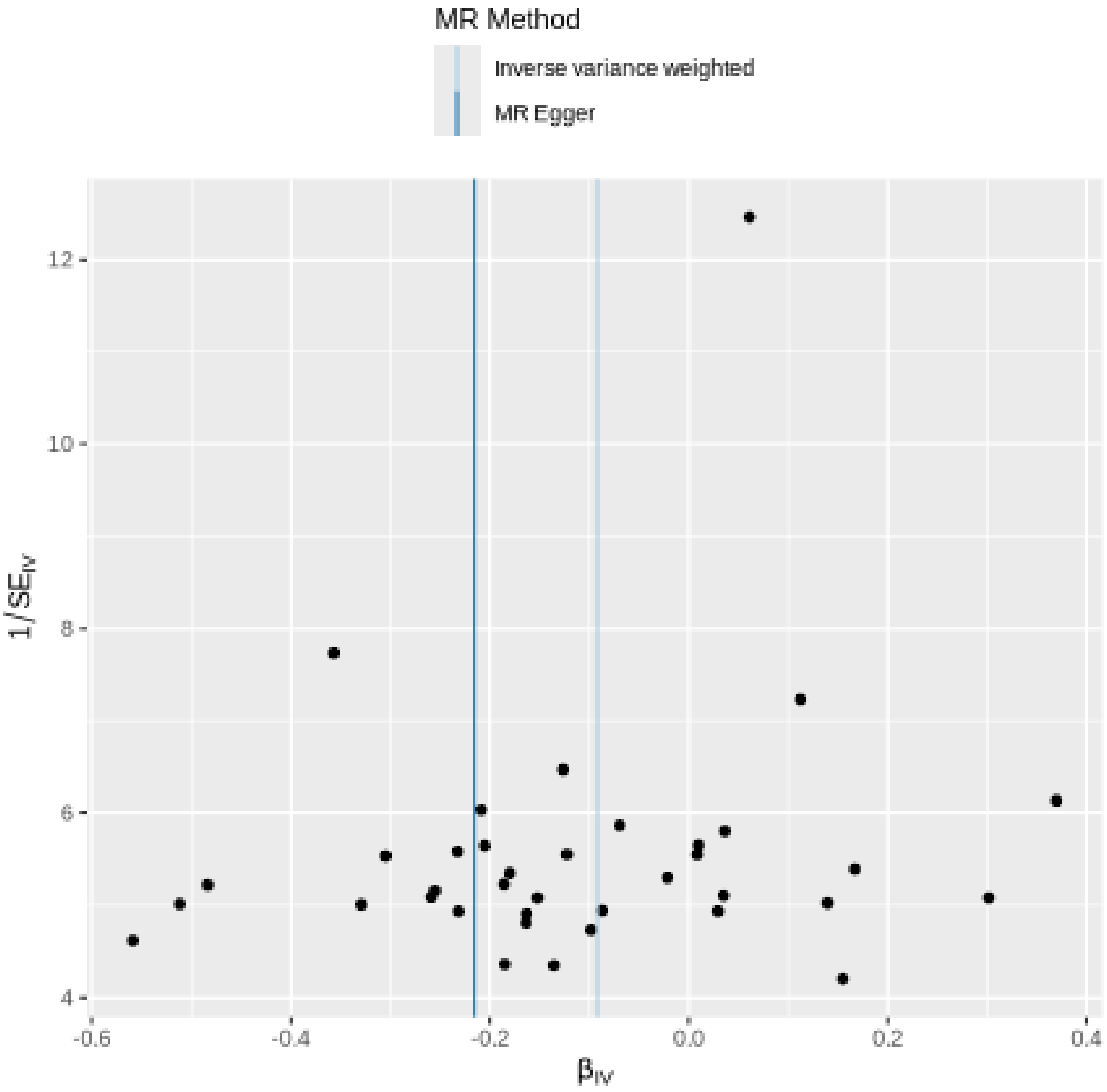
MR Method
Inverse variance weighted
MR Egger
12
10
8
6
4
$1/SE_{IV}$
-0.6
-0.4
-0.2
0.0
0.2
0.4
$\beta_{IV}$

Leave-one-out sensitivity analysis

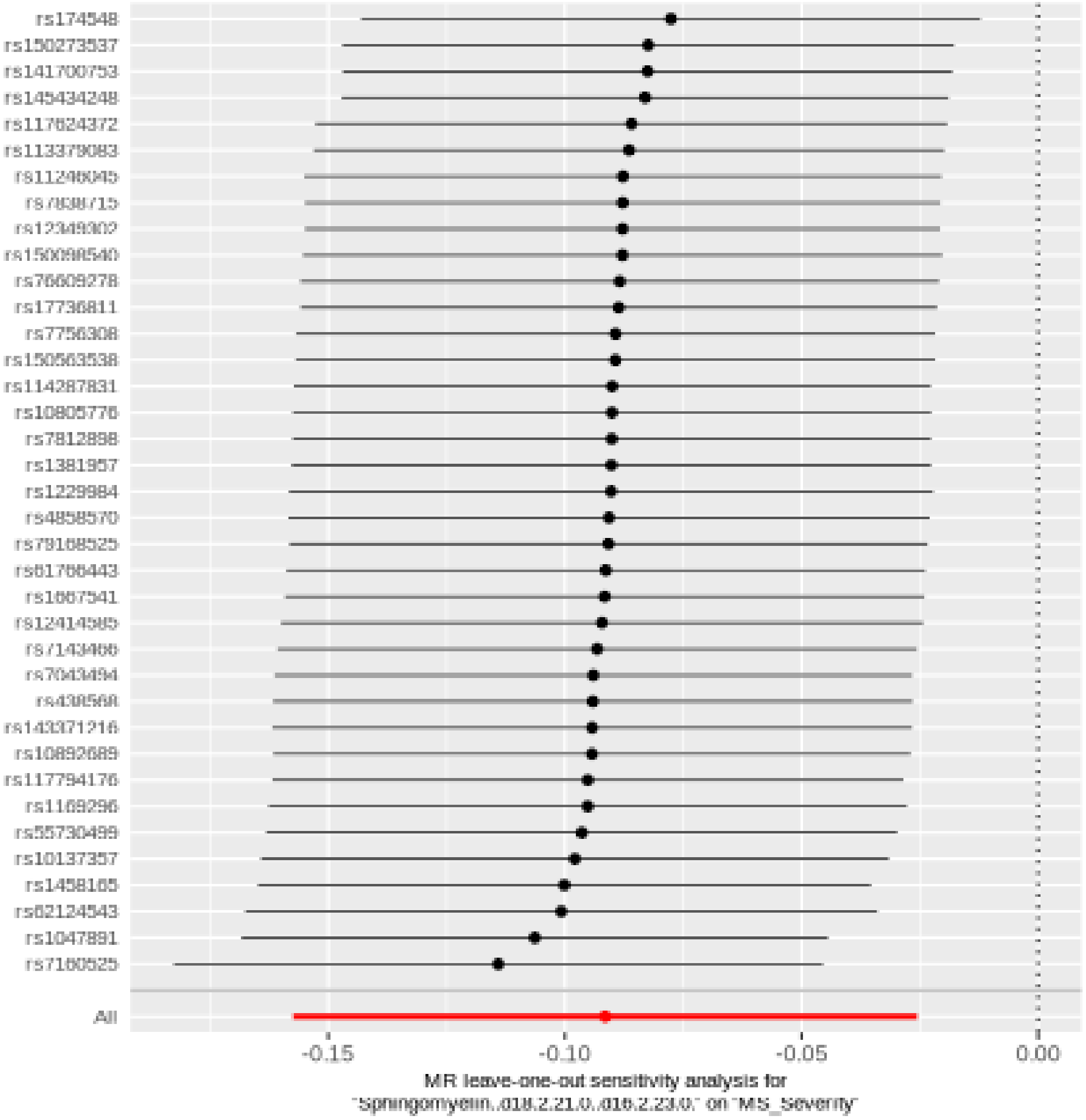

#### Lignoceroylcarnitine (C24)

| method | nsnp | b | se | pval |
|---|---|---|---|---|
| MR Egger | 26 | 0.0842382 | 0.0526150 | 0.1224528 |
| Weighted median | 26 | 0.0299719 | 0.0376718 | 0.4262614 |
| Inverse variance weighted | 26 | 0.0545883 | 0.0237927 | 0.0217714 |
| Simple mode | 26 | 0.0083092 | 0.0724011 | 0.9095477 |
| Weighted mode | 26 | 0.0300103 | 0.0422313 | 0.4838975 |

##### Heterogeneity tests

| method | Q | Q_df | Q_pval |
|---|---|---|---|
| MR Egger | 25.06932 | 24 | 0.4020015 |
| Inverse variance weighted | 25.48894 | 25 | 0.4352572 |

##### Test for directional horizontal pleiotropy

| egger_intercept | se | pval |
|---|---|---|
| -0.0055249 | 0.0087168 | 0.5321959 |

#### Forest plot of single SNP MR

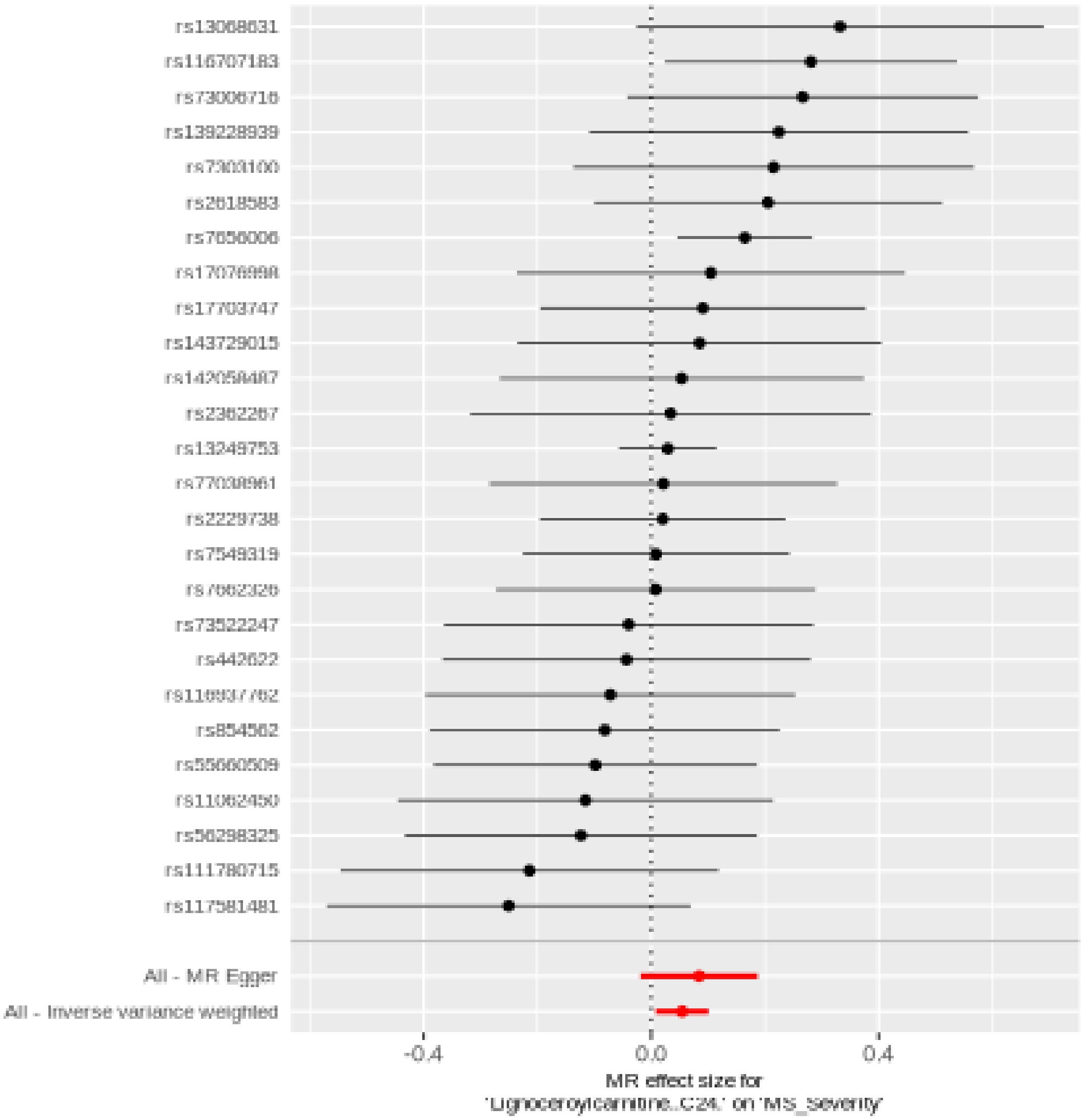

Comparison of results using different MR methods

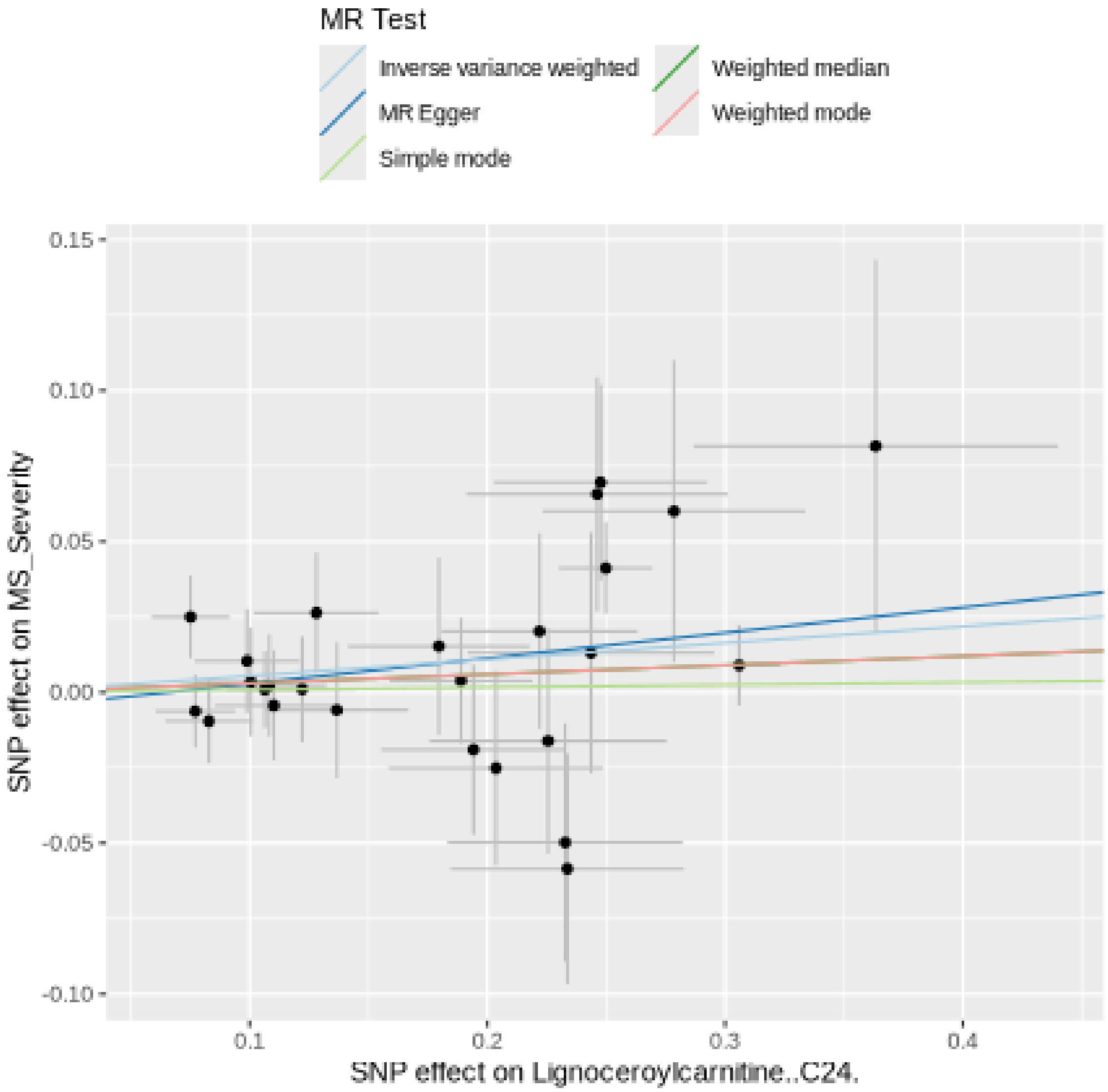

Funnel plot

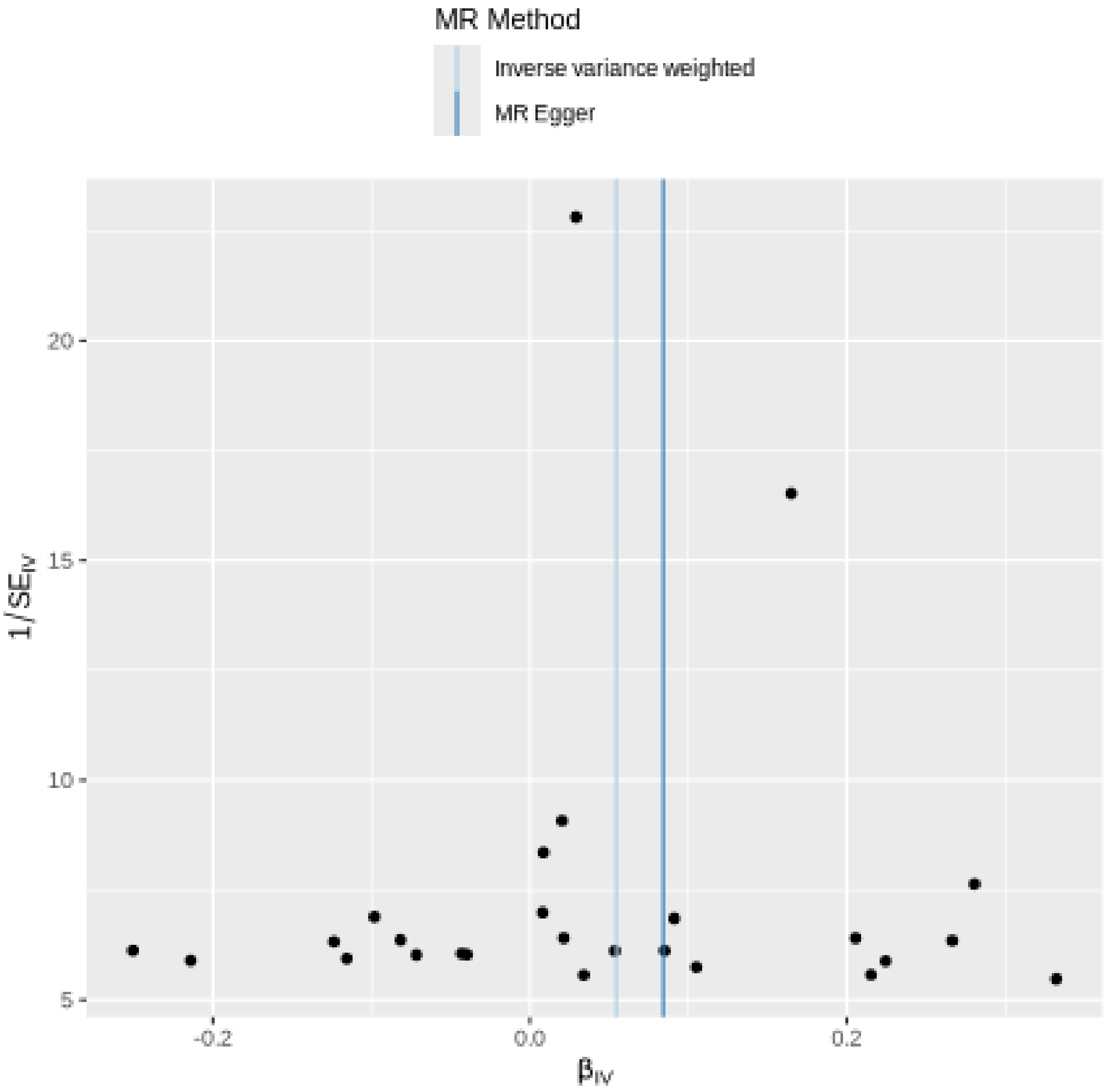
MR Method
Inverse variance weighted
MR Egger
20
15
10
5
$1/SE_{IV}$
-0.2
0.0
0.2
$\beta_{IV}$

Leave-one-out sensitivity analysis

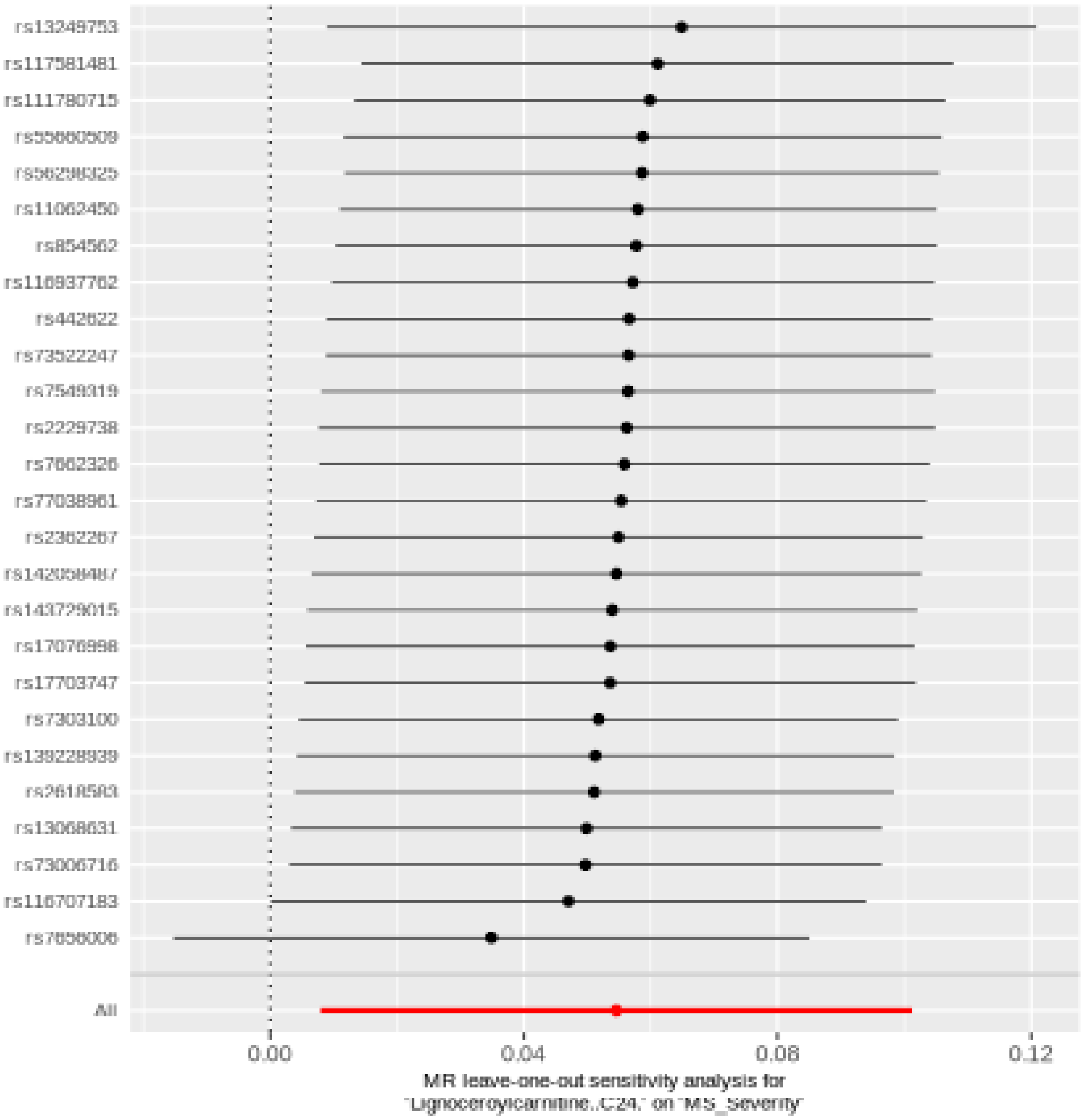

#### Arachidonoylcarnitine (C20:4)

| method | nsnp | b | se | pval |
|---|---|---|---|---|
| MR Egger | 45 | -0.0548922 | 0.0271954 | 0.0498109 |
| Weighted median | 45 | -0.0333299 | 0.0240131 | 0.1651406 |
| Inverse variance weighted | 45 | -0.0356091 | 0.0156910 | 0.0232443 |
| Simple mode | 45 | -0.0640065 | 0.0507124 | 0.2135454 |
| Weighted mode | 45 | -0.0390483 | 0.0253002 | 0.1298965 |

##### Heterogeneity tests

| method | Q | Q_df | Q_pval |
|---|---|---|---|
| MR Egger | 42.89848 | 43 | 0.4756693 |
| Inverse variance weighted | 43.65213 | 44 | 0.4864352 |

##### Test for directional horizontal pleiotropy

| egger_intercept | se | pval |
|---|---|---|
| 0.0037946 | 0.004371 | 0.3901433 |

Forest plot of single SNP MR

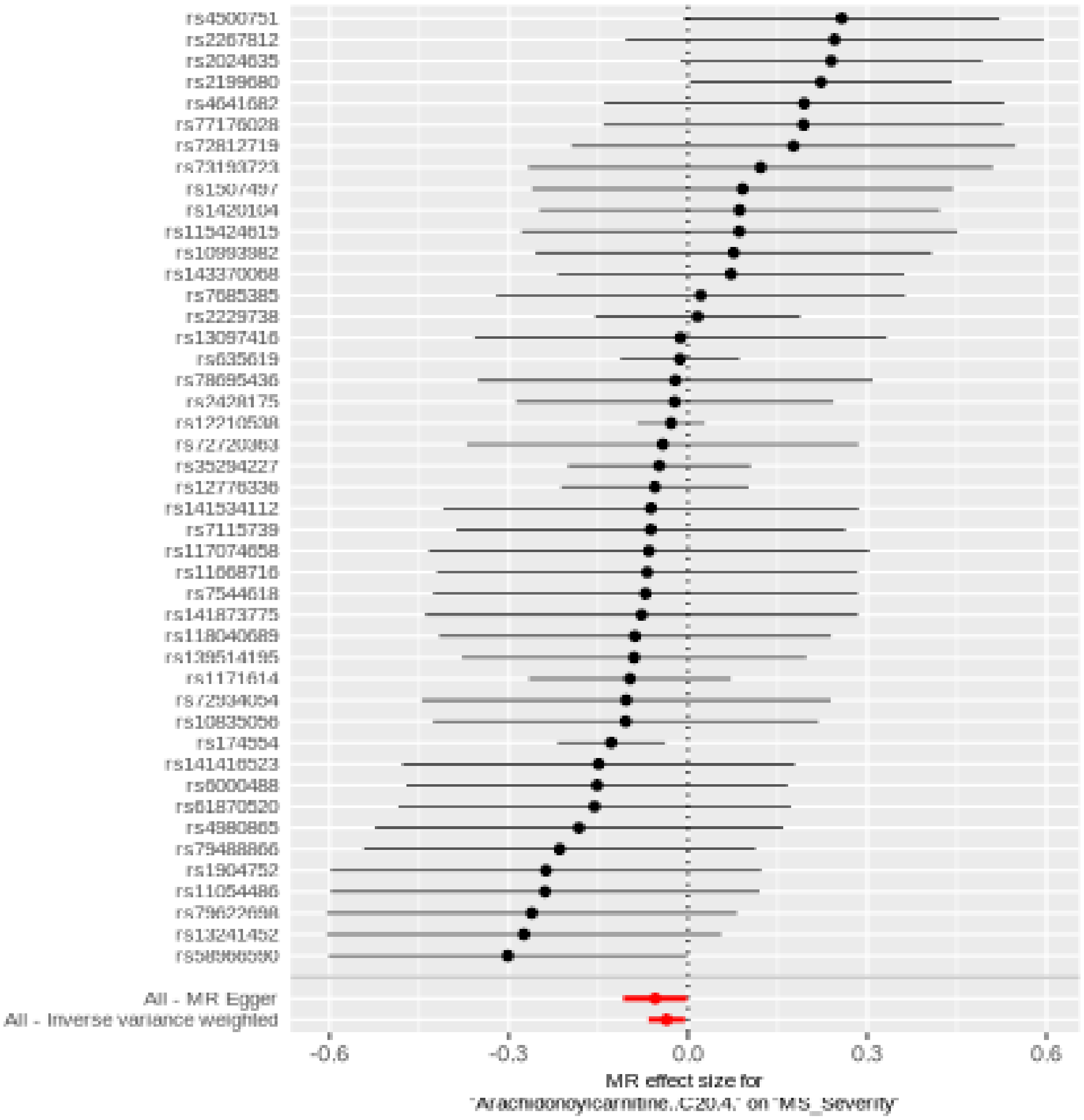

Comparison of results using different MR methods

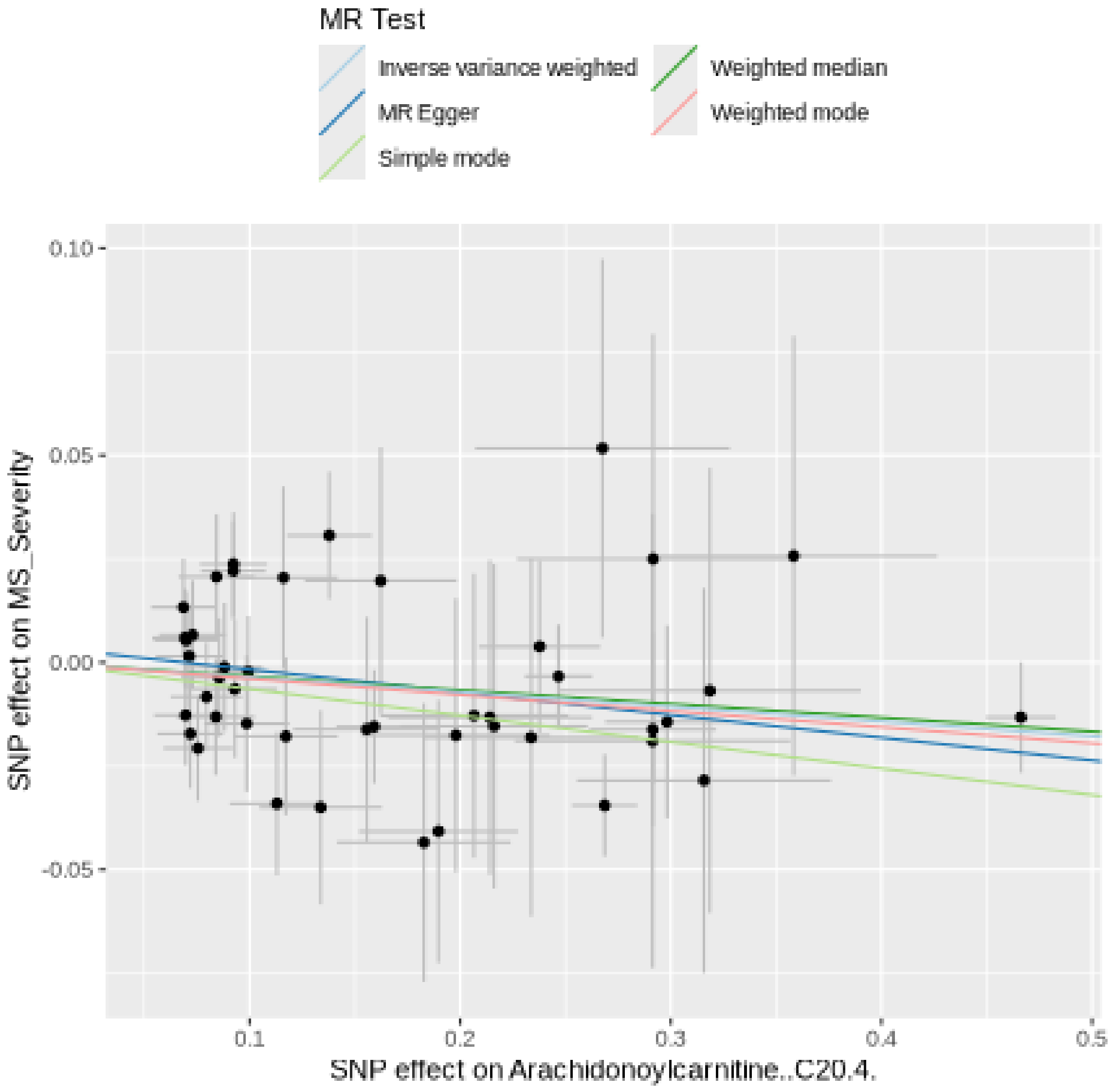

Funnel plot

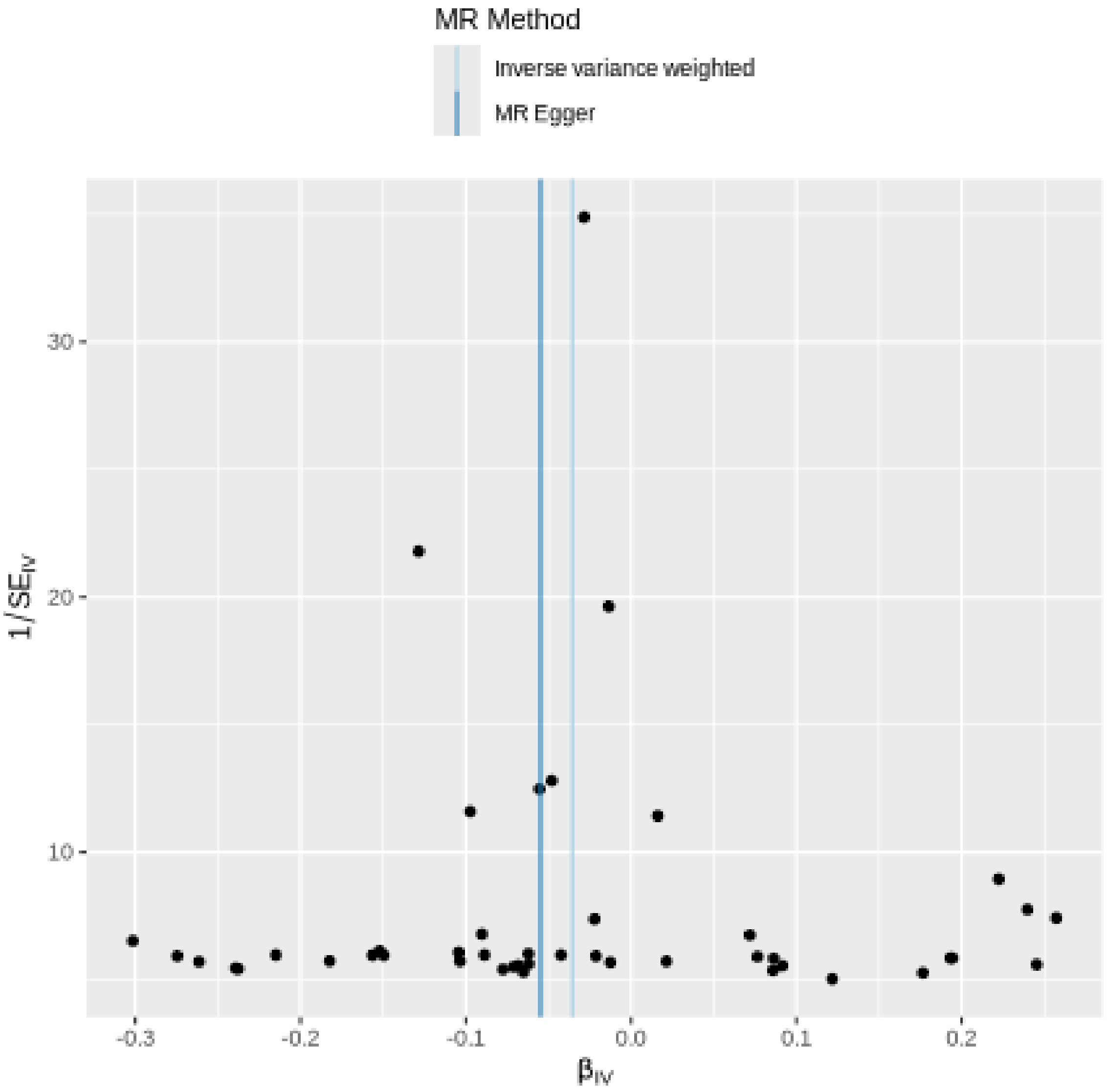

MR Method
Inverse variance weighted
MR Egger
30
20
10
1/SE_IV
-0.3
-0.2
-0.1
0.0
0.1
0.2
β_IV

#### Leave-one-out sensitivity analysis

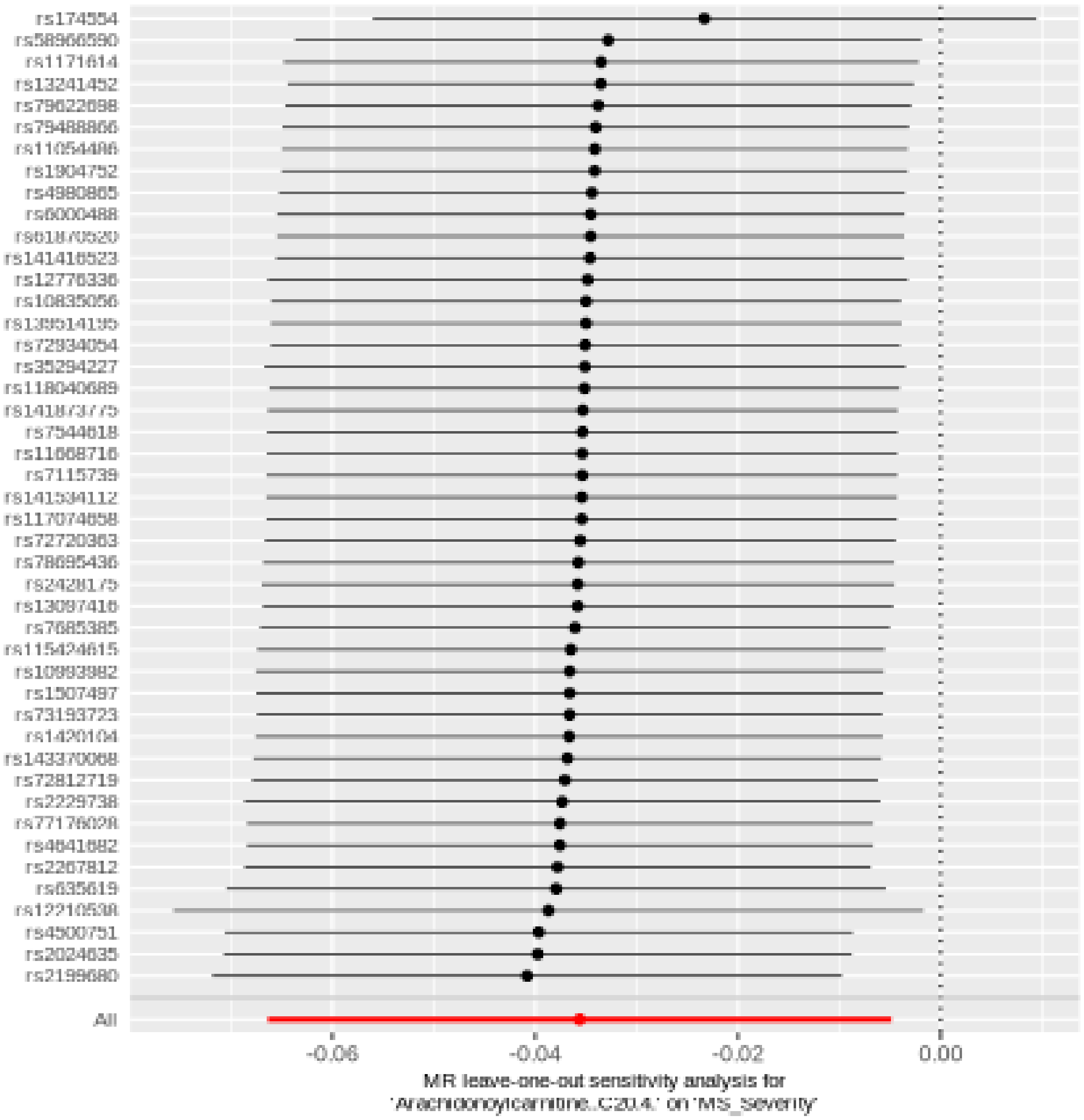

#### Glucuronide of piperine metabolite C17H21NO3 (3)

| method | nsnp | b | se | pval |
|---|---|---|---|---|
| MR Egger | 18 | 0.0200986 | 0.0893830 | 0.8249351 |
| Weighted median | 18 | -0.0841111 | 0.0477906 | 0.0784092 |
| Inverse variance weighted | 18 | -0.0709953 | 0.0341955 | 0.0378791 |
| Simple mode | 18 | -0.1058235 | 0.0822299 | 0.2153694 |
| Weighted mode | 18 | -0.0900247 | 0.0614352 | 0.1610738 |

##### Heterogeneity tests

| method | Q | Q_df | Q_pval |
|---|---|---|---|
| MR Egger | 18.86470 | 16 | 0.2757418 |
| Inverse variance weighted | 20.29623 | 17 | 0.2593589 |

##### Test for directional horizontal pleiotropy

| egger_intercept | se | pval |
|---|---|---|
| -0.0135551 | 0.0123018 | 0.2868134 |

Forest plot of single SNP MR

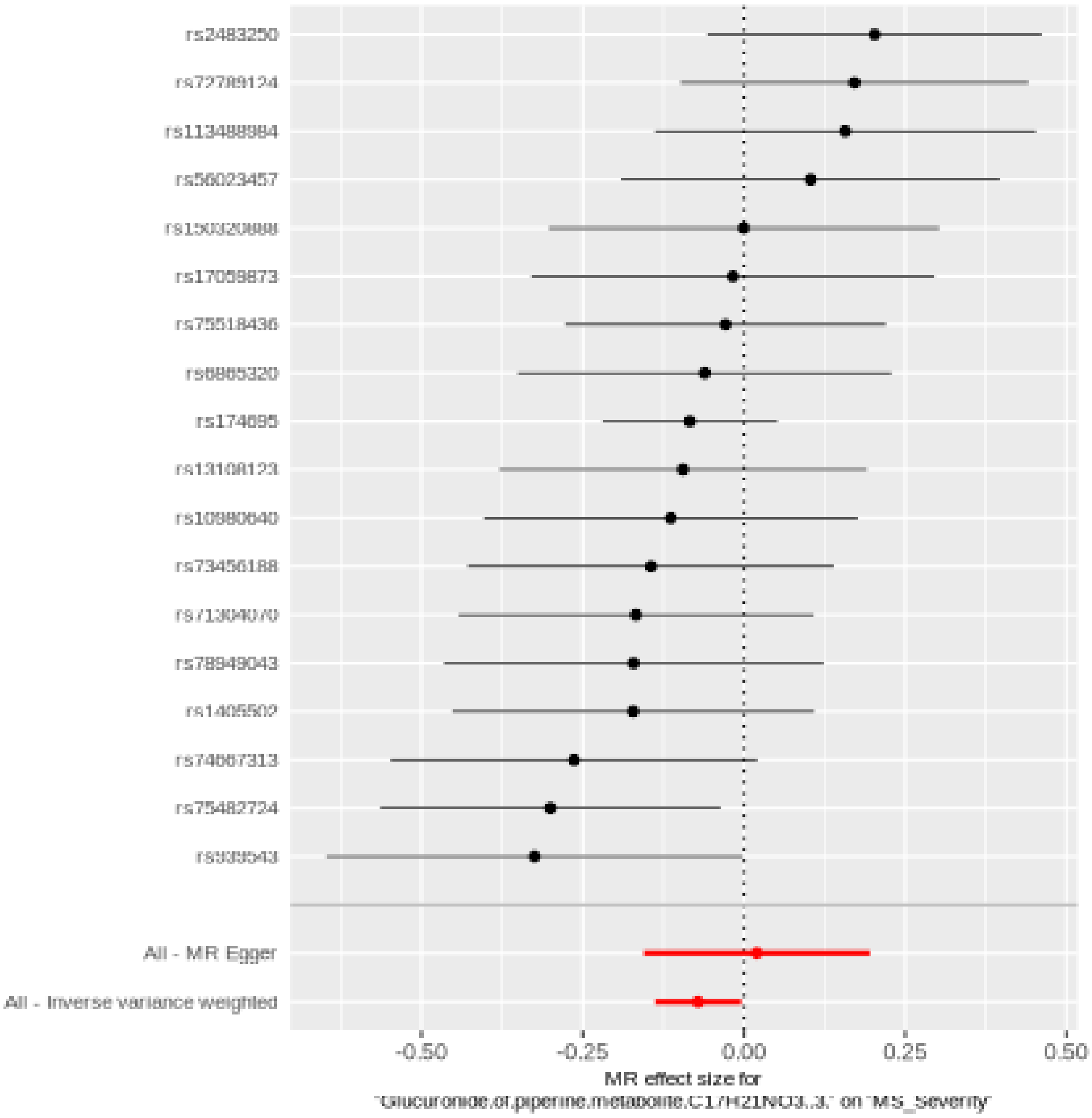

Comparison of results using different MR methods

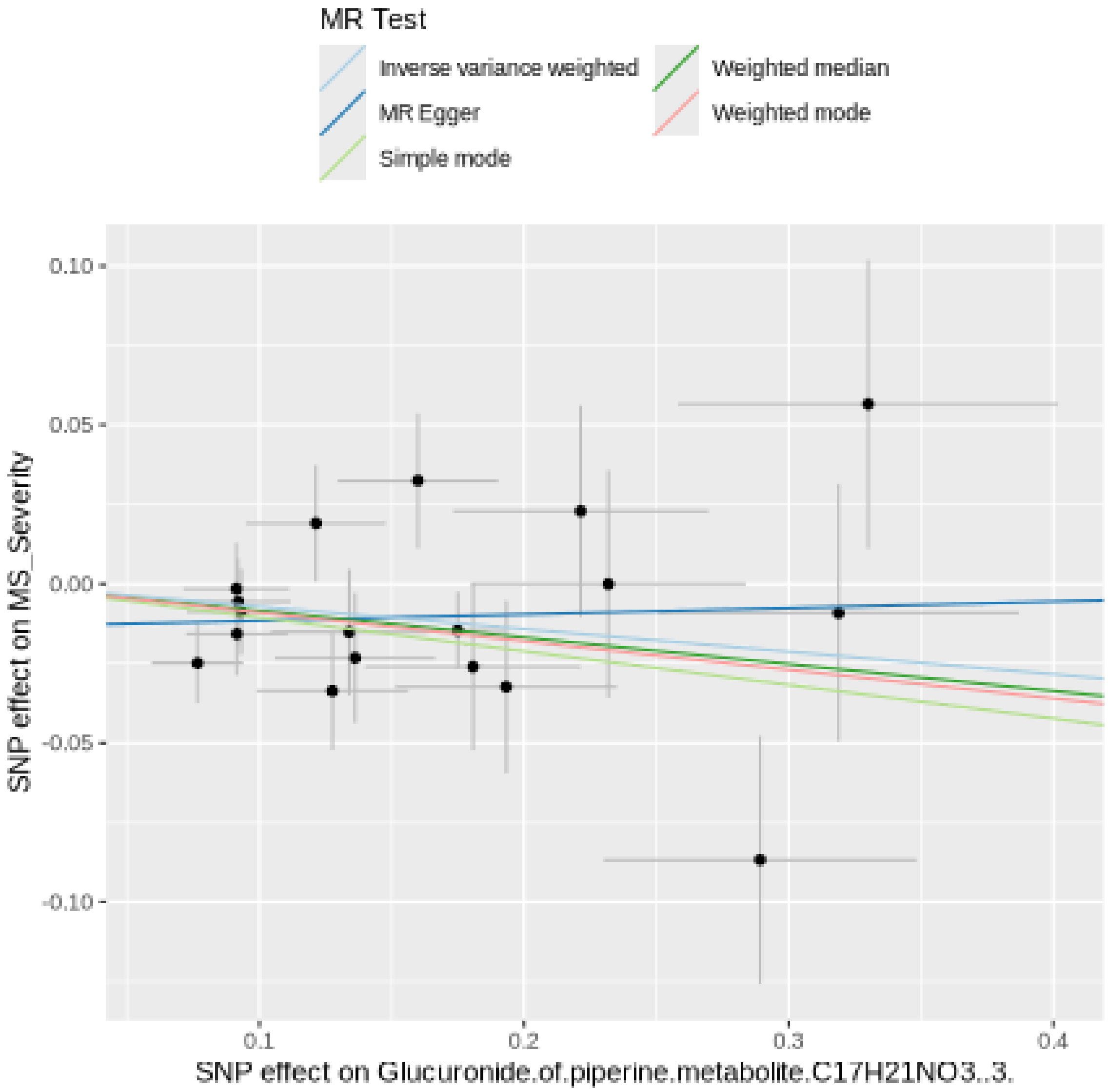

Funnel plot

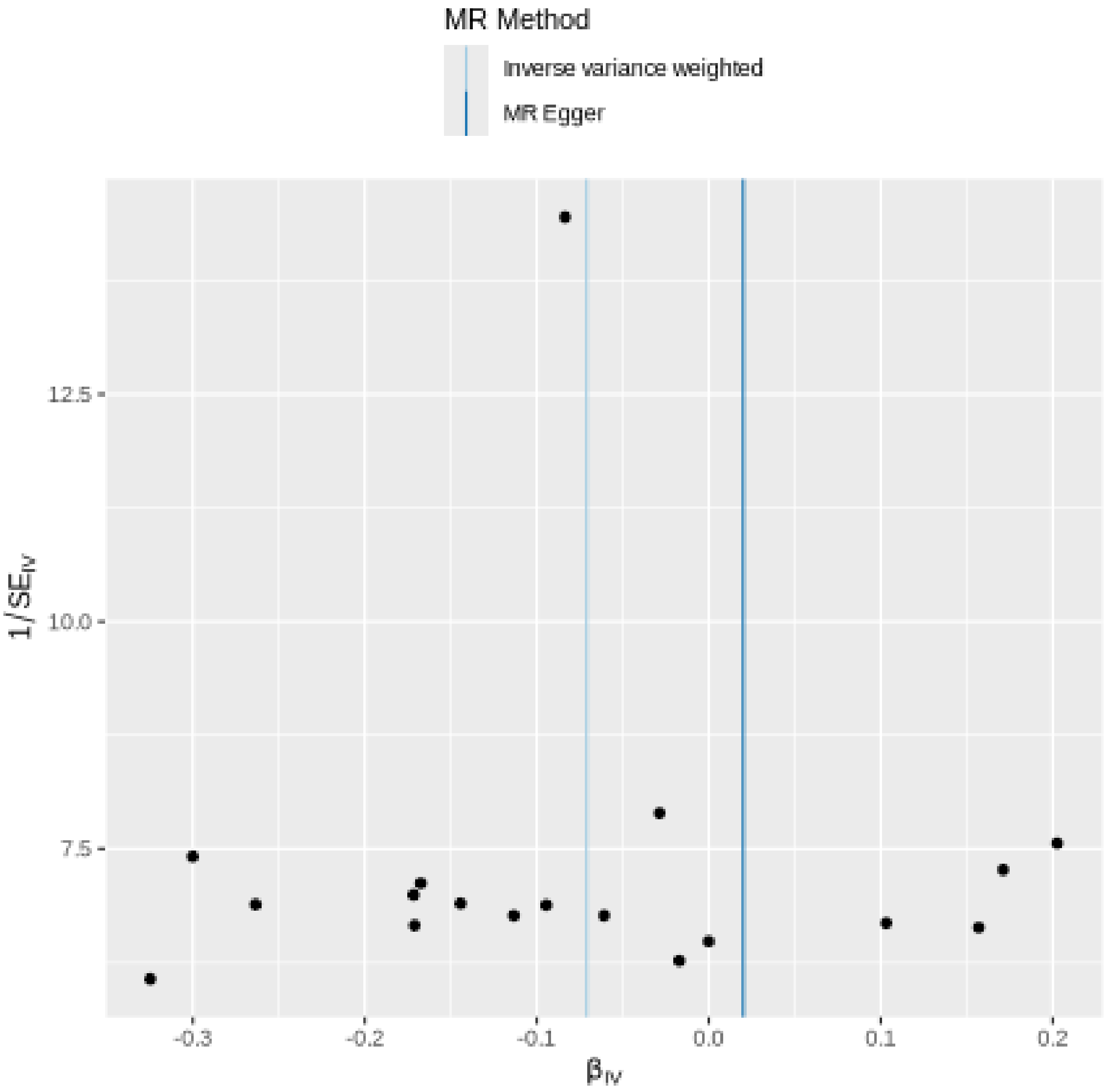

MR Method
Inverse variance weighted
MR Egger
12.5
10.0
7.5
1/SE_IV
-0.3
-0.2
-0.1
0.0
0.1
0.2
β_IV

Leave-one-out sensitivity analysis

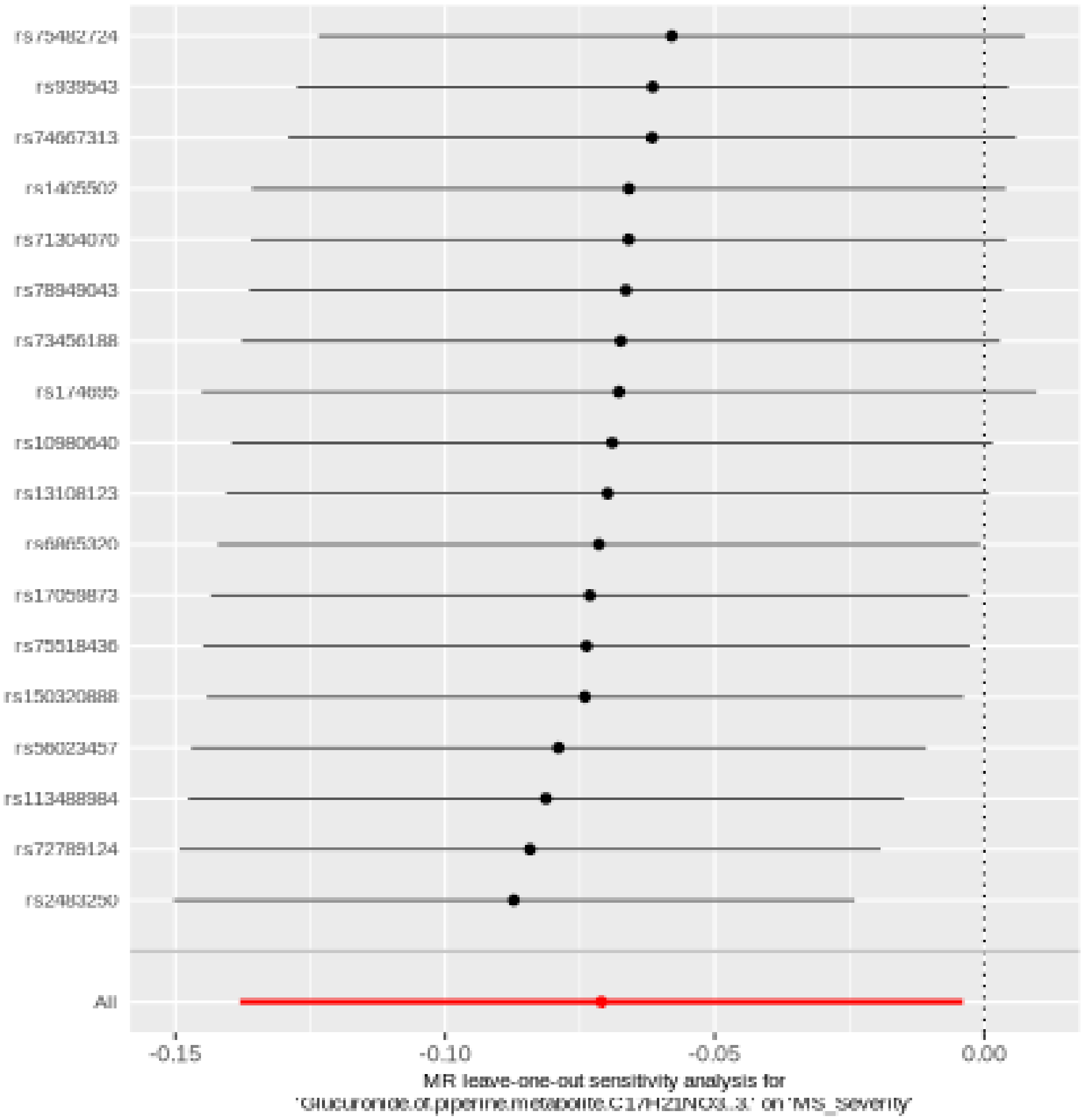

#### Succinoyltaurine

| method | nsnp | b | se | pval |
|---|---|---|---|---|
| MR Egger | 31 | -0.0363422 | 0.0348166 | 0.3051934 |
| Weighted median | 31 | -0.0245594 | 0.0304766 | 0.4203320 |
| Inverse variance weighted | 31 | -0.0404073 | 0.0193179 | 0.0364646 |
| Simple mode | 31 | -0.0317017 | 0.0600477 | 0.6014235 |
| Weighted mode | 31 | -0.0200868 | 0.0302876 | 0.5122652 |

Heterogeneity tests

| method | Q | Q_df | Q_pval |
|---|---|---|---|
| MR Egger | 25.27311 | 29 | 0.6640011 |
| Inverse variance weighted | 25.29281 | 30 | 0.7106808 |

Test for directional horizontal pleiotropy

| egger_intercept | se | pval |
|---|---|---|
| -0.0008356 | 0.0059536 | 0.8893585 |

#### Forest plot of single SNP MR

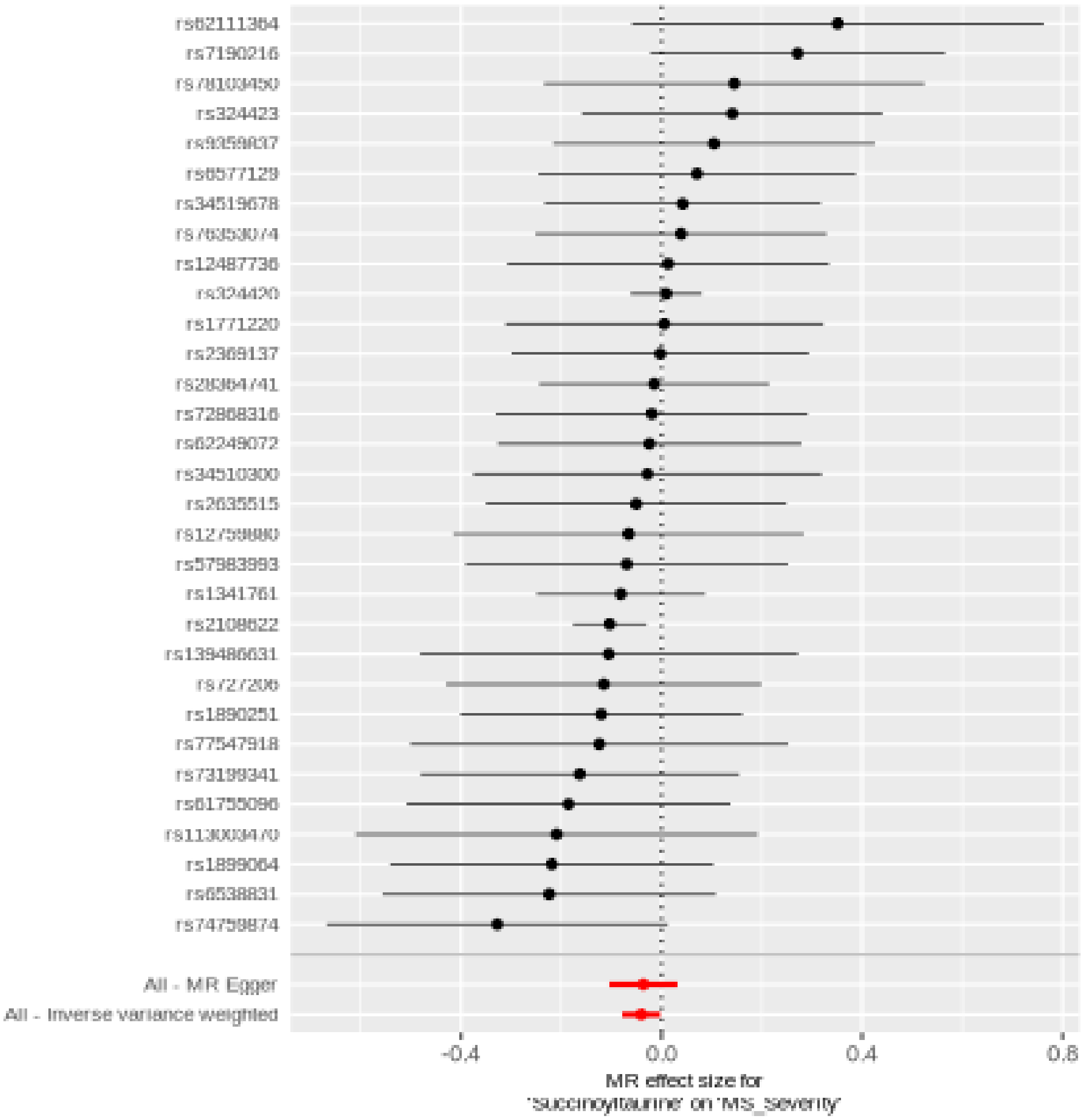

Comparison of results using different MR methods

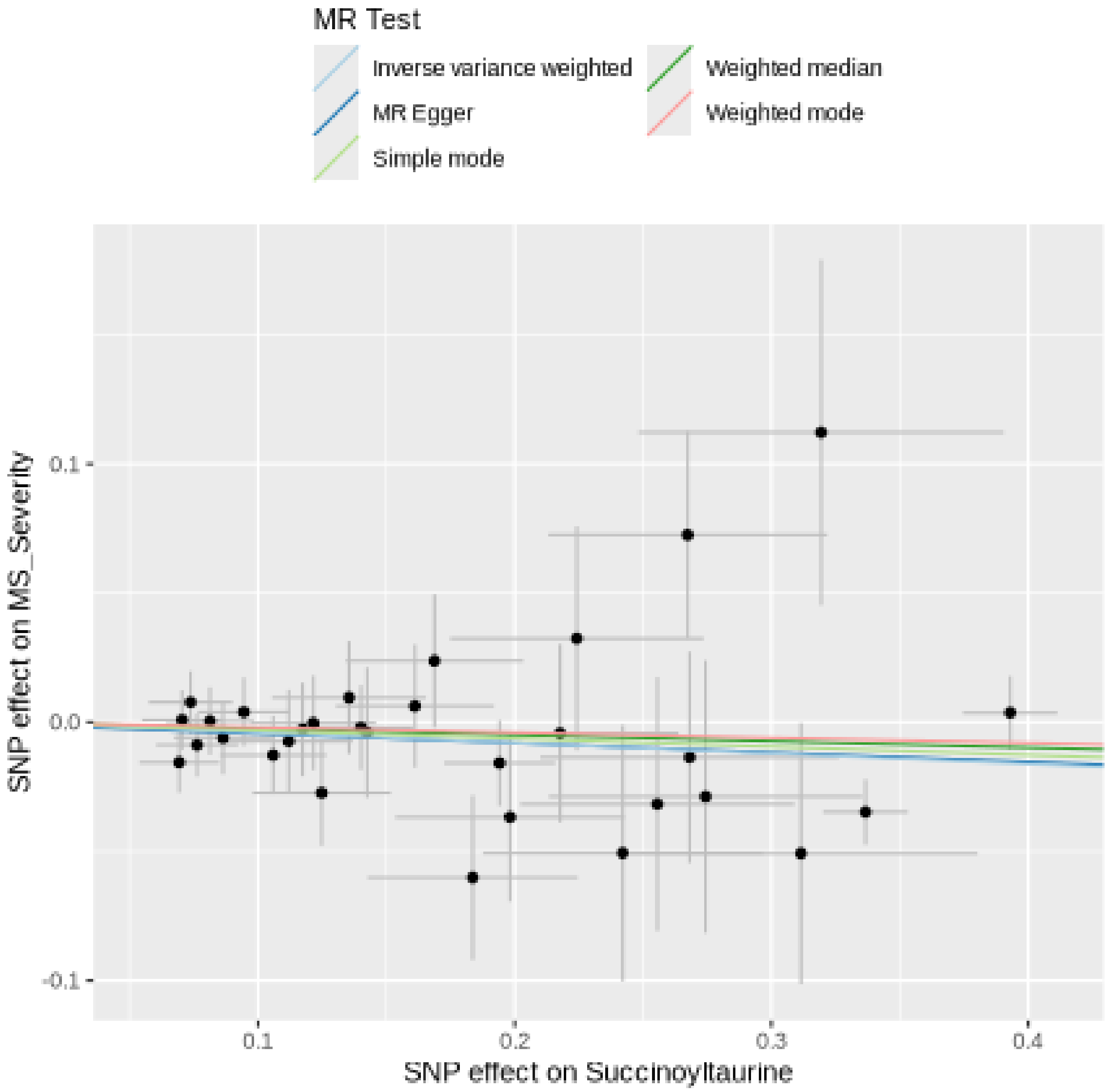

Funnel plot

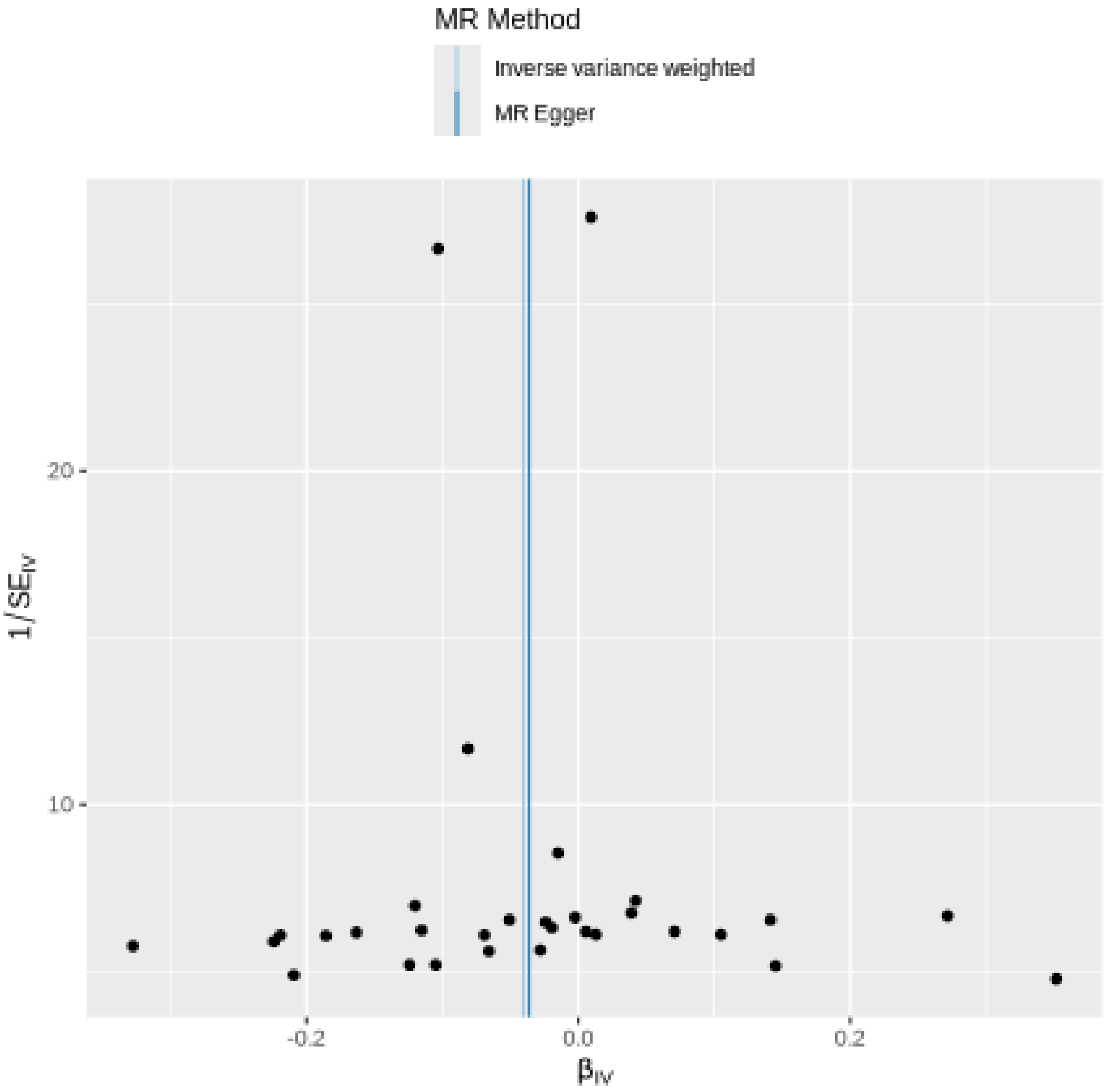
MR Method
Inverse variance weighted
MR Egger
20
10
-0.2
0.0
0.2
1/SE_IV
β_IV

Leave-one-out sensitivity analysis

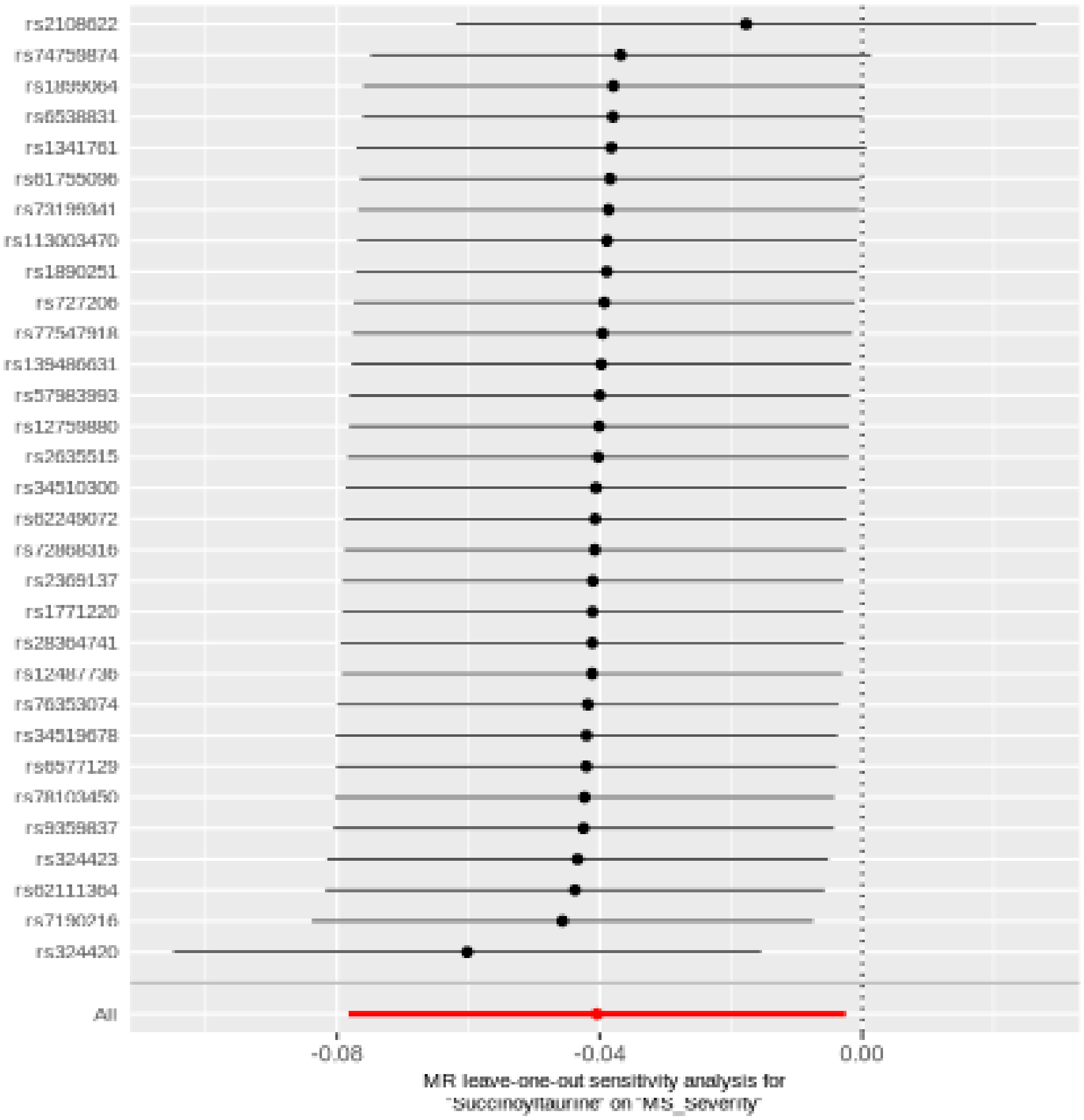

#### Cis 3,4-methyleneheptanoate

| method | nsnp | b | se | pval |
|---|---|---|---|---|
| MR Egger | 25 | -0.0529681 | 0.0753168 | 0.4889450 |
| Weighted median | 25 | -0.0360952 | 0.0437913 | 0.4097949 |
| Inverse variance weighted | 25 | -0.0668963 | 0.0319726 | 0.0364114 |
| Simple mode | 25 | -0.0379794 | 0.0816718 | 0.6461052 |
| Weighted mode | 25 | -0.0280466 | 0.0769314 | 0.7186283 |

##### Heterogeneity tests

| method | Q | Q_df | Q_pval |
|---|---|---|---|
| MR Egger | 21.64583 | 23 | 0.5417099 |
| Inverse variance weighted | 21.68754 | 24 | 0.5979100 |

##### Test for directional horizontal pleiotropy

| egger_intercept | se | pval |
|---|---|---|
| -0.0017574 | 0.0086046 | 0.8399592 |

#### Forest plot of single SNP MR

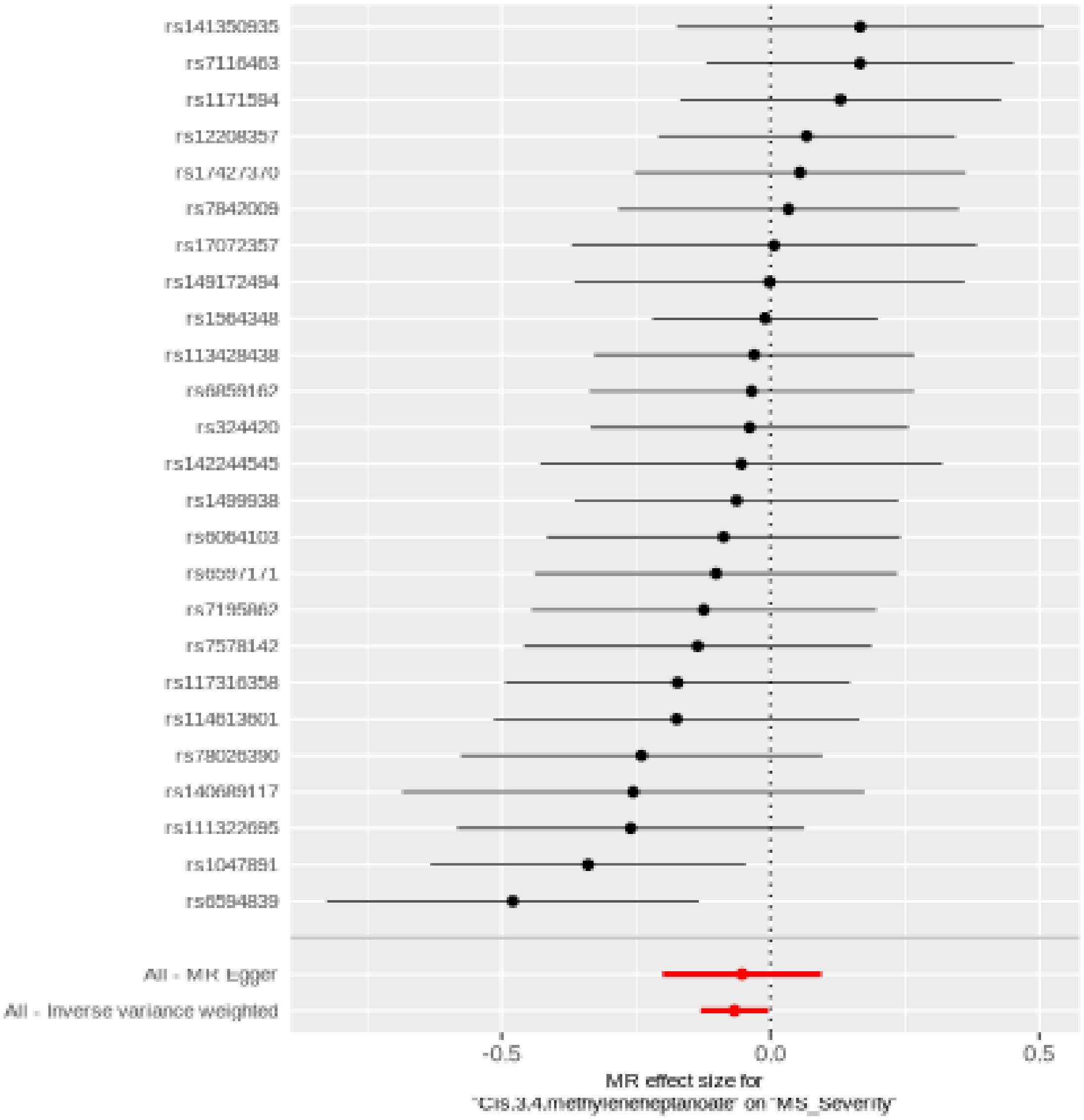

Comparison of results using different MR methods

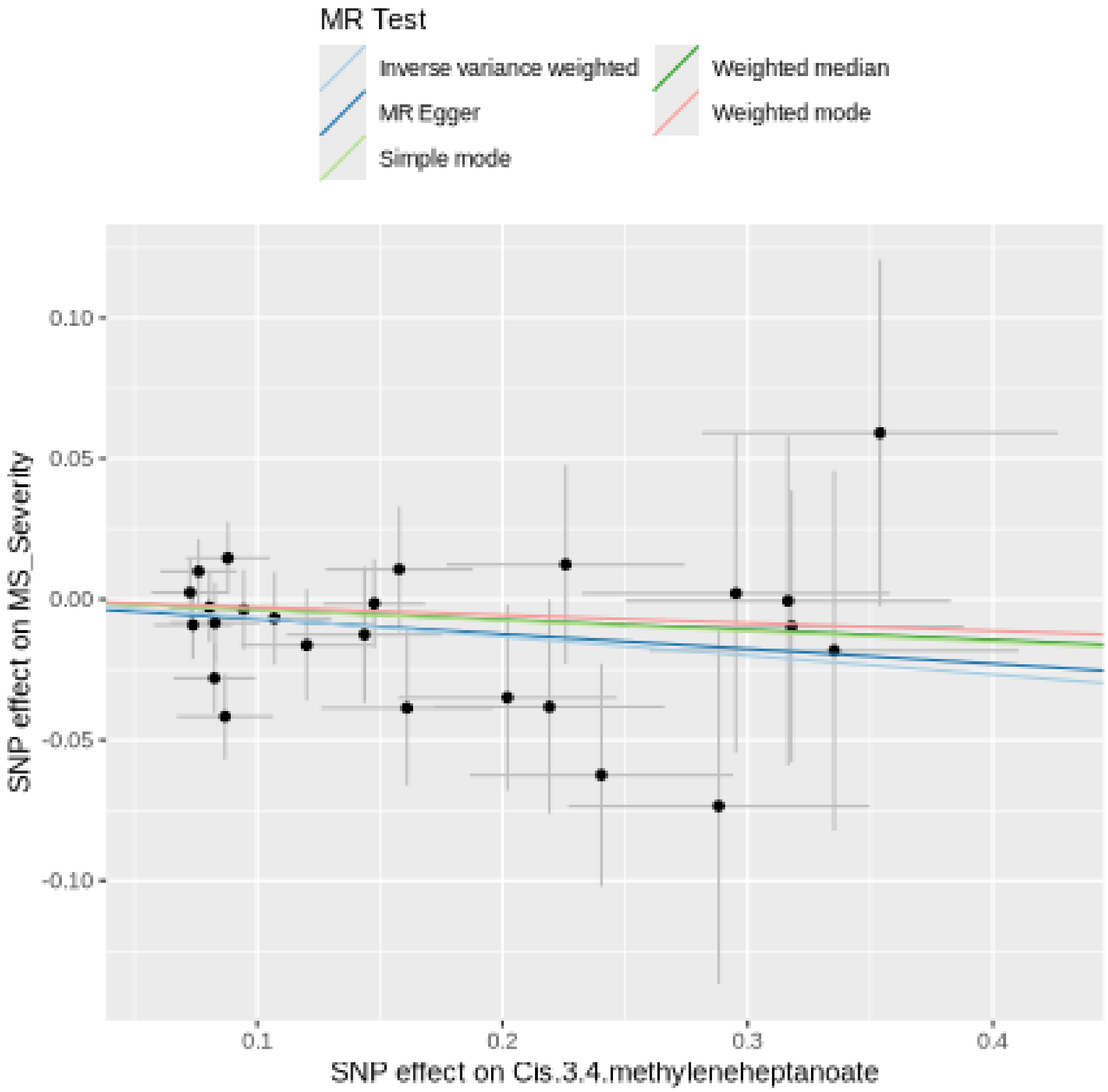

Funnel plot

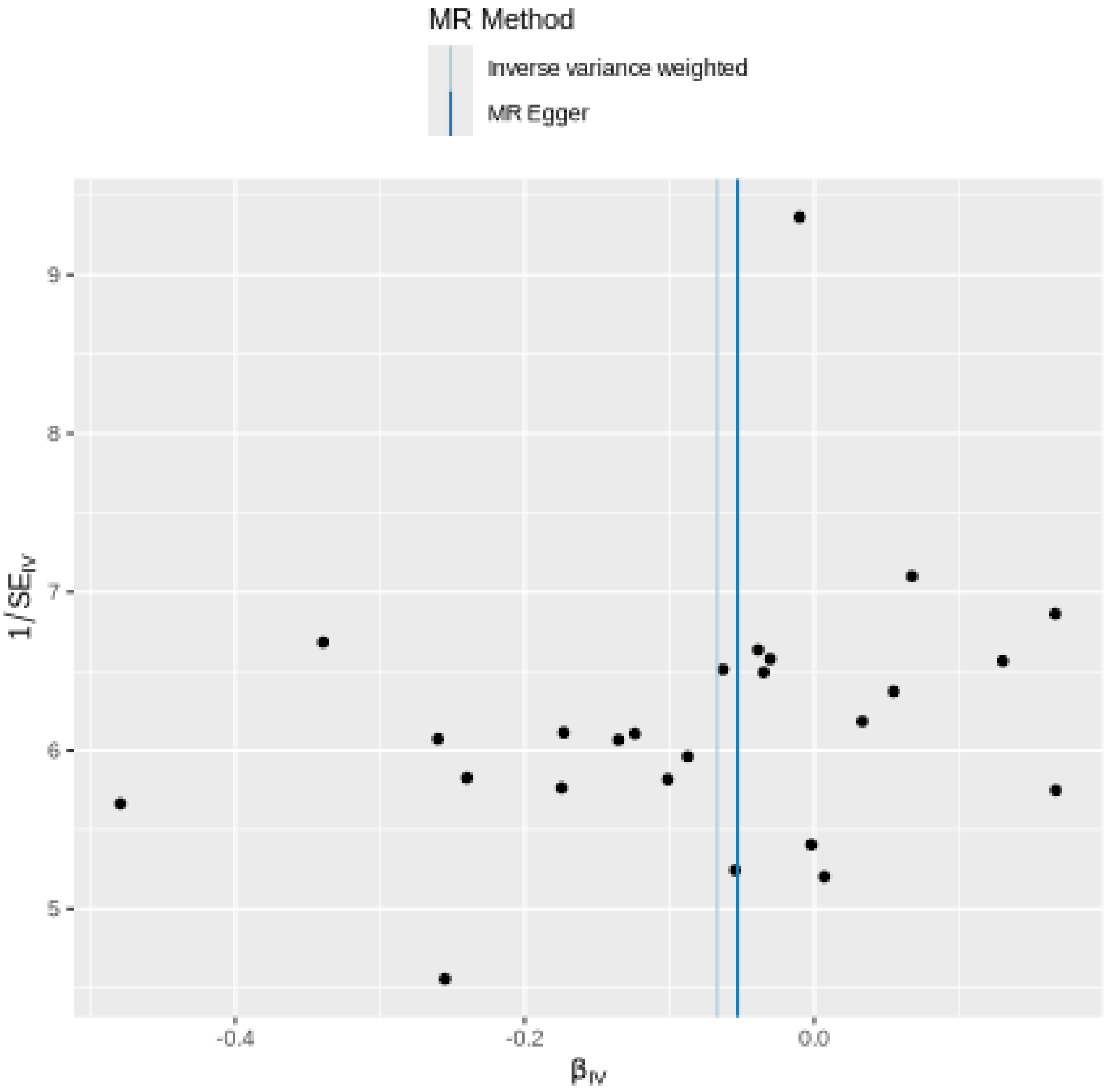
MR Method
Inverse variance weighted
MR Egger
9
8
7
6
5
1/SE$_{IV}$
-0.4
-0.2
0.0
$\beta_{IV}$

#### Leave-one-out sensitivity analysis

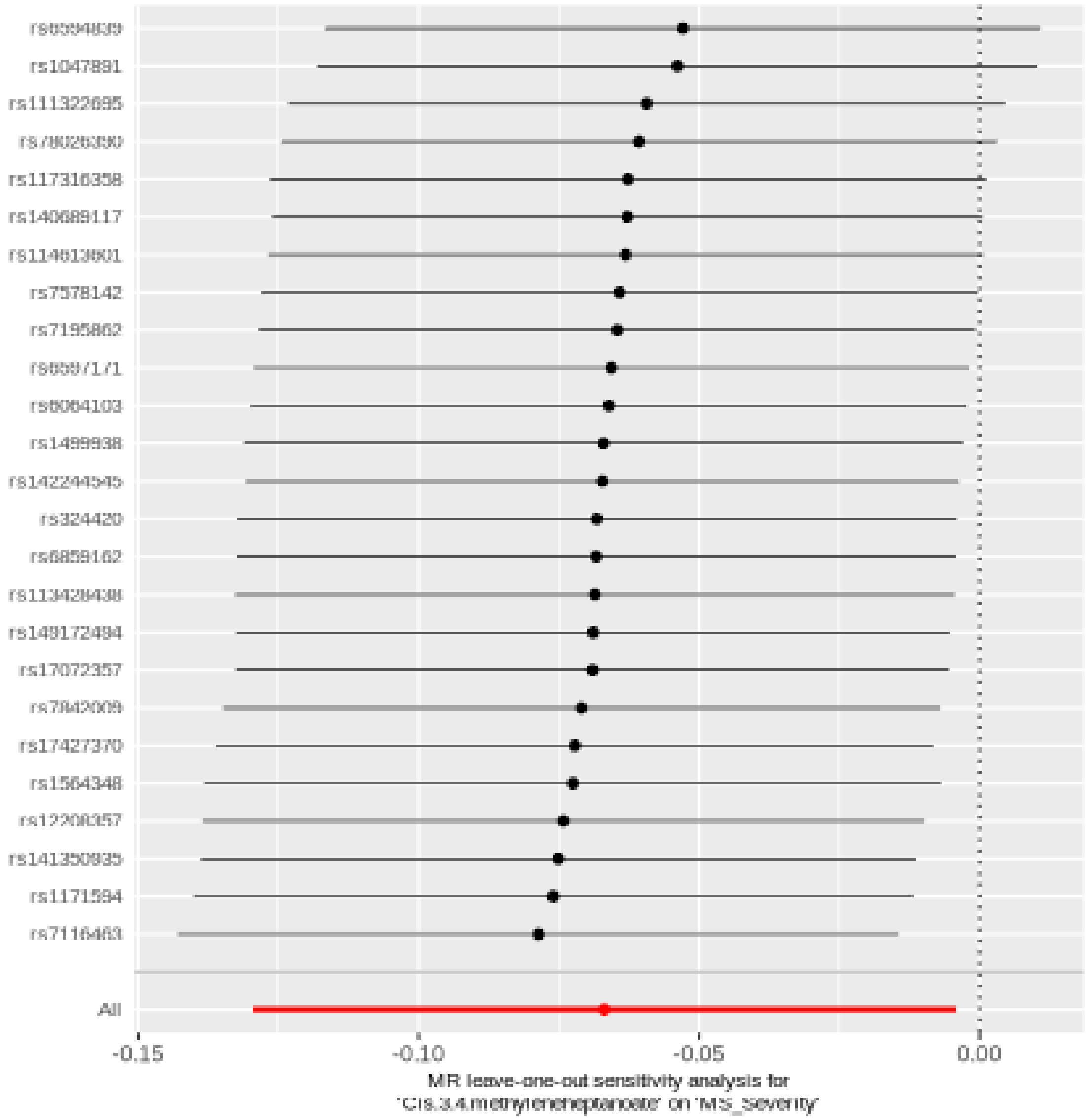

#### Cis-3,4-methyleneheptanoylglycine

| method | nsnp | b | se | pval |
|---|---|---|---|---|
| MR Egger | 28 | 0.0752859 | 0.0397195 | 0.0692033 |
| Weighted median | 28 | 0.0725627 | 0.0299942 | 0.0155538 |
| Inverse variance weighted | 28 | 0.0767337 | 0.0214965 | 0.0003575 |
| Simple mode | 28 | 0.0922881 | 0.0556304 | 0.1086993 |
| Weighted mode | 28 | 0.0746951 | 0.0291366 | 0.0162438 |

##### Heterogeneity tests

| method | Q | Q_df | Q_pval |
|---|---|---|---|
| MR Egger | 21.68661 | 26 | 0.7057151 |
| Inverse variance weighted | 21.68849 | 27 | 0.7530335 |

##### Test for directional horizontal pleiotropy

| egger_intercept | se | pval |
|---|---|---|
| 0.000276 | 0.0063664 | 0.9657536 |

Forest plot of single SNP MR

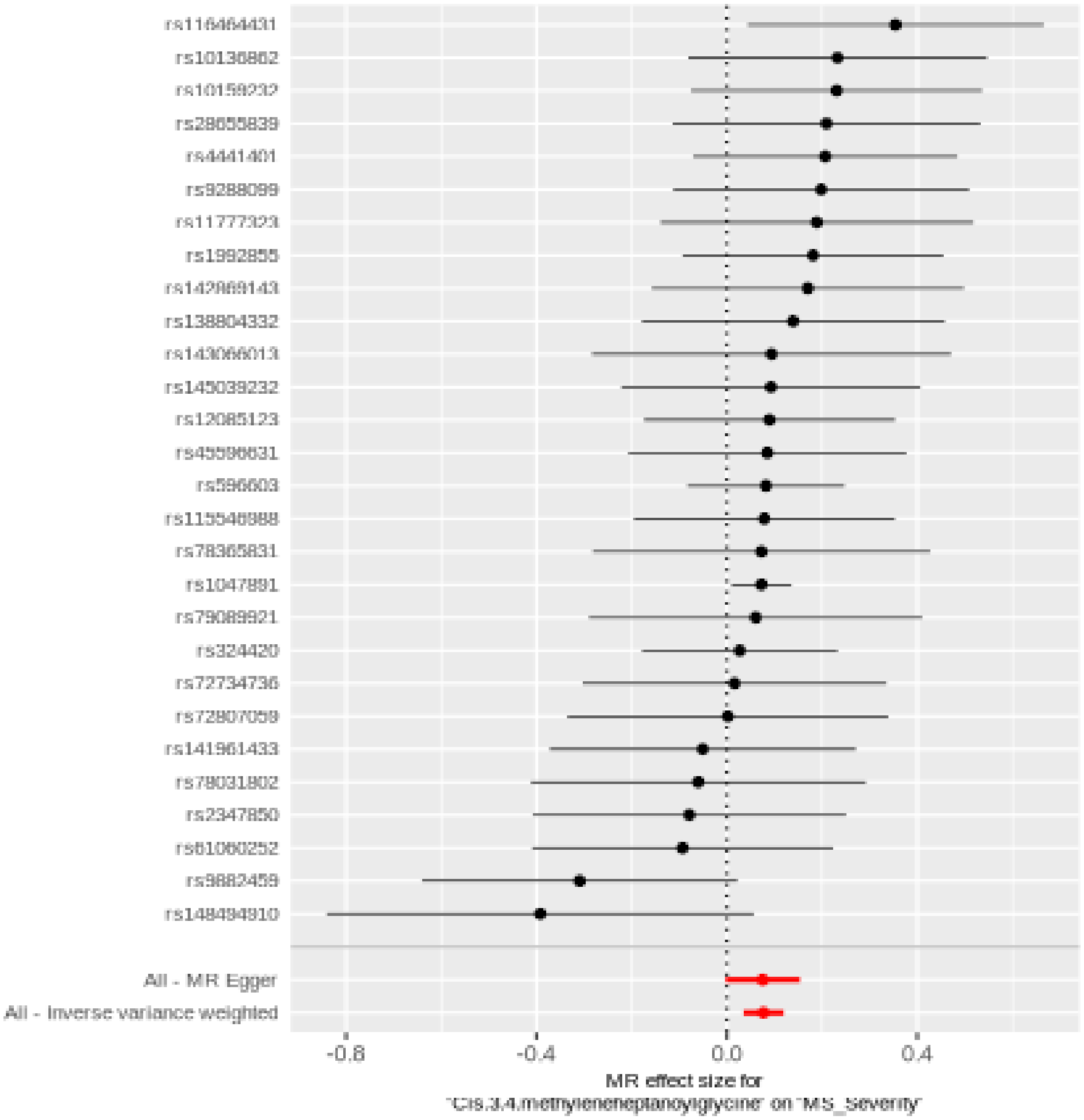

Comparison of results using different MR methods

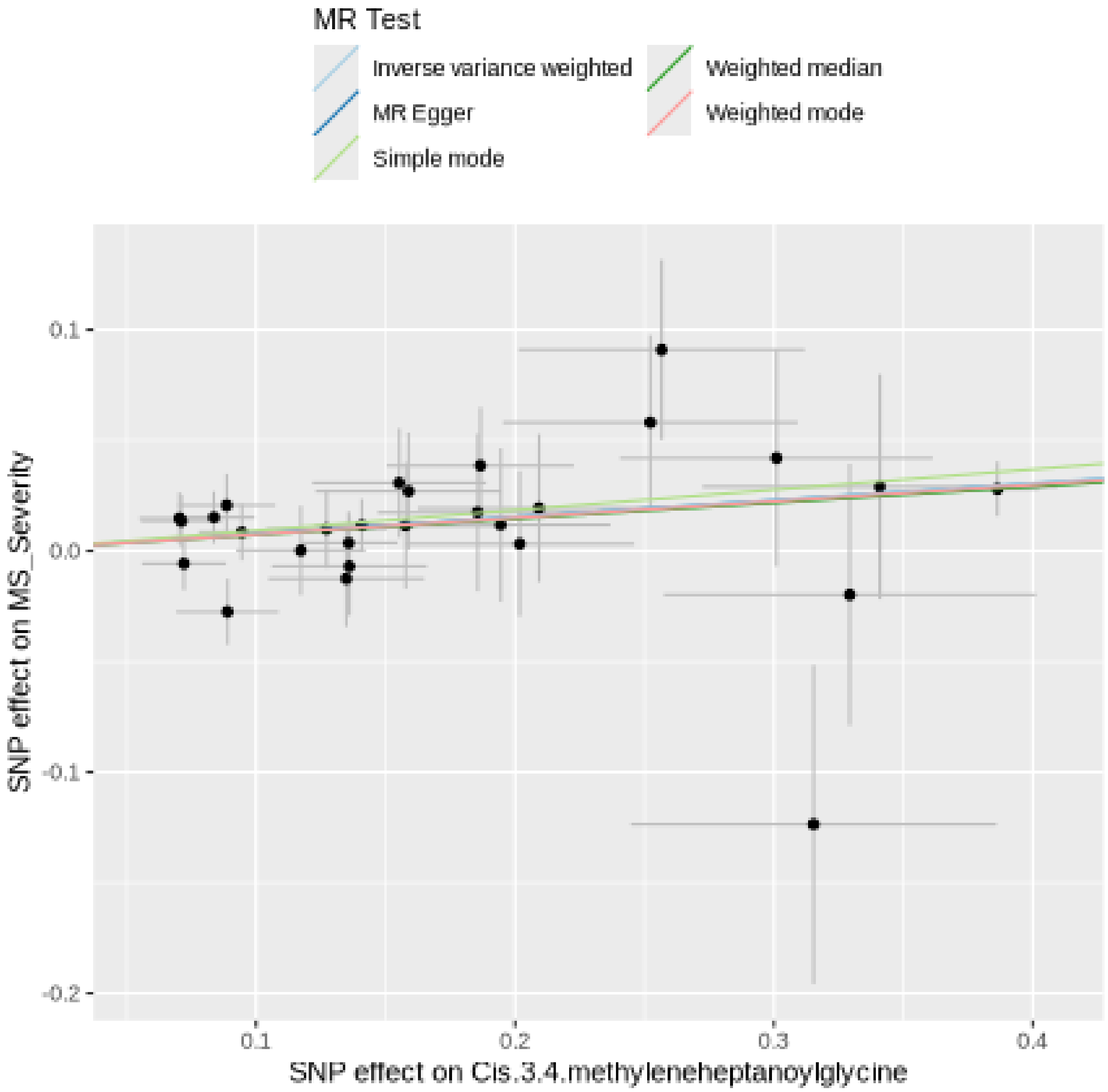

Funnel plot

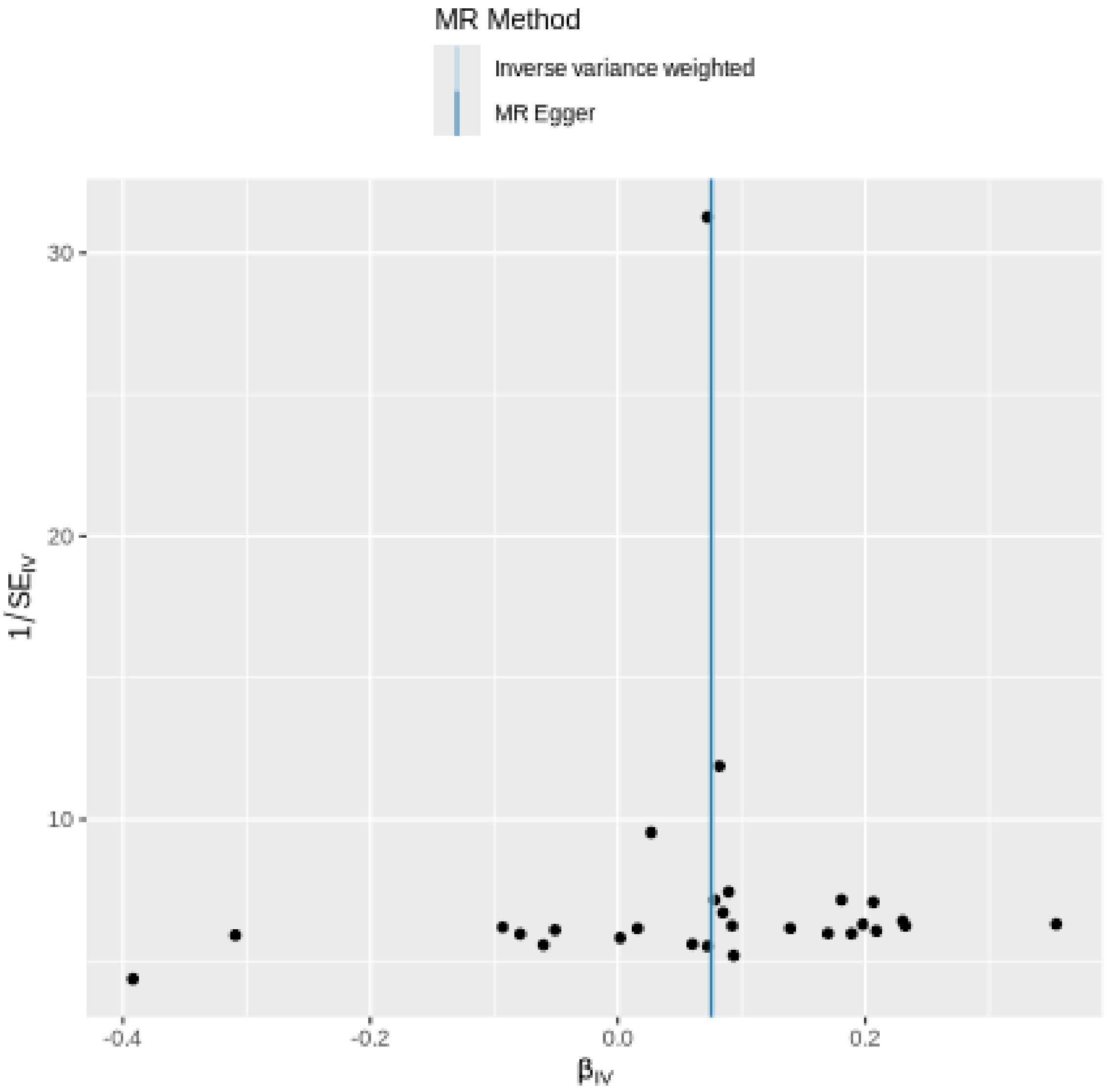

MR Method
Inverse variance weighted
MR Egger
30
20
10
1/SE_IV
-0.4
-0.2
0.0
0.2
β_IV

#### Leave-one-out sensitivity analysis

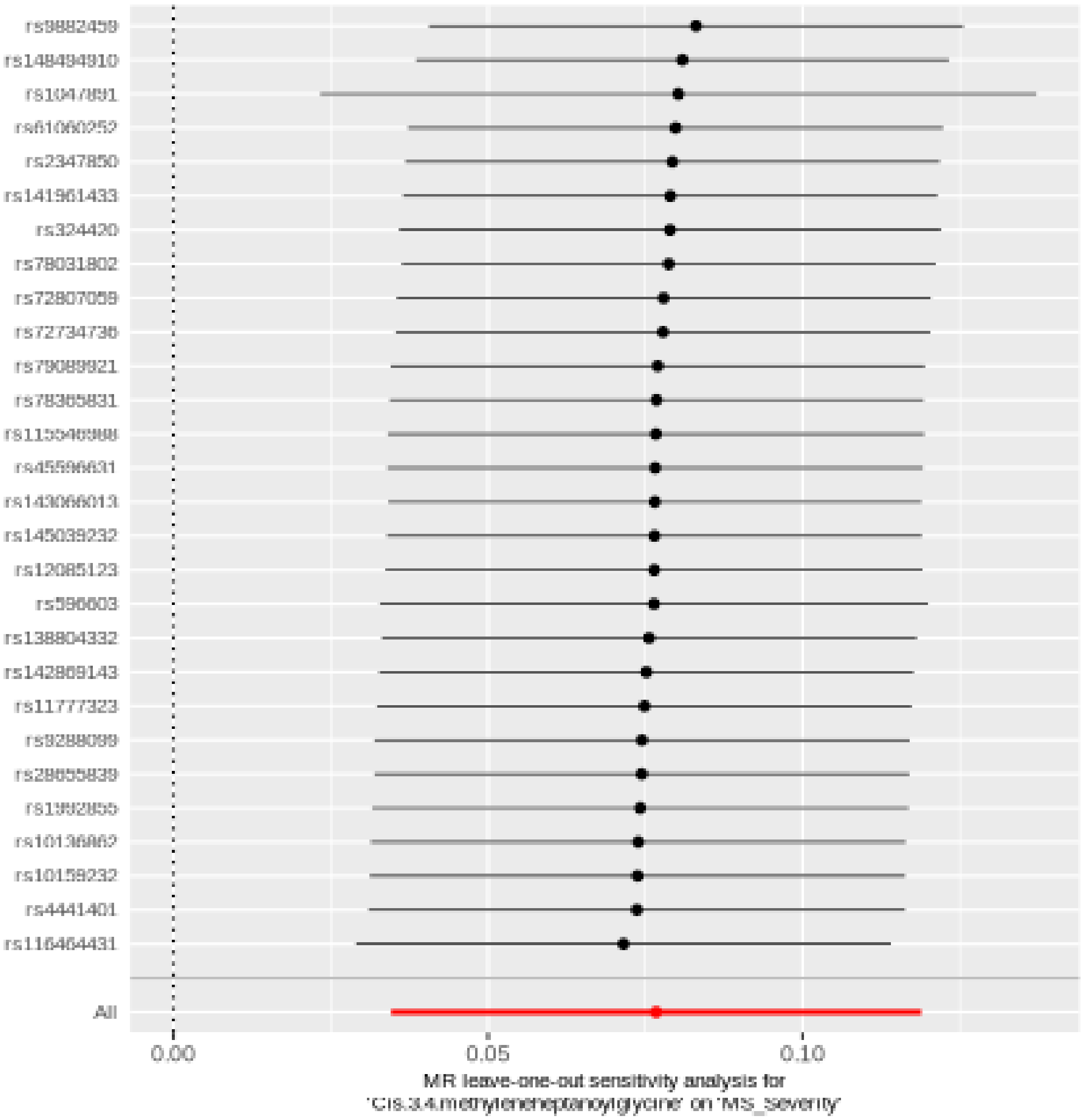

#### Arachidonate (20:4n6)

| method | nsnp | b | se | pval |
|---|---|---|---|---|
| MR Egger | 31 | -0.1085186 | 0.0459187 | 0.0250321 |
| Weighted median | 31 | -0.0940564 | 0.0368575 | 0.0107138 |
| Inverse variance weighted | 31 | -0.0482975 | 0.0240480 | 0.0446031 |
| Simple mode | 31 | 0.0885901 | 0.0894675 | 0.3299993 |
| Weighted mode | 31 | -0.0970377 | 0.0362245 | 0.0118747 |

##### Heterogeneity tests

| method | Q | Q_df | Q_pval |
|---|---|---|---|
| MR Egger | 26.64242 | 29 | 0.5909907 |
| Inverse variance weighted | 29.01239 | 30 | 0.5169458 |

##### Test for directional horizontal pleiotropy

| egger_intercept | se | pval |
|---|---|---|
| 0.0093754 | 0.00609 | 0.1345309 |

#### Forest plot of single SNP MR

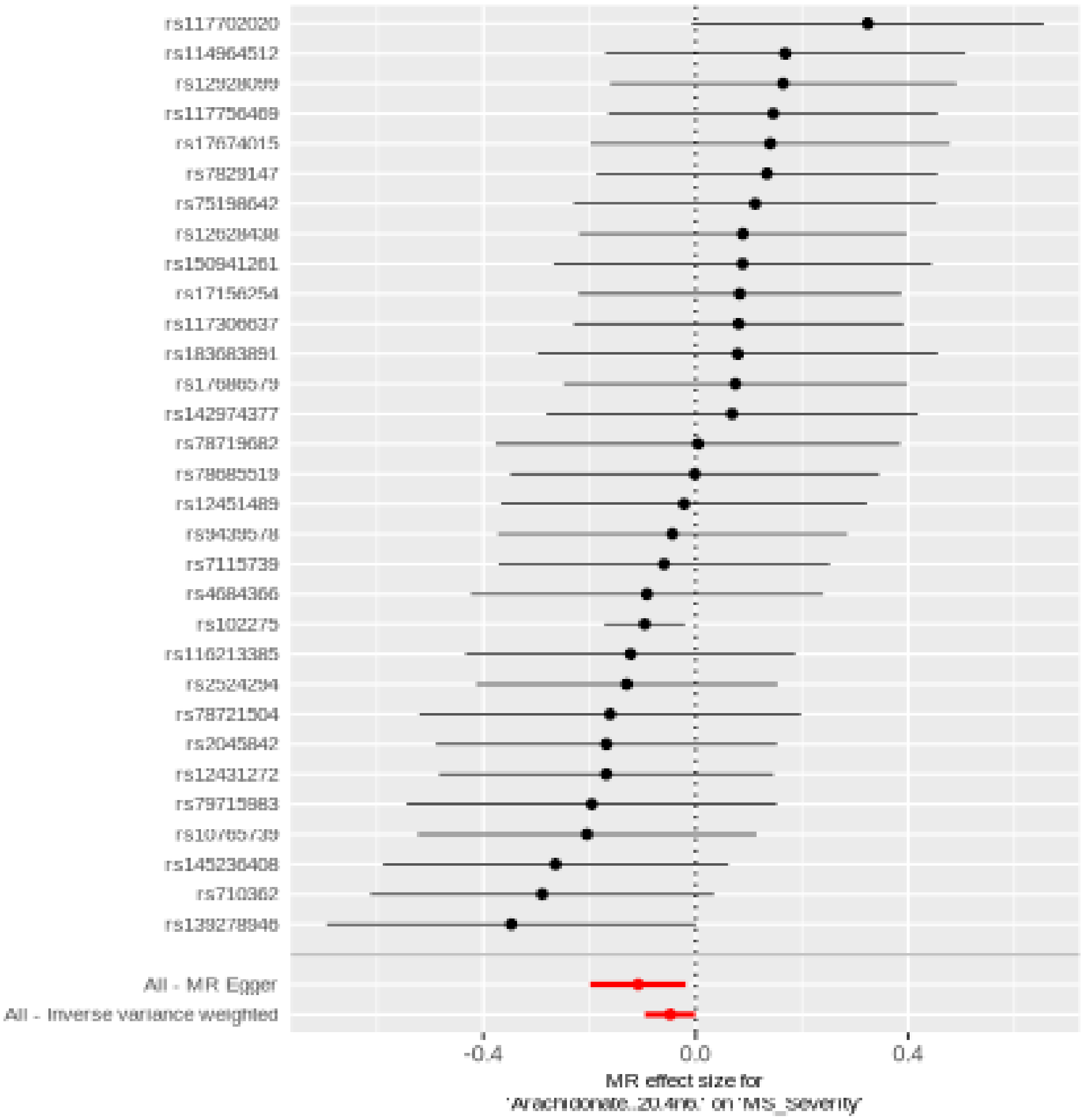

Comparison of results using different MR methods

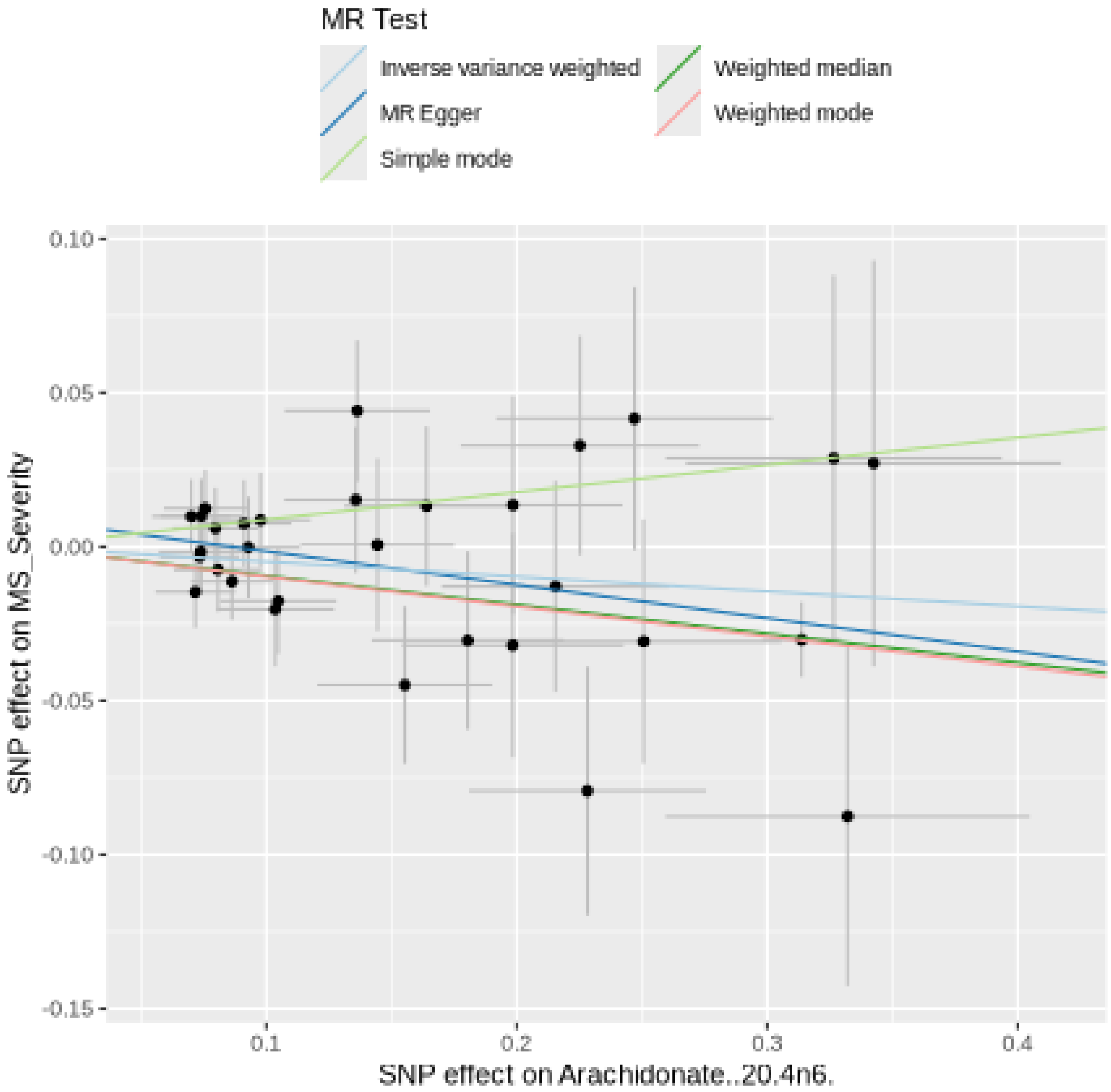

Funnel plot

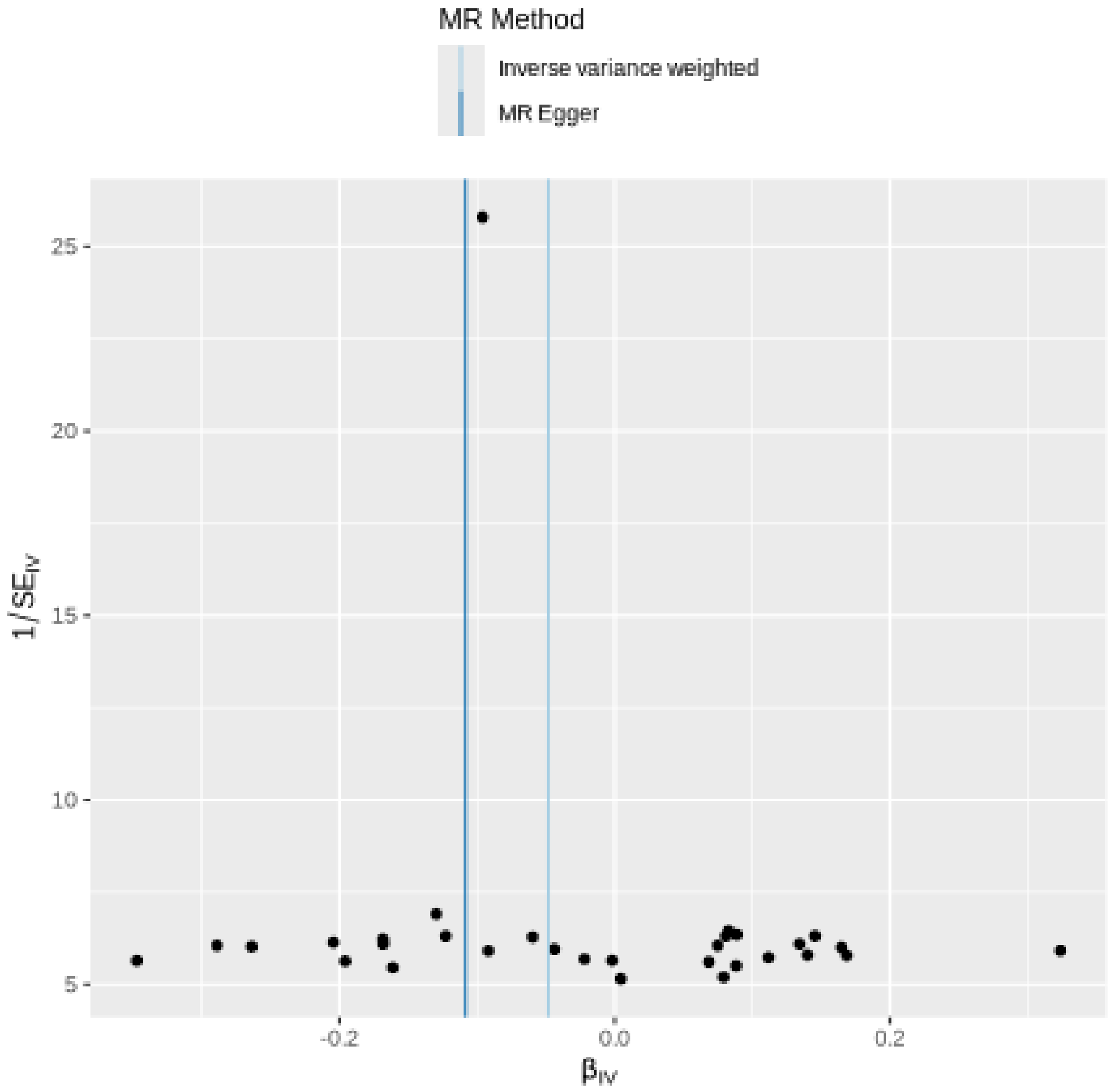

MR Method
Inverse variance weighted
MR Egger
25
20
15
10
5
1/SE$_{IV}$
-0.2
0.0
0.2
β$_{IV}$

Leave-one-out sensitivity analysis

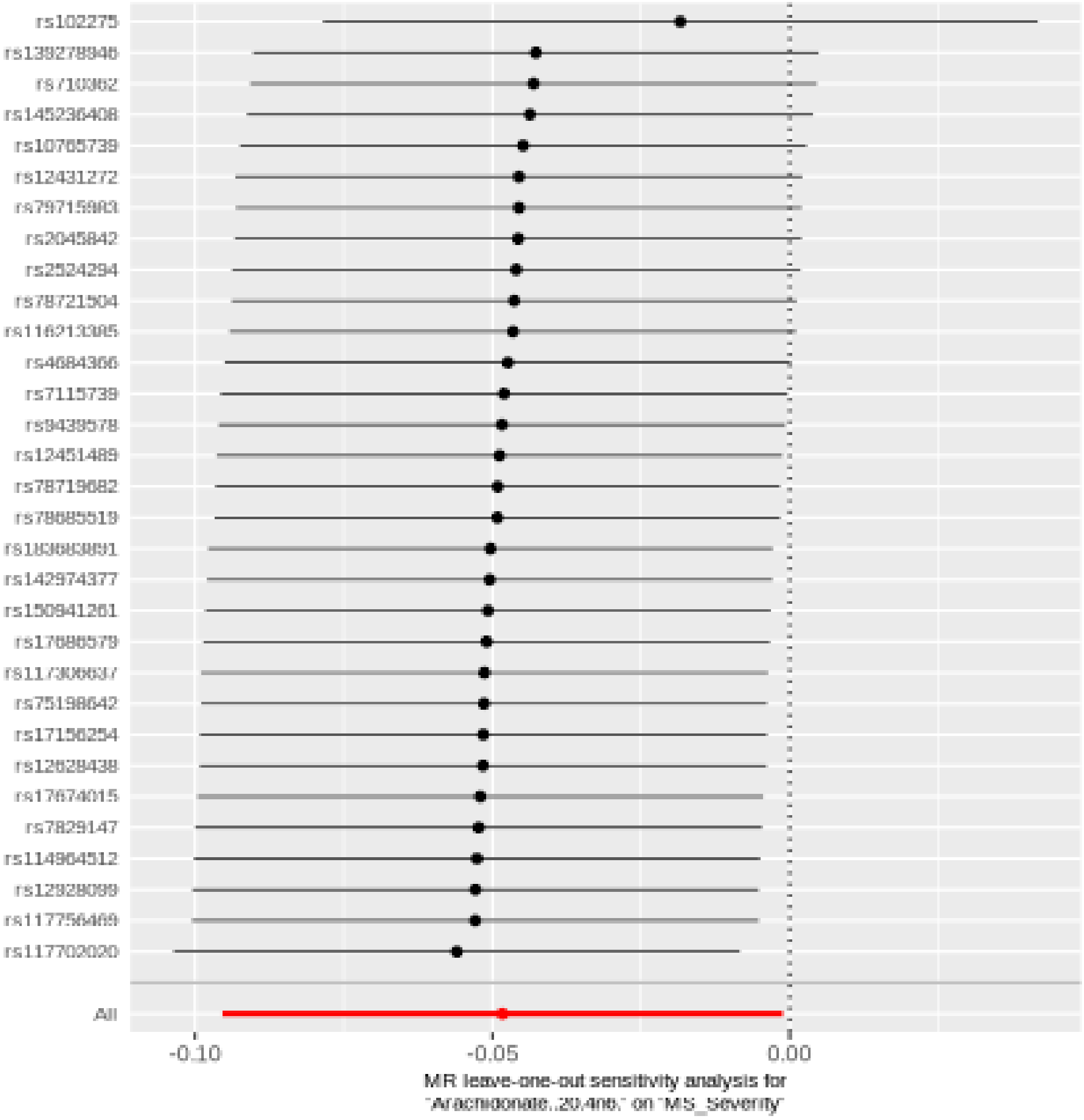

#### Arginine

| method | nsnp | b | se | pval |
|---|---|---|---|---|
| MR Egger | 25 | 0.0865493 | 0.0723350 | 0.2436878 |
| Weighted median | 25 | 0.0679985 | 0.0401801 | 0.0905807 |
| Inverse variance weighted | 25 | 0.0560486 | 0.0283587 | 0.0481080 |
| Simple mode | 25 | 0.1285832 | 0.0712016 | 0.0834920 |
| Weighted mode | 25 | 0.1020558 | 0.0584455 | 0.0935710 |

##### Heterogeneity tests

| method | Q | Q_df | Q_pval |
|---|---|---|---|
| MR Egger | 20.53832 | 23 | 0.6092590 |
| Inverse variance weighted | 20.74841 | 24 | 0.6535194 |

##### Test for directional horizontal pleiotropy

| egger_intercept | se | pval |
|---|---|---|
| -0.0043549 | 0.0095012 | 0.6510011 |

#### Forest plot of single SNP MR

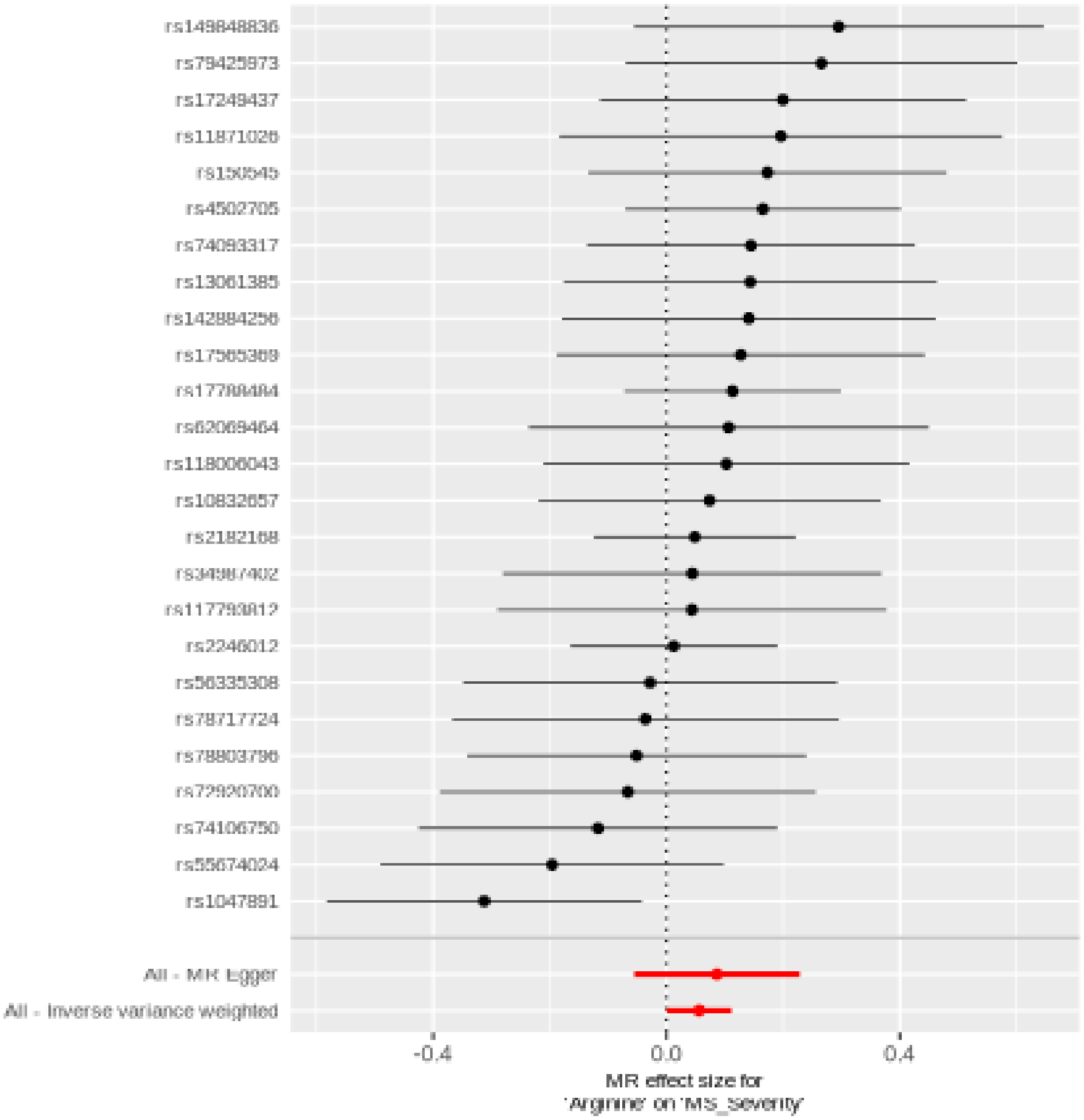

Comparison of results using different MR methods

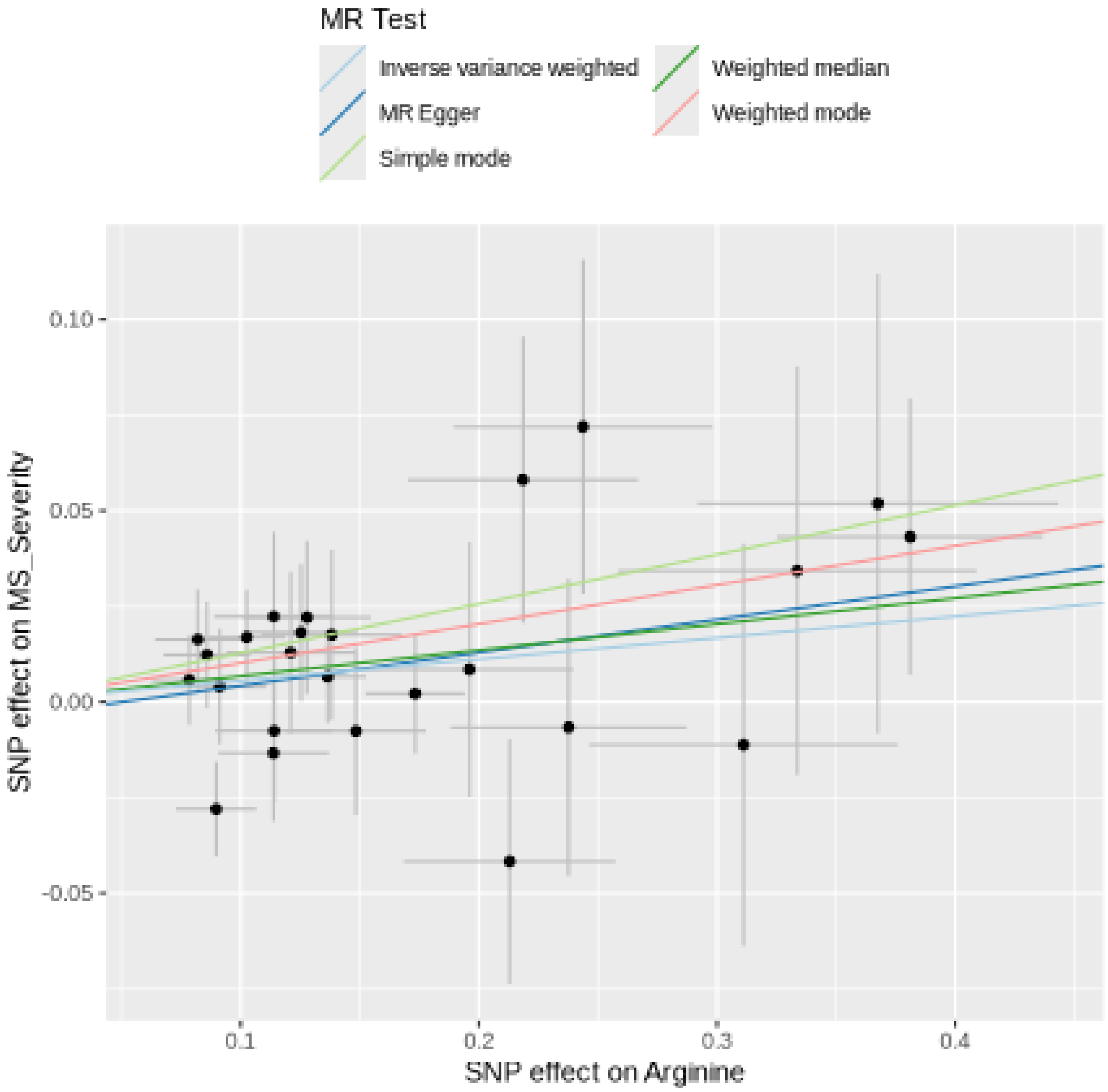

Funnel plot

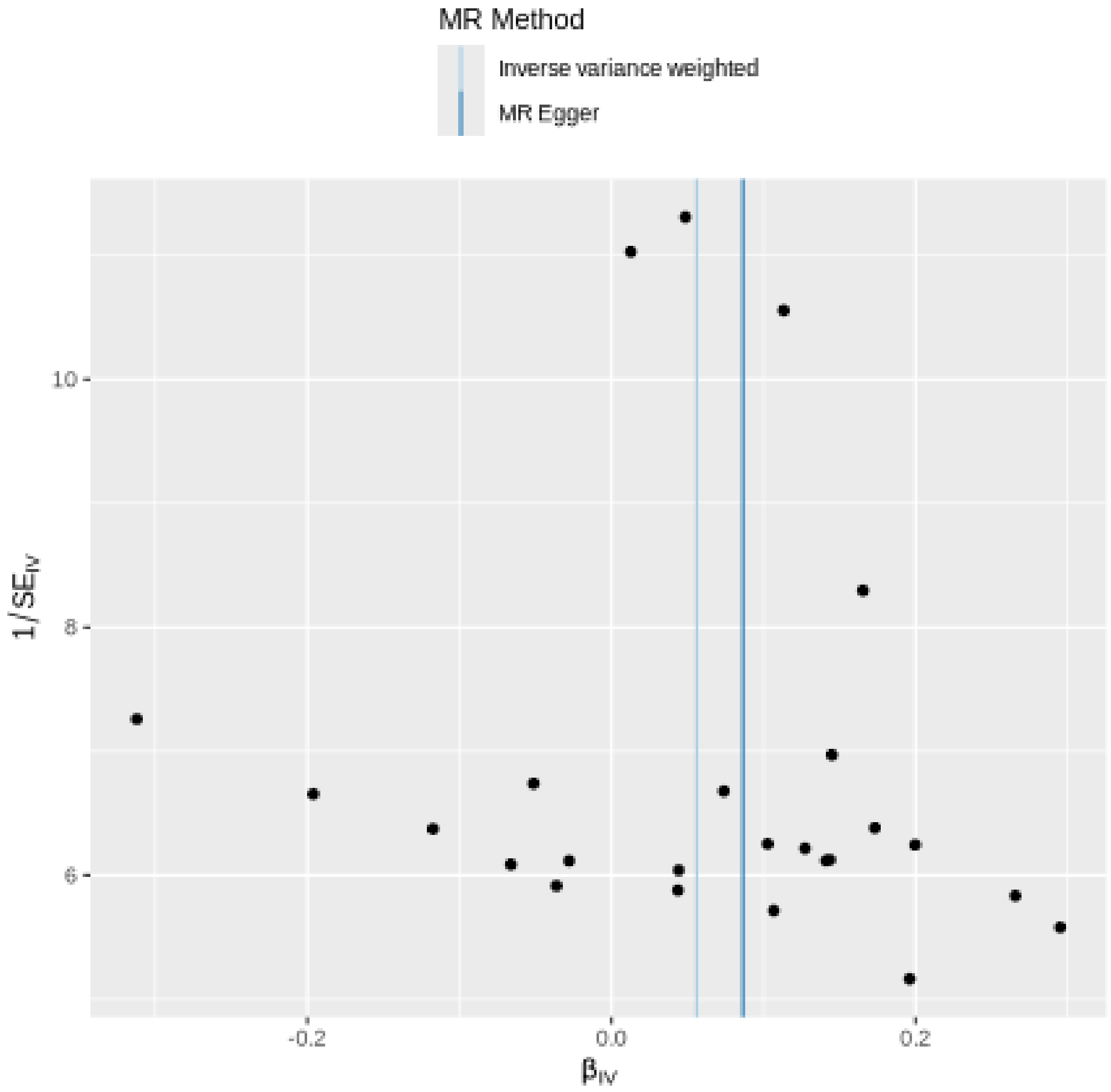

MR Method
Inverse variance weighted
MR Egger
10
8
6
1/SE$_{IV}$
-0.2
0.0
0.2
β$_{IV}$

#### Leave-one-out sensitivity analysis

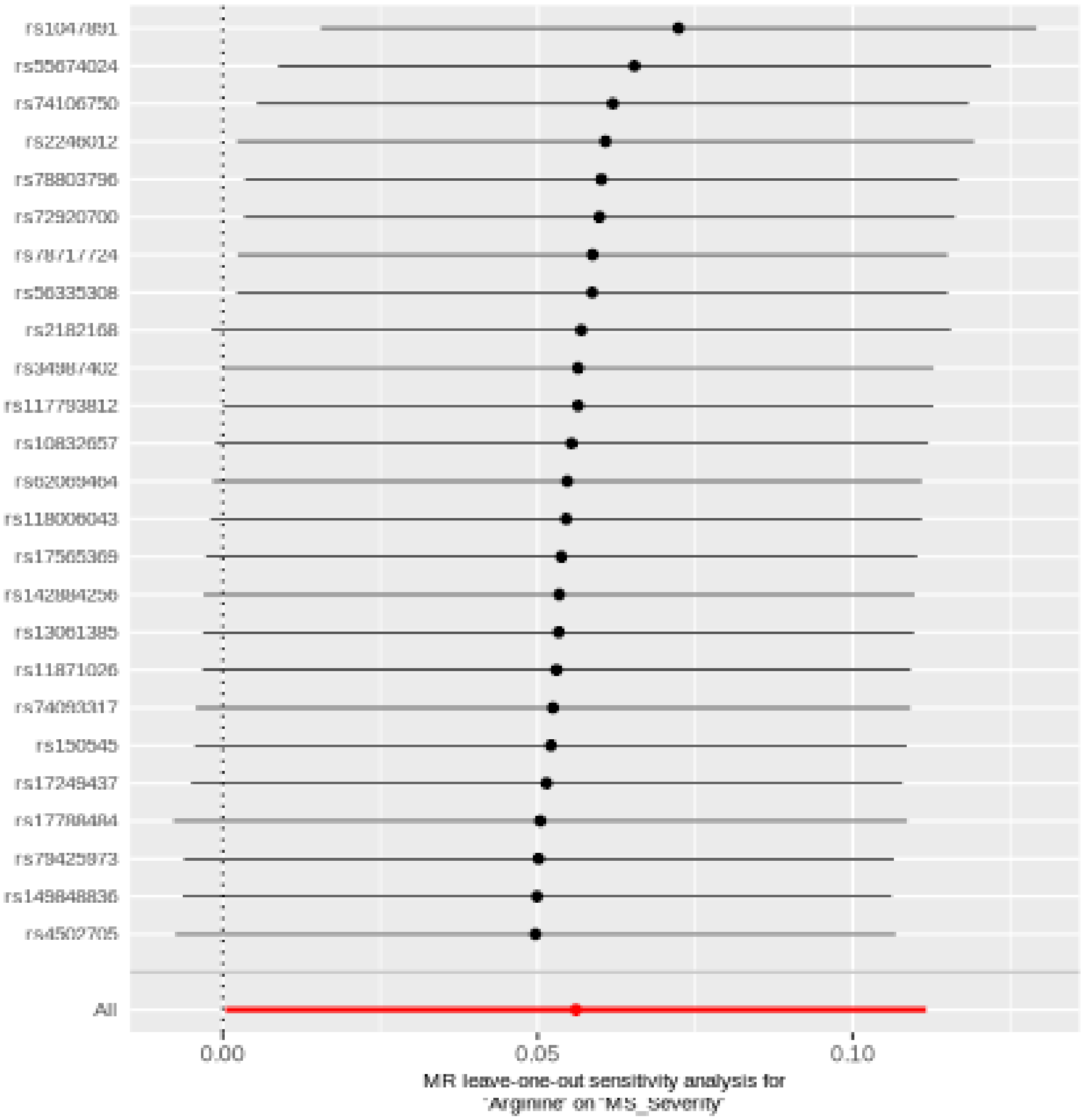

#### Betaine

| method | nsnp | b | se | pval |
|---|---|---|---|---|
| MR Egger | 24 | -0.1261352 | 0.0618965 | 0.0537680 |
| Weighted median | 24 | -0.0617226 | 0.0364778 | 0.0906350 |
| Inverse variance weighted | 24 | -0.0618349 | 0.0248282 | 0.0127559 |
| Simple mode | 24 | -0.1503048 | 0.0696389 | 0.0415785 |
| Weighted mode | 24 | -0.0614539 | 0.0390615 | 0.1293147 |

##### Heterogeneity tests

| method | Q | Q_df | Q_pval |
|---|---|---|---|
| MR Egger | 19.68489 | 22 | 0.6027430 |
| Inverse variance weighted | 20.97101 | 23 | 0.5828579 |

##### Test for directional horizontal pleiotropy

| egger_intercept | se | pval |
|---|---|---|
| 0.0102123 | 0.009005 | 0.2689715 |

Forest plot of single SNP MR

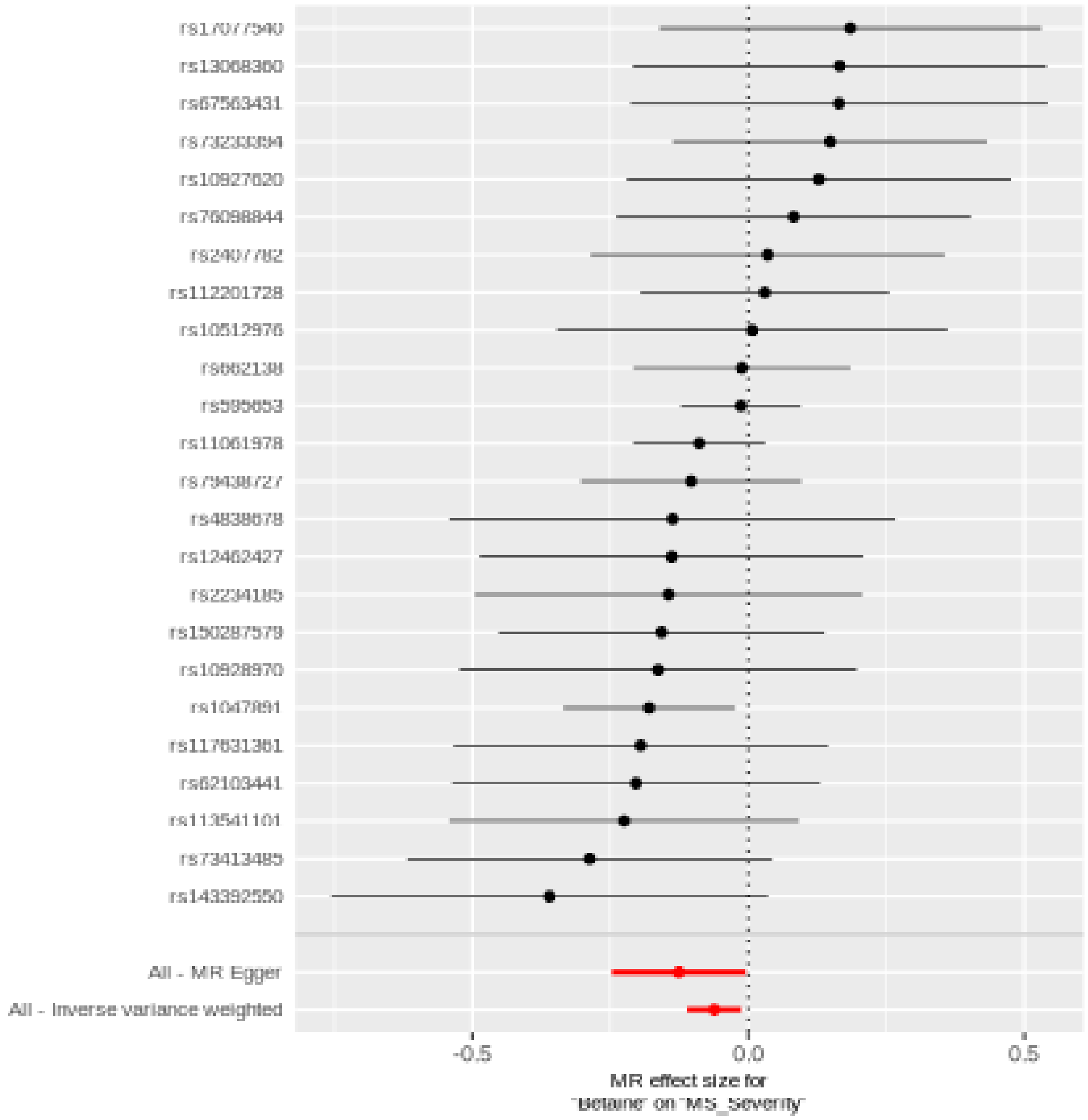

Comparison of results using different MR methods

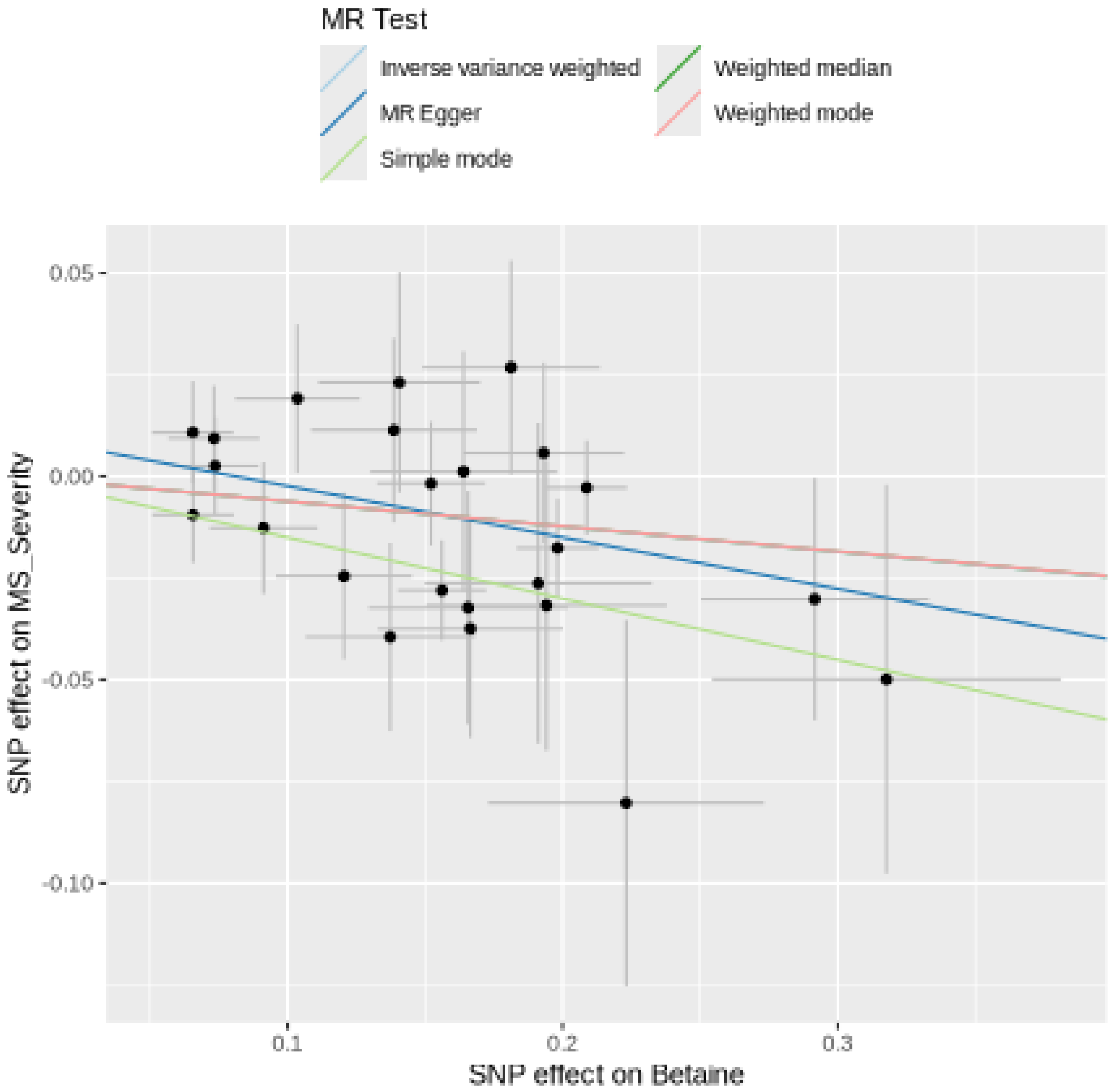

Funnel plot

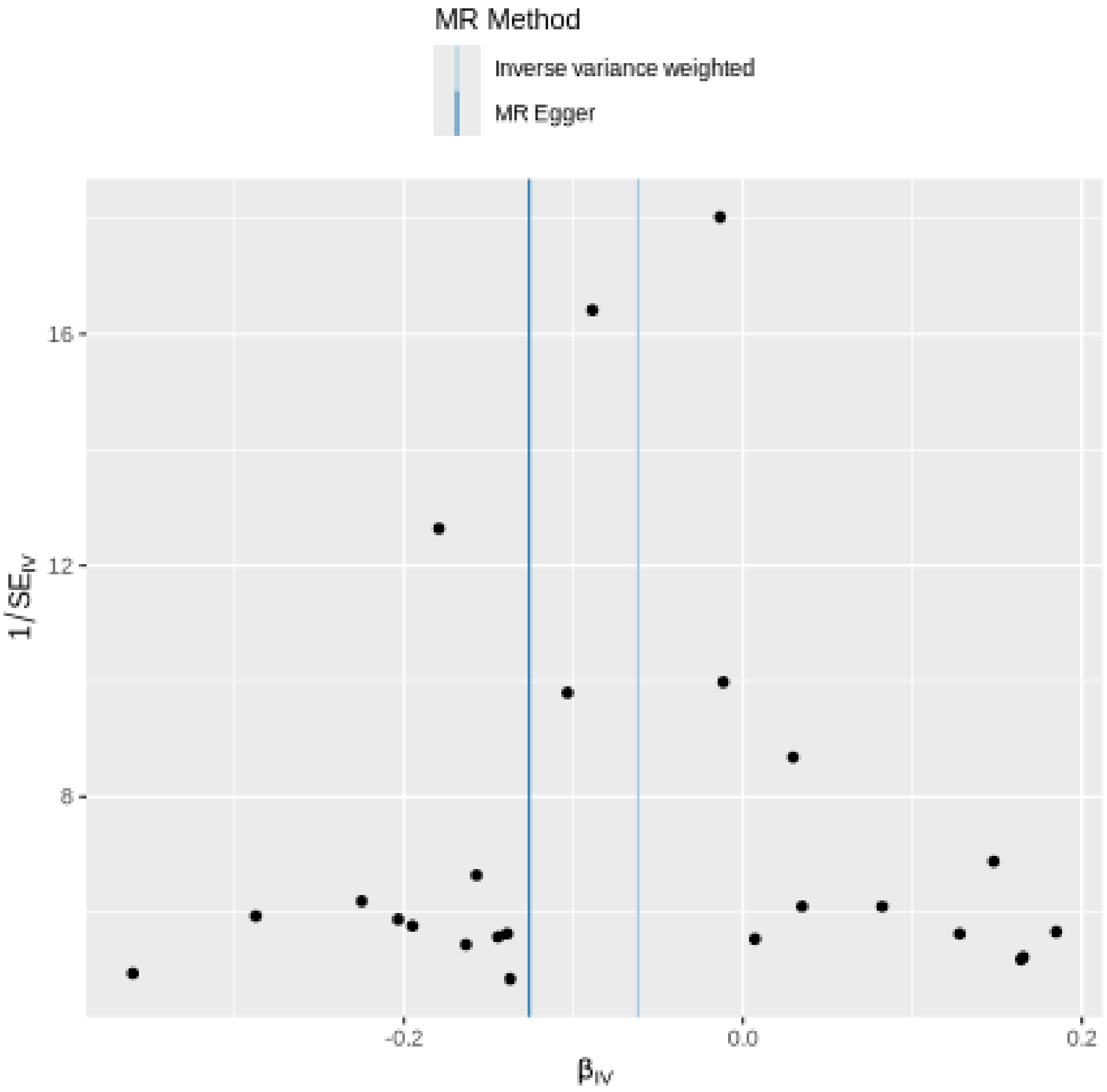

MR Method
Inverse variance weighted
MR Egger
16
12
8
1/SE_IV
-0.2
0.0
0.2
β_IV

#### Leave-one-out sensitivity analysis

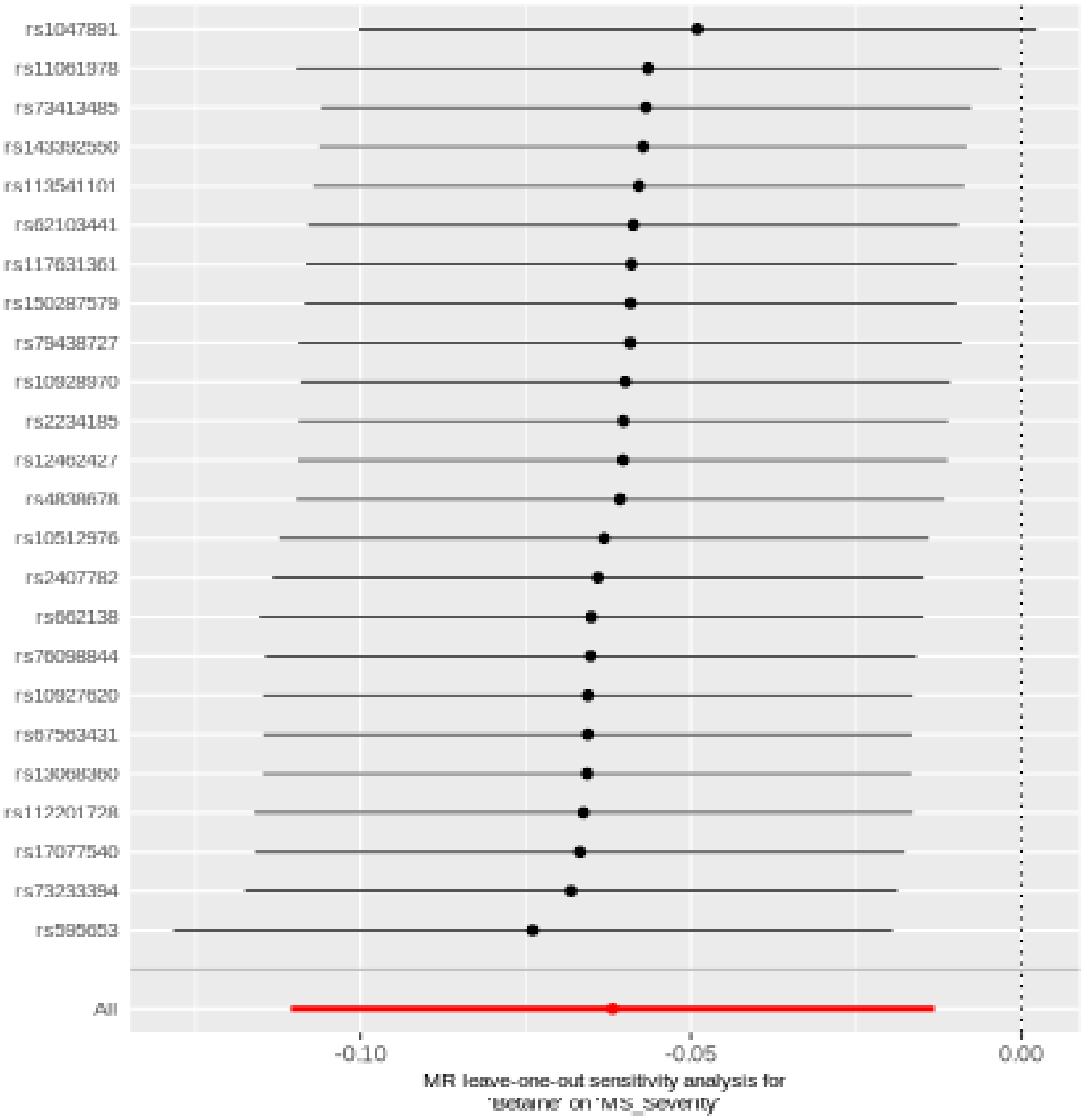

#### Alpha-ketoglutarate

| method | nsnp | b | se | pval |
|---|---|---|---|---|
| MR Egger | 28 | -0.1338525 | 0.0822875 | 0.1158734 |
| Weighted median | 28 | -0.0697091 | 0.0426470 | 0.1021414 |
| Inverse variance weighted | 28 | -0.0867140 | 0.0320748 | 0.0068616 |
| Simple mode | 28 | -0.0552400 | 0.0792062 | 0.4915021 |
| Weighted mode | 28 | -0.0800534 | 0.0565571 | 0.1683702 |

##### Heterogeneity tests

| method | Q | Q_df | Q_pval |
|---|---|---|---|
| MR Egger | 33.95993 | 26 | 0.1360361 |
| Inverse variance weighted | 34.46745 | 27 | 0.1528755 |

##### Test for directional horizontal pleiotropy

| egger_intercept | se | pval |
|---|---|---|
| 0.0065216 | 0.0104622 | 0.5384855 |

#### Forest plot of single SNP MR

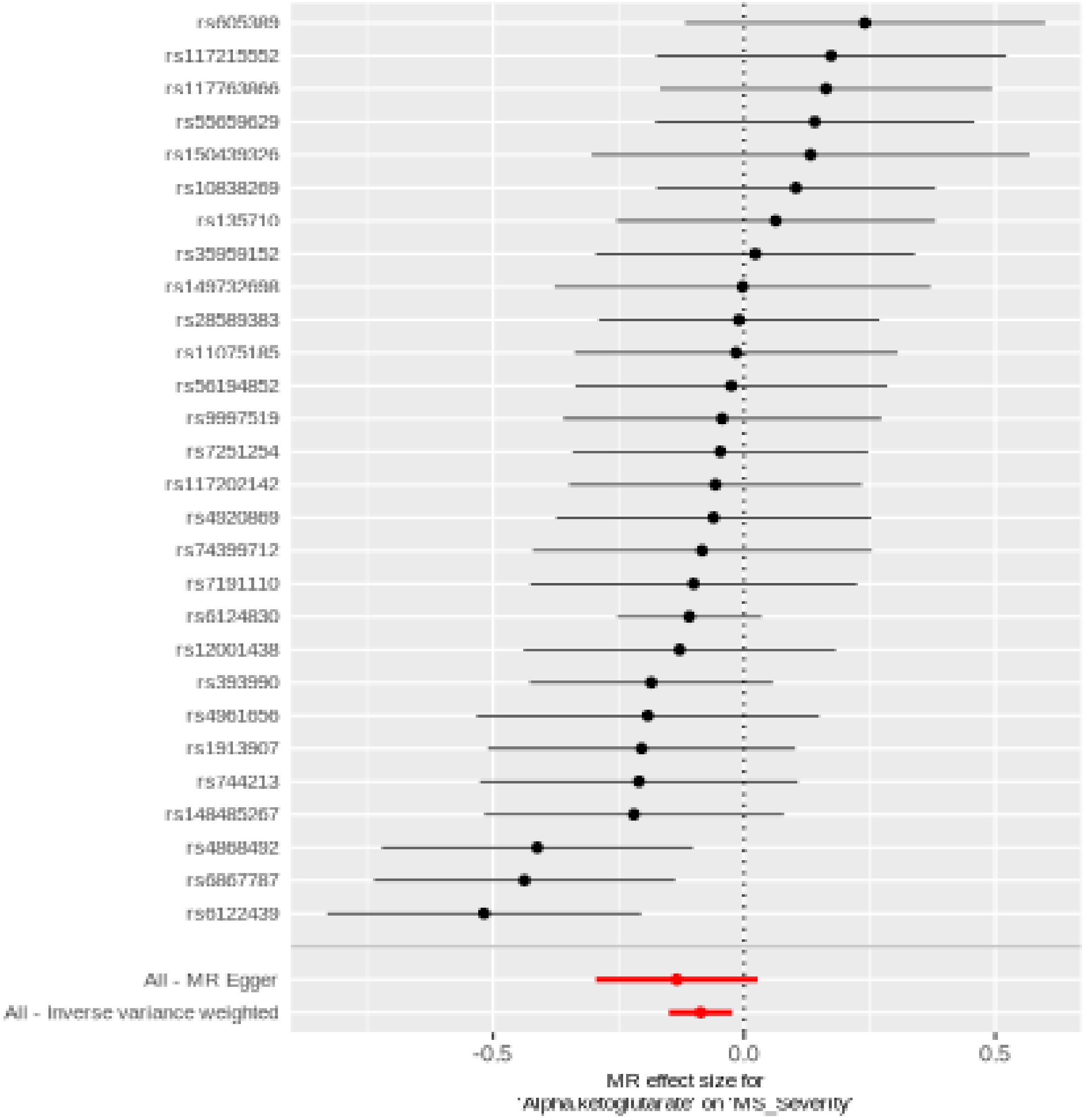

Comparison of results using different MR methods

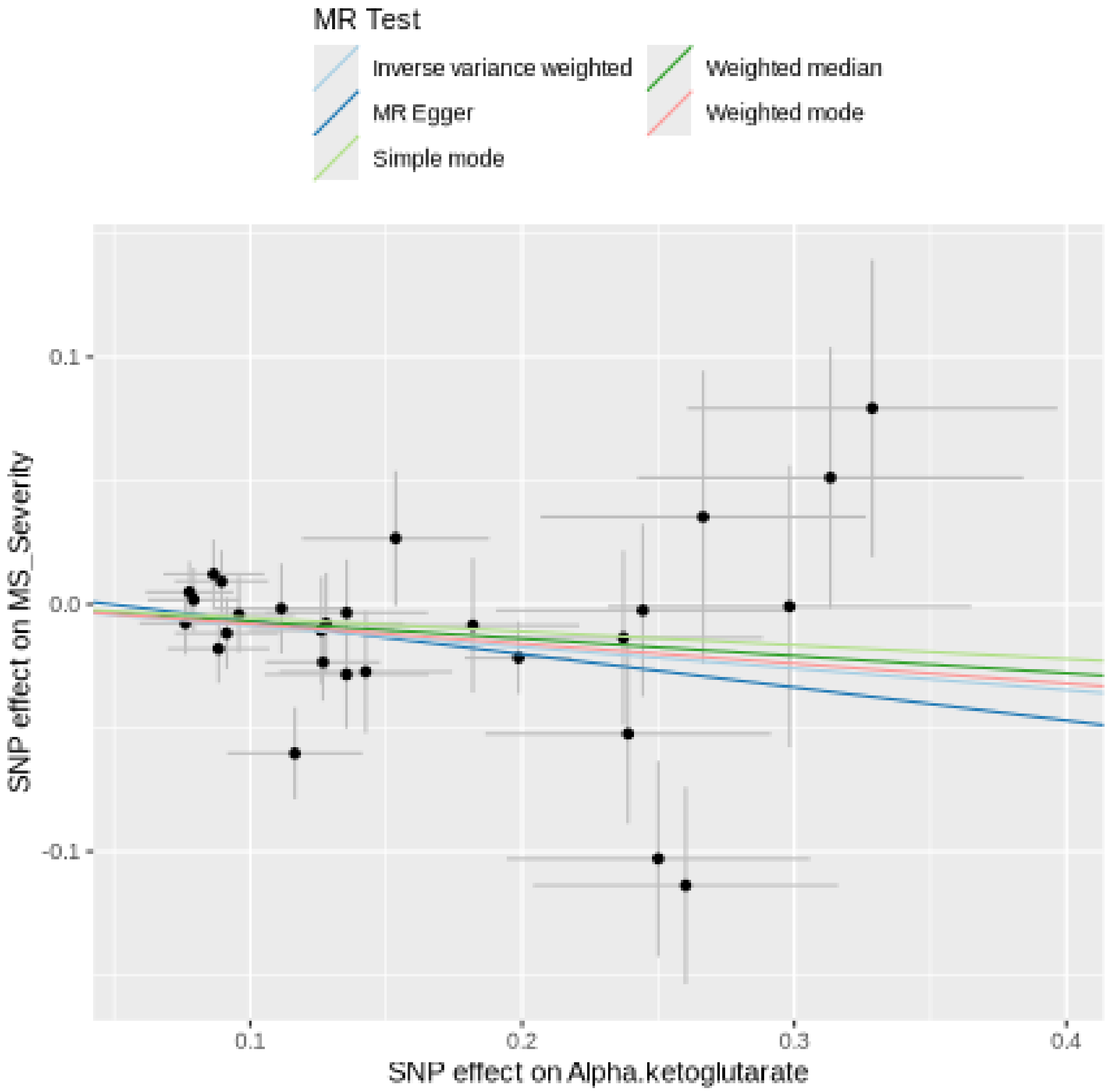

Funnel plot

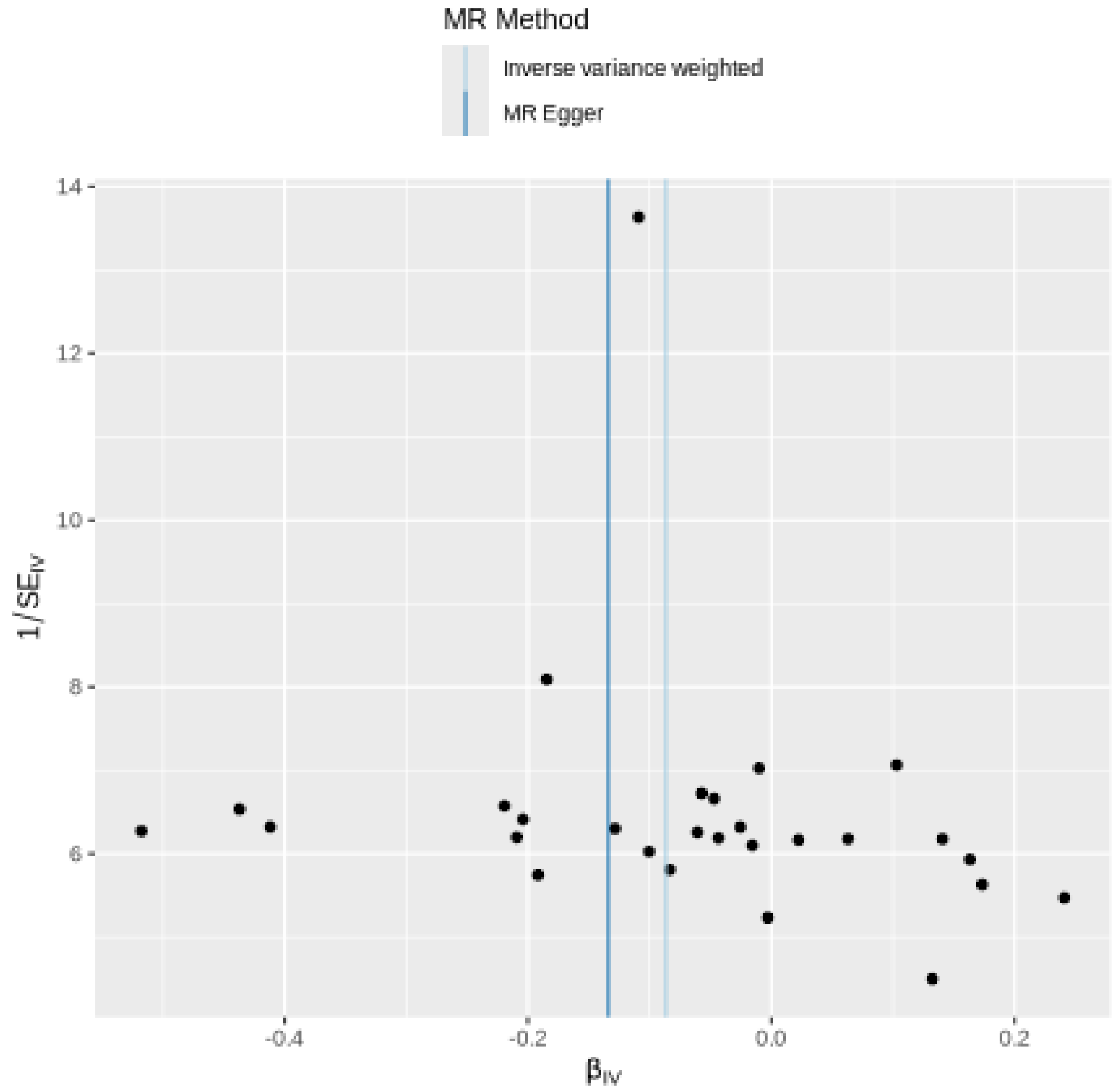

MR Method
Inverse variance weighted
MR Egger
14
12
10
8
6
1/SE$_{IV}$
-0.4
-0.2
0.0
0.2
β$_{IV}$

Leave-one-out sensitivity analysis

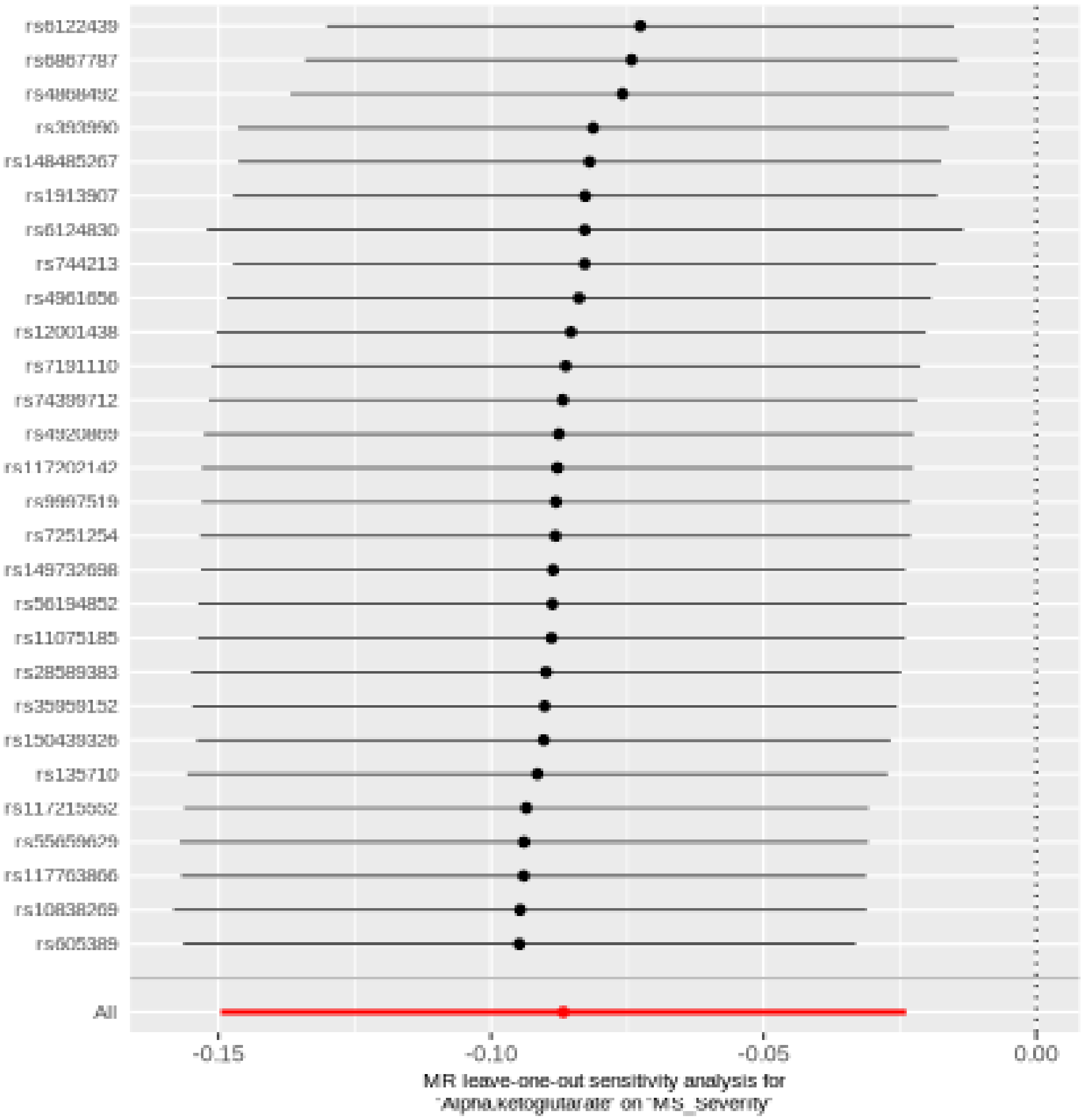

#### Butyrylcarnitine (C4)

| method | nsnp | b | se | pval |
|---|---|---|---|---|
| MR Egger | 59 | -0.0006175 | 0.0224423 | 0.9781438 |
| Weighted median | 59 | -0.0057478 | 0.0204787 | 0.7789628 |
| Inverse variance weighted | 59 | -0.0325559 | 0.0133676 | 0.0148739 |
| Simple mode | 59 | -0.0276721 | 0.0396725 | 0.4882681 |
| Weighted mode | 59 | -0.0125559 | 0.0192701 | 0.5172516 |

##### Heterogeneity tests

| method | Q | Q_df | Q_pval |
|---|---|---|---|
| MR Egger | 51.30624 | 57 | 0.6876192 |
| Inverse variance weighted | 54.44522 | 58 | 0.6082565 |

##### Test for directional horizontal pleiotropy

| egger_intercept | se | pval |
|---|---|---|
| -0.0073785 | 0.0041646 | 0.0817866 |

Forest plot of single SNP MR

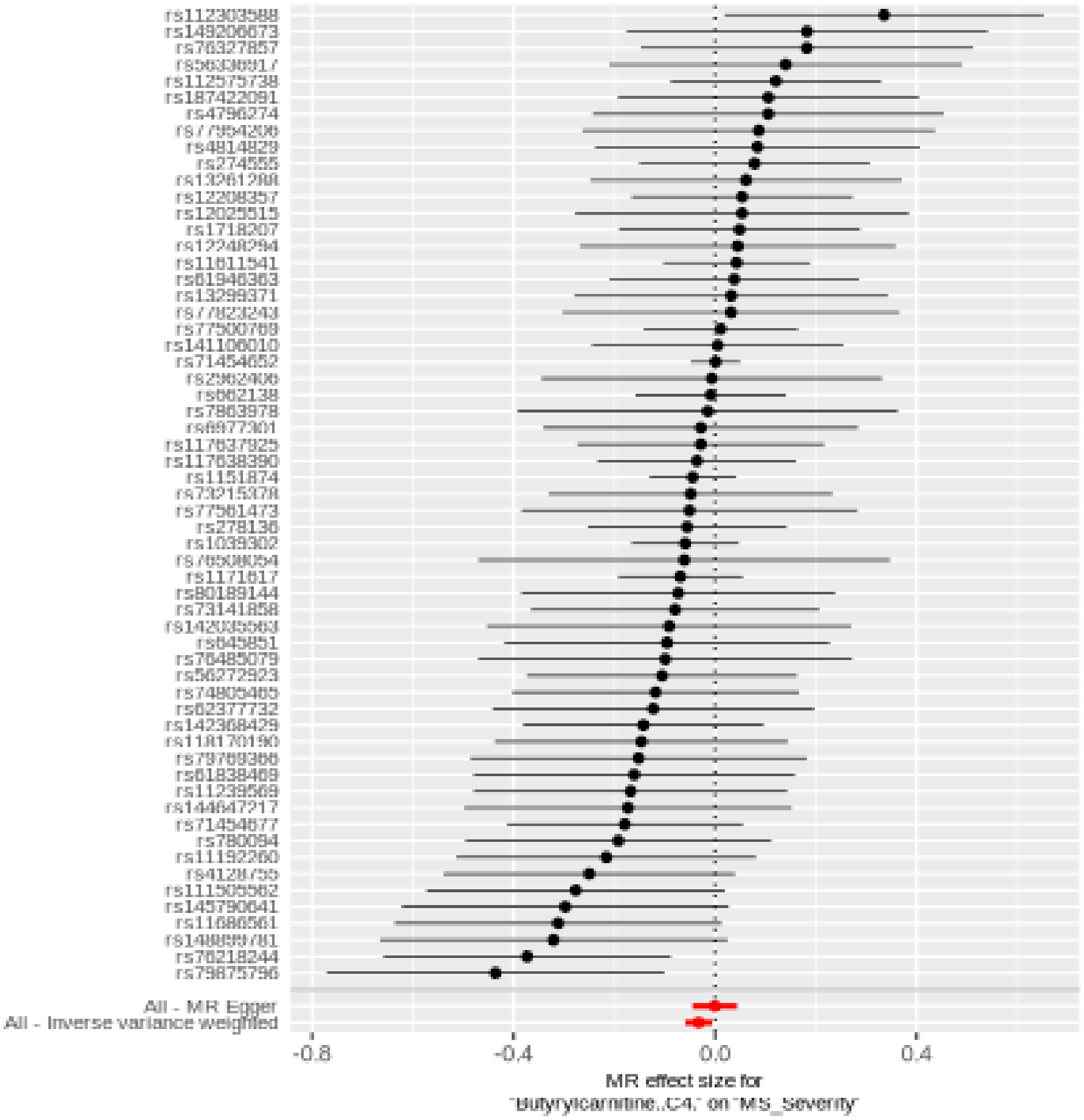

Comparison of results using different MR methods

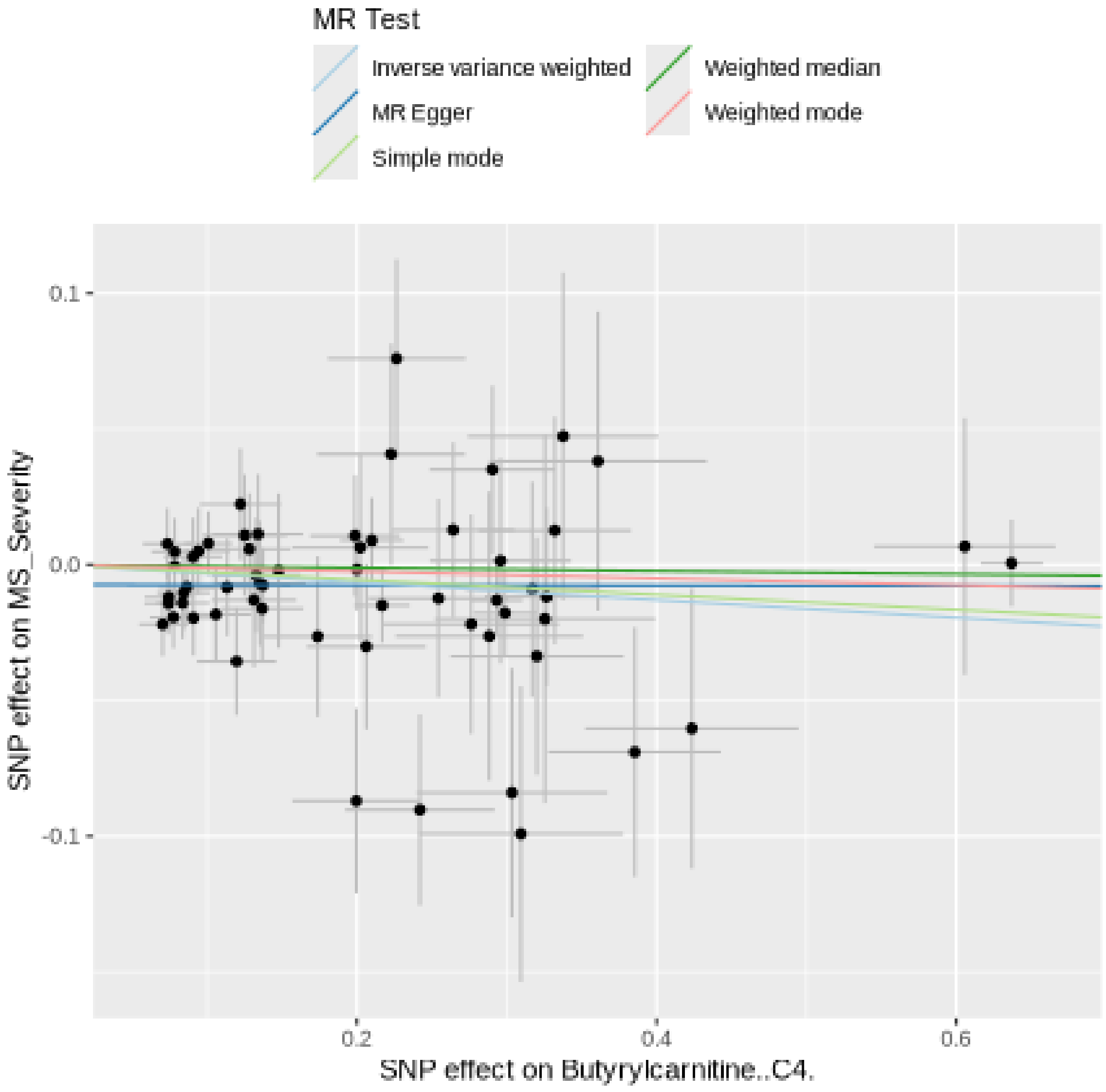

Funnel plot

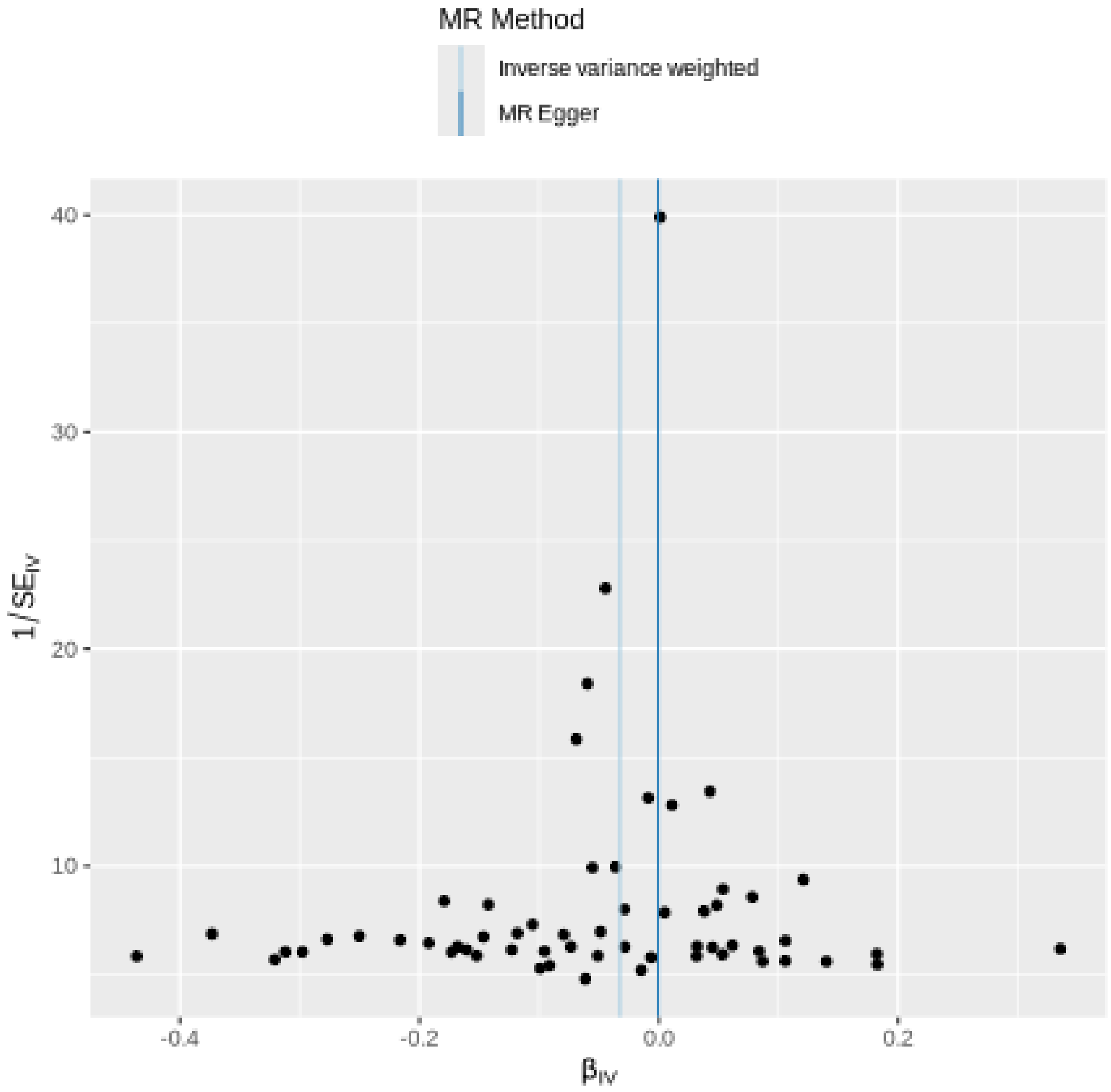
MR Method
Inverse variance weighted
MR Egger
40
30
20
10
1/SE$_{IV}$
-0.4
-0.2
0.0
0.2
β$_{IV}$

### Leave-one-out sensitivity analysis

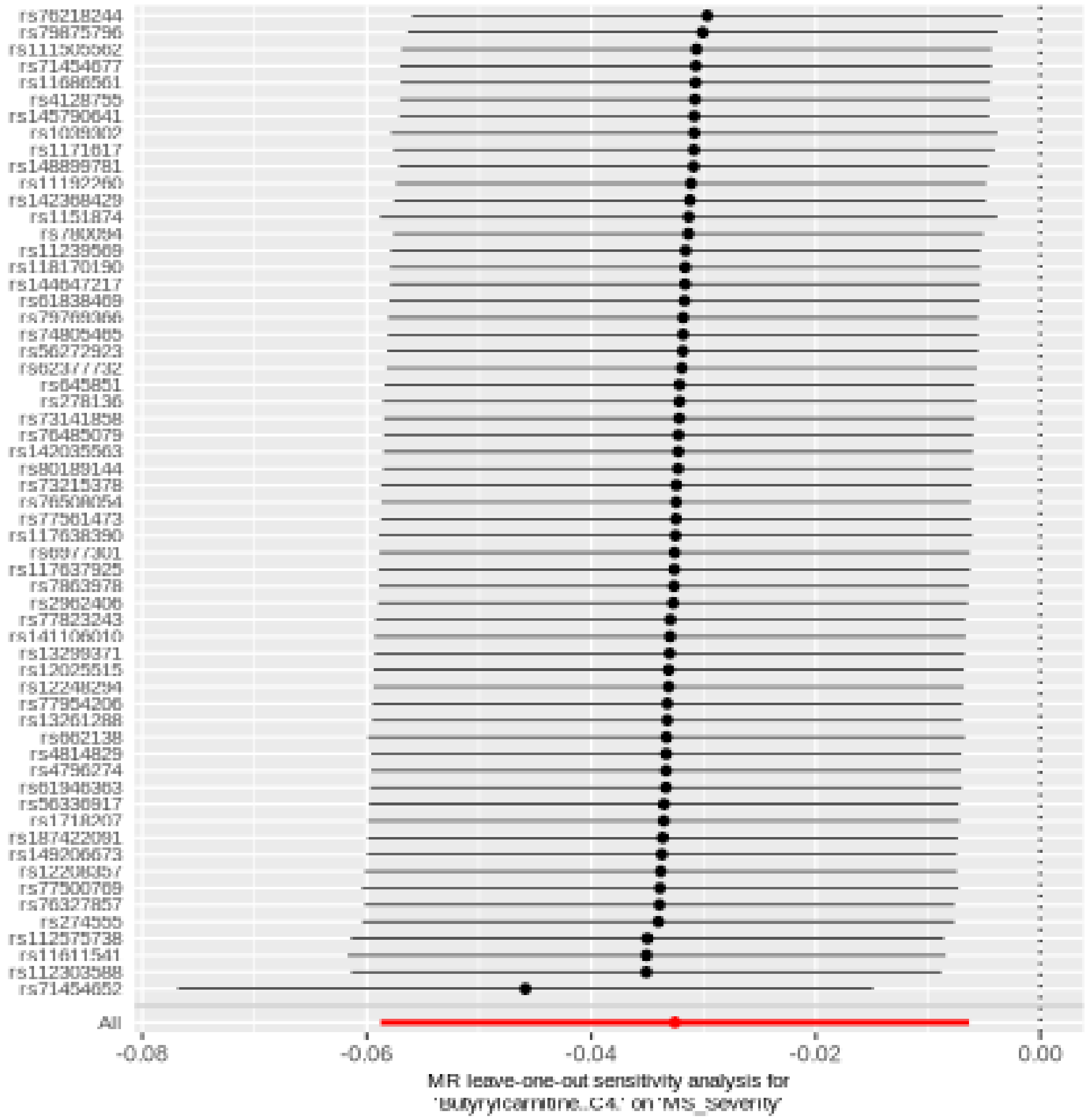

#### Glycochenodeoxycholate glucuronide (1)

| method | nsnp | b | se | pval |
|---|---|---|---|---|
| MR Egger | 34 | 0.0407513 | 0.0236657 | 0.0947325 |
| Weighted median | 34 | 0.0384530 | 0.0175591 | 0.0285301 |
| Inverse variance weighted | 34 | 0.0490606 | 0.0170239 | 0.0039534 |
| Simple mode | 34 | 0.0197047 | 0.0498876 | 0.6953972 |
| Weighted mode | 34 | 0.0398606 | 0.0169419 | 0.0247468 |

##### Heterogeneity tests

| method | Q | Q_df | Q_pval |
|---|---|---|---|
| MR Egger | 47.75099 | 32 | 0.0362576 |
| Inverse variance weighted | 48.14181 | 33 | 0.0430472 |

##### Test for directional horizontal pleiotropy

| egger_intercept | se | pval |
|---|---|---|
| 0.0027995 | 0.0054702 | 0.6123269 |

#### Forest plot of single SNP MR

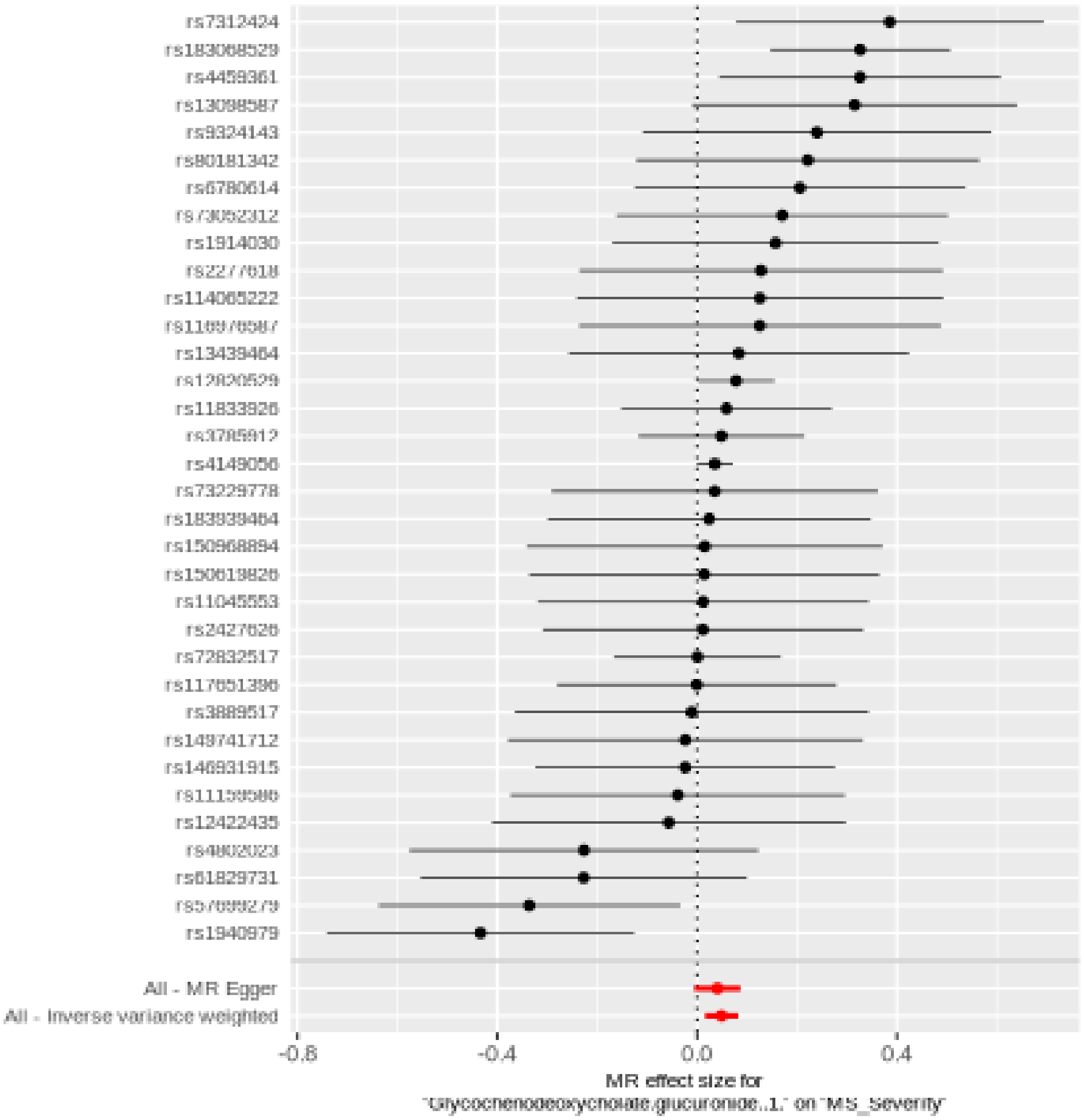

Comparison of results using different MR methods

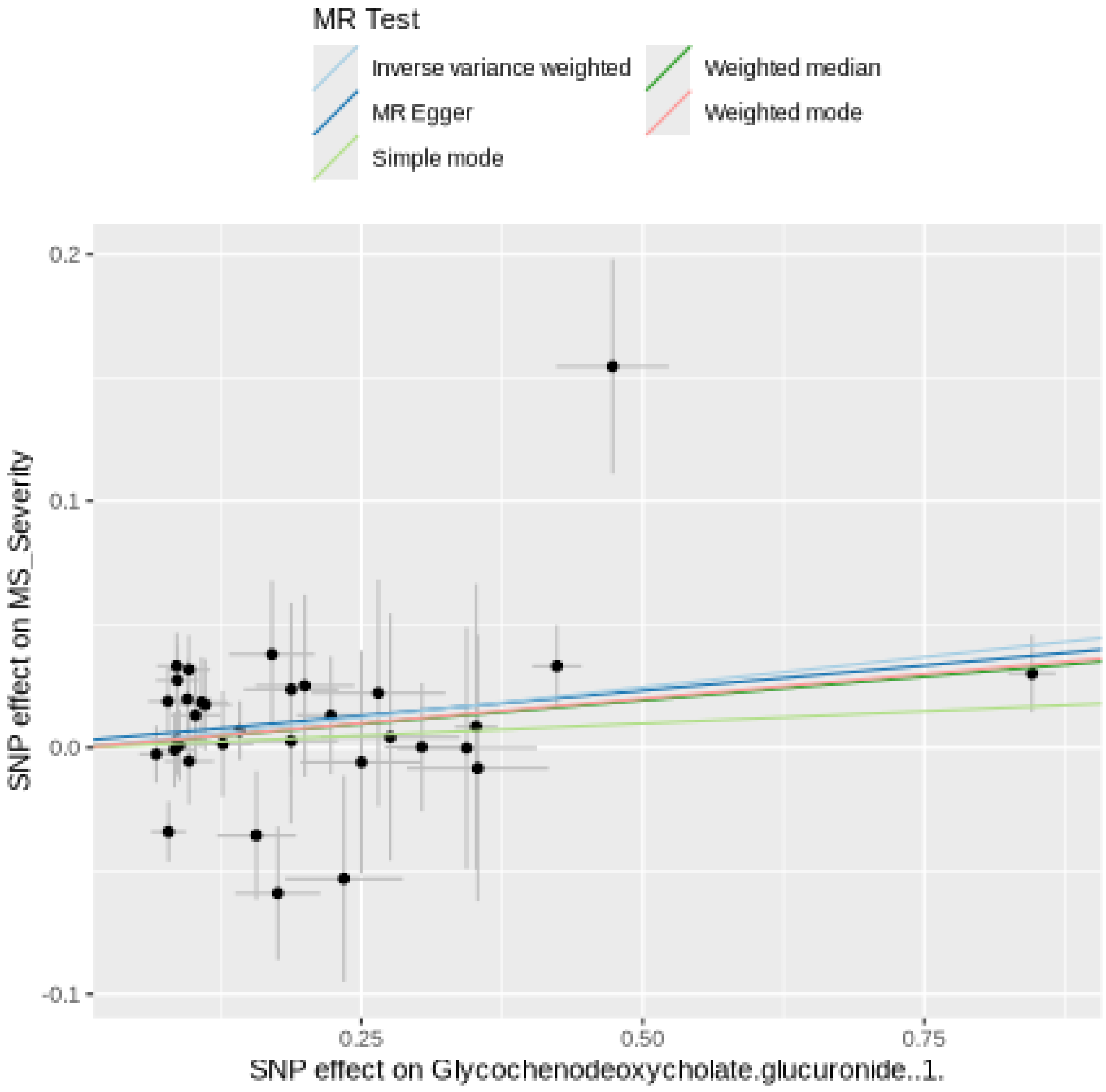

Funnel plot

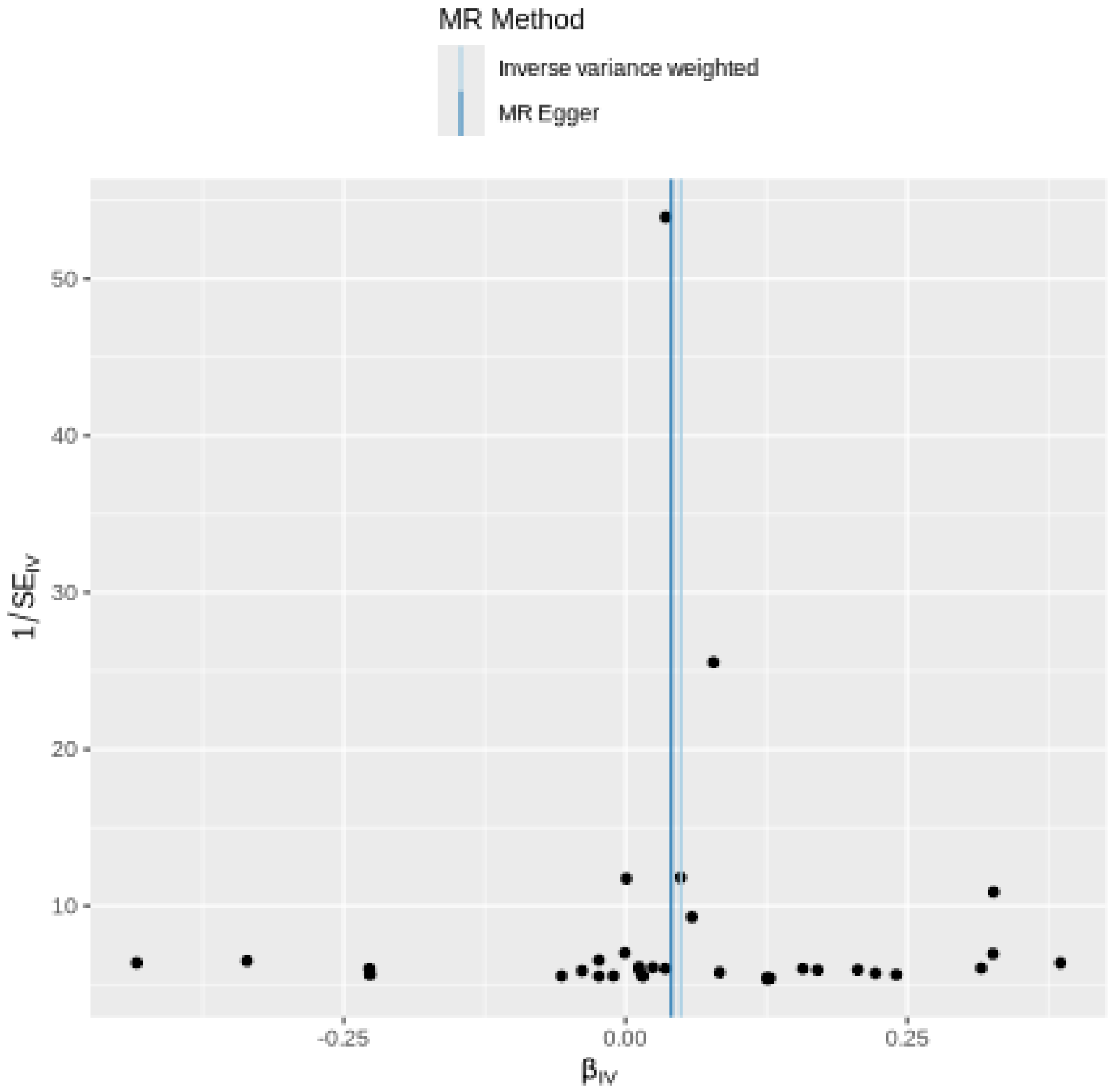

MR Method
Inverse variance weighted
MR Egger
50
40
30
20
10
1/SE_IV
-0.25
0.00
0.25
β_IV

#### Leave-one-out sensitivity analysis

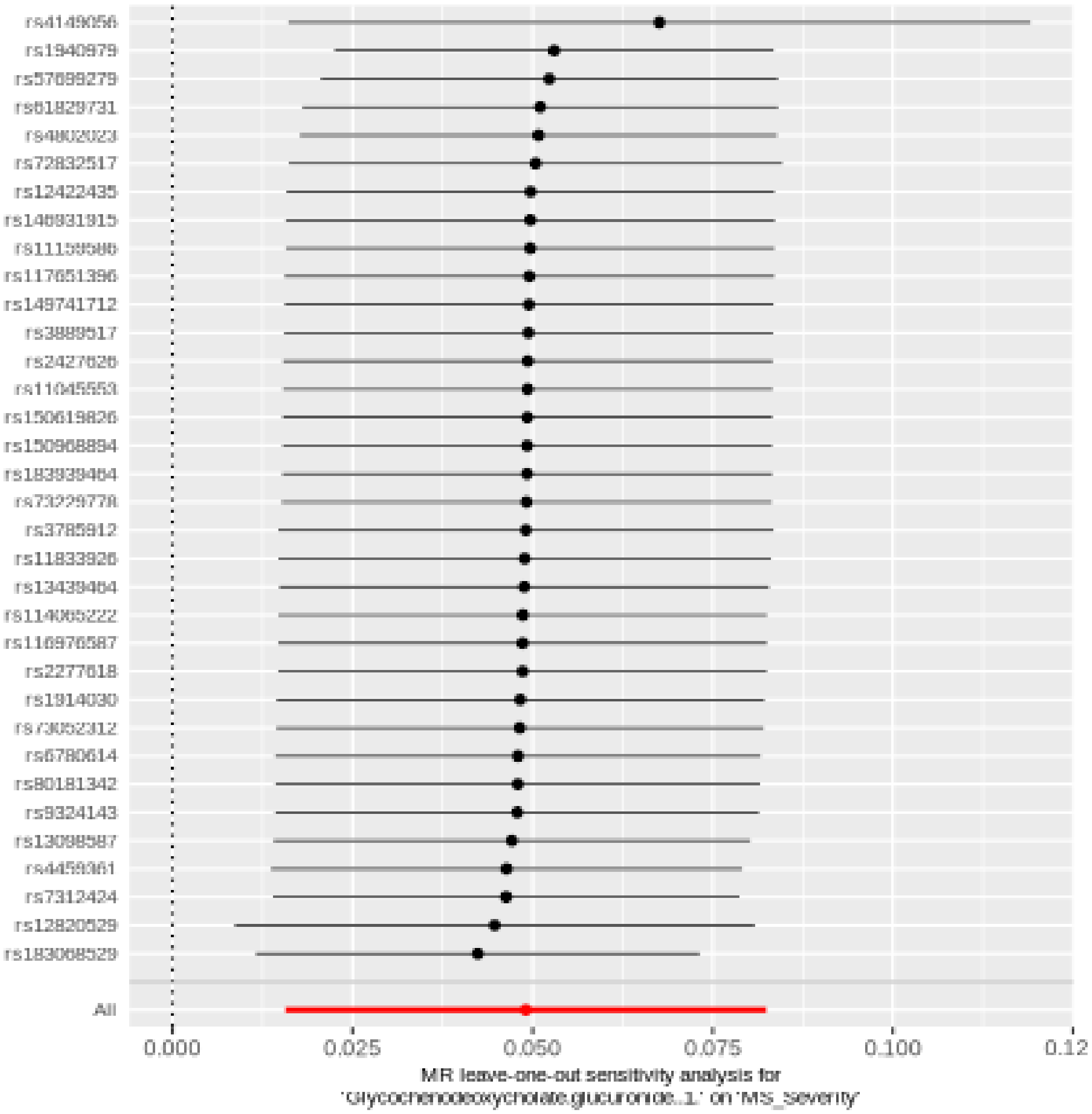

#### Decadienedioic acid (C10:2-DC)

| method | nsnp | b | se | pval |
|---|---|---|---|---|
| MR Egger | 42 | -0.0196551 | 0.0238065 | 0.4139168 |
| Weighted median | 42 | -0.0366350 | 0.0201254 | 0.0687079 |
| Inverse variance weighted | 42 | -0.0300140 | 0.0142714 | 0.0354575 |
| Simple mode | 42 | -0.0743965 | 0.0457576 | 0.1116378 |
| Weighted mode | 42 | -0.0394014 | 0.0193005 | 0.0476747 |

##### Heterogeneity tests

| method | Q | Q_df | Q_pval |
|---|---|---|---|
| MR Egger | 33.40101 | 40 | 0.7602417 |
| Inverse variance weighted | 33.69656 | 41 | 0.7838471 |

##### Test for directional horizontal pleiotropy

| egger_intercept | se | pval |
|---|---|---|
| -0.0029186 | 0.0053686 | 0.5897036 |

Forest plot of single SNP MR

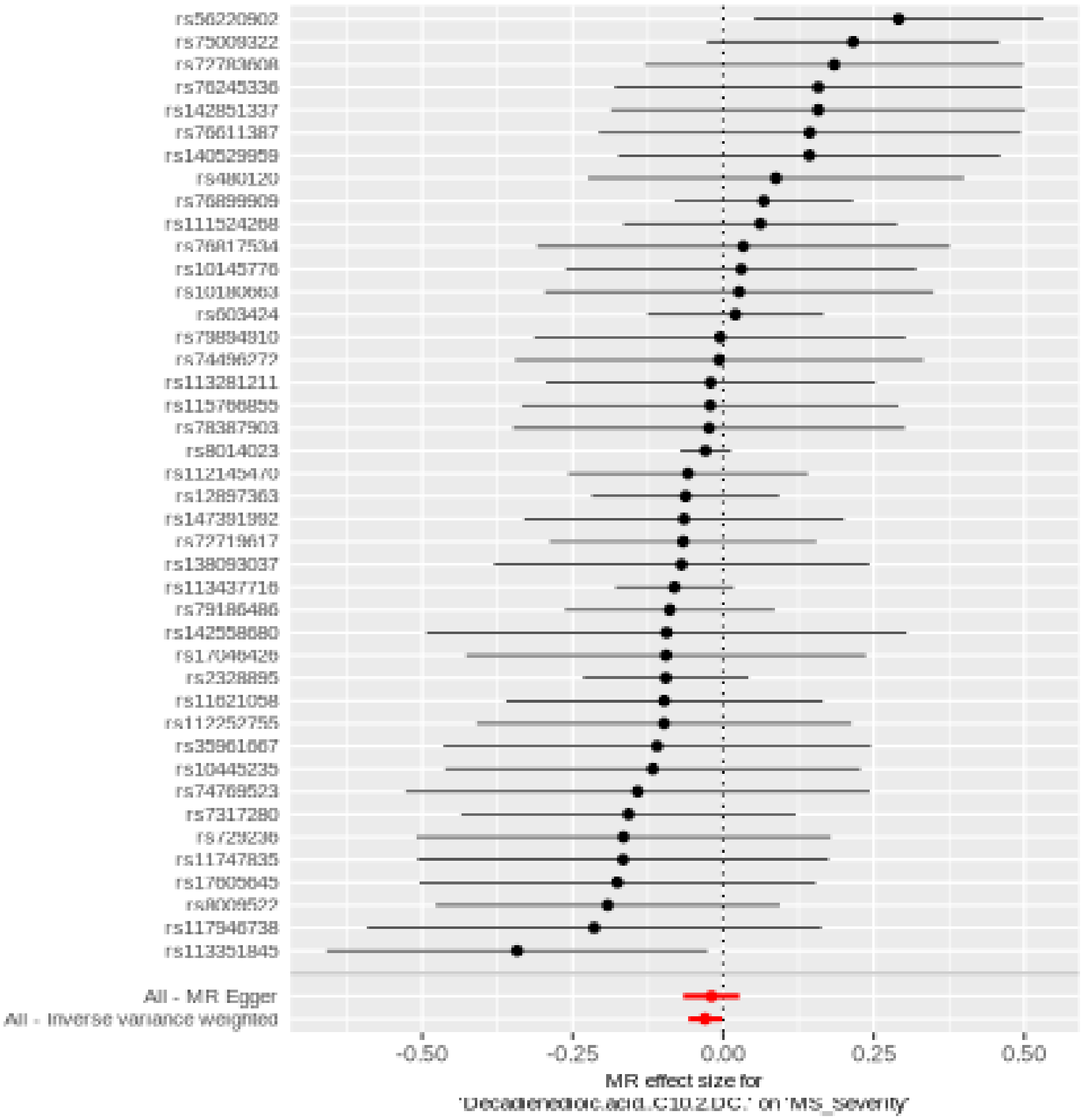

Comparison of results using different MR methods

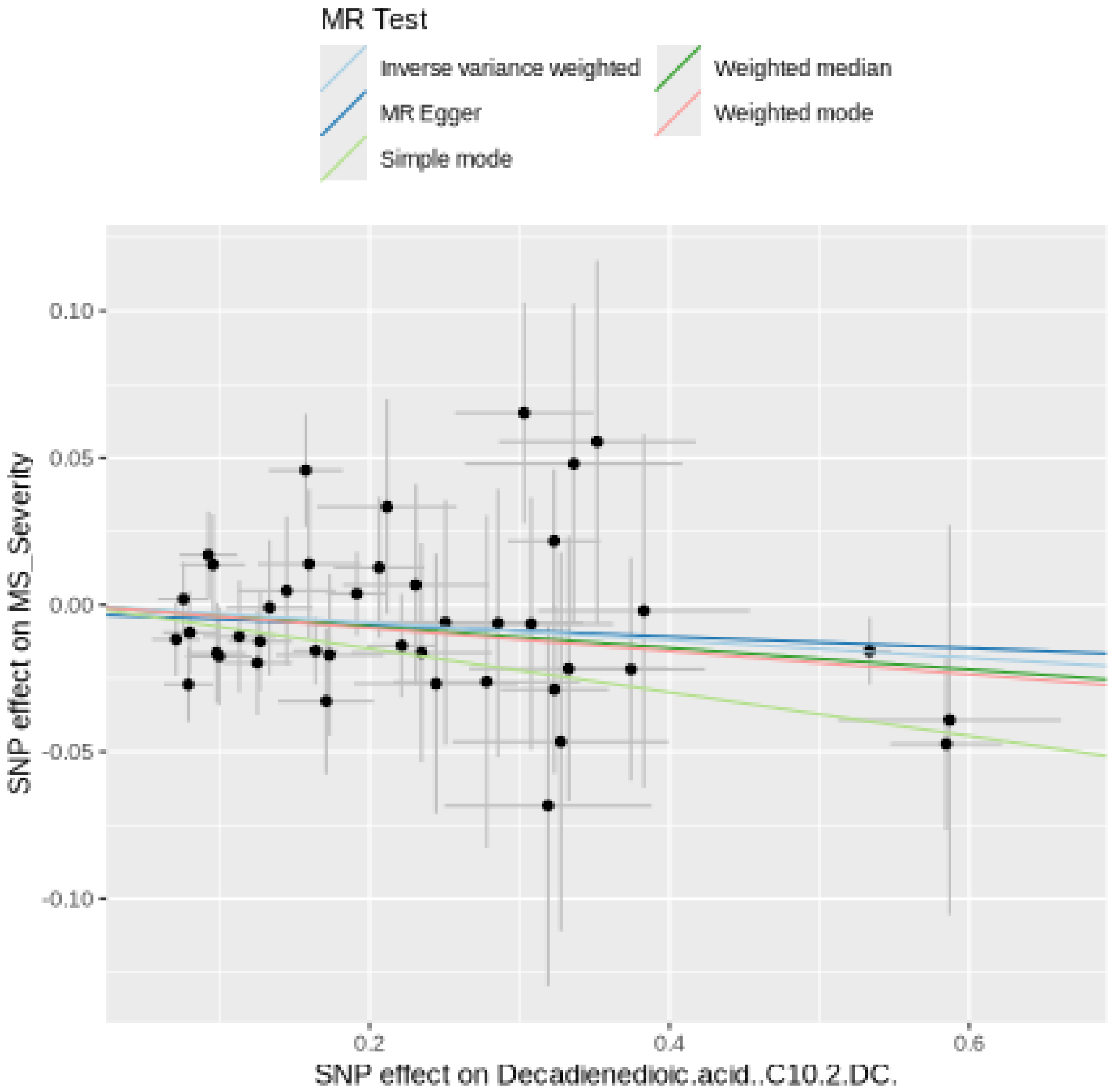

Funnel plot

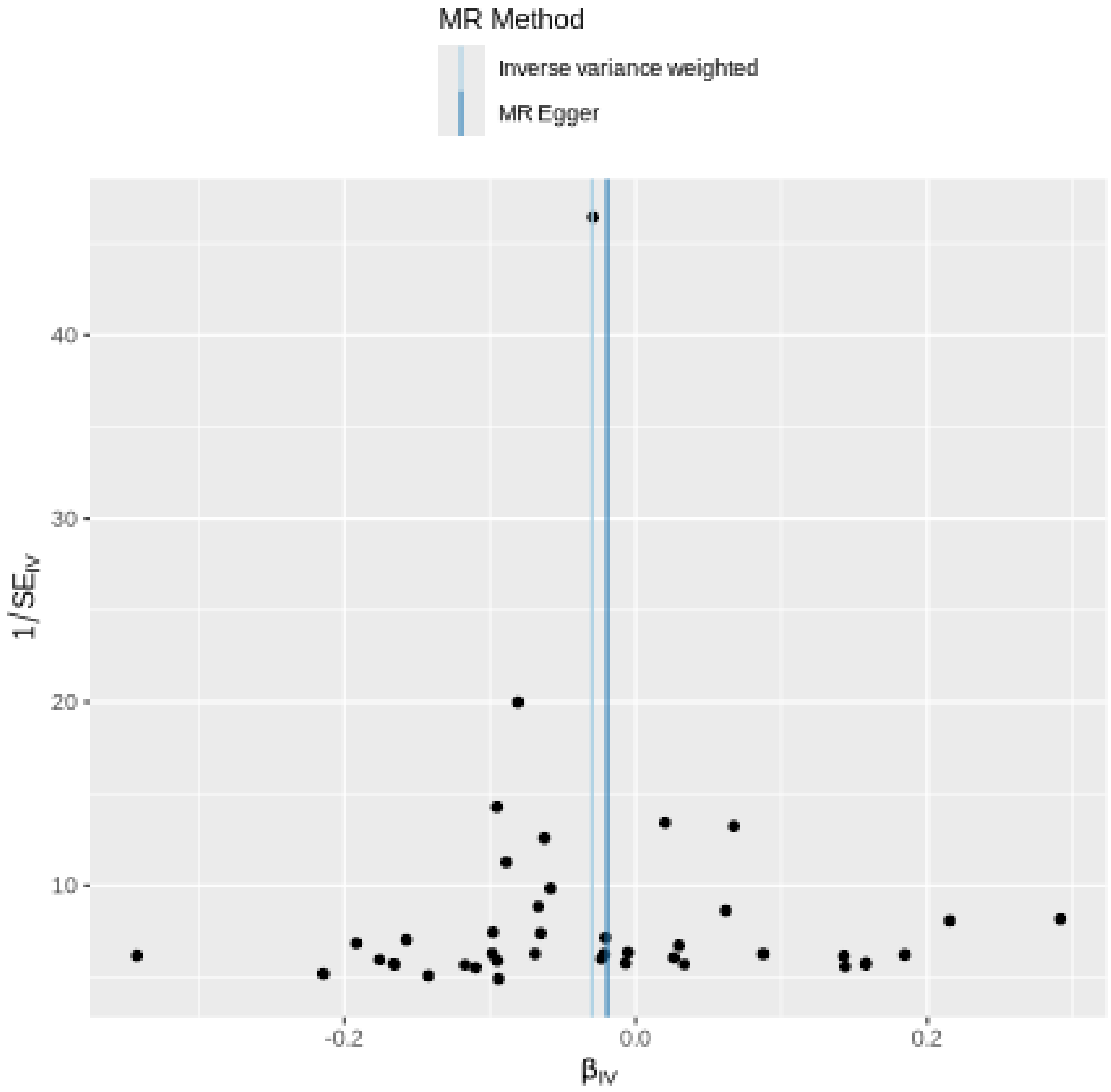

MR Method
Inverse variance weighted
MR Egger
40
30
20
10
1/SE$_{IV}$
-0.2
0.0
0.2
$\beta_{IV}$

### Leave-one-out sensitivity analysis

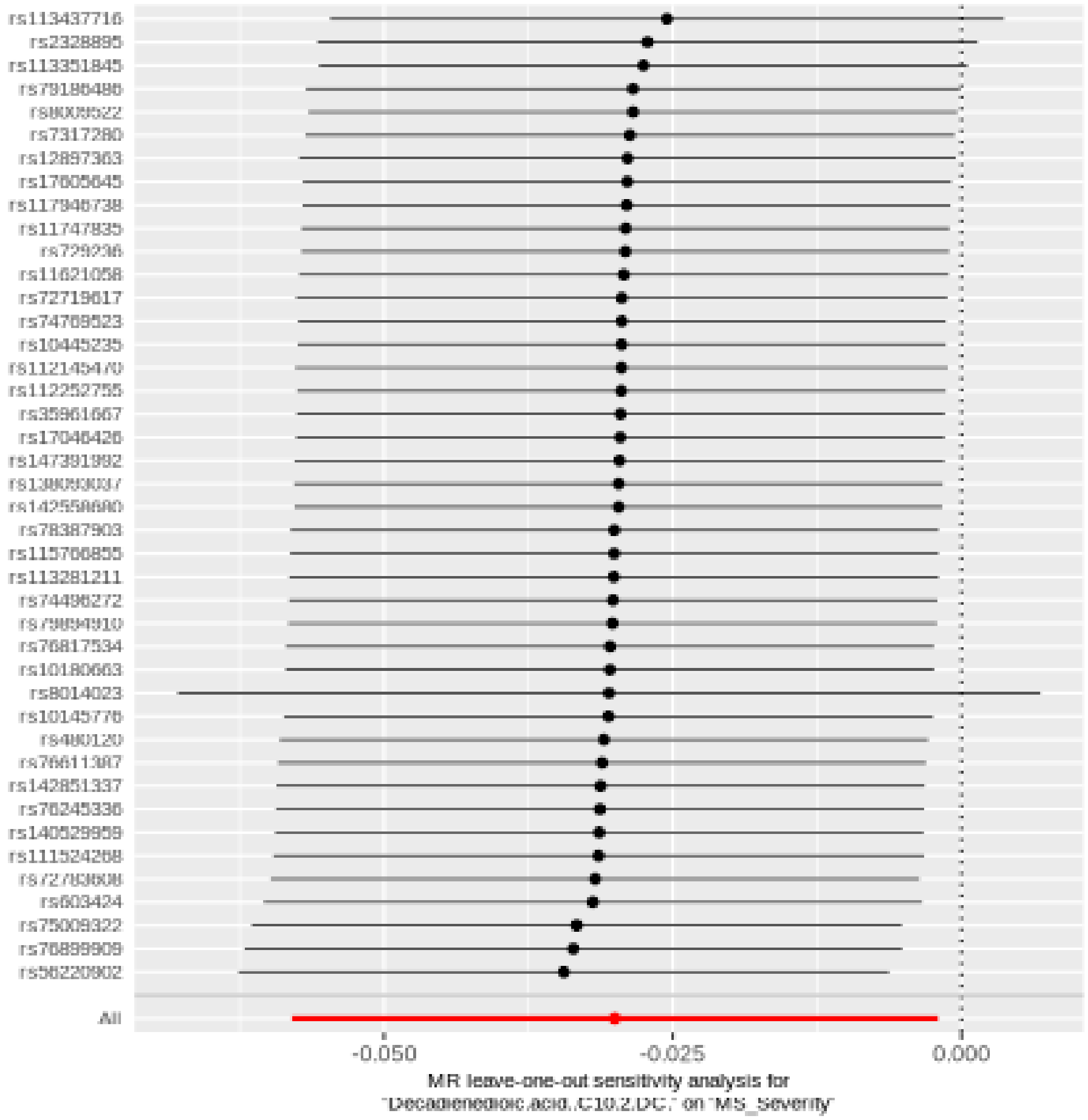

#### Arachidonate (20:4n6) to oleate to vaccenate (18:1) ratio

| method | nsnp | b | se | pval |
|---|---|---|---|---|
| MR Egger | 22 | -0.0696628 | 0.0357293 | 0.0653677 |
| Weighted median | 22 | -0.0665516 | 0.0266592 | 0.0125466 |
| Inverse variance weighted | 22 | -0.0454240 | 0.0218790 | 0.0378799 |
| Simple mode | 22 | 0.0409172 | 0.0745179 | 0.5887337 |
| Weighted mode | 22 | -0.0676841 | 0.0266998 | 0.0192611 |

#### Heterogeneity tests

| method | Q | Q_df | Q_pval |
|---|---|---|---|
| MR Egger | 16.13134 | 20 | 0.7084437 |
| Inverse variance weighted | 16.86767 | 21 | 0.7190961 |

#### Test for directional horizontal pleiotropy

| egger_intercept | se | pval |
|---|---|---|
| 0.0059066 | 0.0068833 | 0.4010051 |

#### Forest plot of single SNP MR

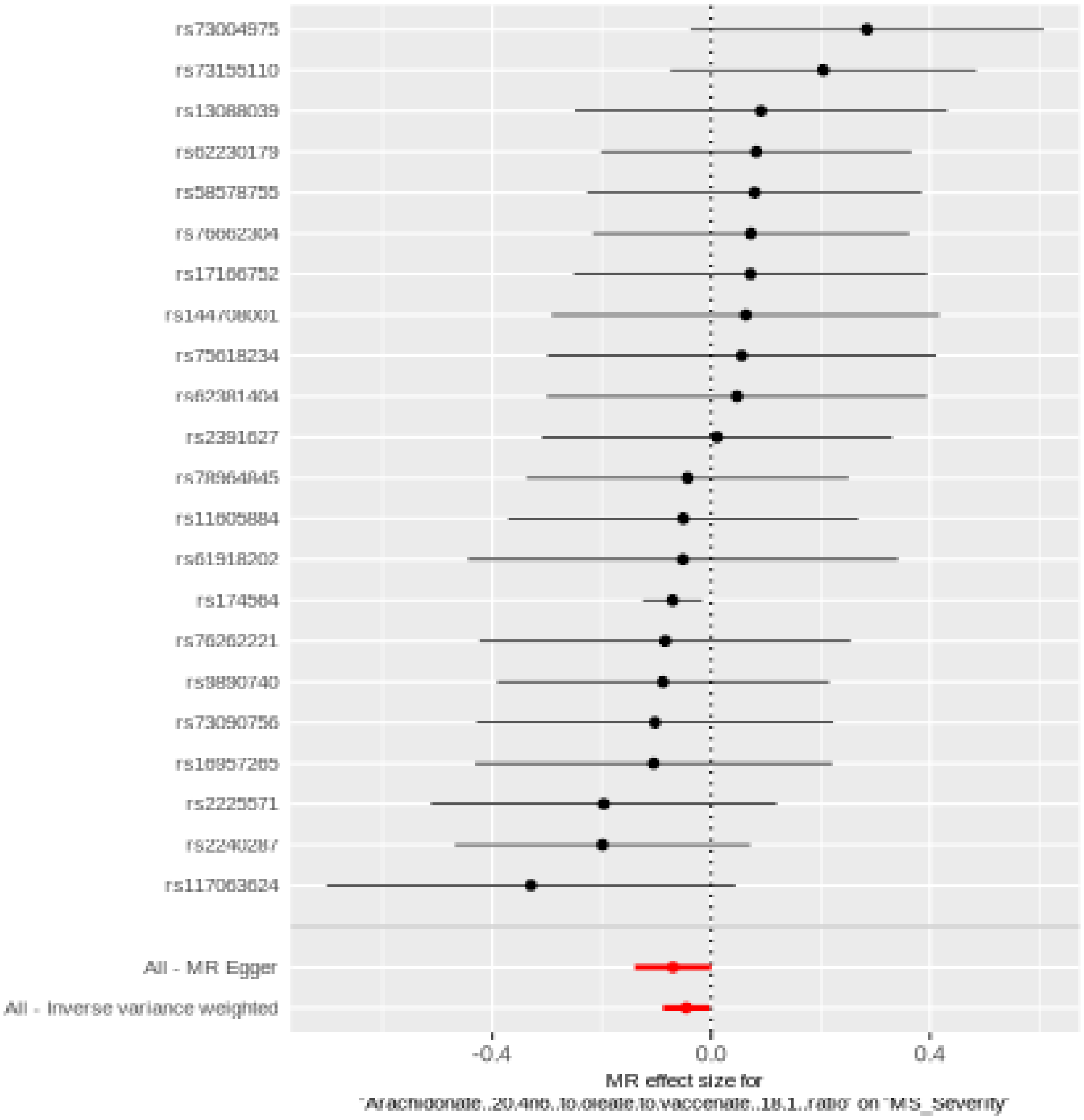

#### Comparison of results using different MR methods

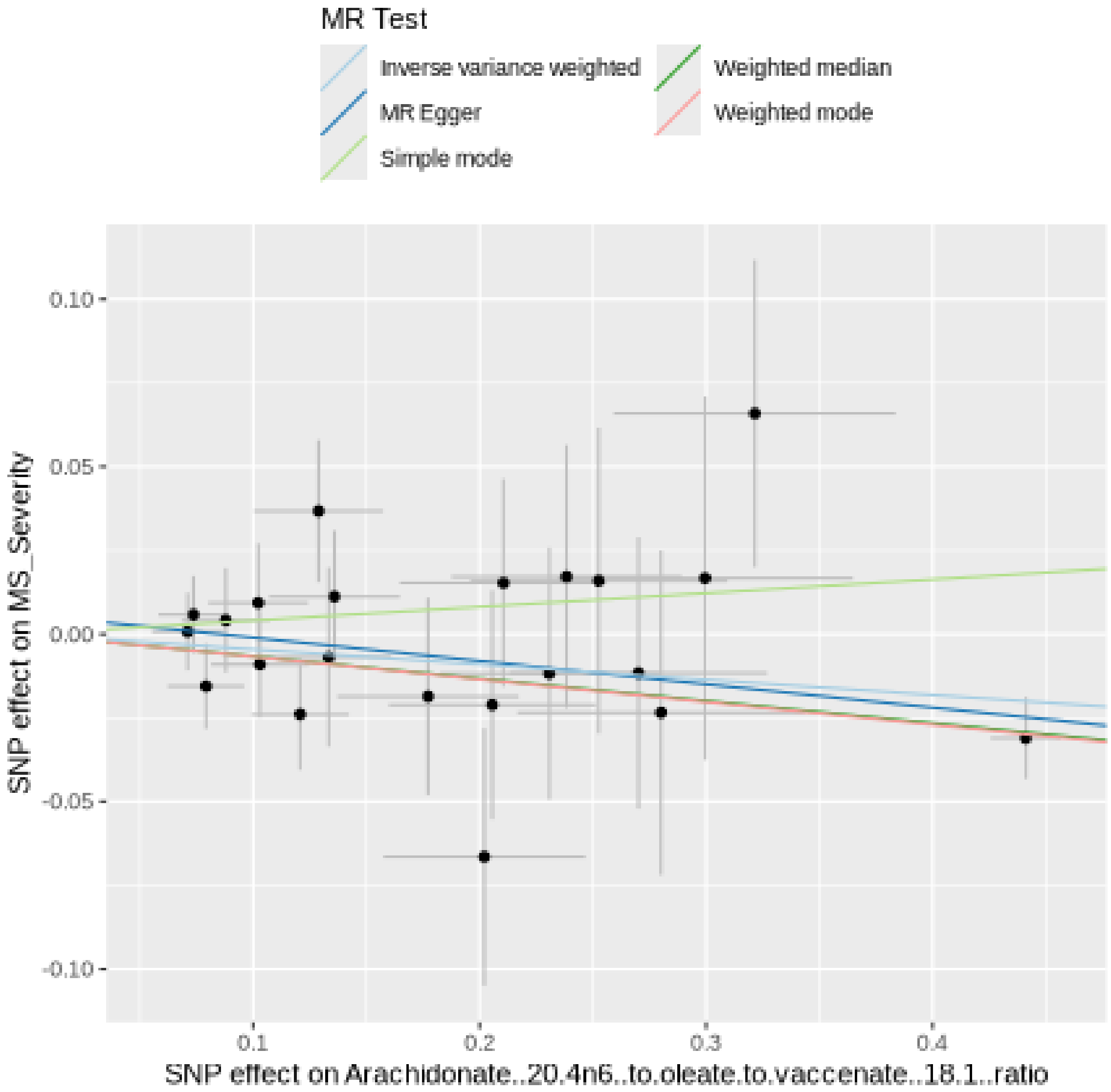

#### Funnel plot

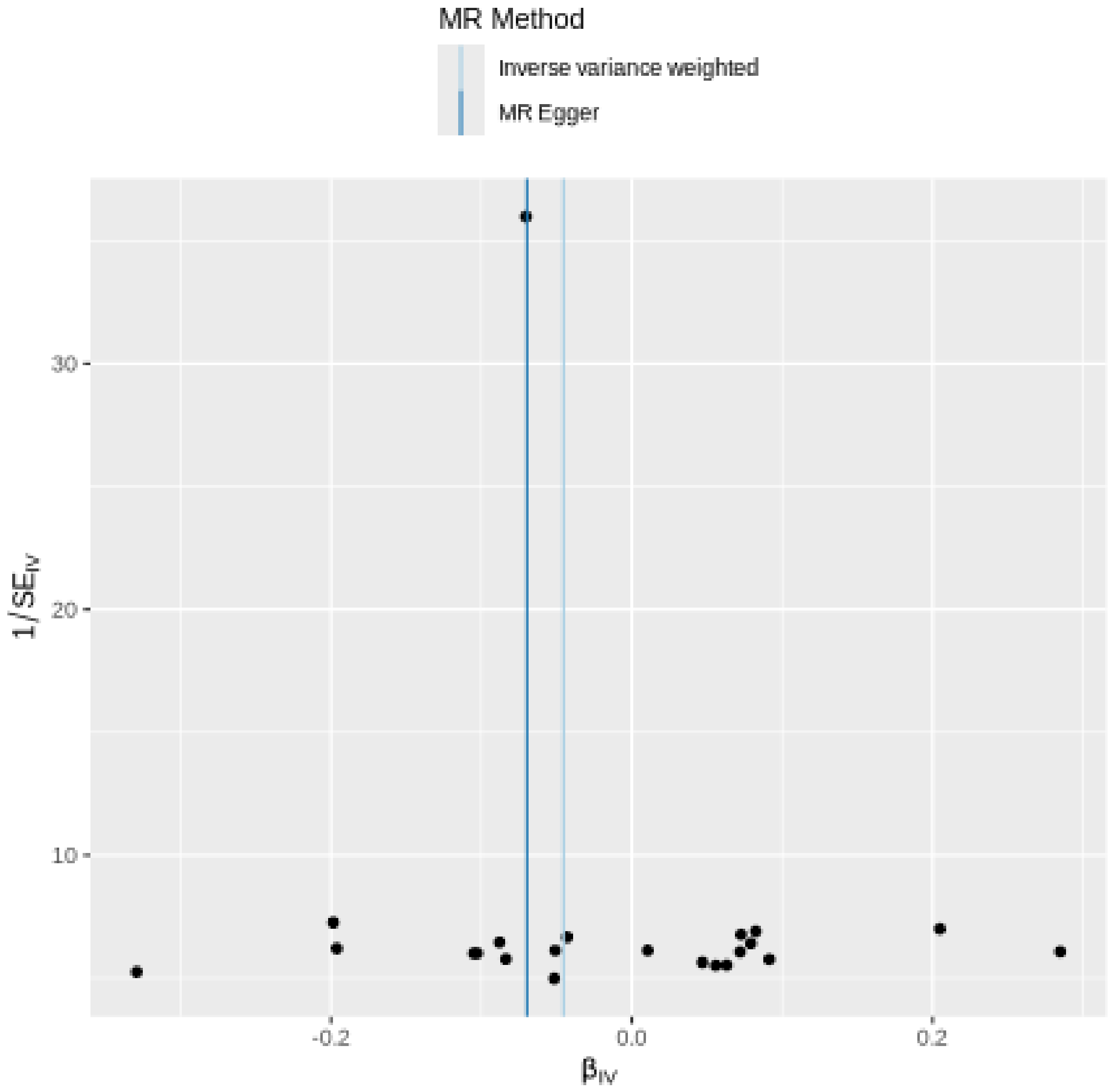
MR Method
Inverse variance weighted
MR Egger
30
20
10
1/SE_IV
-0.2
0.0
0.2
β_IV

### Leave-one-out sensitivity analysis

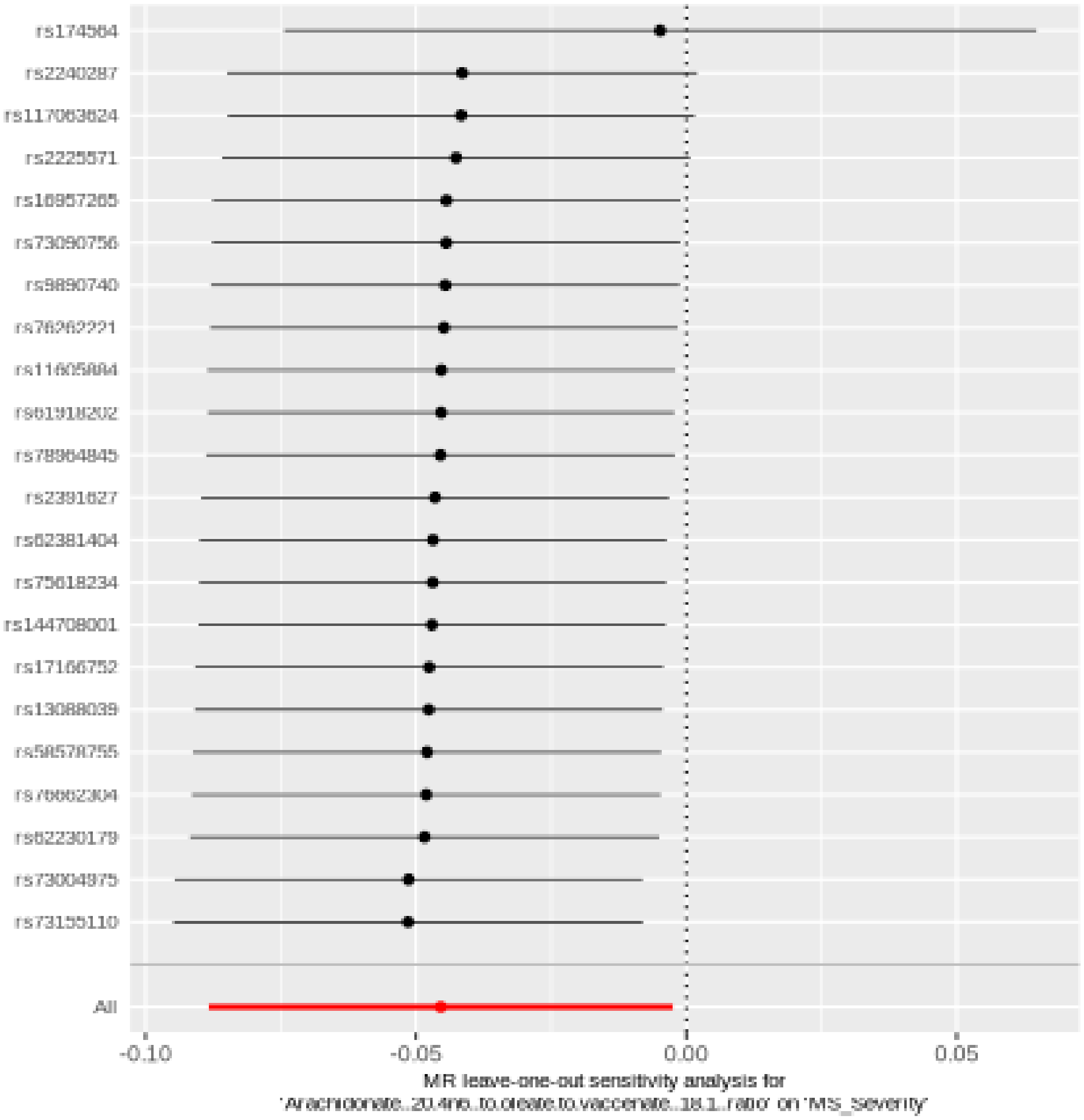

#### Phosphate to citrate ratio

| method | nsnp | b | se | pval |
| --- | --- | --- | --- | --- |
| MR Egger | 35 | 0.0563853 | 0.0623688 | 0.3725153 |
| Weighted median | 35 | -0.0178626 | 0.0379290 | 0.6376771 |
| Inverse variance weighted | 35 | -0.0574511 | 0.0268331 | 0.0322696 |
| Simple mode | 35 | 0.0045050 | 0.0728692 | 0.9510661 |
| Weighted mode | 35 | -0.0013622 | 0.0696062 | 0.9845003 |

#### Heterogeneity tests

| method | Q | Q_df | Q_pval |
| --- | --- | --- | --- |
| MR Egger | 24.99197 | 33 | 0.8401608 |
| Inverse variance weighted | 29.08007 | 34 | 0.7075150 |

#### Test for directional horizontal pleiotropy

| egger_intercept | se | pval |
| --- | --- | --- |
| -0.0150434 | 0.0074402 | 0.0513551 |

### Forest plot of single SNP MR

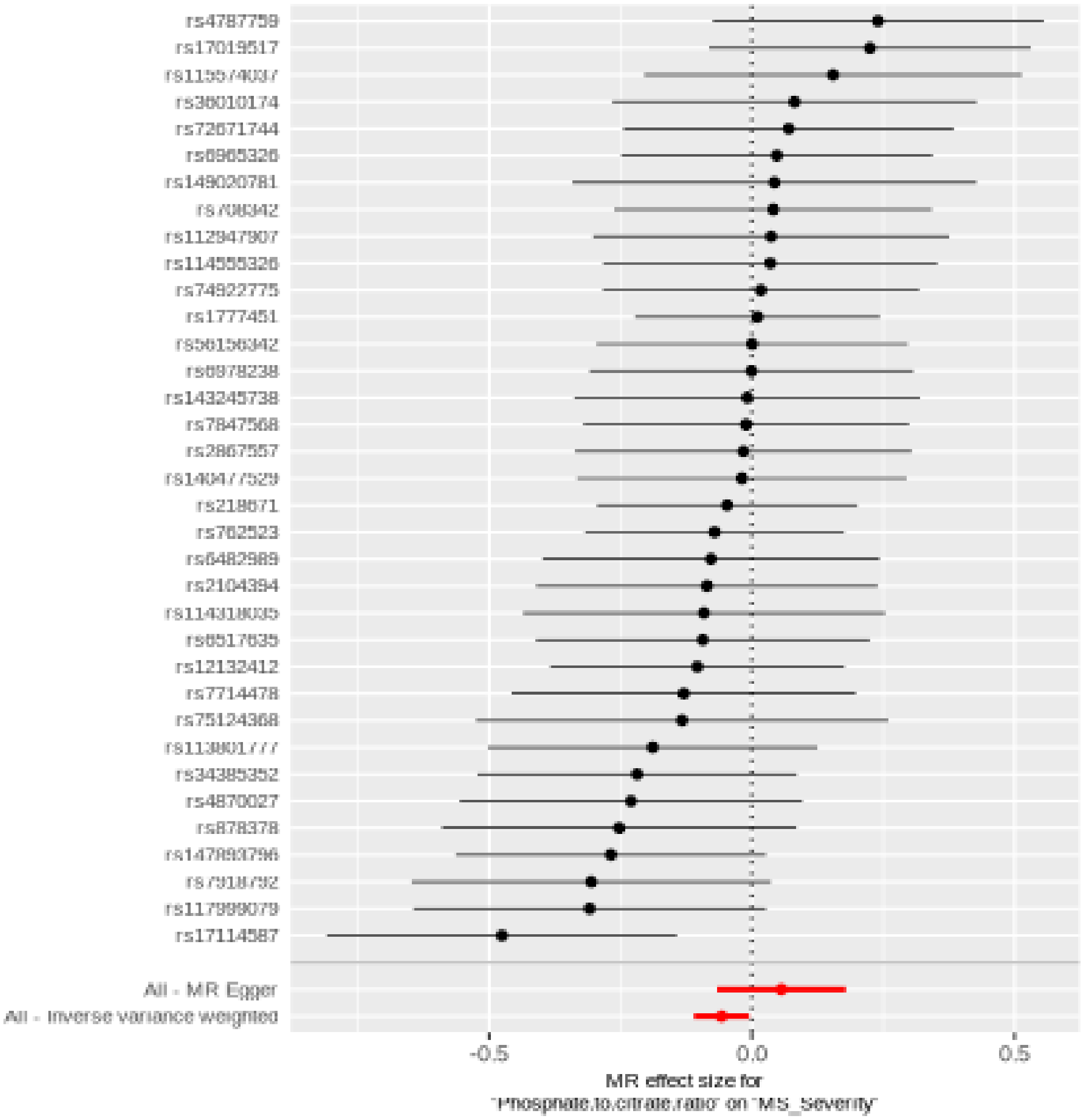

#### Comparison of results using different MR methods

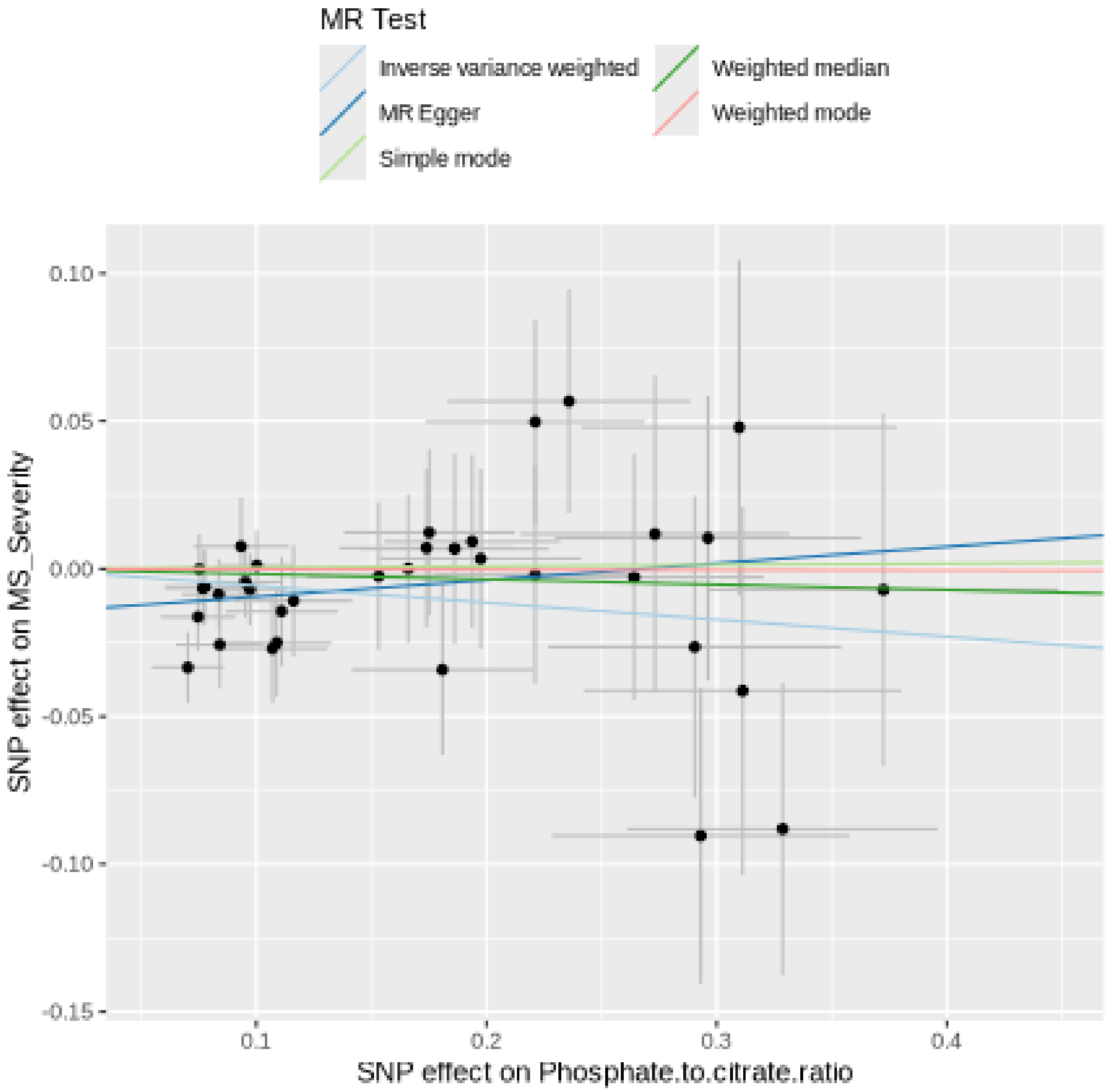

**Funnel plot**

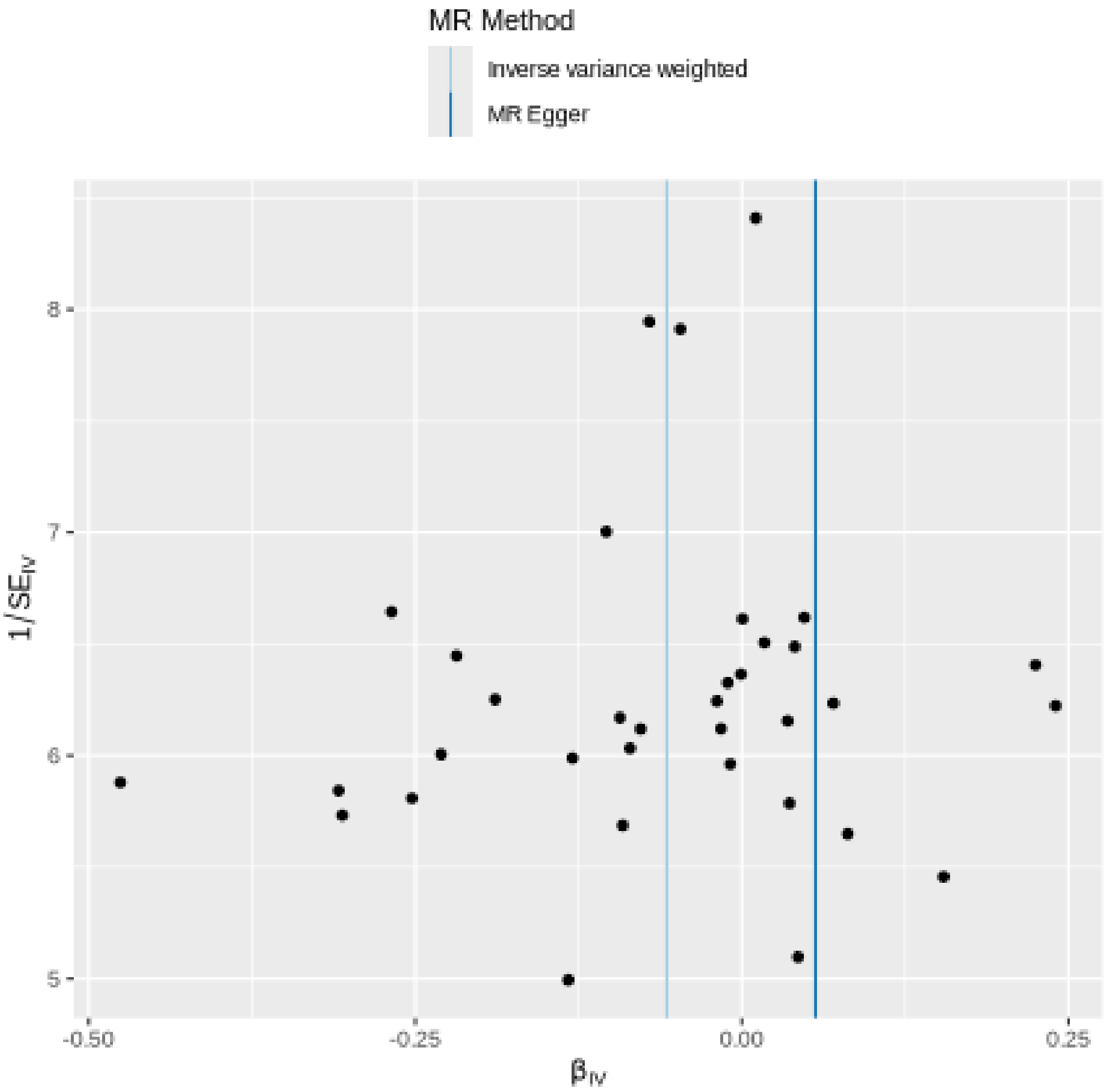

MR Method
Inverse variance weighted
MR Egger
8
7
6
5
$1/SE_{IV}$
-0.50
-0.25
0.00
0.25
$\beta_{IV}$

#### Leave-one-out sensitivity analysis

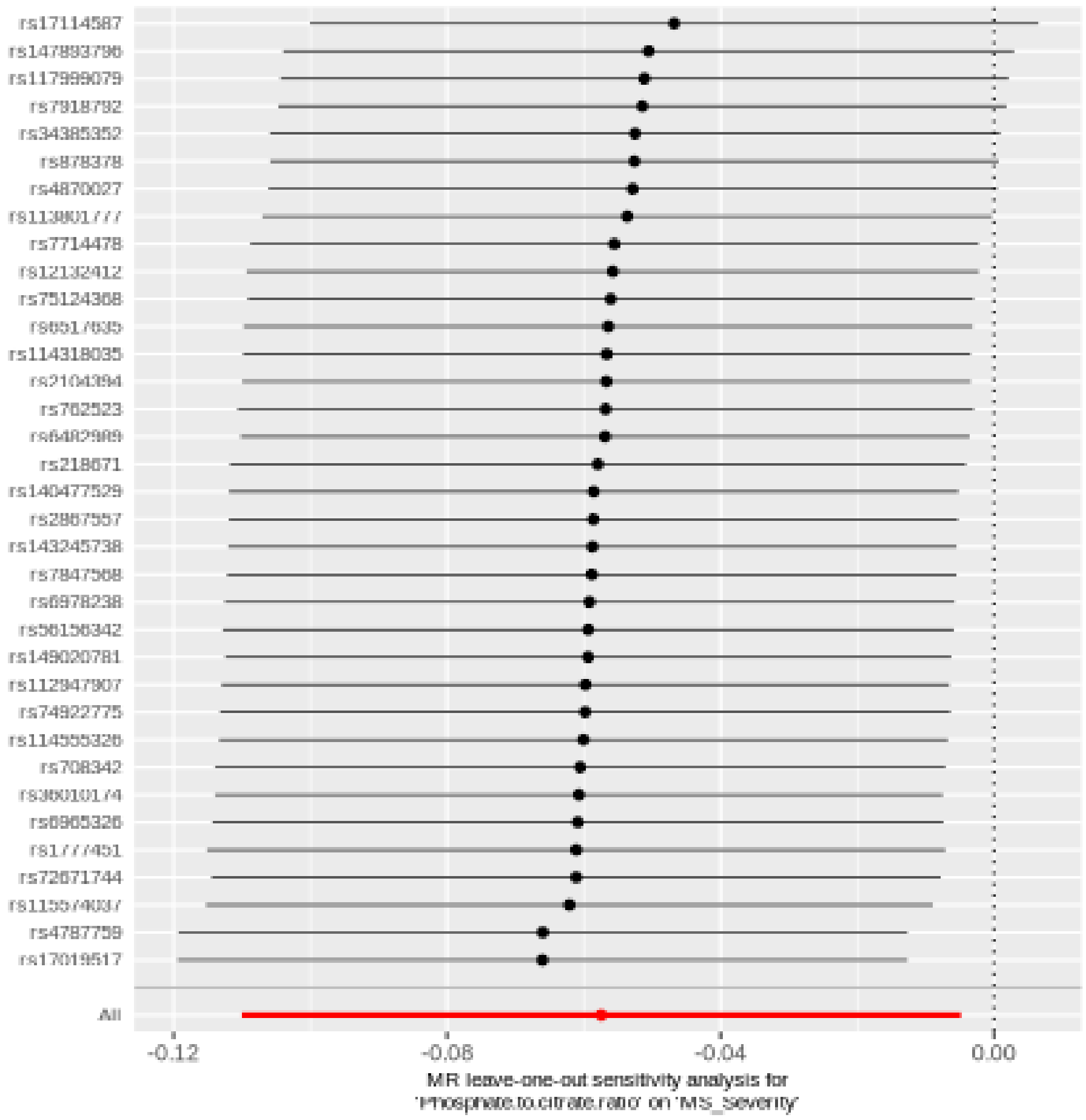

#### Serine to pyruvate ratio

| method | nsnp | b | se | pval |
|---|---|---|---|---|
| MR Egger | 25 | 0.1586919 | 0.0776319 | 0.0525516 |
| Weighted median | 25 | 0.0684904 | 0.0414959 | 0.0988335 |
| Inverse variance weighted | 25 | 0.0719696 | 0.0306313 | 0.0187963 |
| Simple mode | 25 | 0.0936987 | 0.0842161 | 0.2769016 |
| Weighted mode | 25 | 0.1108819 | 0.0725288 | 0.1393886 |

##### Heterogeneity tests

| method | Q | Q_df | Q_pval |
|---|---|---|---|
| MR Egger | 17.11015 | 23 | 0.8038114 |
| Inverse variance weighted | 18.58816 | 24 | 0.7735815 |

##### Test for directional horizontal pleiotropy

| egger_intercept | se | pval |
|---|---|---|
| -0.0111036 | 0.0091333 | 0.236418 |

#### Forest plot of single SNP MR

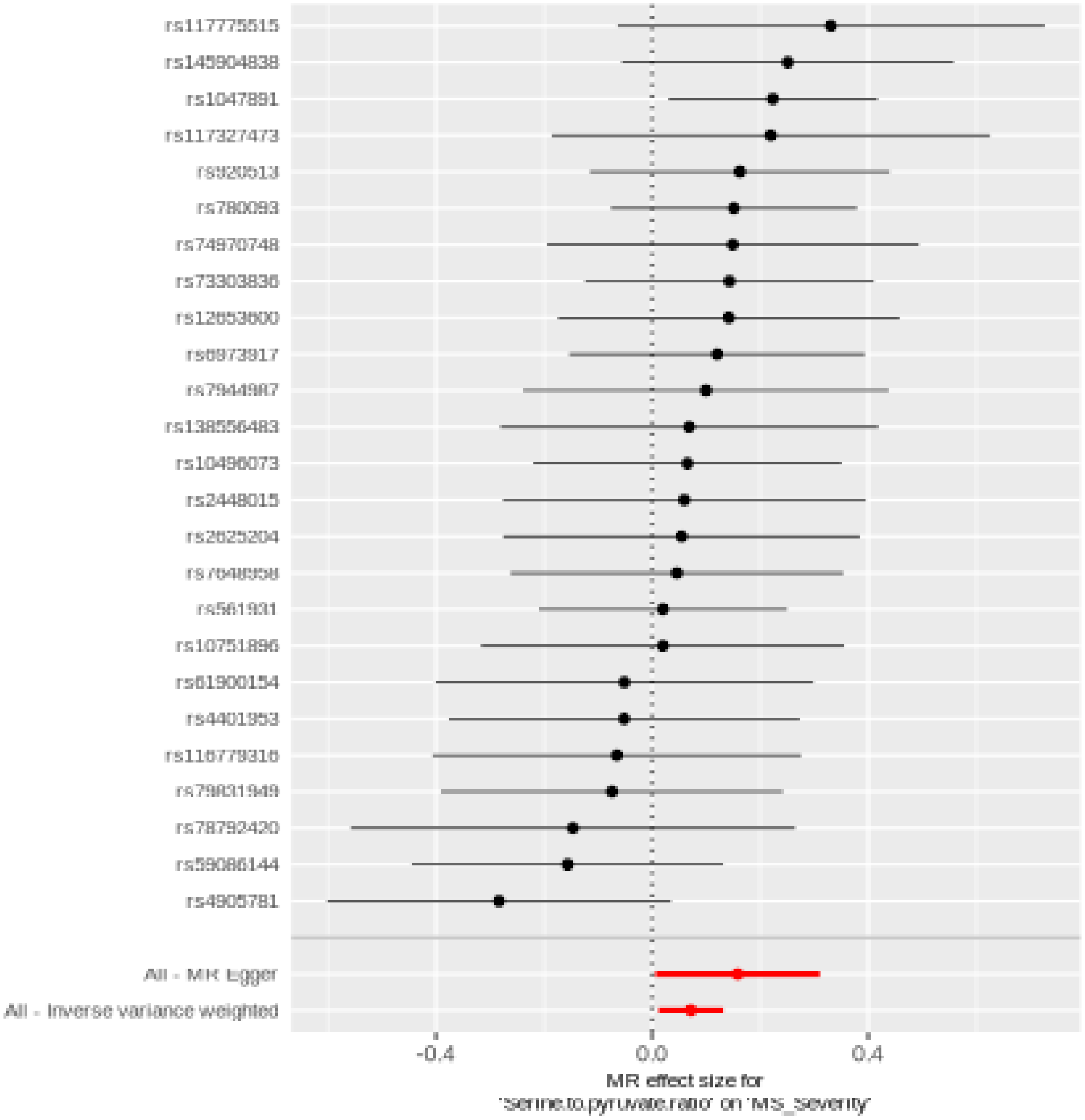

#### Comparison of results using different MR methods

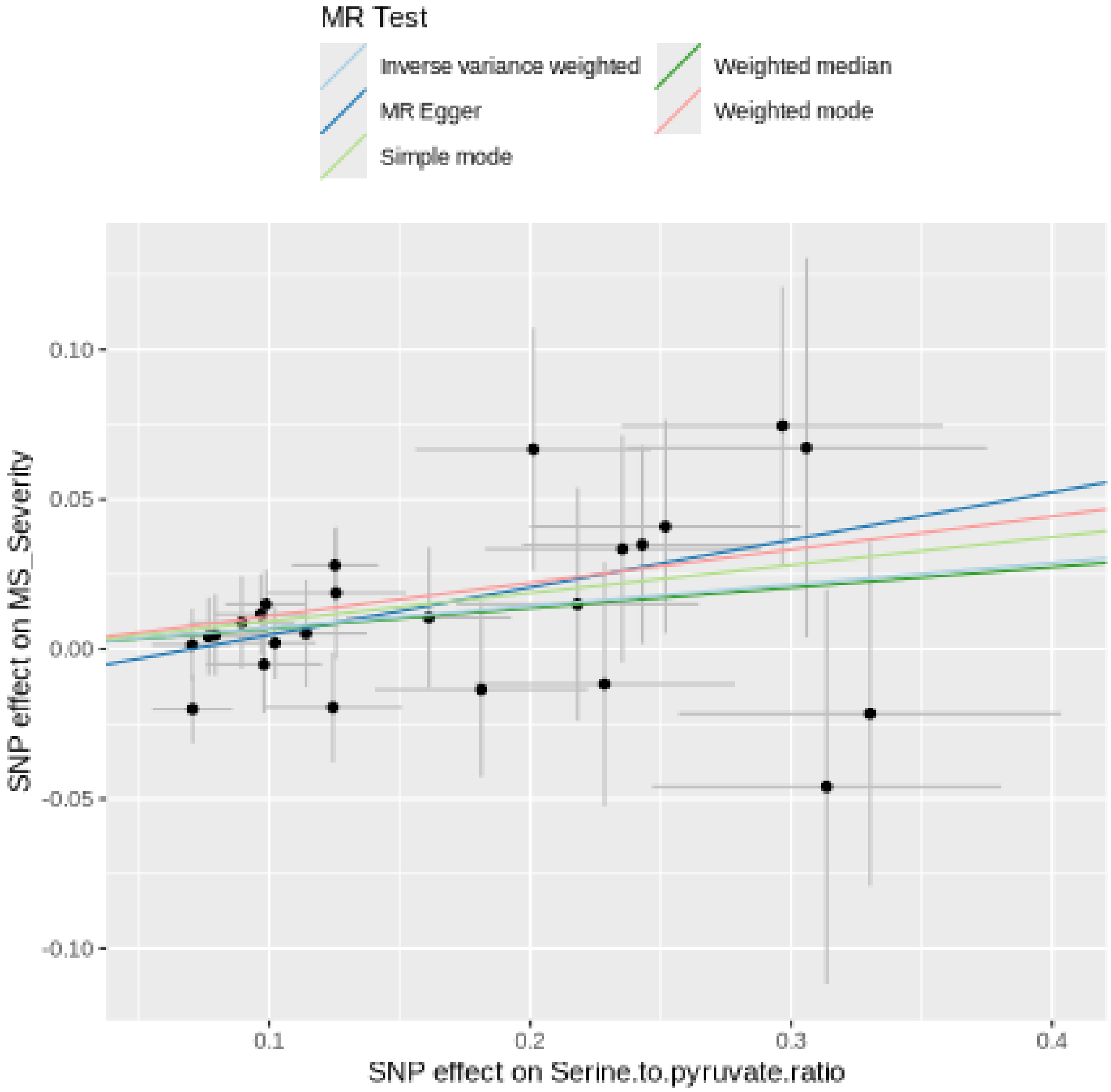
MR Test
Inverse variance weighted
MR Egger
Simple mode
Weighted median
Weighted mode
0.10
0.05
0.00
-0.05
-0.10
SNP effect on MS_Severity
0.1
0.2
0.3
0.4
SNP effect on Serine.to.pyruvate.ratio

#### Funnel plot

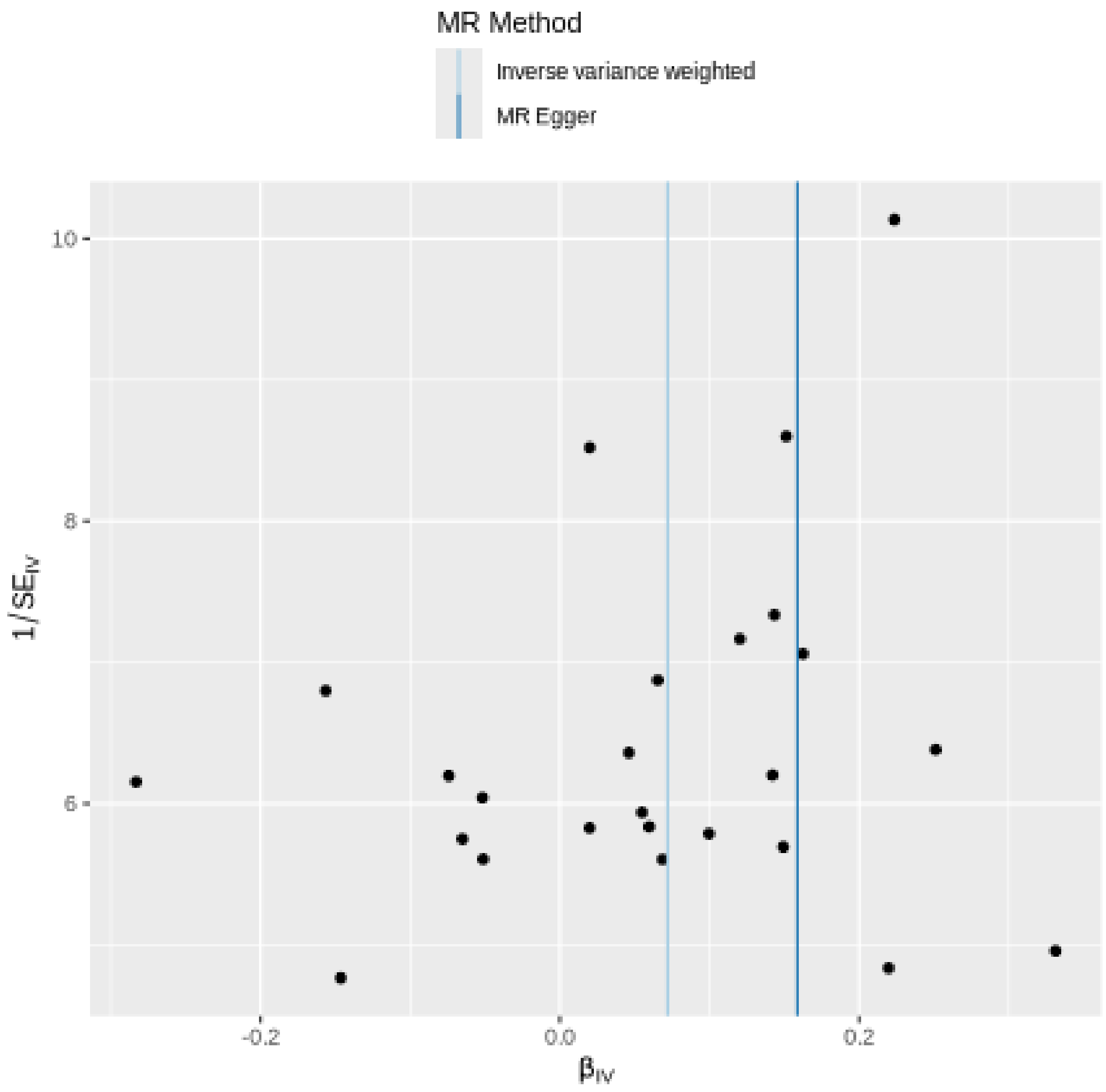

MR Method
Inverse variance weighted
MR Egger
10
8
6
1/SE$_{IV}$
-0.2
0.0
0.2
β$_{IV}$

### Leave-one-out sensitivity analysis

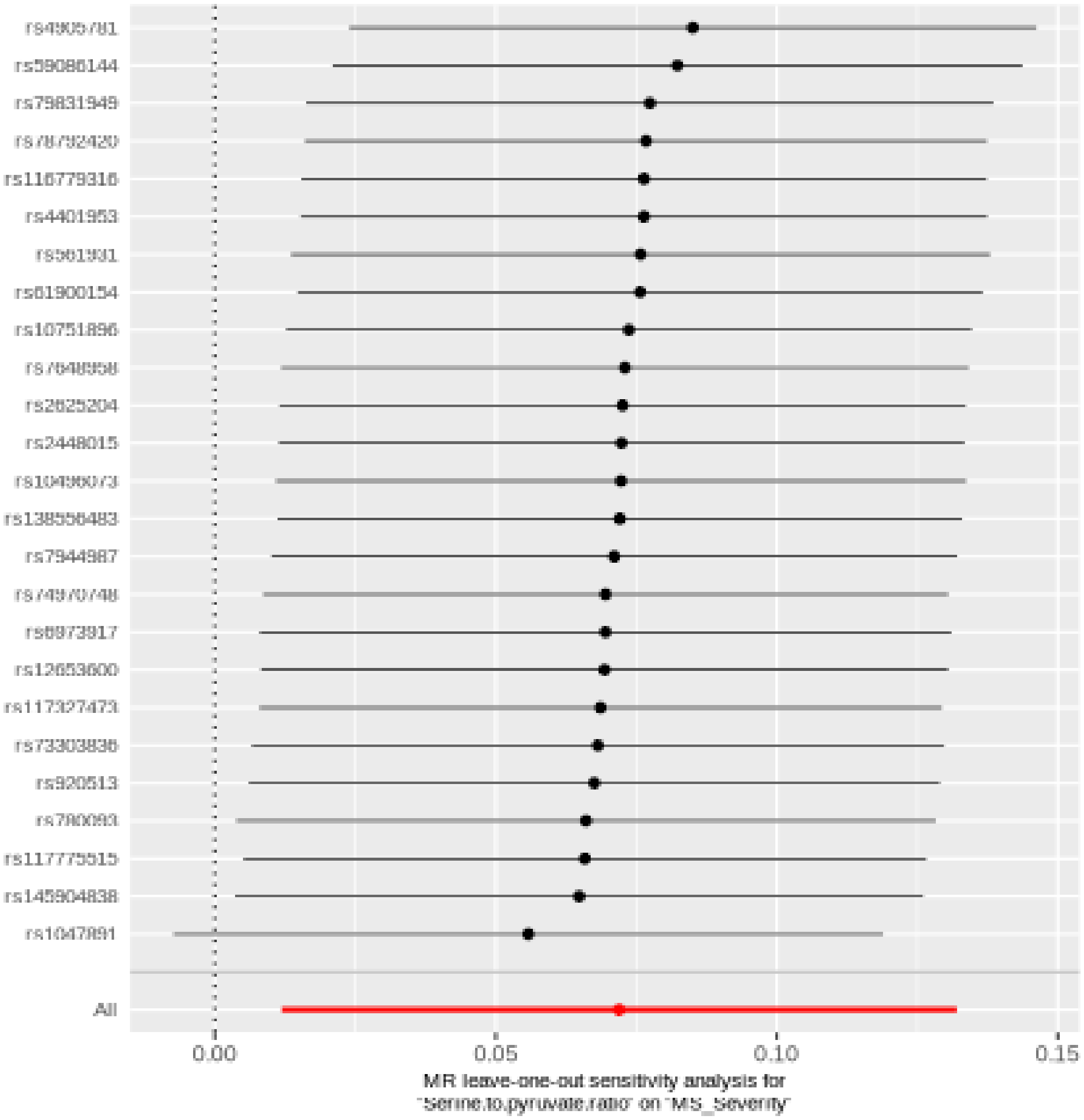

#### Oleoyl-linoleoyl-glycerol (18:1 to 18:2) [2] to linoleoyl-arachidonoyl-glycerol (18:2 to 20:4) [2] ratio

| method | nsnp | b | se | pval |
|---|---|---|---|---|
| MR Egger | 31 | 0.0926286 | 0.0321360 | 0.0073581 |
| Weighted median | 31 | 0.0670724 | 0.0274710 | 0.0146237 |
| Inverse variance weighted | 31 | 0.0441054 | 0.0192937 | 0.0222540 |
| Simple mode | 31 | 0.0554633 | 0.0714585 | 0.4437299 |
| Weighted mode | 31 | 0.0657616 | 0.0260444 | 0.0170914 |

##### Heterogeneity tests

| method | Q | Q_df | Q_pval |
|---|---|---|---|
| MR Egger | 25.28653 | 29 | 0.6632976 |
| Inverse variance weighted | 28.85138 | 30 | 0.5254137 |

##### Test for directional horizontal pleiotropy

| egger_intercept | se | pval |
|---|---|---|
| -0.0099322 | 0.0052605 | 0.0690569 |

### Forest plot of single SNP MR

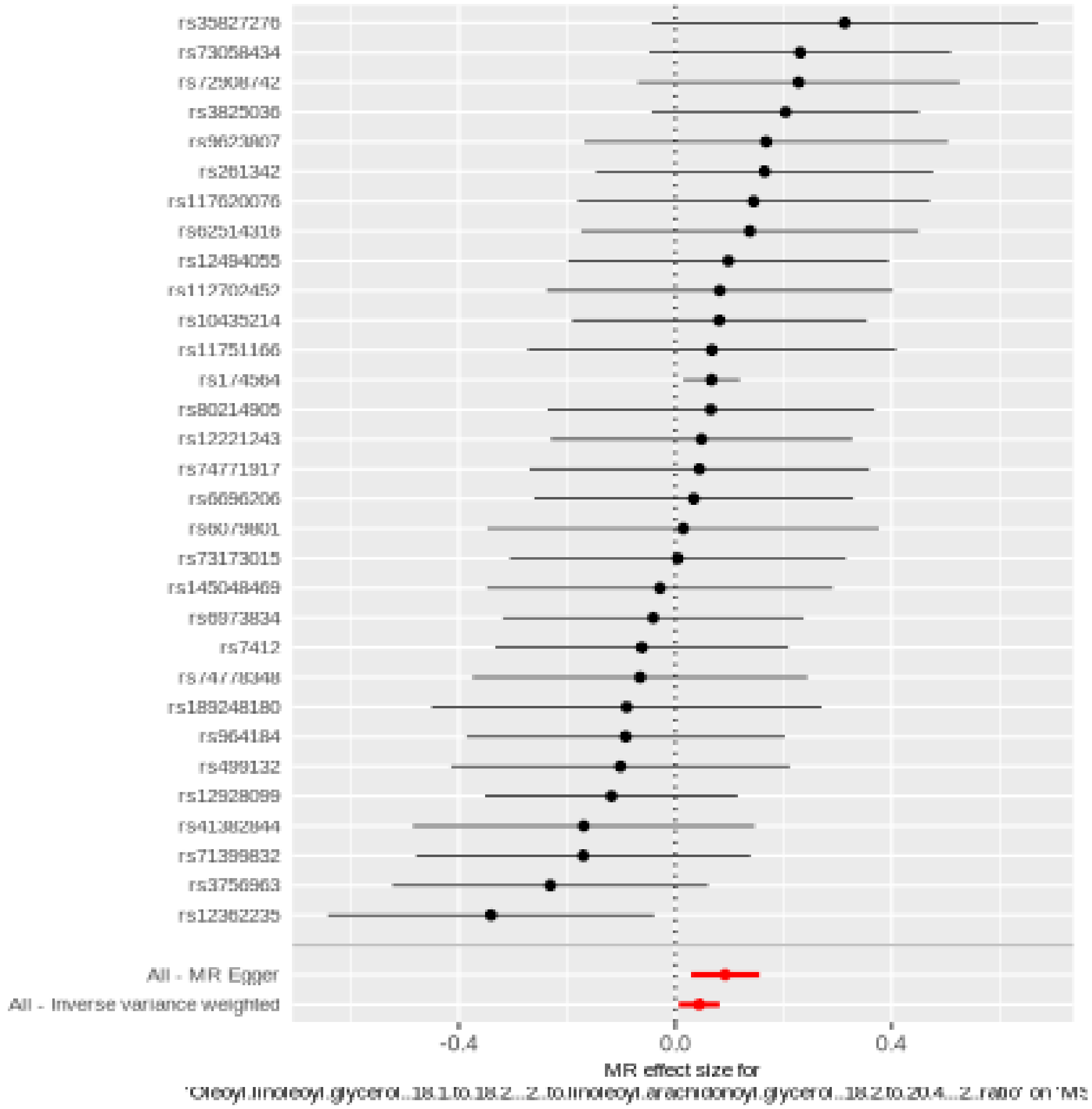

#### Comparison of results using different MR methods

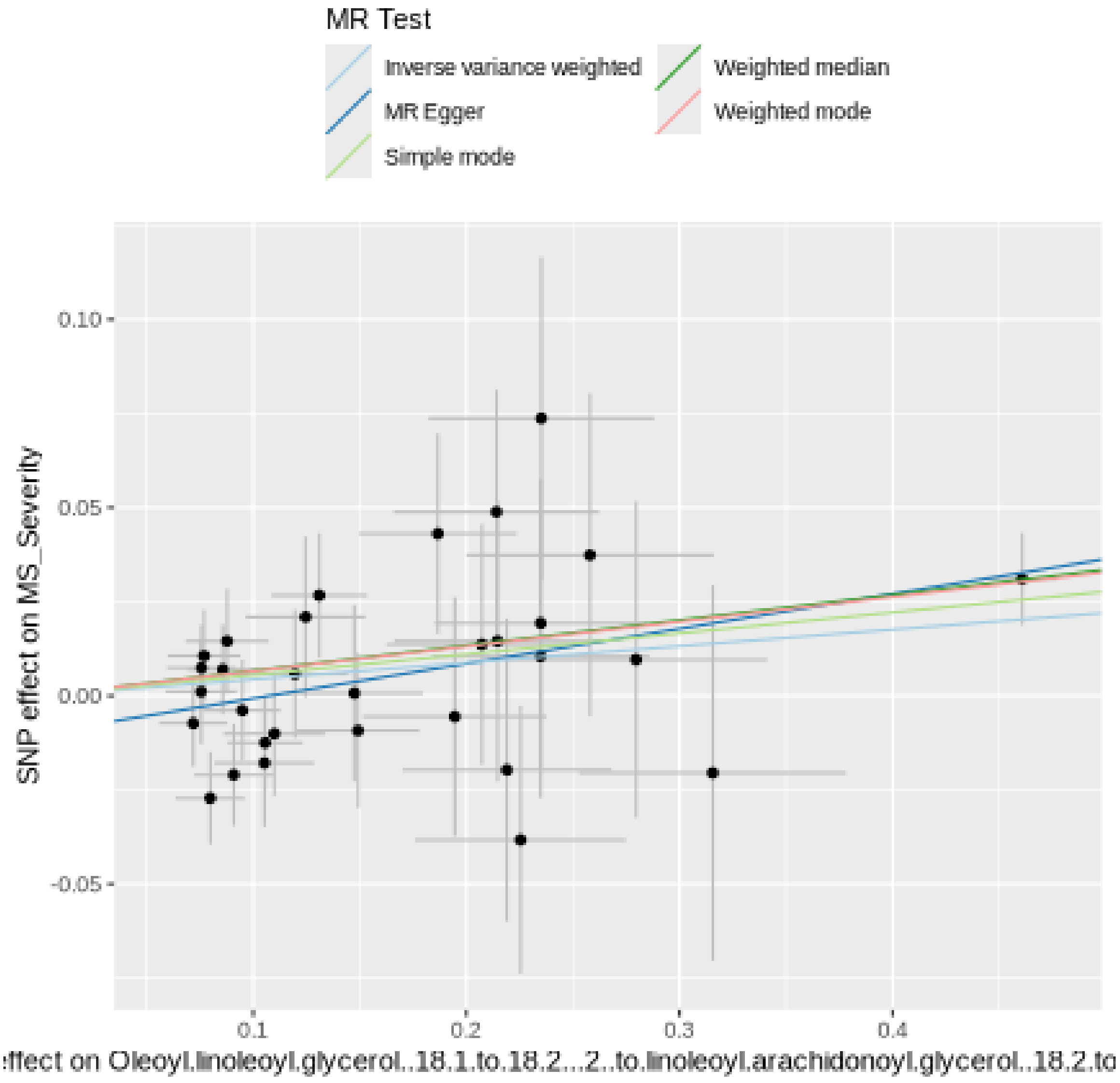

**Funnel plot**

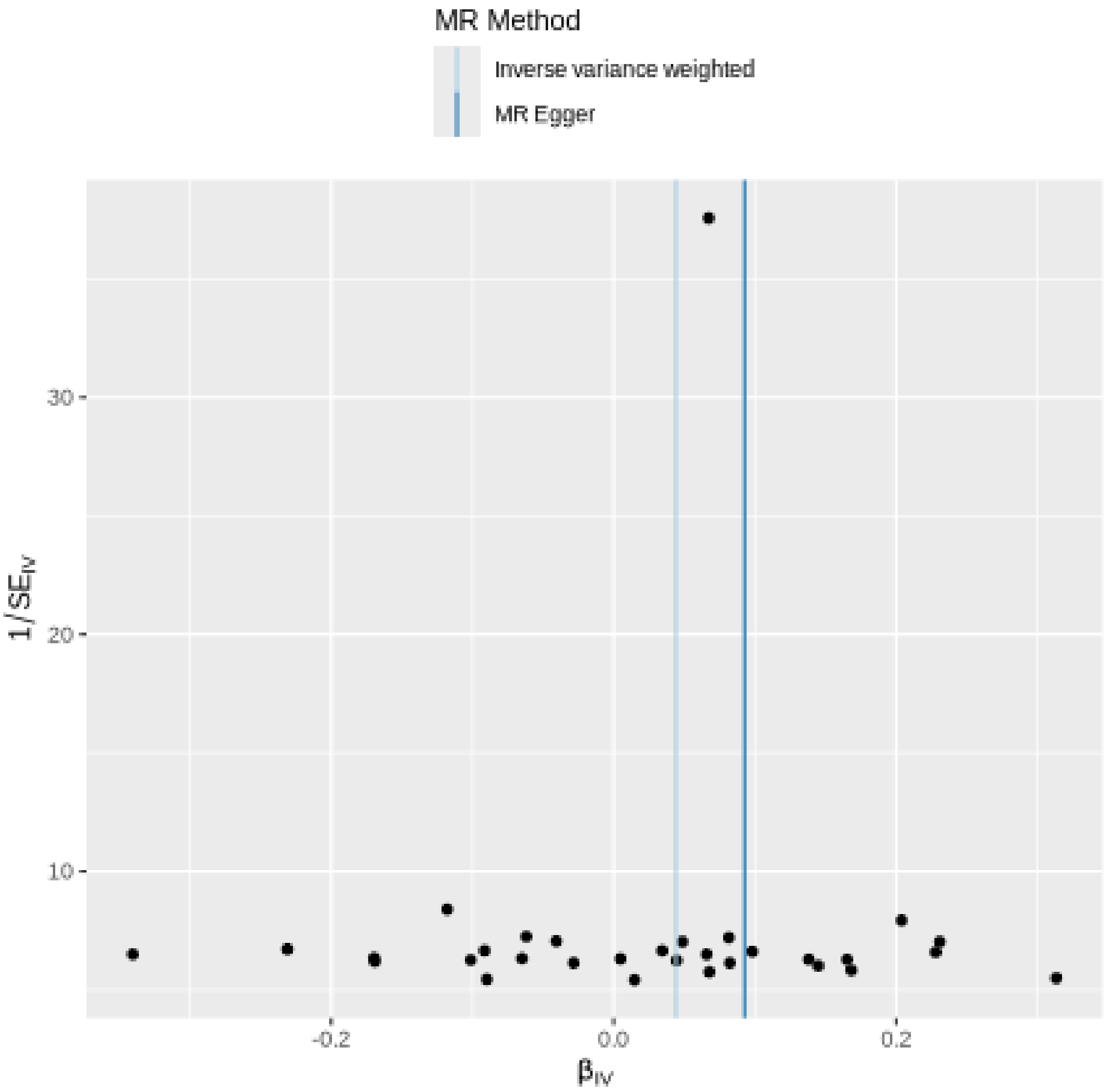
MR Method
Inverse variance weighted
MR Egger
30
20
10
1/SE_IV
-0.2
0.0
0.2
β_IV

#### Leave-one-out sensitivity analysis

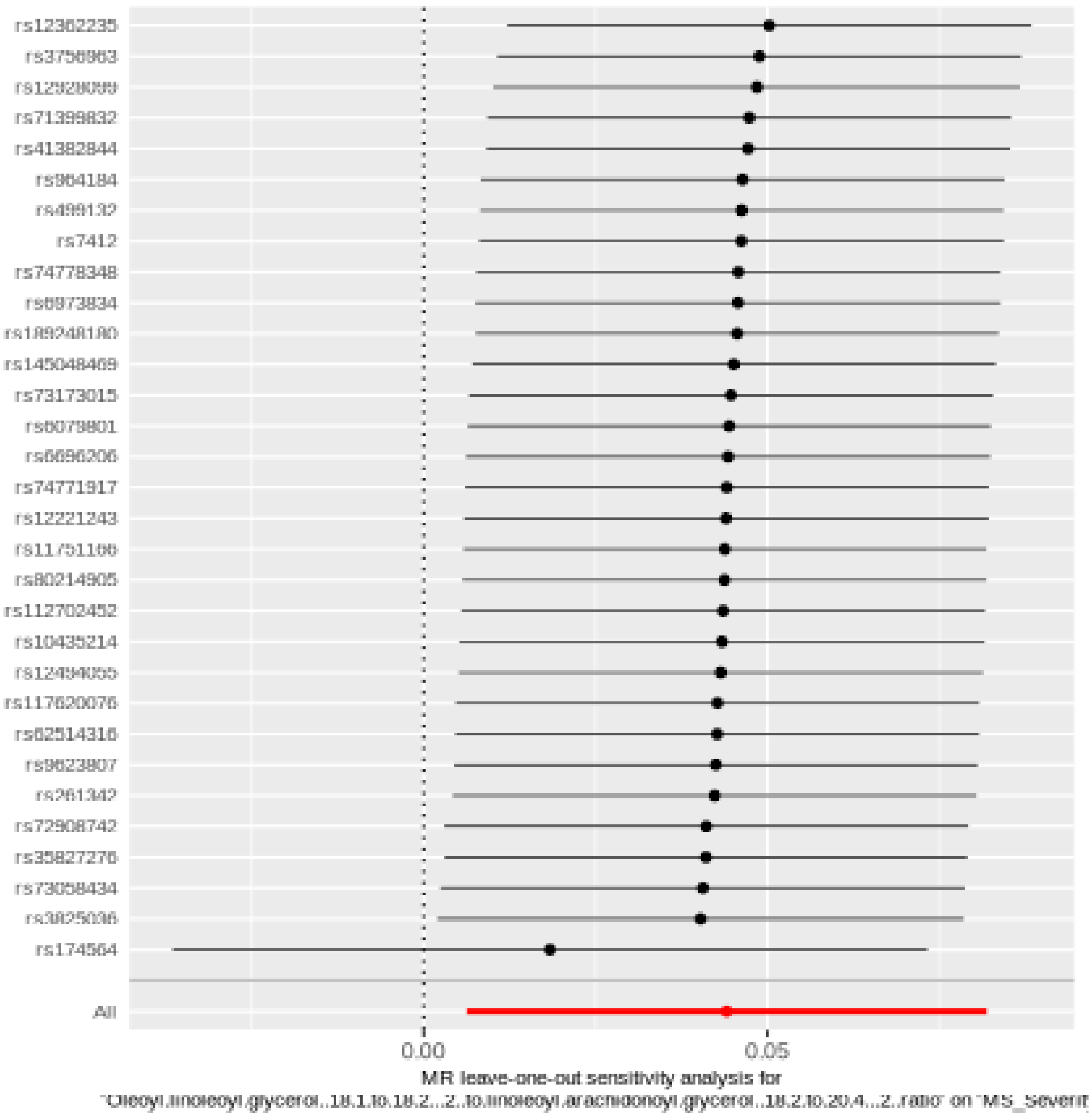

#### Glycine to phosphate ratio

| method | nsnp | b | se | pval |
|---|---|---|---|---|
| MR Egger | 37 | 0.0567782 | 0.0272057 | 0.0442376 |
| Weighted median | 37 | 0.0533088 | 0.0232418 | 0.0218098 |
| Inverse variance weighted | 37 | 0.0417710 | 0.0175454 | 0.0172776 |
| Simple mode | 37 | 0.0711956 | 0.0683517 | 0.3045391 |
| Weighted mode | 37 | 0.0575440 | 0.0233305 | 0.0185399 |

#### Heterogeneity tests

| method | Q | Q_df | Q_pval |
|---|---|---|---|
| MR Egger | 32.23045 | 35 | 0.6025137 |
| Inverse variance weighted | 32.75140 | 36 | 0.6239000 |

#### Test for directional horizontal pleiotropy

| egger_intercept | se | pval |
|---|---|---|
| -0.0032385 | 0.0044868 | 0.4752288 |

### Forest plot of single SNP MR

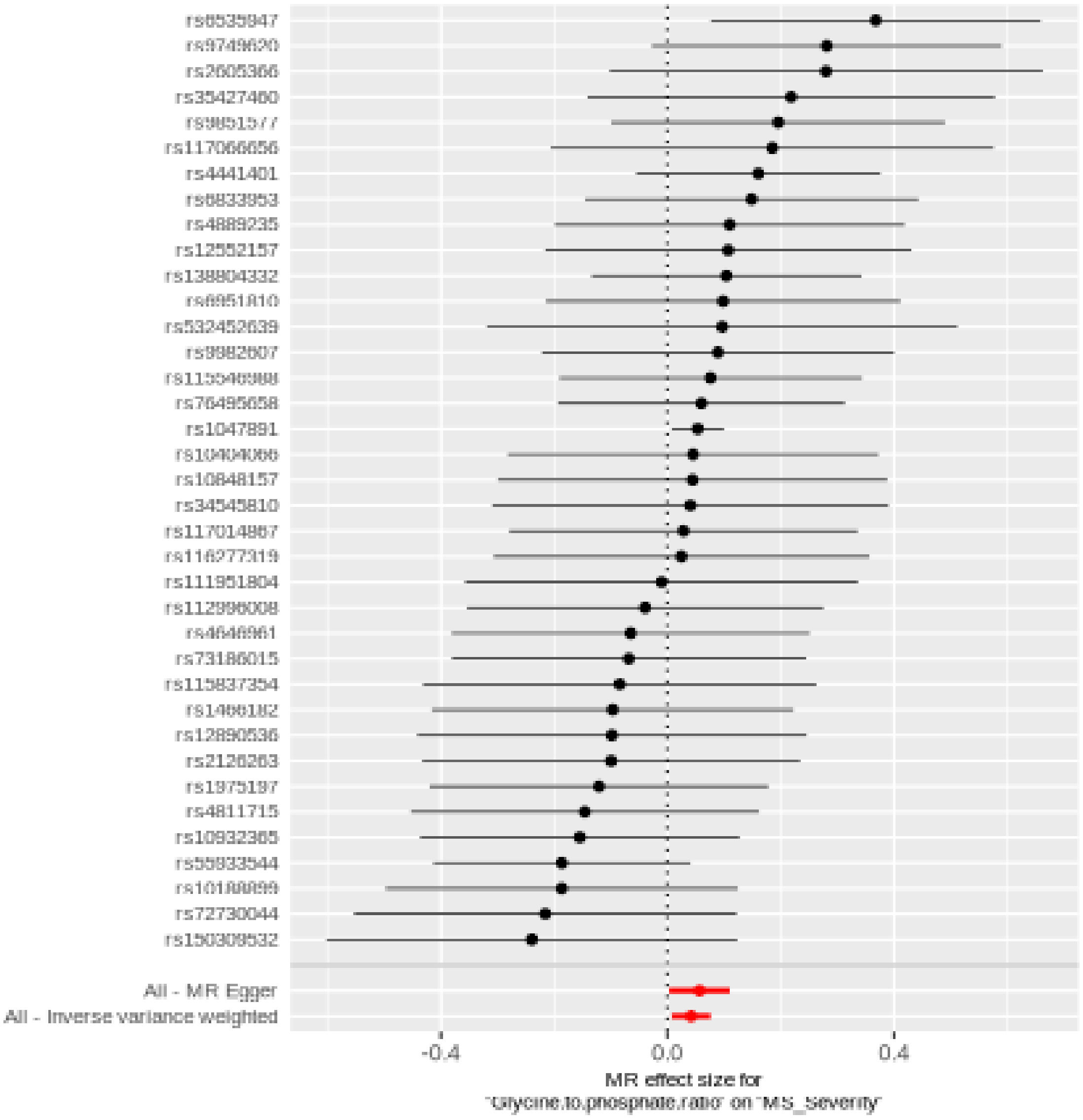

#### Comparison of results using different MR methods

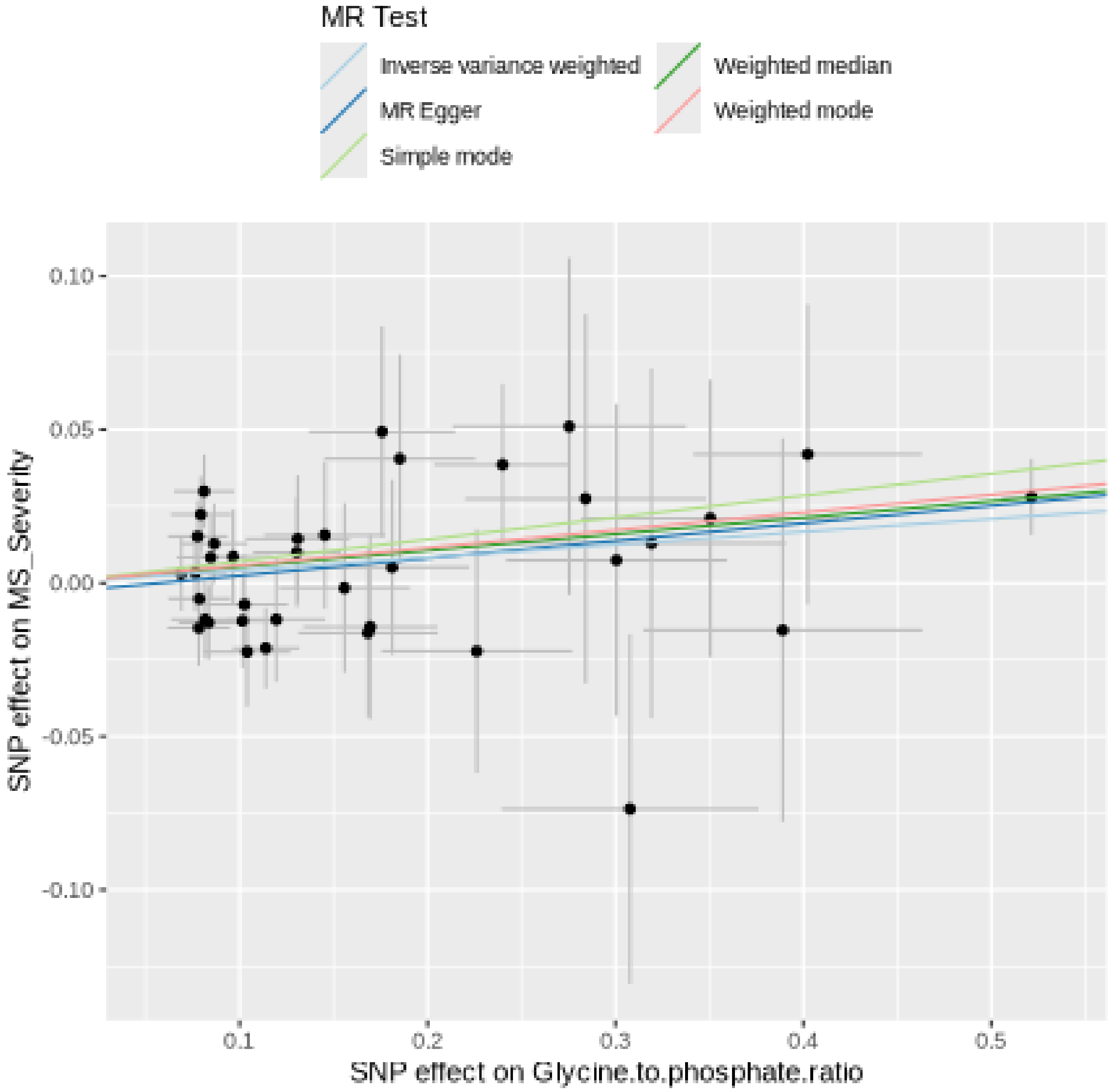

#### Funnel plot

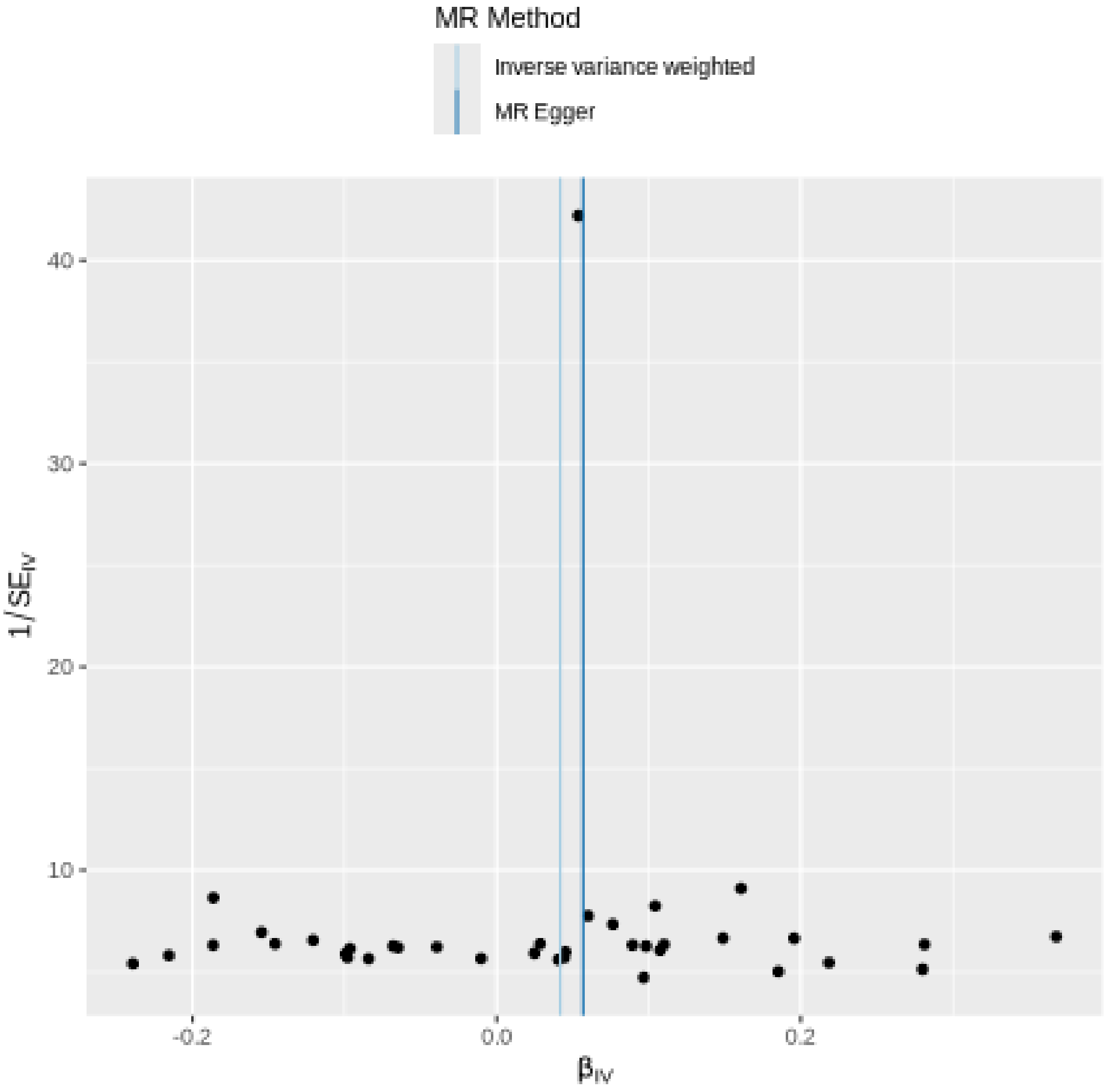

MR Method
Inverse variance weighted
MR Egger
40
30
20
10
1/SE$_{IV}$
-0.2
0.0
0.2
β$_{IV}$

#### Leave-one-out sensitivity analysis

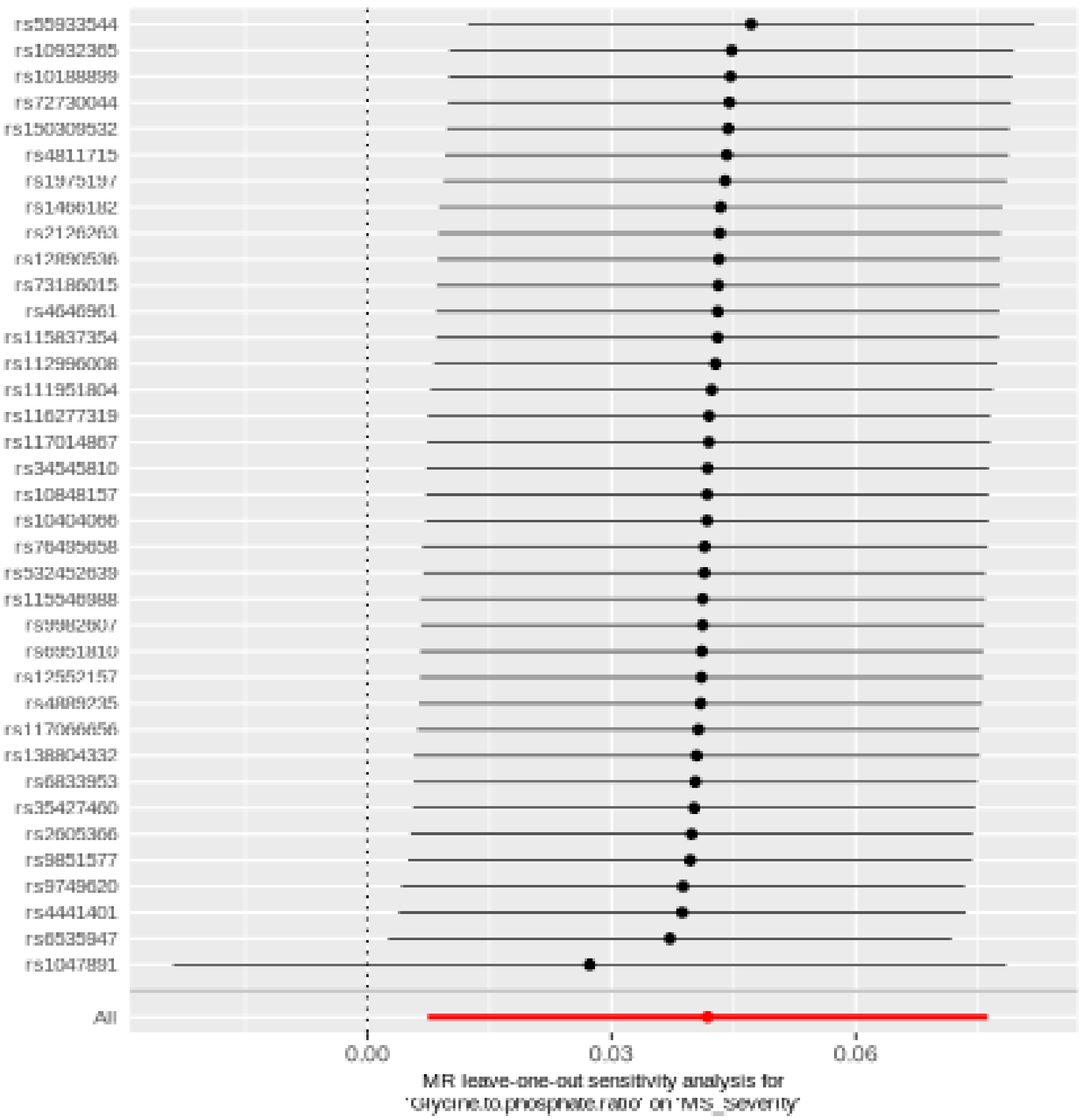

rs55933544
rs10932365
rs10188899
rs72730044
rs150308532
rs4811715
rs1975197
rs1466182
rs2126263
rs12890536
rs73186015
rs4646961
rs115837354
rs112996008
rs111951804
rs116277319
rs117014867
rs34545810
rs10848157
rs10404066
rs76485658
rs532452639
rs115546988
rs9982607
rs6851810
rs12552157
rs4889235
rs117066656
rs138804332
rs6833953
rs35427460
rs2605366
rs9851577
rs9749620
rs4441401
rs6535947
rs1047891
All
0.00
0.03
0.06
MR leave-one-out sensitivity analysis for
'Glycine.to.phosphate.ratio' on 'MS_Severity'

#### Alpha-ketoglutarate to proline ratio

| method | nsnp | b | se | pval |
|---|---|---|---|---|
| MR Egger | 27 | -0.0685069 | 0.0600417 | 0.2646870 |
| Weighted median | 27 | -0.0319613 | 0.0404996 | 0.4300089 |
| Inverse variance weighted | 27 | -0.0720328 | 0.0287358 | 0.0121854 |
| Simple mode | 27 | -0.0387442 | 0.0738690 | 0.6043754 |
| Weighted mode | 27 | -0.0387442 | 0.0498335 | 0.4438972 |

#### Heterogeneity tests

| method | Q | Q_df | Q_pval |
|---|---|---|---|
| MR Egger | 28.94994 | 25 | 0.2660141 |
| Inverse variance weighted | 28.95518 | 26 | 0.3131243 |

#### Test for directional horizontal pleiotropy

| egger_intercept | se | pval |
|---|---|---|
| -0.0005353 | 0.0079563 | 0.9468935 |

### Forest plot of single SNP MR

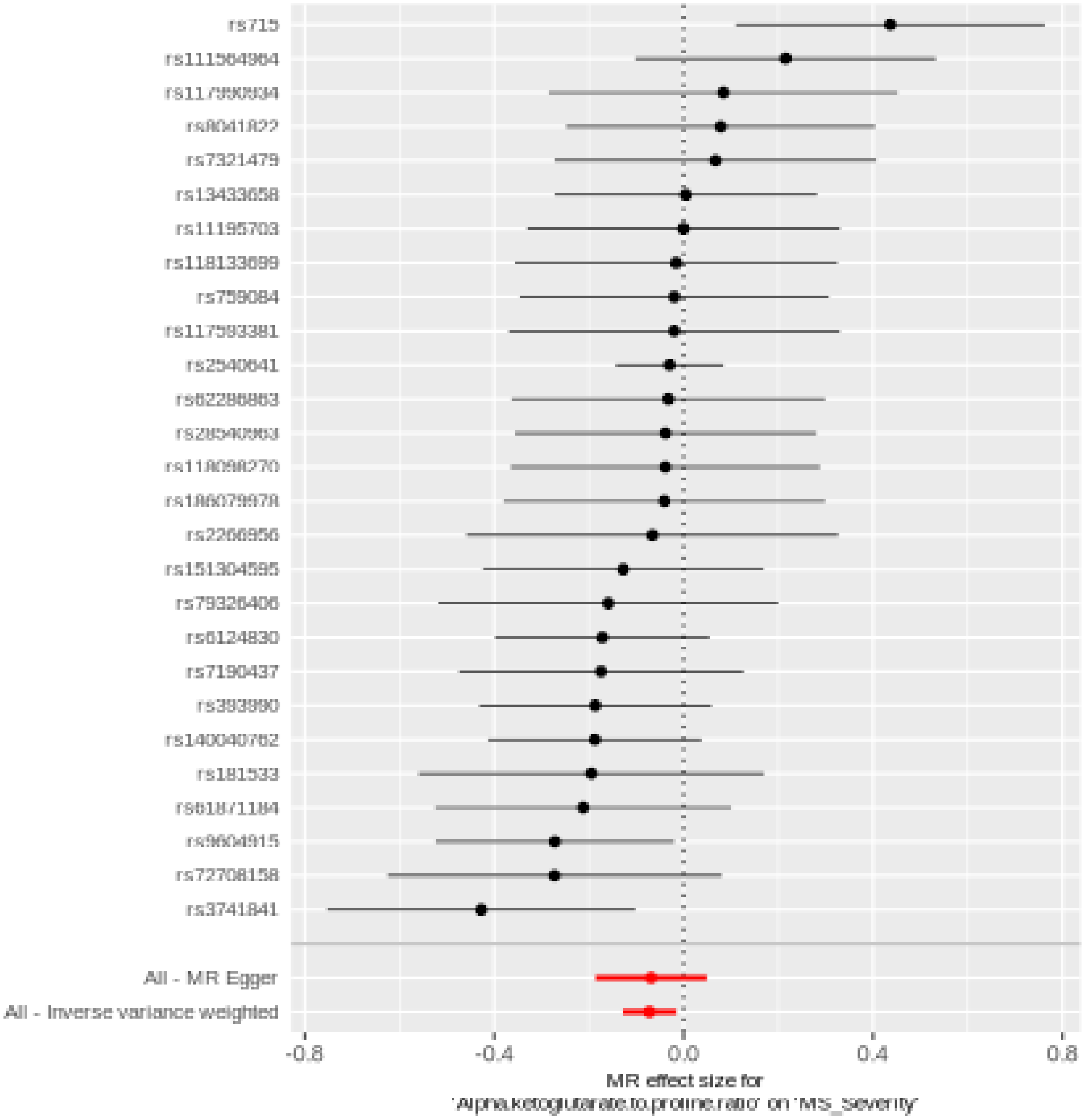

#### Comparison of results using different MR methods

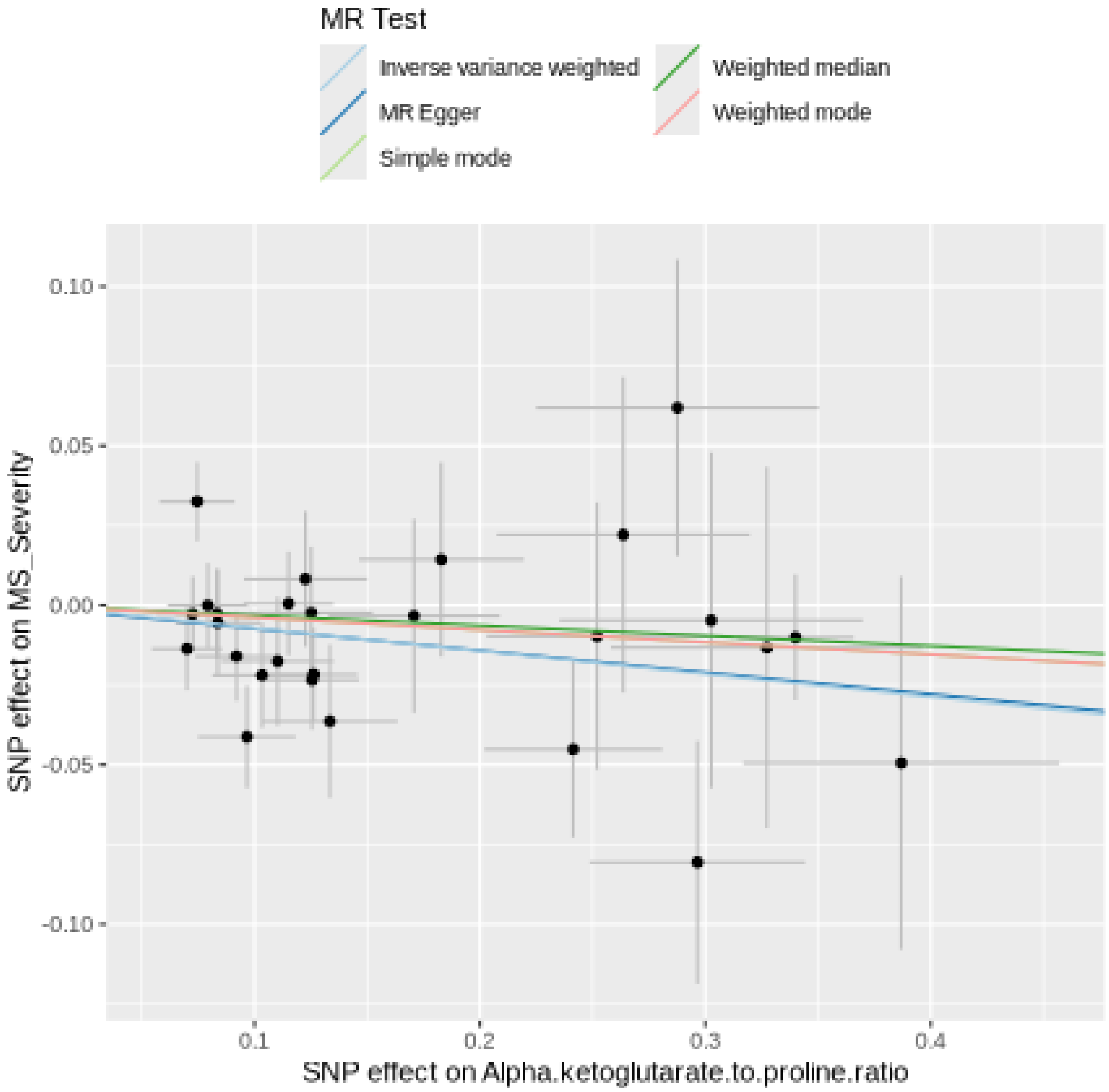

#### Funnel plot

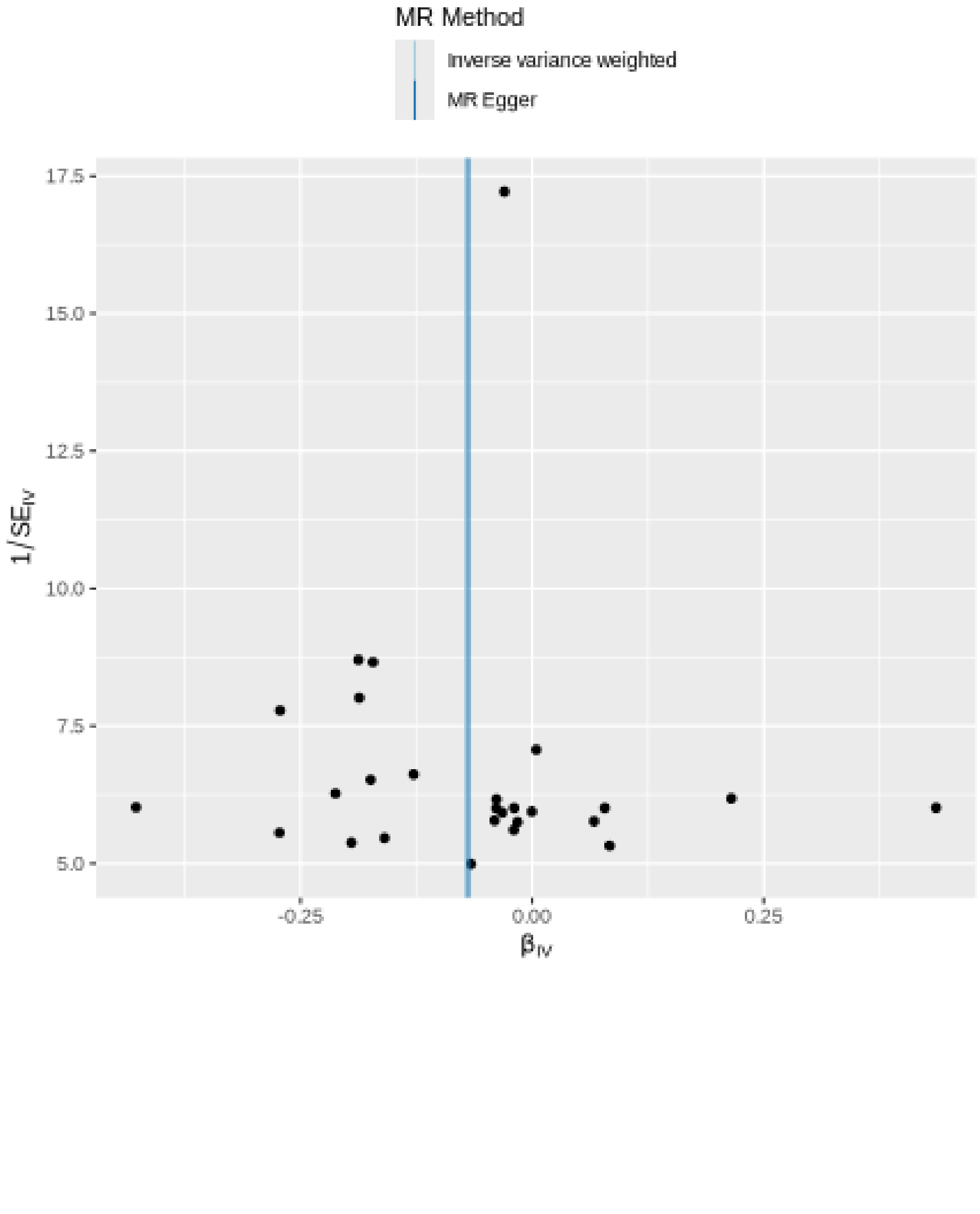
MR Method
Inverse variance weighted
MR Egger
17.5
15.0
12.5
10.0
7.5
5.0
1/SE$_{IV}$
-0.25
0.00
0.25
β$_{IV}$

#### Leave-one-out sensitivity analysis

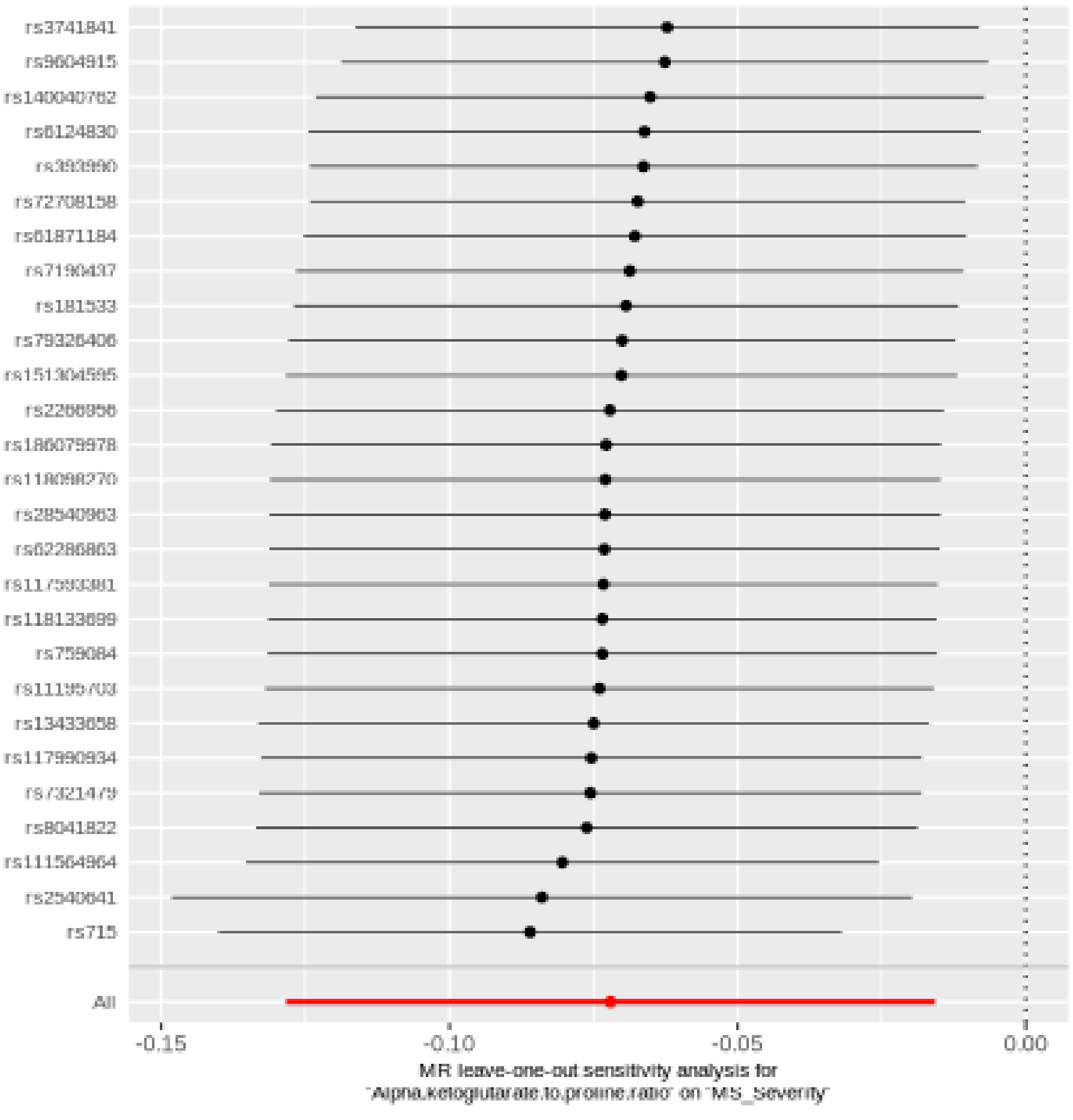

#### Arachidonate (20:4n6) to linoleate (18:2n6) ratio

| method | nsnp | b | se | pval |
|---|---|---|---|---|
| MR Egger | 27 | -0.0826390 | 0.0362025 | 0.0312208 |
| Weighted median | 27 | -0.0720660 | 0.0294452 | 0.0143864 |
| Inverse variance weighted | 27 | -0.0465452 | 0.0219682 | 0.0341114 |
| Simple mode | 27 | -0.0069505 | 0.0725887 | 0.9244516 |
| Weighted mode | 27 | -0.0695039 | 0.0293510 | 0.0256023 |

#### Heterogeneity tests

| method | Q | Q_df | Q_pval |
|---|---|---|---|
| MR Egger | 24.47597 | 25 | 0.4920271 |
| Inverse variance weighted | 26.04765 | 26 | 0.4604880 |

#### Test for directional horizontal pleiotropy

| egger_intercept | se | pval |
|---|---|---|
| 0.0069122 | 0.0055136 | 0.2215611 |

### Forest plot of single SNP MR

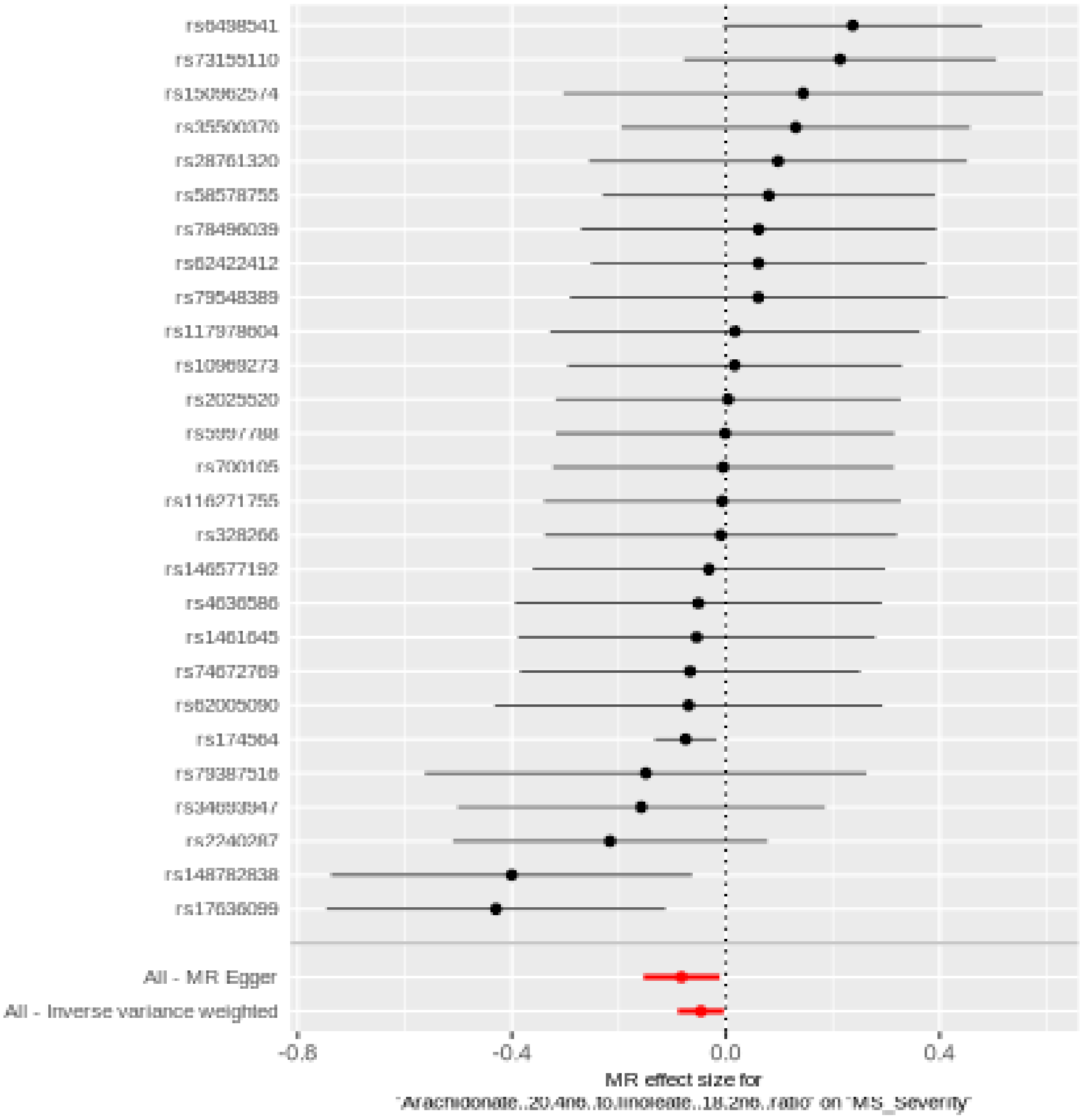

#### Comparison of results using different MR methods

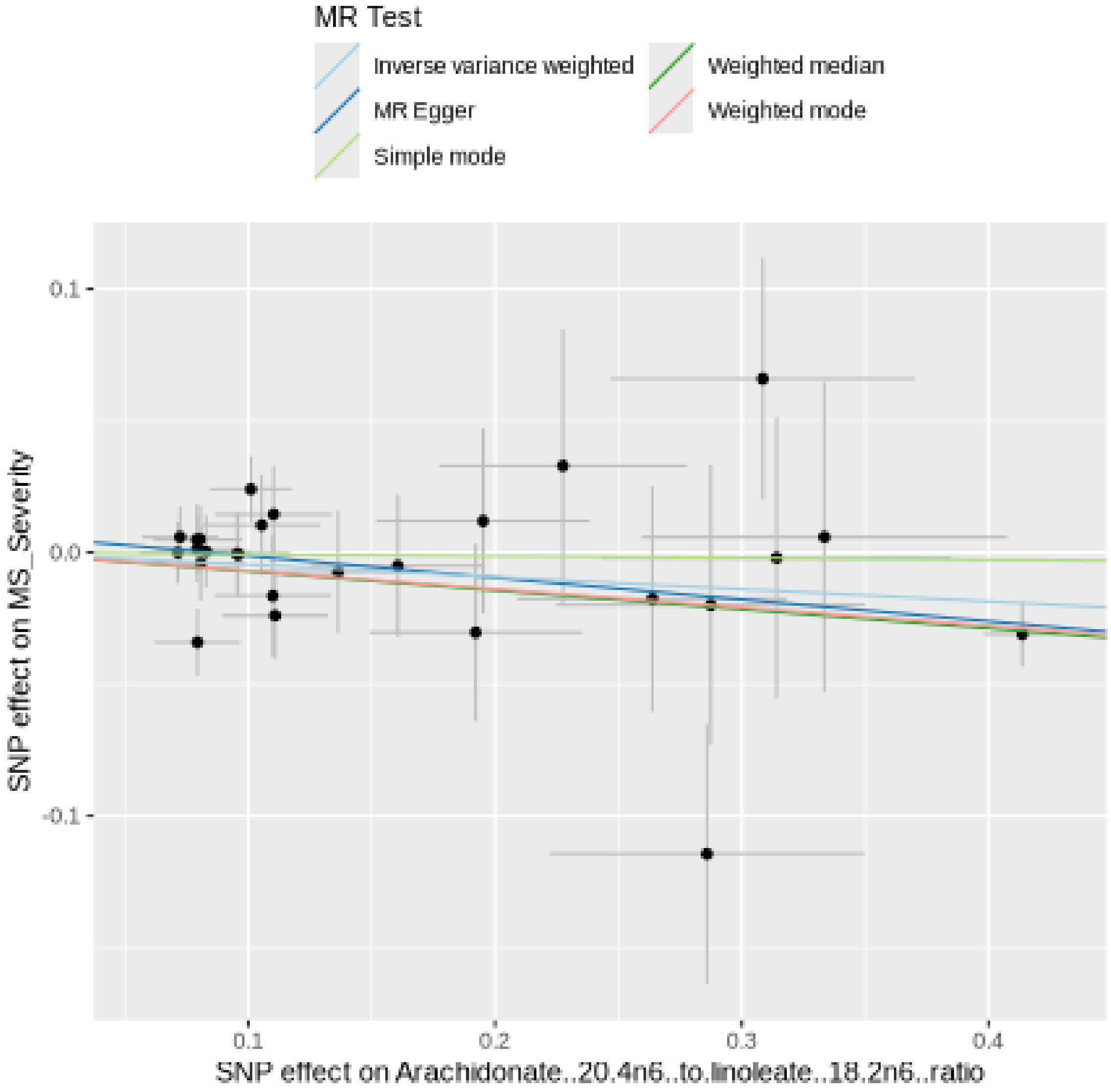

#### Funnel plot

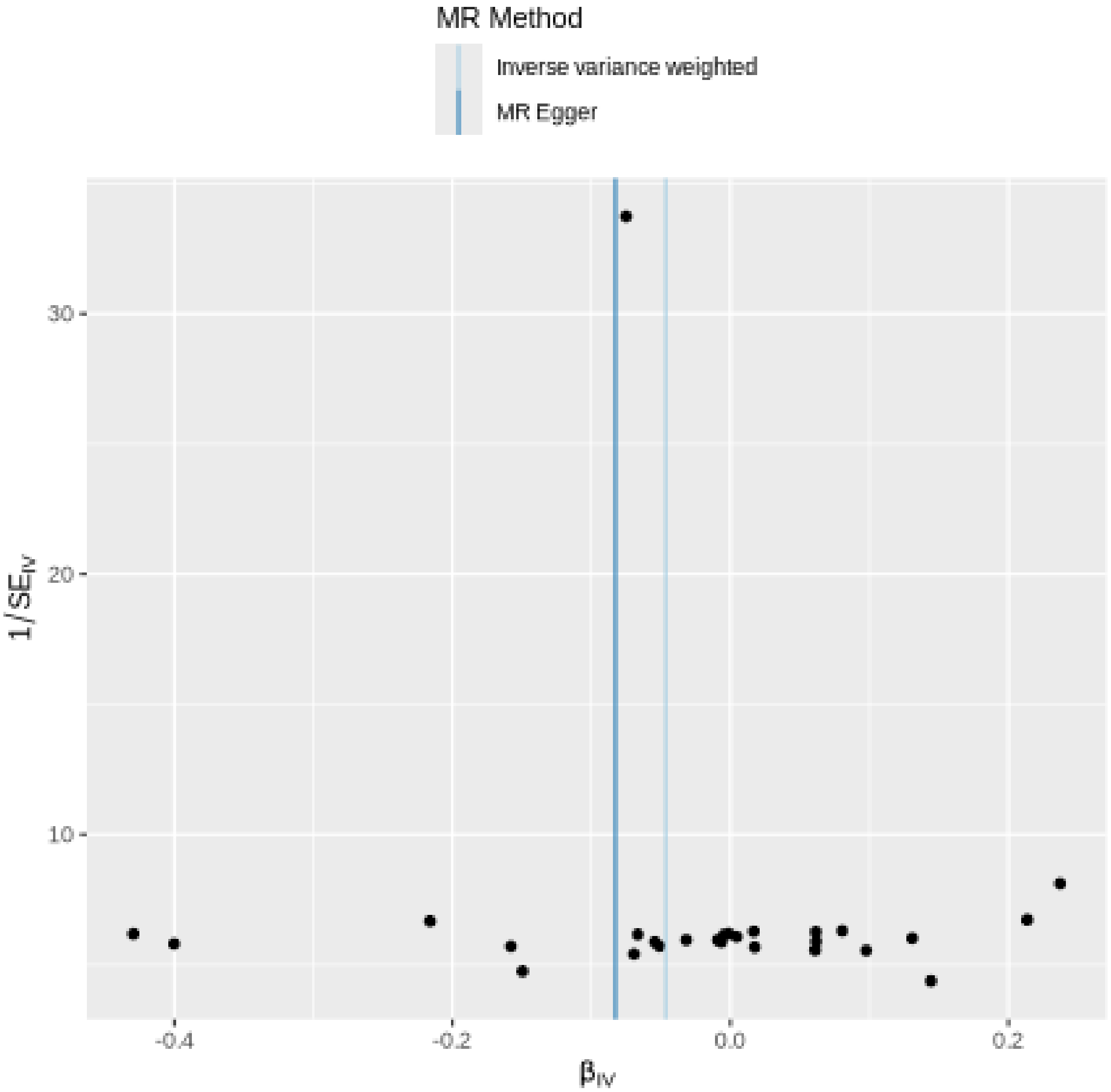
MR Method
Inverse variance weighted
MR Egger
30
20
10
1/SE$_{IV}$
-0.4
-0.2
0.0
0.2
β$_{IV}$

#### Leave-one-out sensitivity analysis

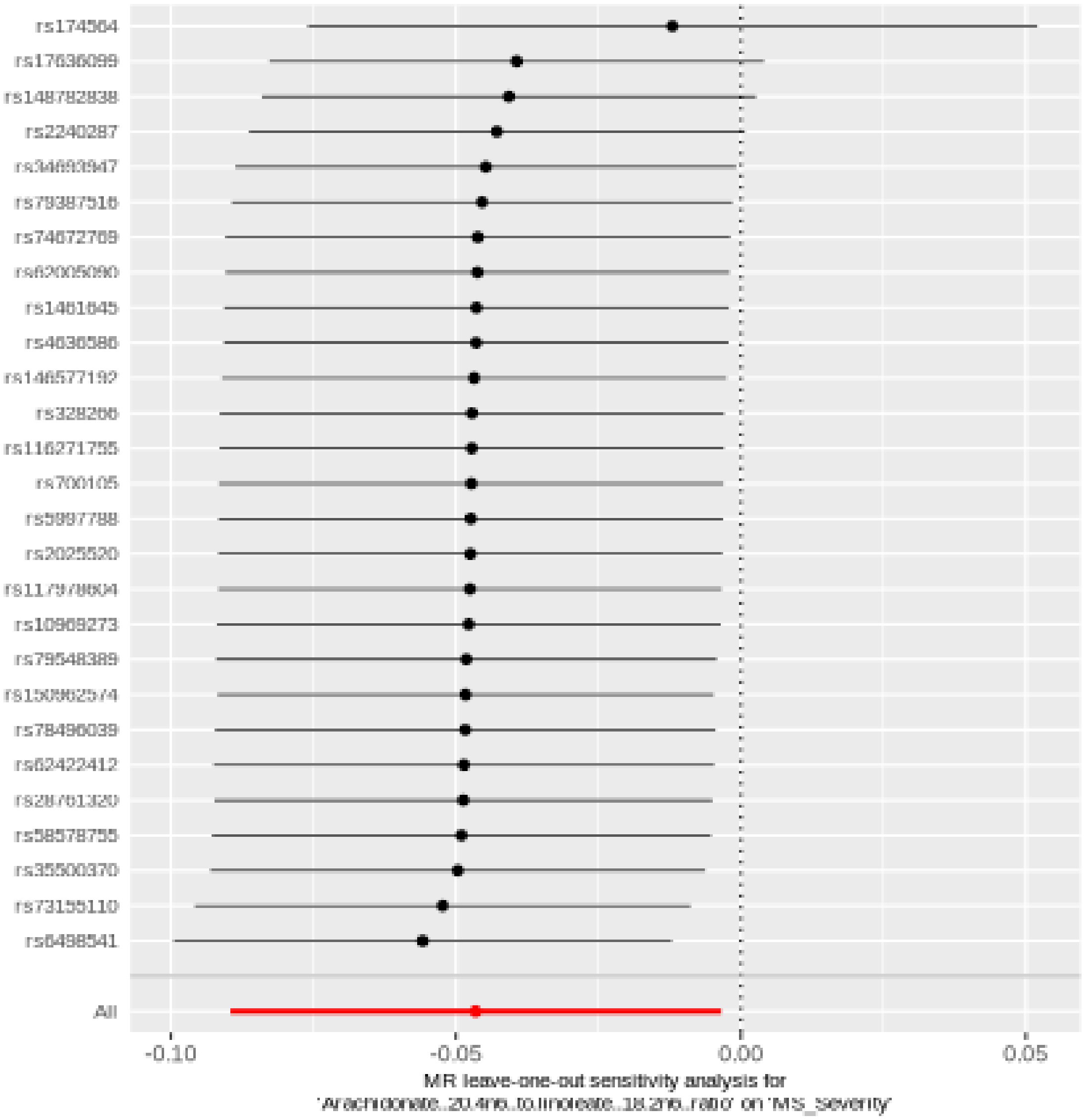
rs174564
rs17636099
rs148782838
rs2240287
rs79387516
rs74672769
rs1461645
rs4636586
rs146577192
rs328266
rs116271755
rs700105
rs5997788
rs2025520
rs10969273
rs79548389
rs78496039
rs62422412
rs58578755
rs35500370
rs73155110
rs6498541
All
-0.10
-0.05
0.00
0.05
MR leave-one-out sensitivity analysis for
'Arachidonate..20.4n6..to.linoleate..18.2n6..ratio' on 'MS_Severity'

## X.16124

| method | nsnp | b | se | pval |
|---|---|---|---|---|
| MR Egger | 28 | 0.0617585 | 0.0455311 | 0.1866343 |
| Weighted median | 28 | 0.0304252 | 0.0356496 | 0.3934082 |
| Inverse variance weighted | 28 | 0.0500591 | 0.0251331 | 0.0463974 |
| Simple mode | 28 | 0.0935904 | 0.0656362 | 0.1653588 |
| Weighted mode | 28 | 0.0386776 | 0.0414135 | 0.3586141 |

#### Heterogeneity tests

| method | Q | Q_df | Q_pval |
|---|---|---|---|
| MR Egger | 35.02360 | 26 | 0.1111395 |
| Inverse variance weighted | 35.15349 | 27 | 0.1350204 |

#### Test for directional horizontal pleiotropy

| egger_intercept | se | pval |
|---|---|---|
| -0.0024088 | 0.0077574 | 0.7586419 |

#### Forest plot of single SNP MR

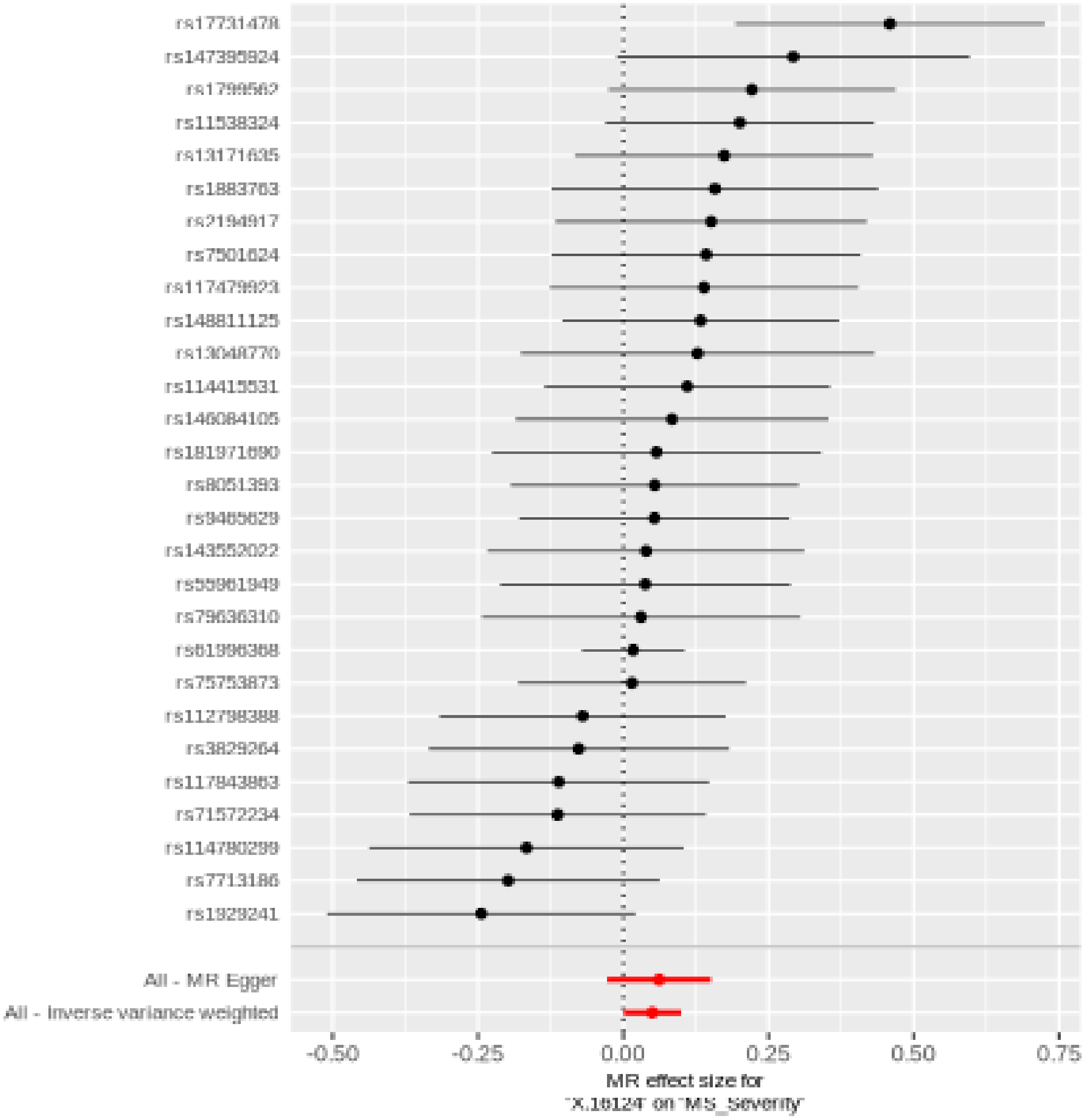

#### Comparison of results using different MR methods

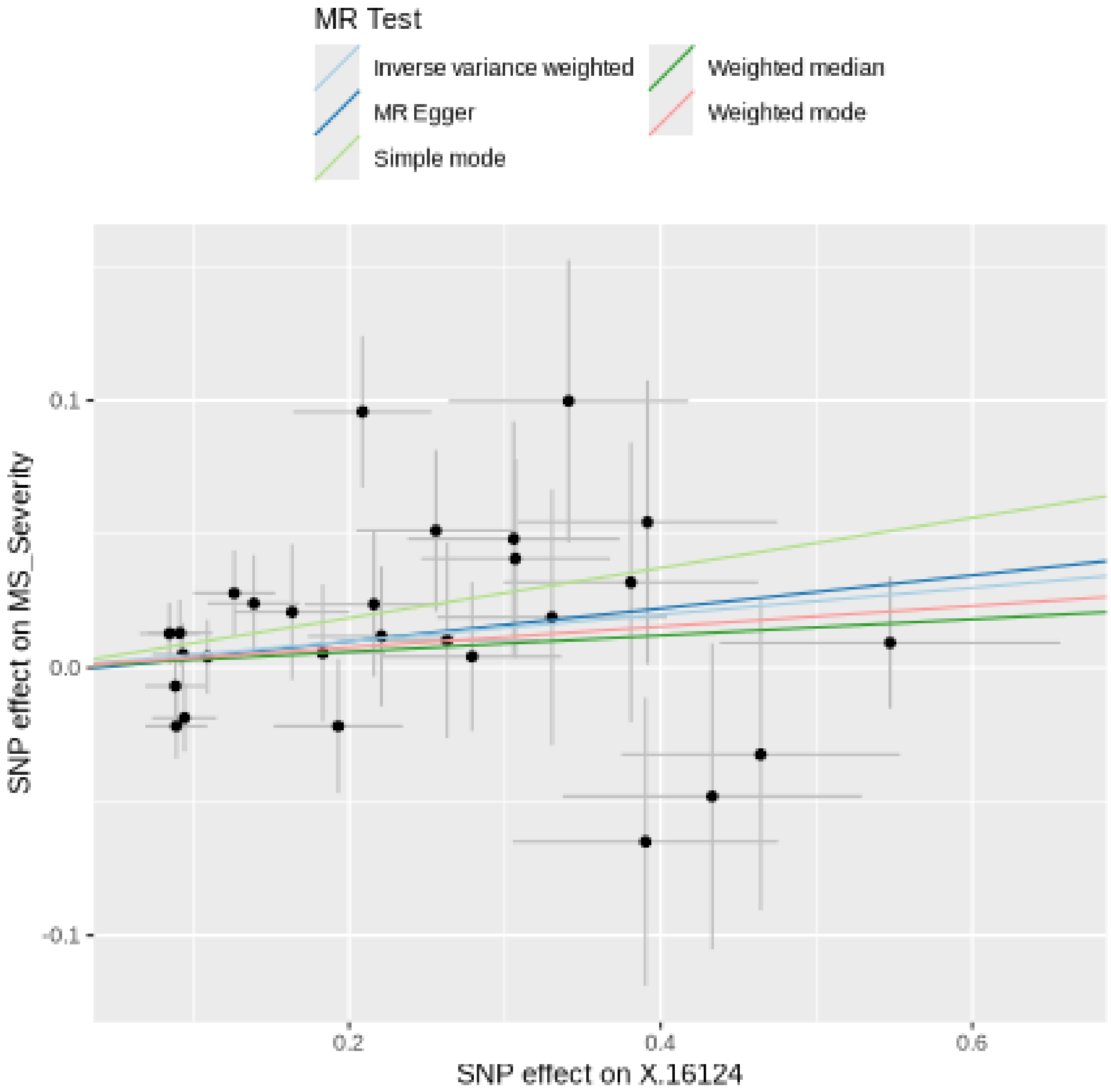

#### Funnel plot

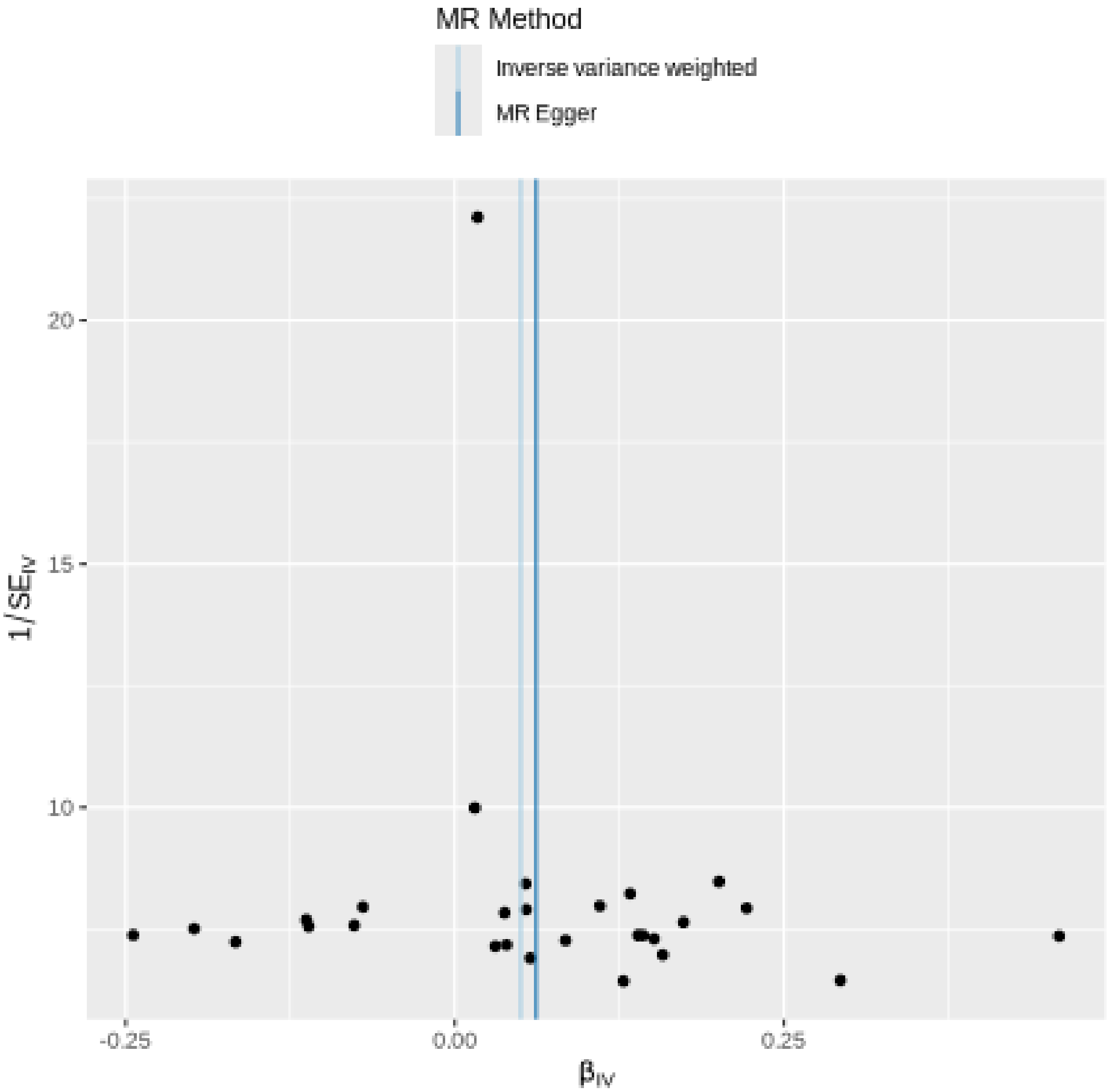

MR Method
Inverse variance weighted
MR Egger
20
15
10
1/SE_IV
-0.25
0.00
0.25
β_IV

### Leave-one-out sensitivity analysis

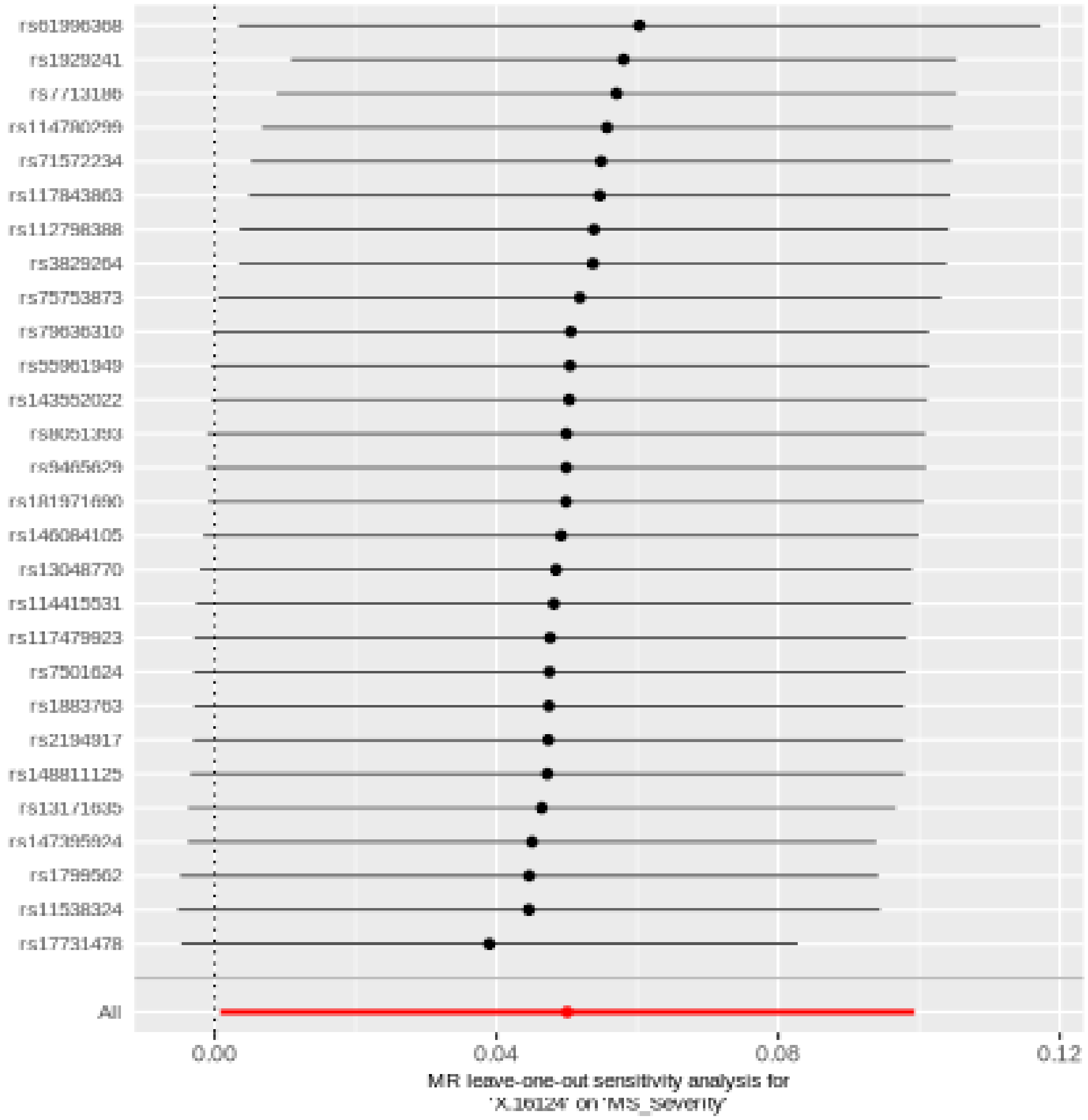

## X.17654

| method | nsnp | b | se | pval |
|---|---|---|---|---|
| MR Egger | 33 | 0.0467404 | 0.0335733 | 0.1737734 |
| Weighted median | 33 | 0.0855975 | 0.0301565 | 0.0045334 |
| Inverse variance weighted | 33 | 0.0408662 | 0.0197333 | 0.0383662 |
| Simple mode | 33 | 0.1031737 | 0.0588032 | 0.0889070 |
| Weighted mode | 33 | 0.0912460 | 0.0363518 | 0.0173219 |

#### Heterogeneity tests

| method | Q | Q_df | Q_pval |
|---|---|---|---|
| MR Egger | 29.52581 | 31 | 0.5418689 |
| Inverse variance weighted | 29.57258 | 32 | 0.5899700 |

#### Test for directional horizontal pleiotropy

| egger_intercept | se | pval |
|---|---|---|
| -0.0013076 | 0.0060461 | 0.8301974 |

### Forest plot of single SNP MR

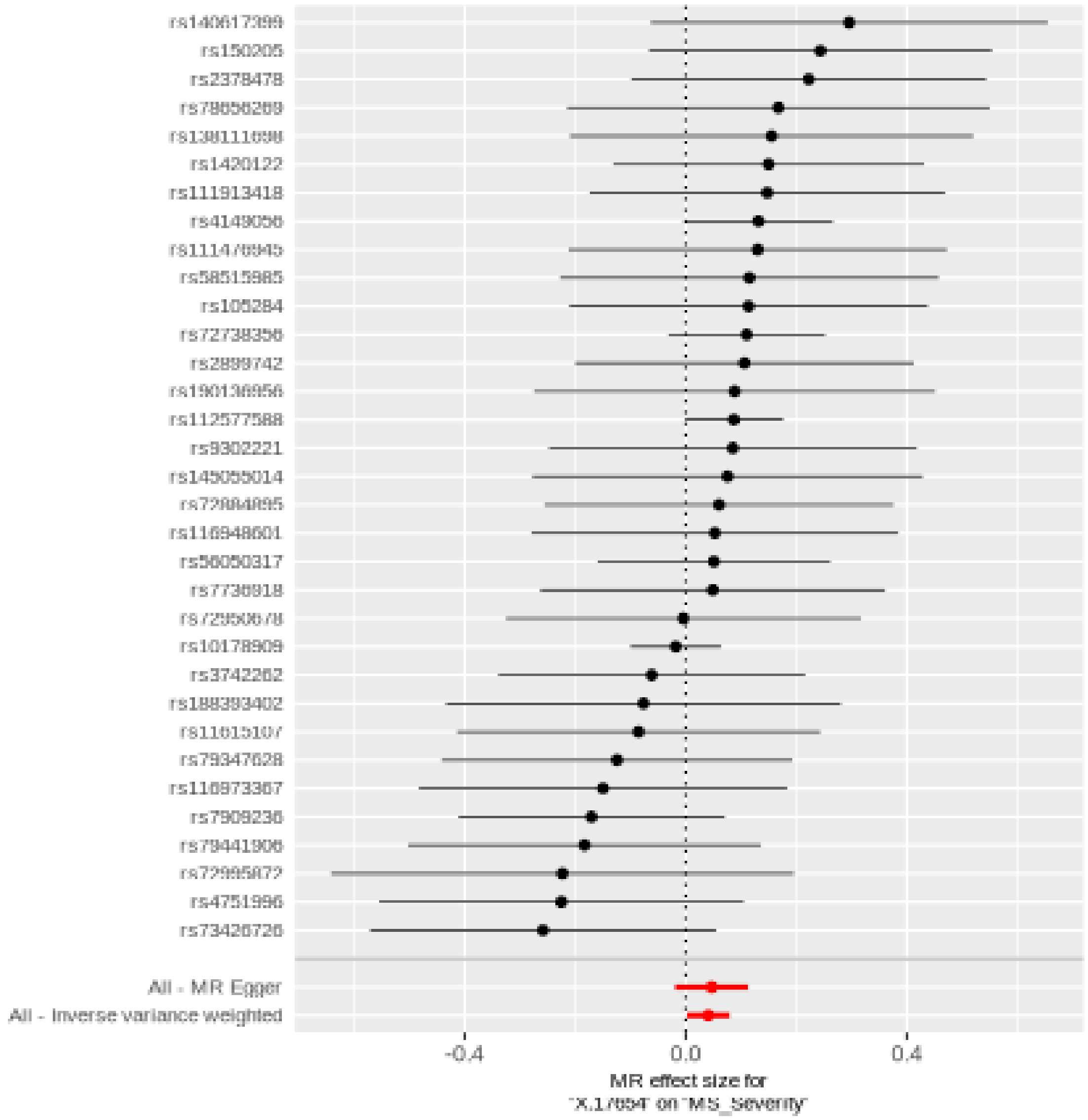

#### Comparison of results using different MR methods

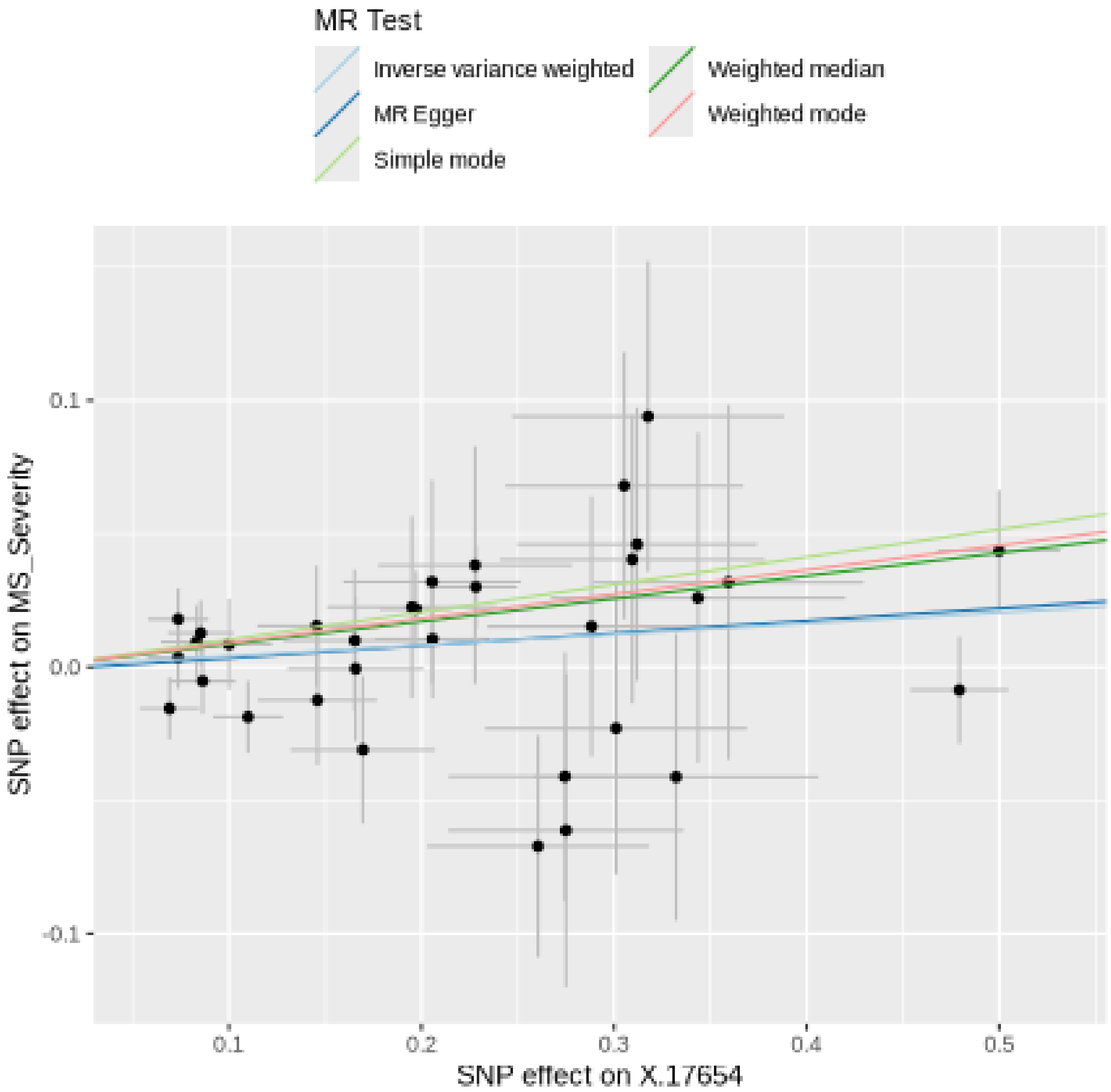

**Funnel plot**

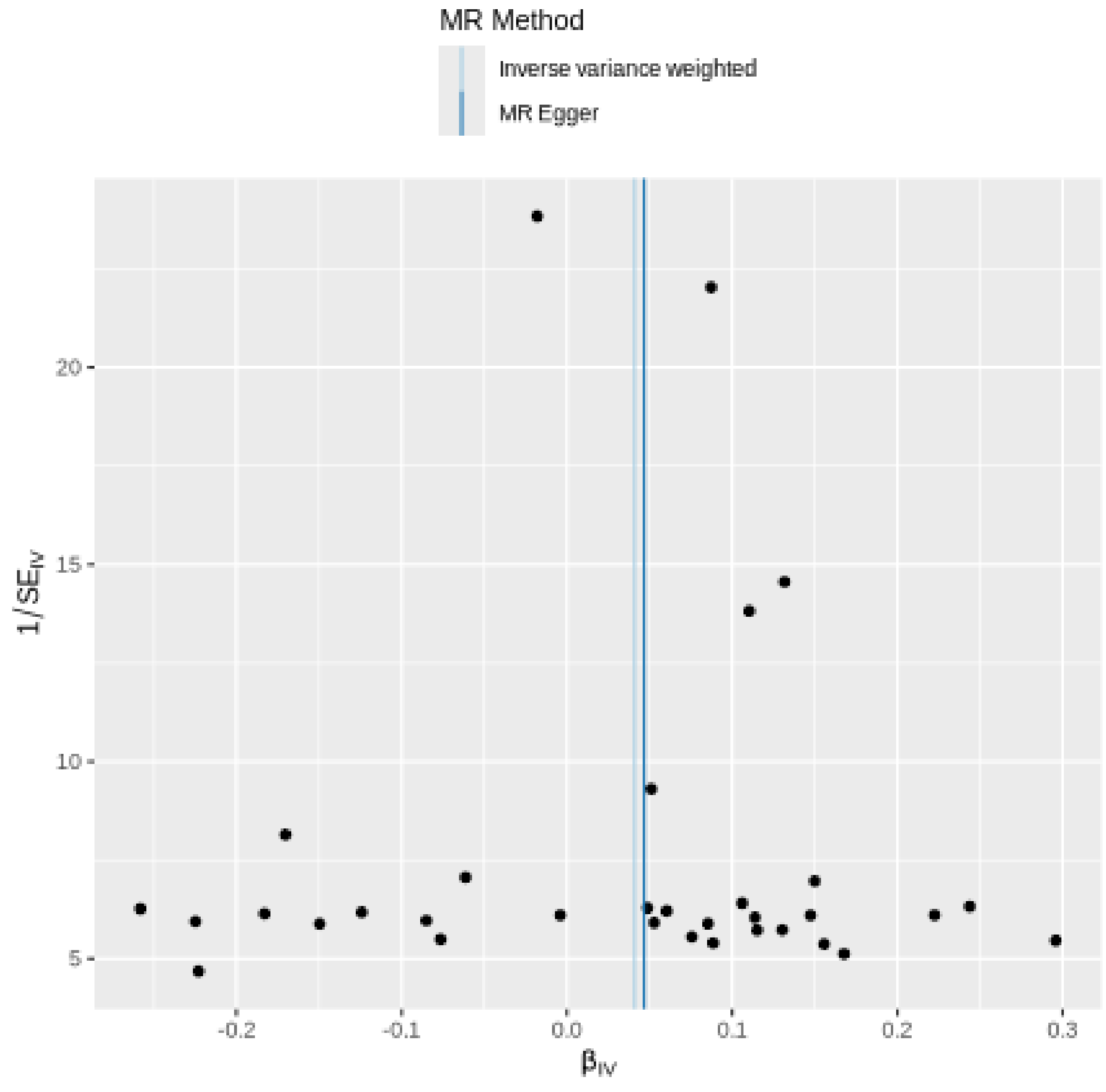

MR Method
Inverse variance weighted
MR Egger
20
15
10
5
$1/SE_{IV}$
-0.2
-0.1
0.0
0.1
0.2
0.3
$\beta_{IV}$

### Leave-one-out sensitivity analysis

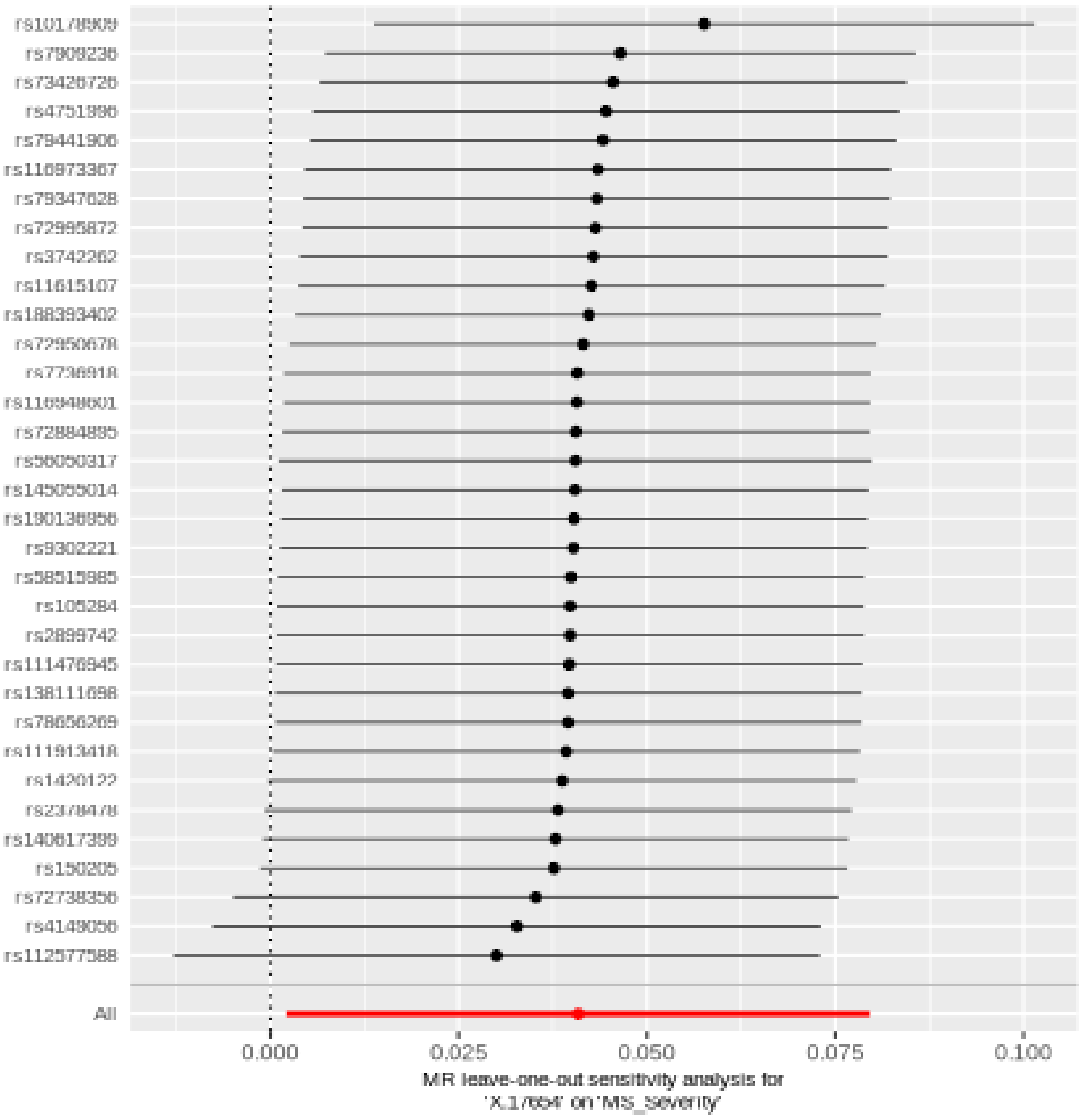

## X.17653

| method | nsnp | b | se | pval |
|---|---|---|---|---|
| MR Egger | 27 | 0.0965810 | 0.0561240 | 0.0976337 |
| Weighted median | 27 | 0.0927339 | 0.0403248 | 0.0214668 |
| Inverse variance weighted | 27 | 0.0678928 | 0.0273509 | 0.0130542 |
| Simple mode | 27 | 0.0763970 | 0.0601903 | 0.2155937 |
| Weighted mode | 27 | 0.0964480 | 0.0425593 | 0.0319944 |

#### Heterogeneity tests

| method | Q | Q_df | Q_pval |
|---|---|---|---|
| MR Egger | 29.93645 | 25 | 0.2266775 |
| Inverse variance weighted | 30.35009 | 26 | 0.2533616 |

#### Test for directional horizontal pleiotropy

| egger_intercept | se | pval |
|---|---|---|
| -0.0050171 | 0.0085363 | 0.5619788 |

### Forest plot of single SNP MR

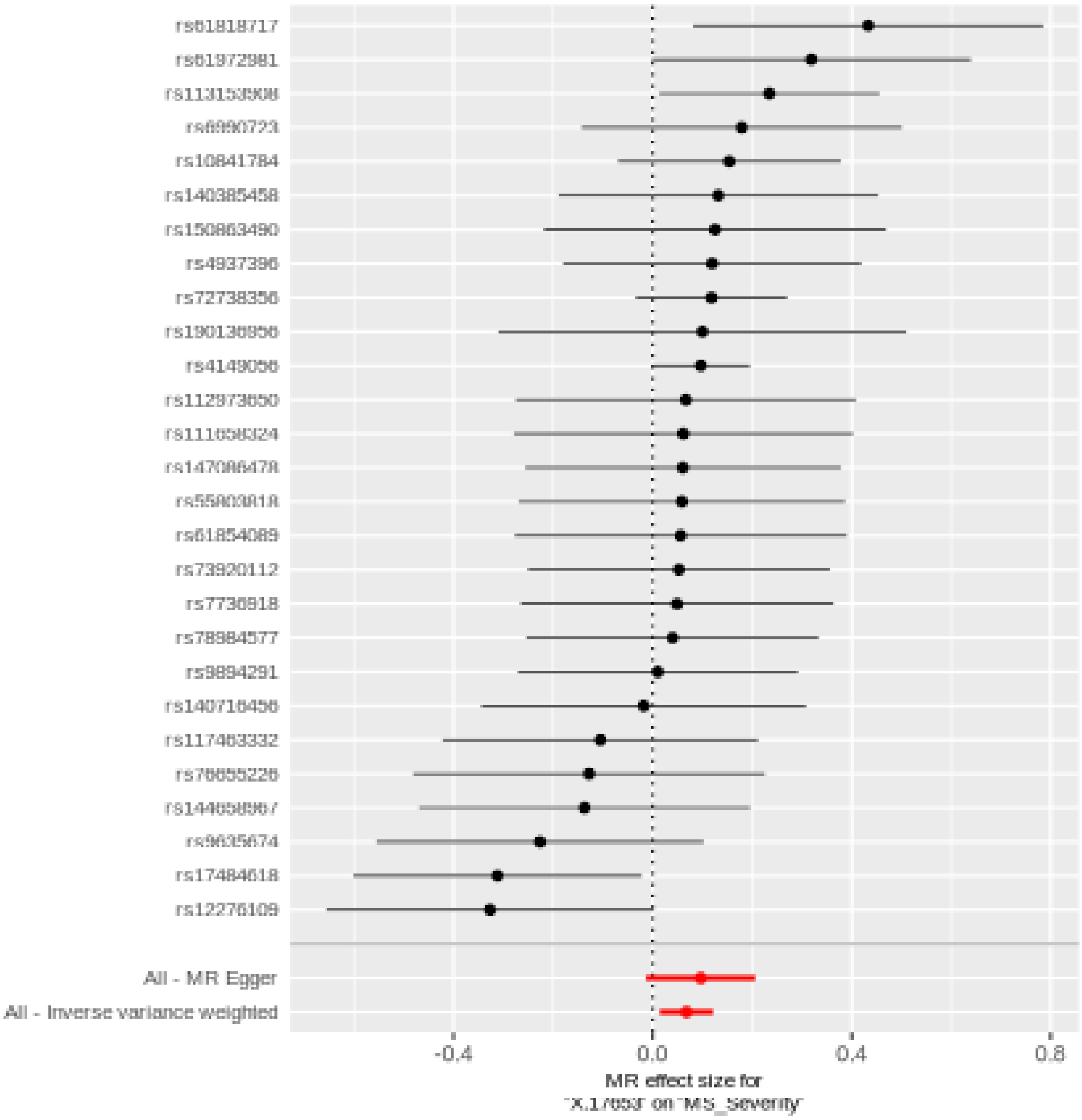

#### Comparison of results using different MR methods

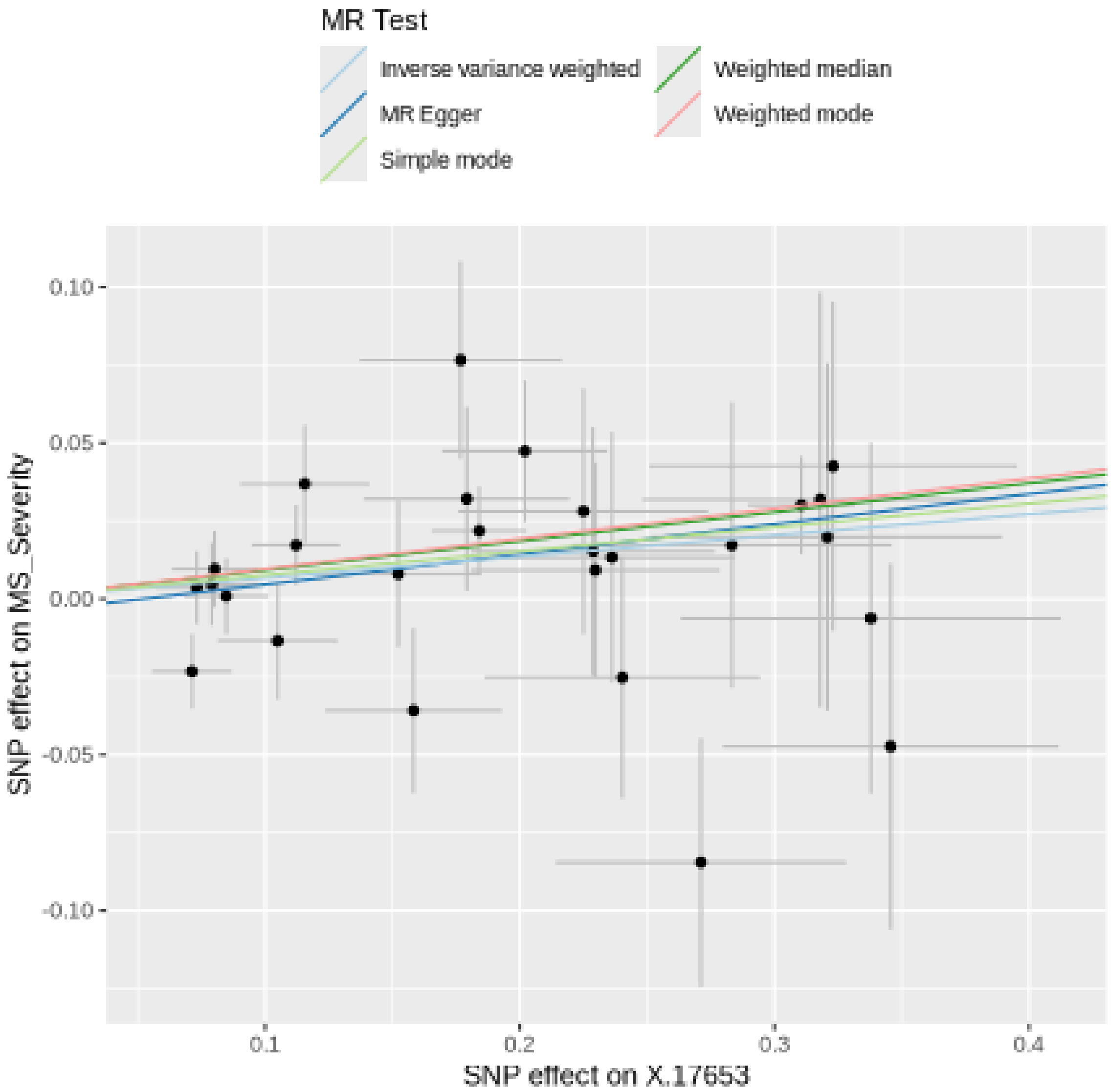

**Funnel plot**

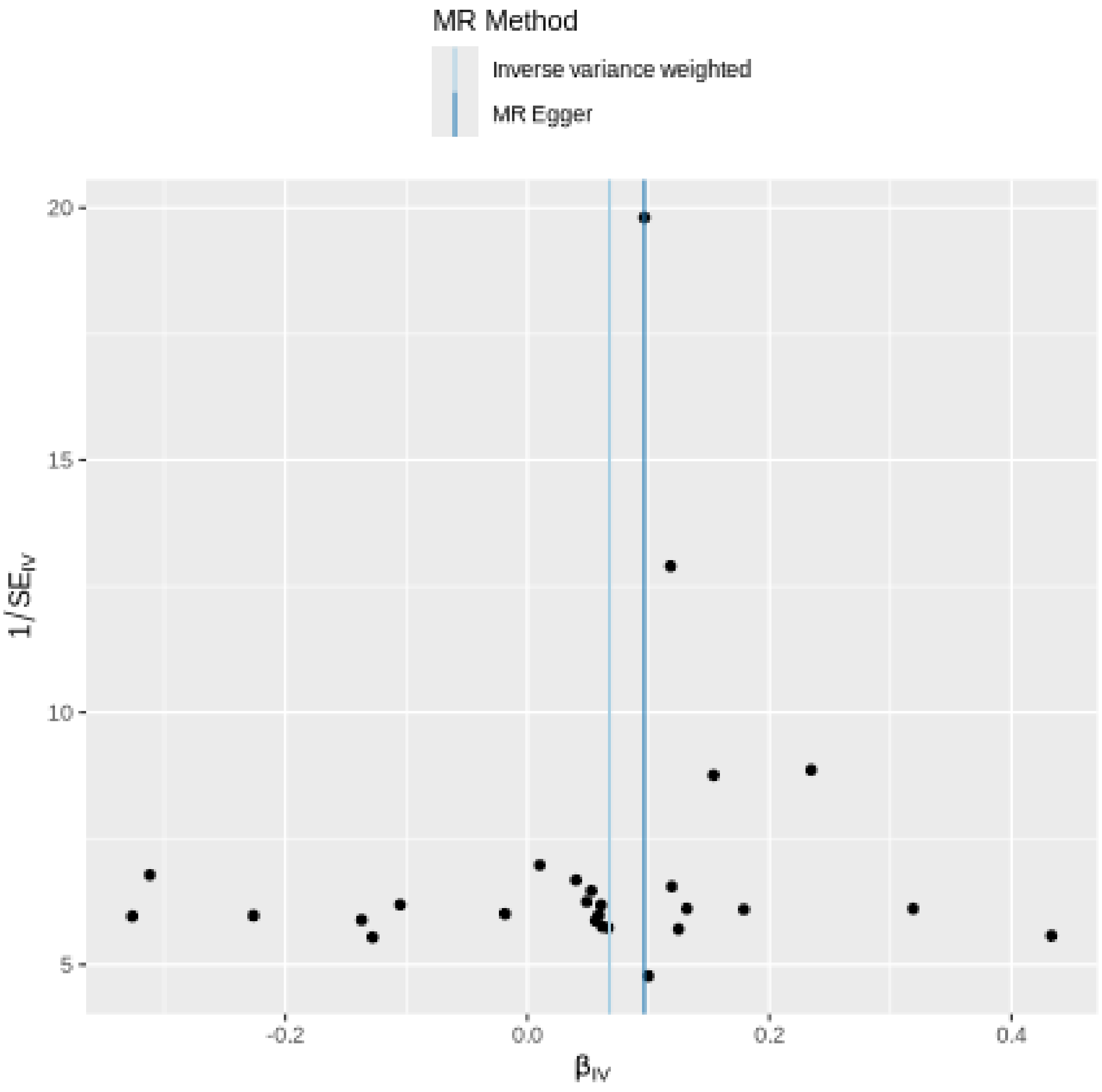

MR Method
Inverse variance weighted
MR Egger
20
15
10
5
$1/SE_{IV}$
-0.2
0.0
0.2
0.4
$\beta_{IV}$

#### Leave-one-out sensitivity analysis

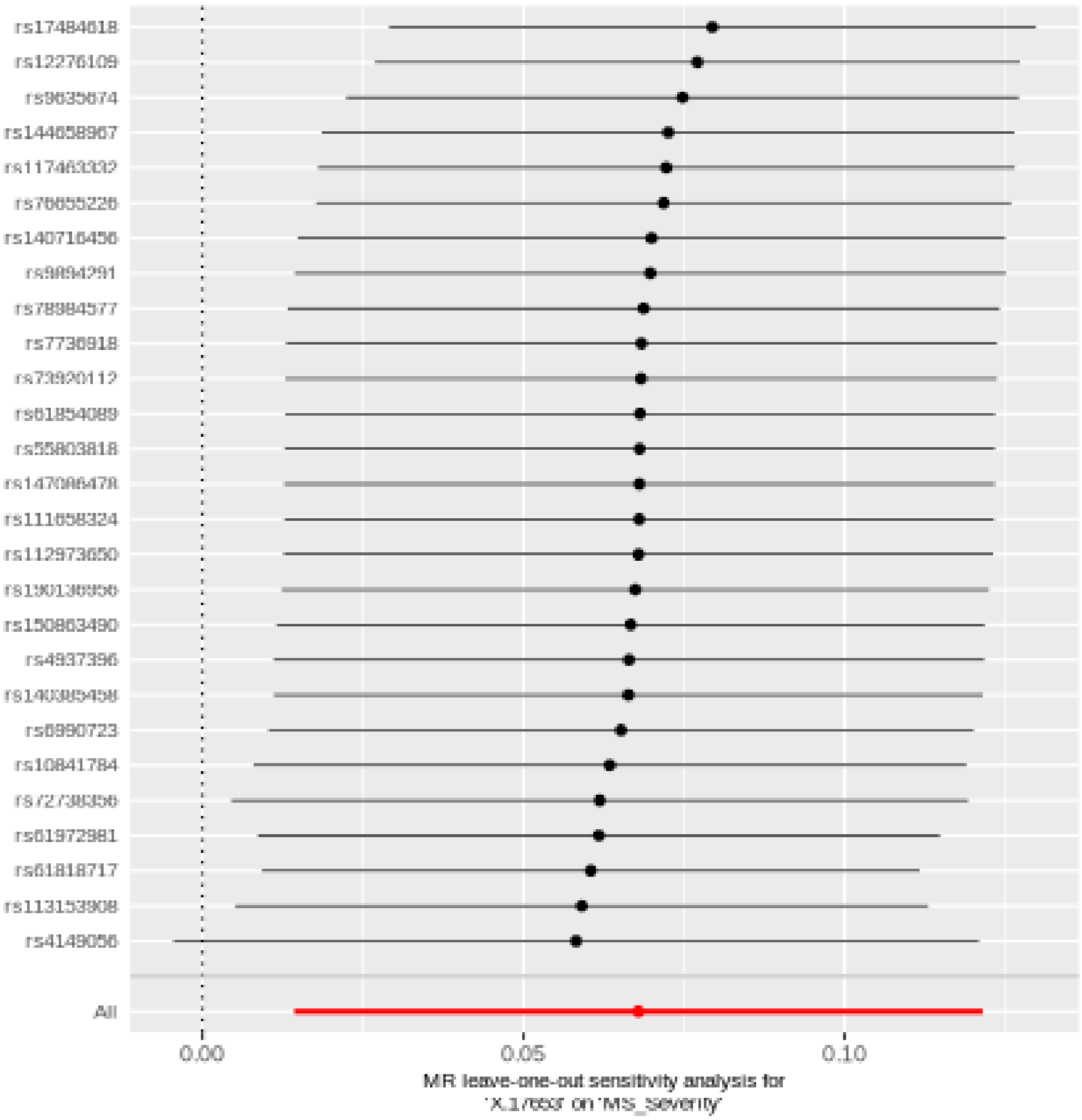

## X.21467

| method | nsnp | b | se | pval |
|---|---|---|---|---|
| MR Egger | 37 | 0.0547878 | 0.0359517 | 0.1365123 |
| Weighted median | 37 | 0.0535449 | 0.0270187 | 0.0475051 |
| Inverse variance weighted | 37 | 0.0509659 | 0.0202469 | 0.0118285 |
| Simple mode | 37 | 0.0285670 | 0.0530834 | 0.5937833 |
| Weighted mode | 37 | 0.0517484 | 0.0261825 | 0.0558043 |

##### Heterogeneity tests

| method | Q | Q_df | Q_pval |
|---|---|---|---|
| MR Egger | 44.40453 | 35 | 0.1324566 |
| Inverse variance weighted | 44.42581 | 36 | 0.1582005 |

##### Test for directional horizontal pleiotropy

| egger_intercept | se | pval |
|---|---|---|
| -0.0006968 | 0.0053811 | 0.8977079 |

#### Forest plot of single SNP MR

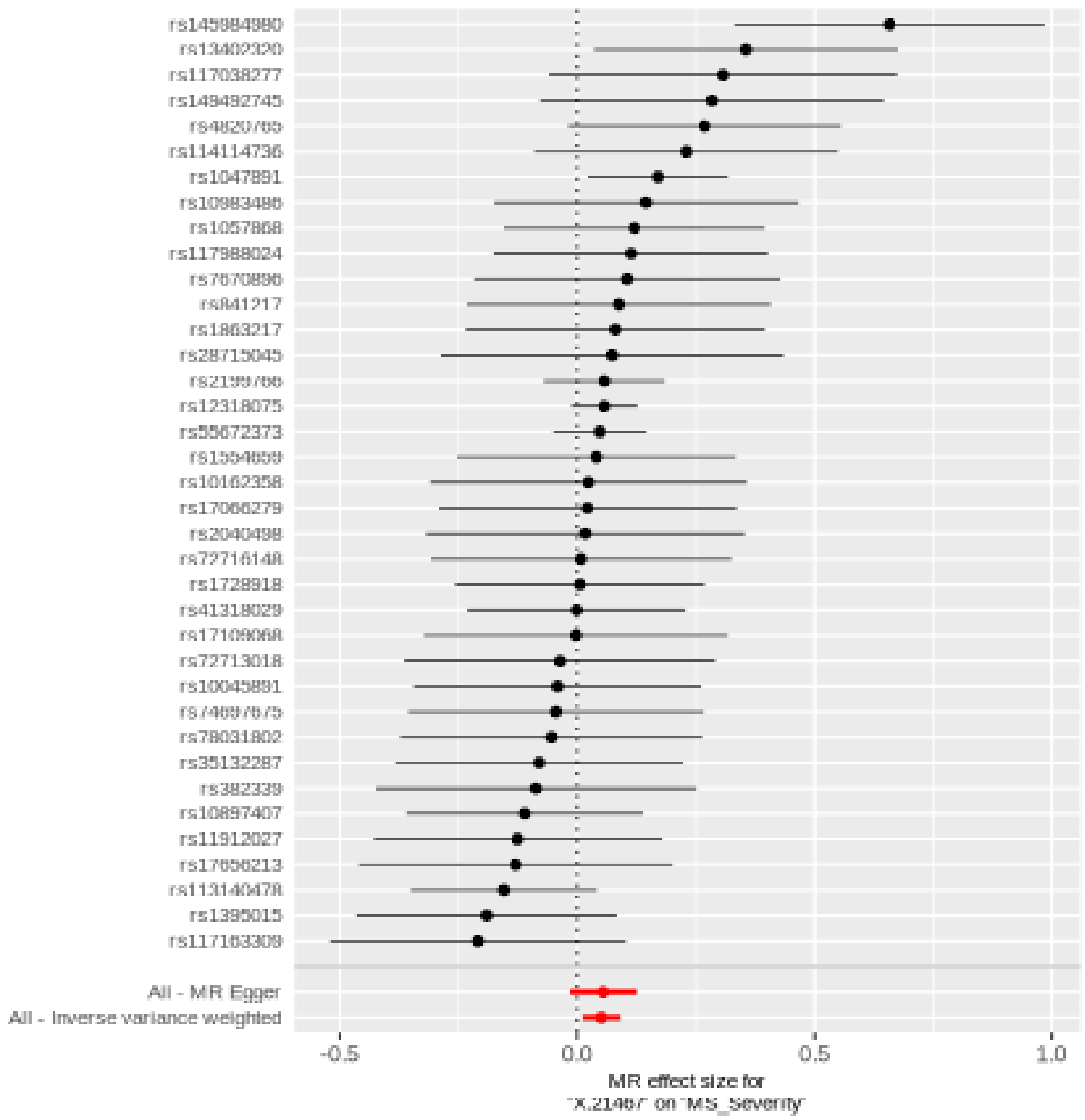

#### Comparison of results using different MR methods

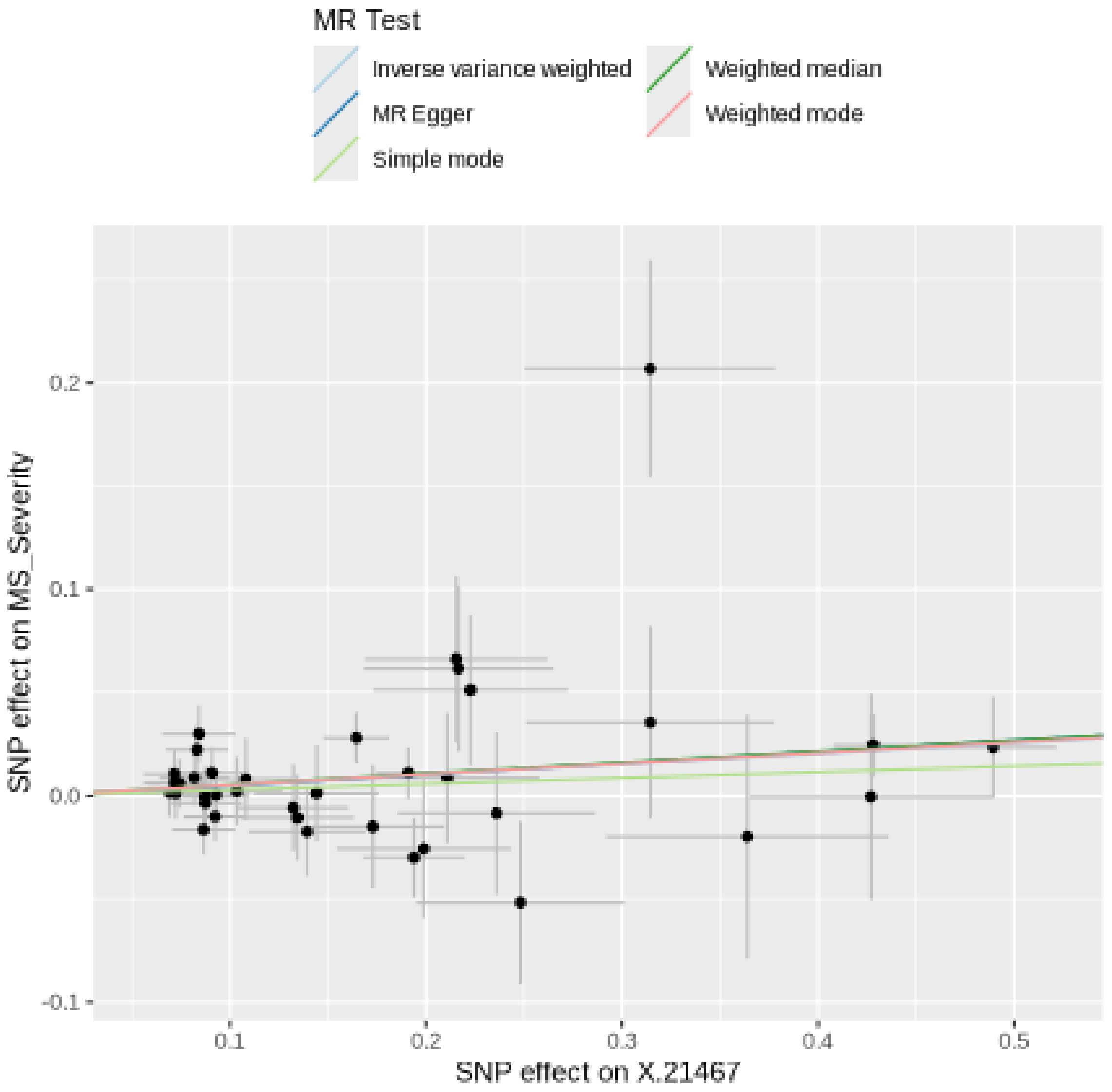
MR Test
Inverse variance weighted
MR Egger
Simple mode
Weighted median
Weighted mode
SNP effect on MS_Severity
0.2
0.1
0.0
-0.1
0.1
0.2
0.3
0.4
0.5
SNP effect on X.21467

#### Funnel plot

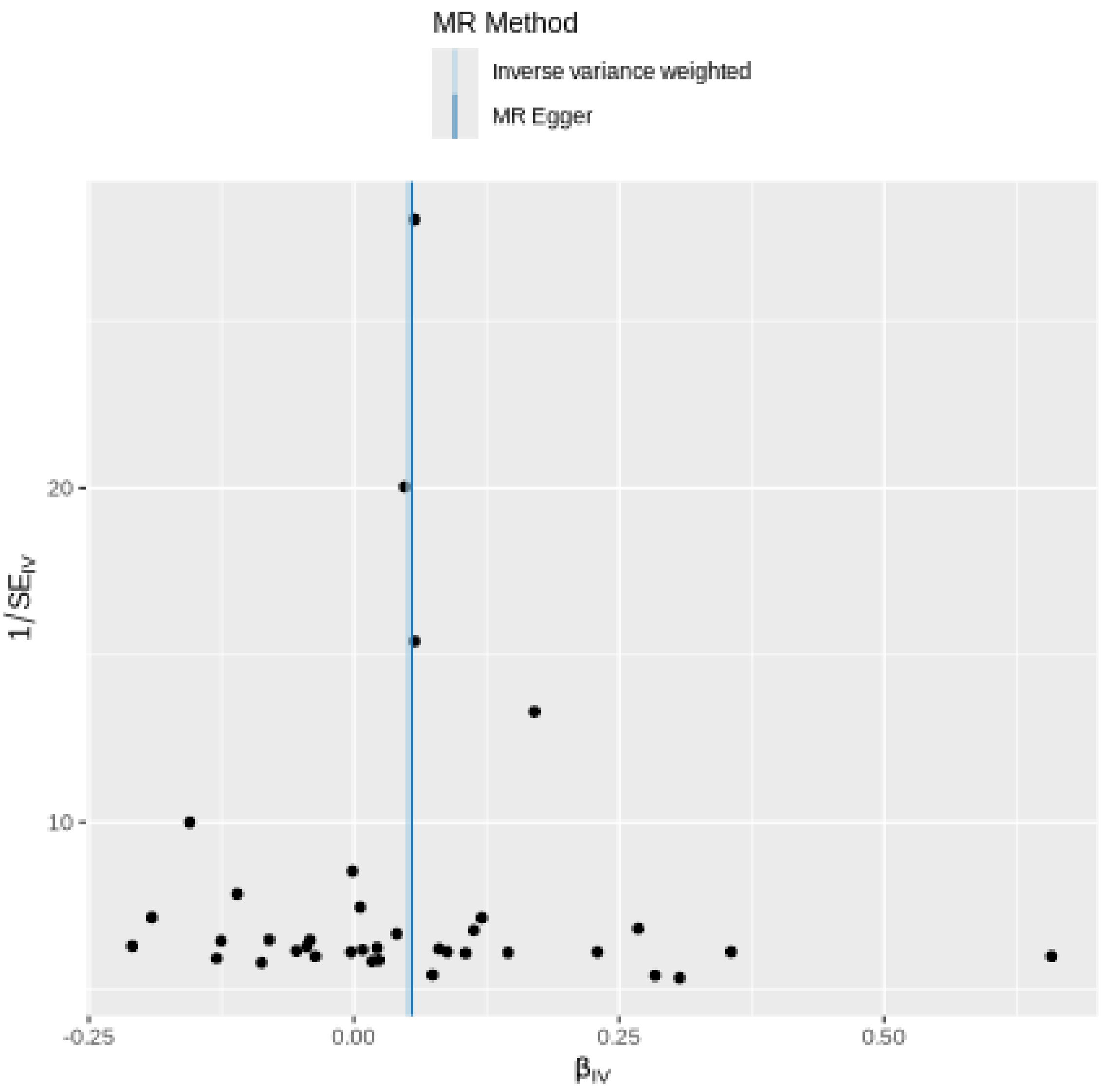
MR Method
Inverse variance weighted
MR Egger
20
10
1/SE$_{IV}$
-0.25
0.00
0.25
0.50
β$_{IV}$

#### **Leave-one-out sensitivity analysis**

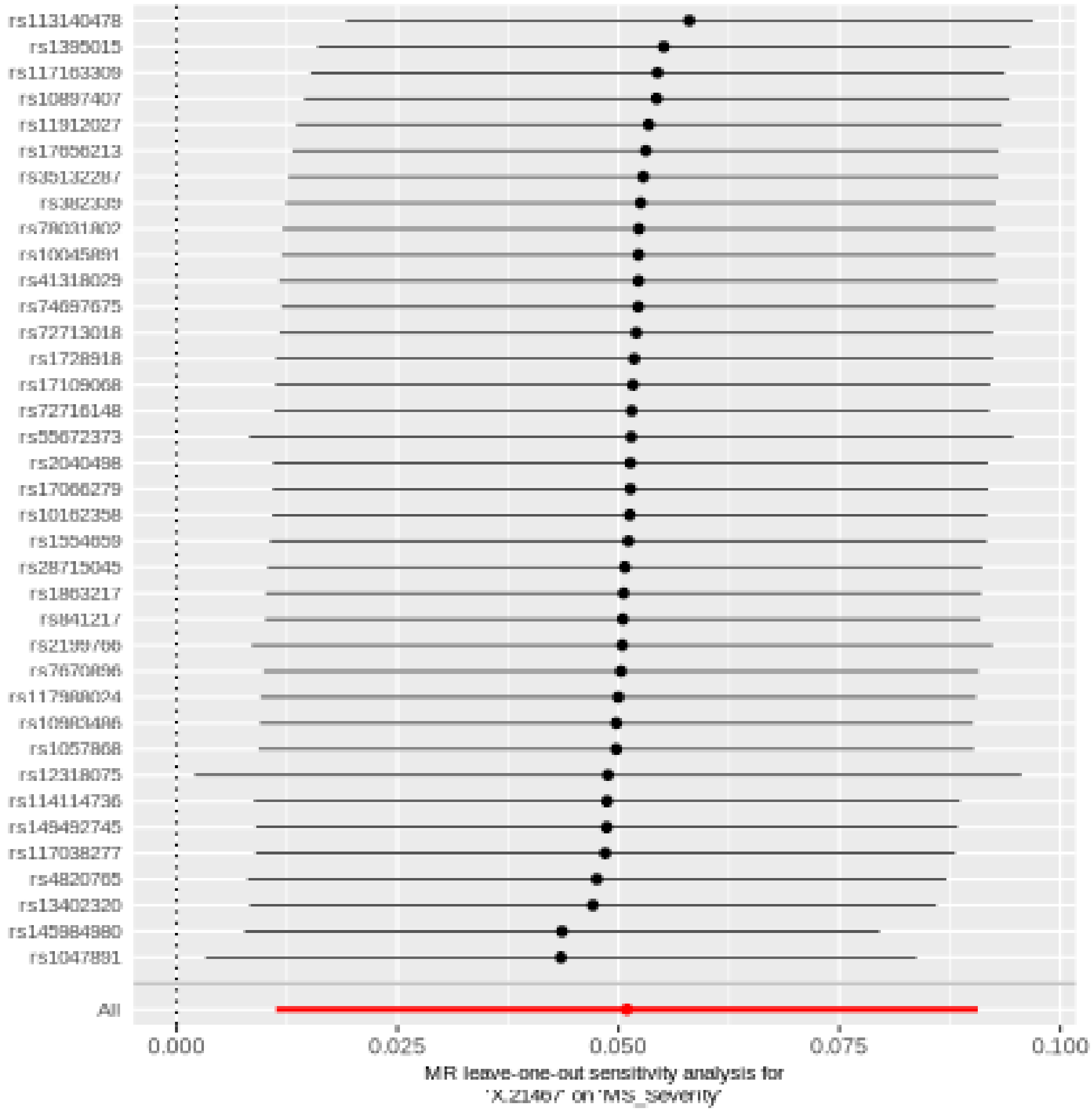

## X.22520

| method | nsnp | b | se | pval |
|---|---|---|---|---|
| MR Egger | 21 | 0.0789389 | 0.0706642 | 0.2778800 |
| Weighted median | 21 | 0.0658977 | 0.0358356 | 0.0659313 |
| Inverse variance weighted | 21 | 0.0660812 | 0.0265049 | 0.0126609 |
| Simple mode | 21 | 0.0524089 | 0.0689444 | 0.4560264 |
| Weighted mode | 21 | 0.0733601 | 0.0668791 | 0.2857149 |

##### Heterogeneity tests

| method | Q | Q_df | Q_pval |
|---|---|---|---|
| MR Egger | 16.16769 | 19 | 0.6460618 |
| Inverse variance weighted | 16.20622 | 20 | 0.7037512 |

##### Test for directional horizontal pleiotropy

| egger_intercept | se | pval |
|---|---|---|
| -0.0020114 | 0.0102474 | 0.846471 |

#### Forest plot of single SNP MR

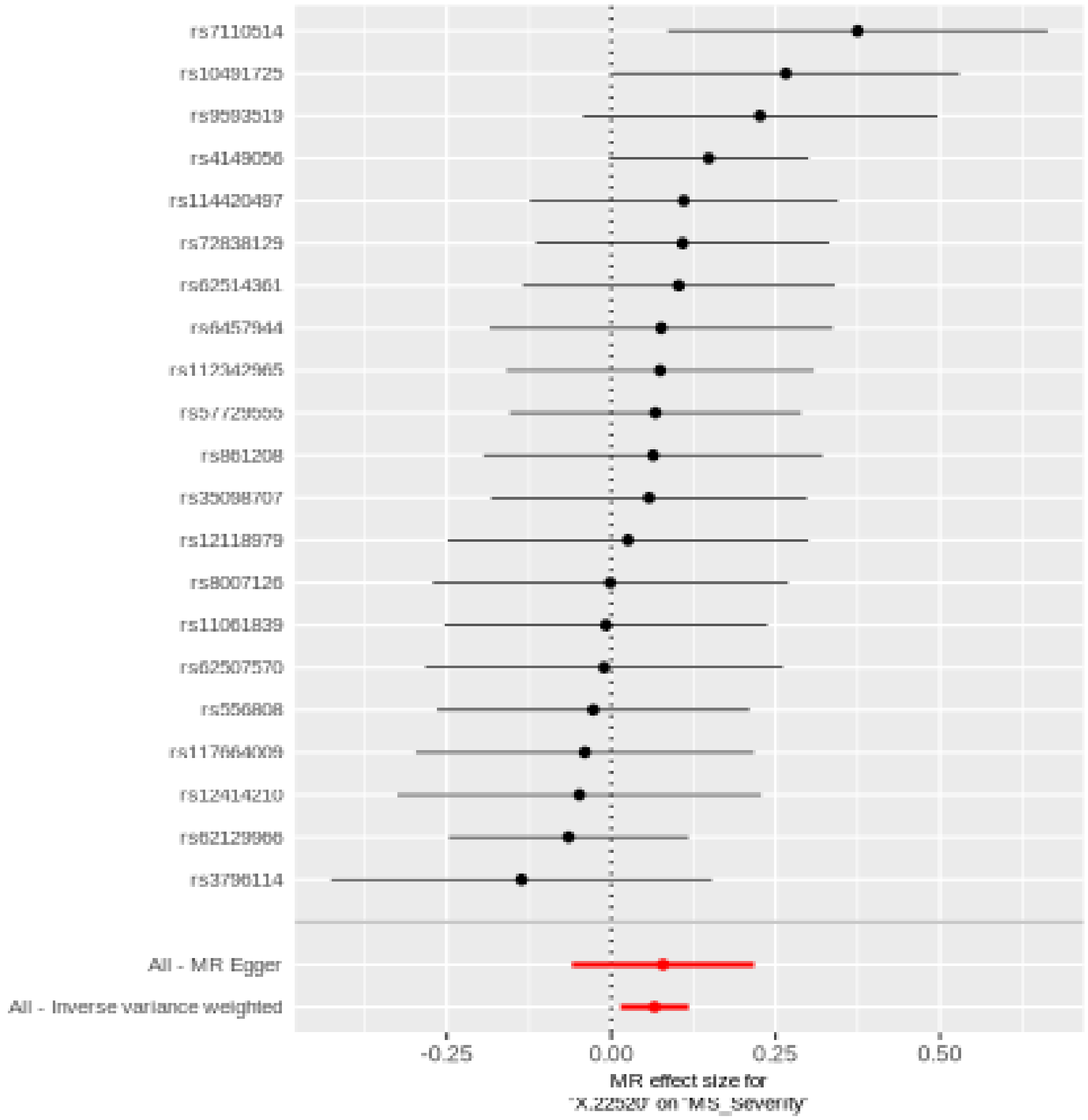

#### Comparison of results using different MR methods

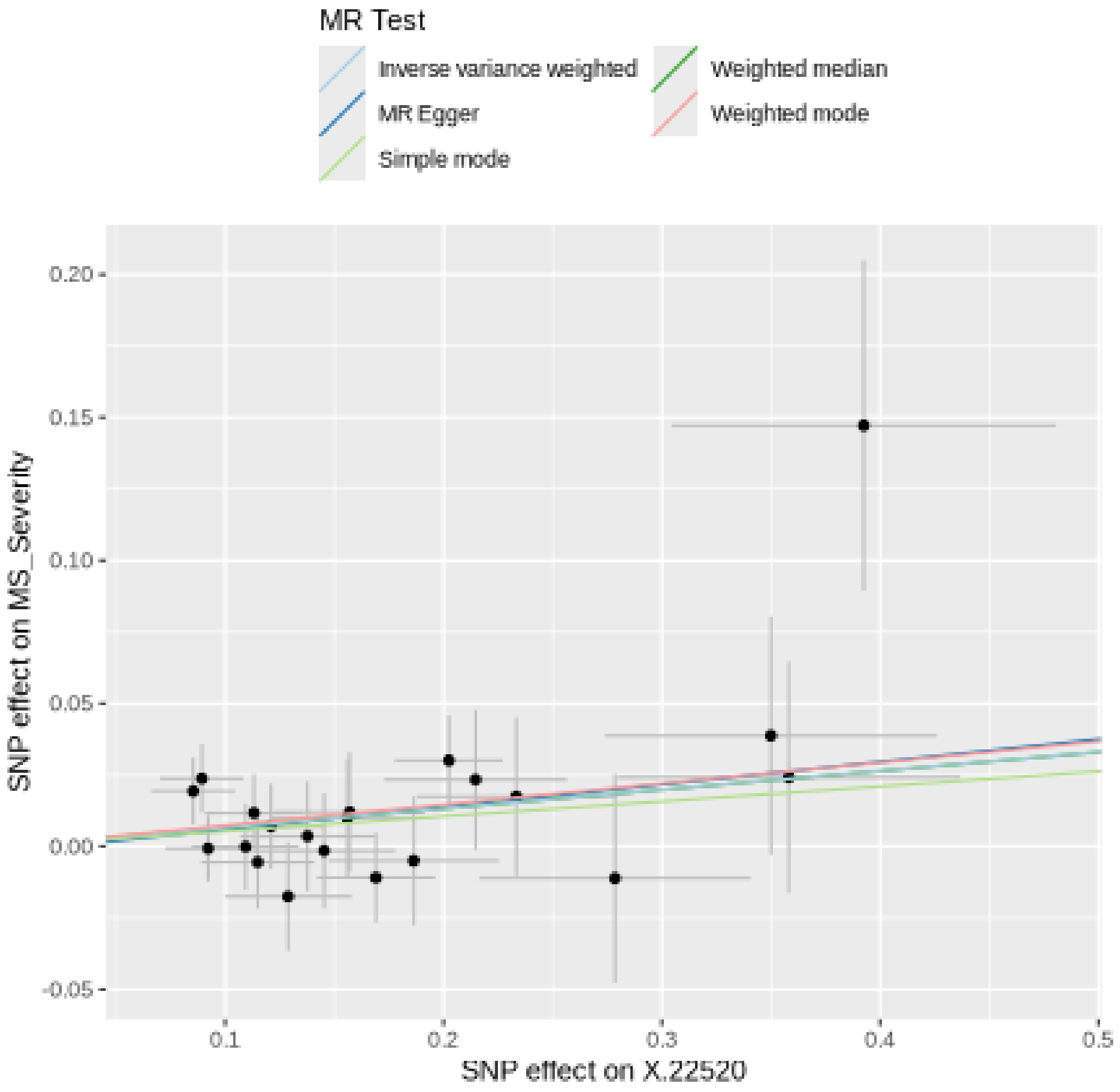

**Funnel plot**

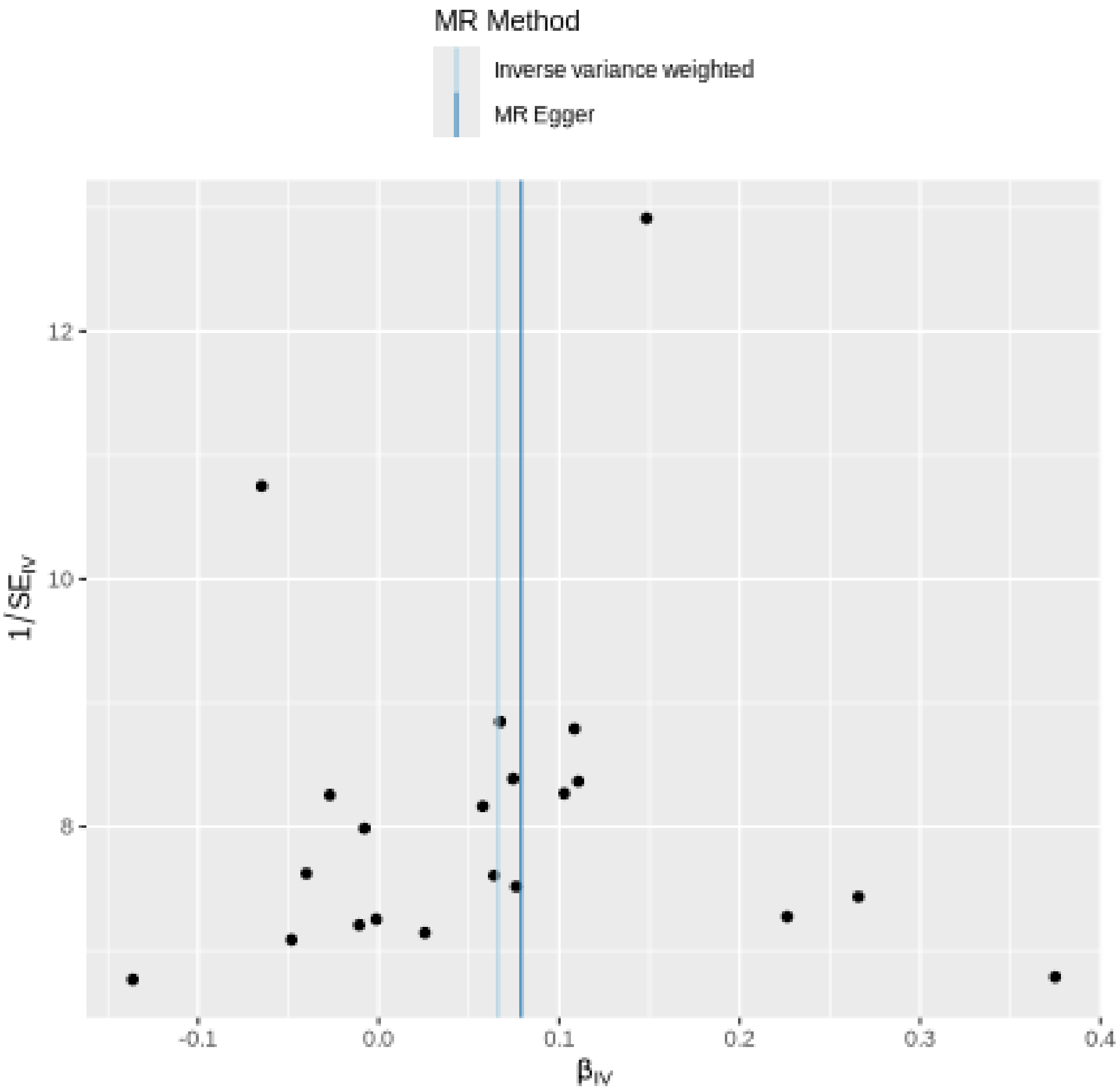

MR Method
Inverse variance weighted
MR Egger
12
10
8
$1/SE_{IV}$
-0.1
0.0
0.1
0.2
0.3
0.4
$\beta_{IV}$

### Leave-one-out sensitivity analysis

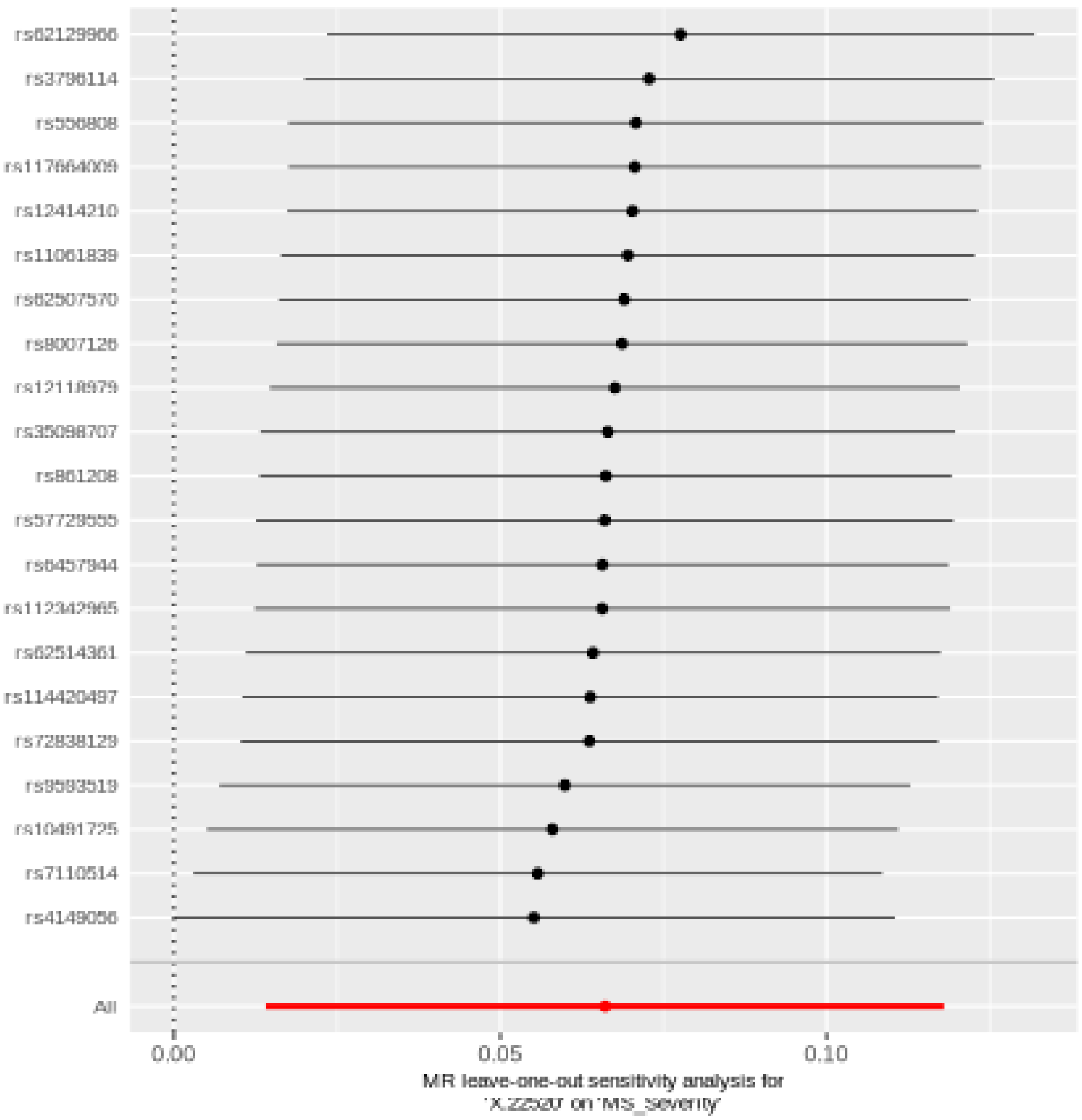

## X.22509

| method | nsnp | b | se | pval |
|---|---|---|---|---|
| MR Egger | 20 | -0.0754027 | 0.0725234 | 0.3122436 |
| Weighted median | 20 | -0.0803569 | 0.0397907 | 0.0434362 |
| Inverse variance weighted | 20 | -0.0582733 | 0.0294524 | 0.0478658 |
| Simple mode | 20 | -0.0817976 | 0.0690621 | 0.2508578 |
| Weighted mode | 20 | -0.0804775 | 0.0667736 | 0.2429116 |

#### Heterogeneity tests

| method | Q | Q_df | Q_pval |
|---|---|---|---|
| MR Egger | 13.15529 | 18 | 0.7822660 |
| Inverse variance weighted | 13.22210 | 19 | 0.8270096 |

#### Test for directional horizontal pleiotropy

| egger_intercept | se | pval |
|---|---|---|
| 0.0026119 | 0.0101056 | 0.7989808 |

#### Forest plot of single SNP MR

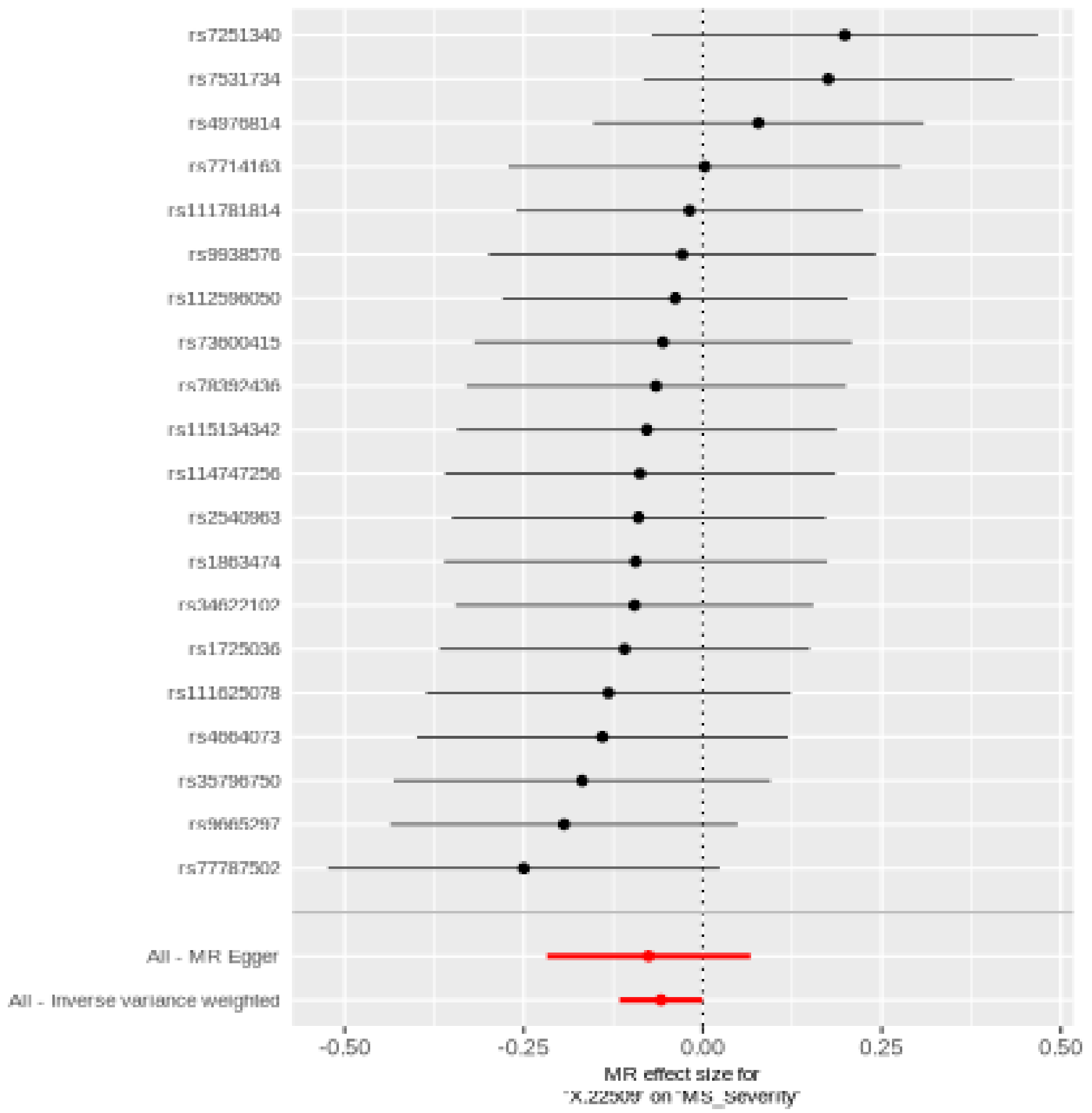

#### Comparison of results using different MR methods

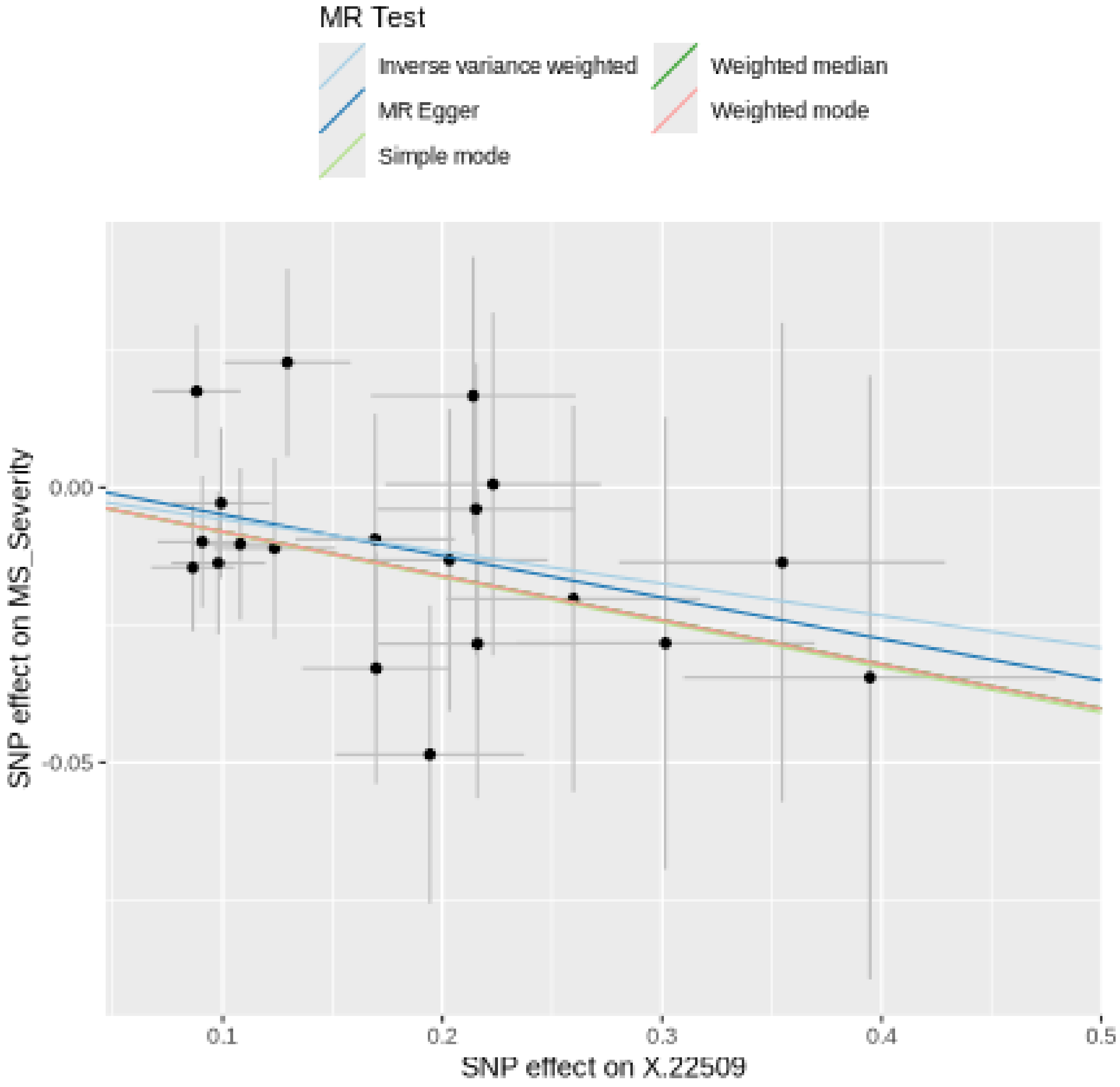
MR Test
Inverse variance weighted
MR Egger
Simple mode
Weighted median
Weighted mode
0.00
-0.05
SNP effect on MS_Severity
0.1
0.2
0.3
0.4
0.5
SNP effect on X.22509

#### Funnel plot

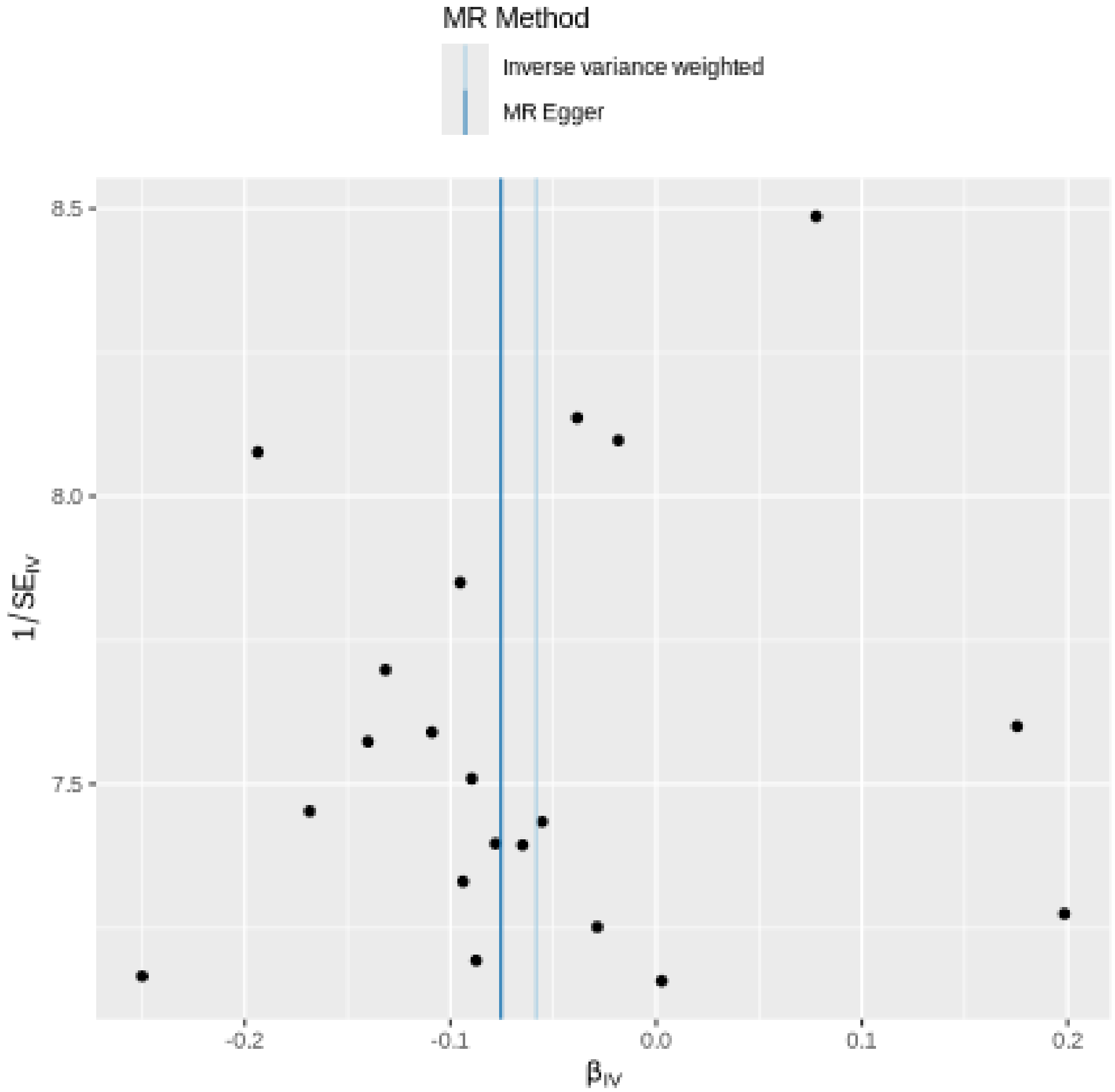

MR Method
Inverse variance weighted
MR Egger
8.5
8.0
7.5
1/SE$_{IV}$
-0.2
-0.1
0.0
0.1
0.2
β$_{IV}$

### Leave-one-out sensitivity analysis

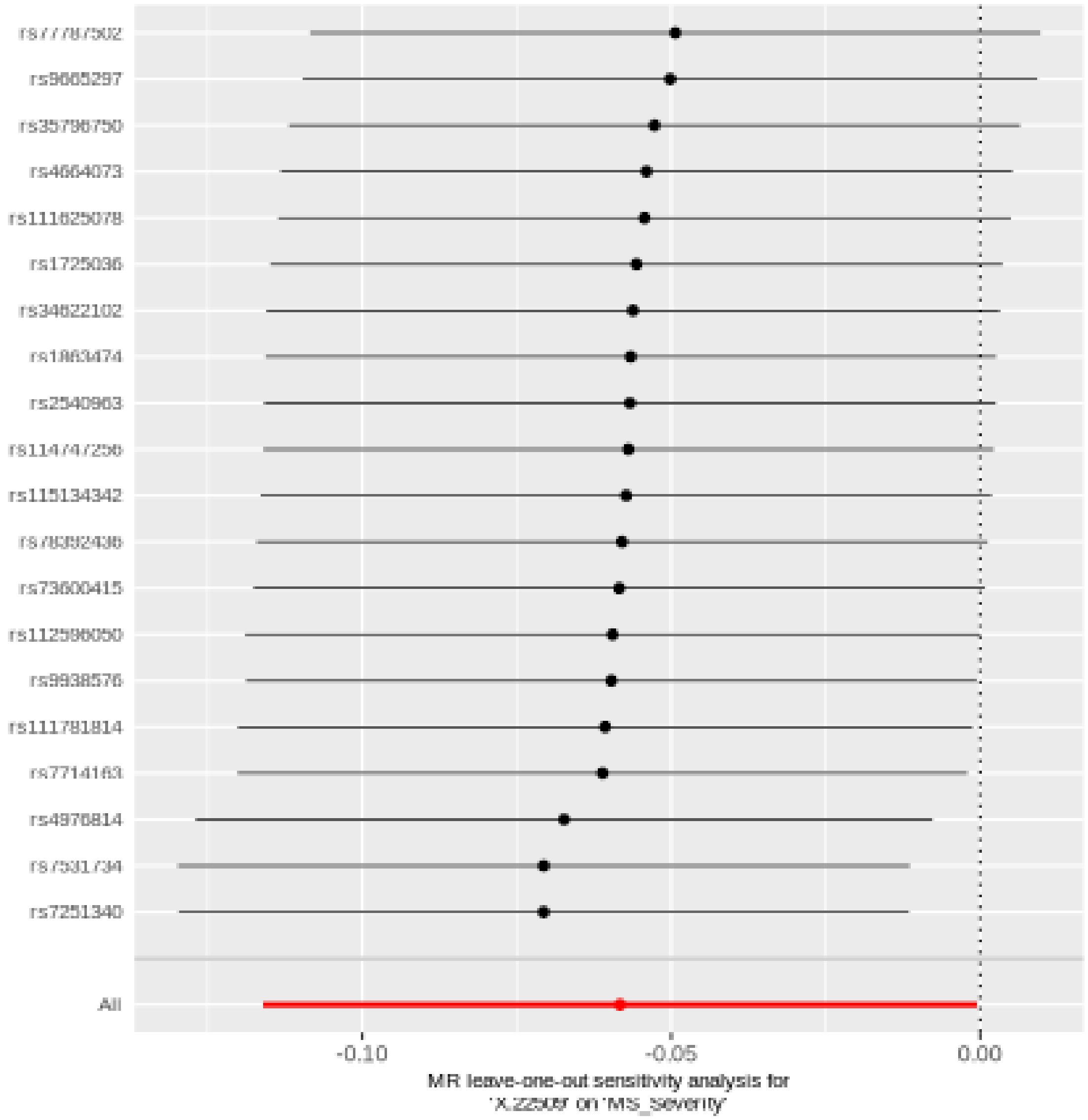

## X.24306

| method | nsnp | b | se | pval |
|---|---|---|---|---|
| MR Egger | 22 | -0.0140952 | 0.0887870 | 0.8754557 |
| Weighted median | 22 | 0.0970335 | 0.0463623 | 0.0363546 |
| Inverse variance weighted | 22 | 0.0815428 | 0.0341226 | 0.0168623 |
| Simple mode | 22 | 0.1066335 | 0.0871069 | 0.2344457 |
| Weighted mode | 22 | 0.1000715 | 0.0796786 | 0.2229317 |

##### Heterogeneity tests

| method | Q | Q_df | Q_pval |
|---|---|---|---|
| MR Egger | 10.51661 | 20 | 0.9578088 |
| Inverse variance weighted | 11.87796 | 21 | 0.9428539 |

##### Test for directional horizontal pleiotropy

| egger_intercept | se | pval |
|---|---|---|
| 0.0111806 | 0.0095826 | 0.2570273 |

#### Forest plot of single SNP MR

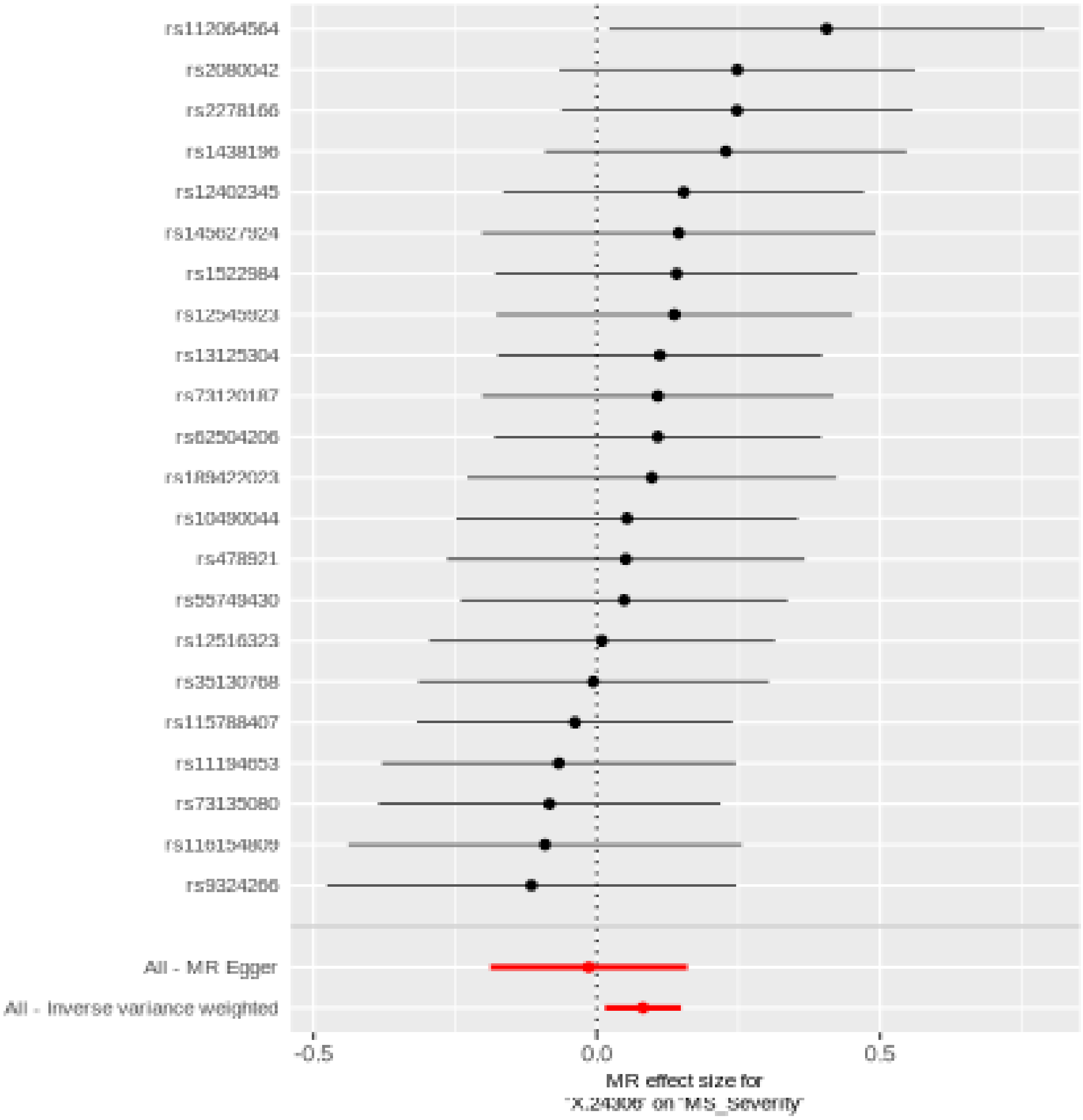

#### Comparison of results using different MR methods

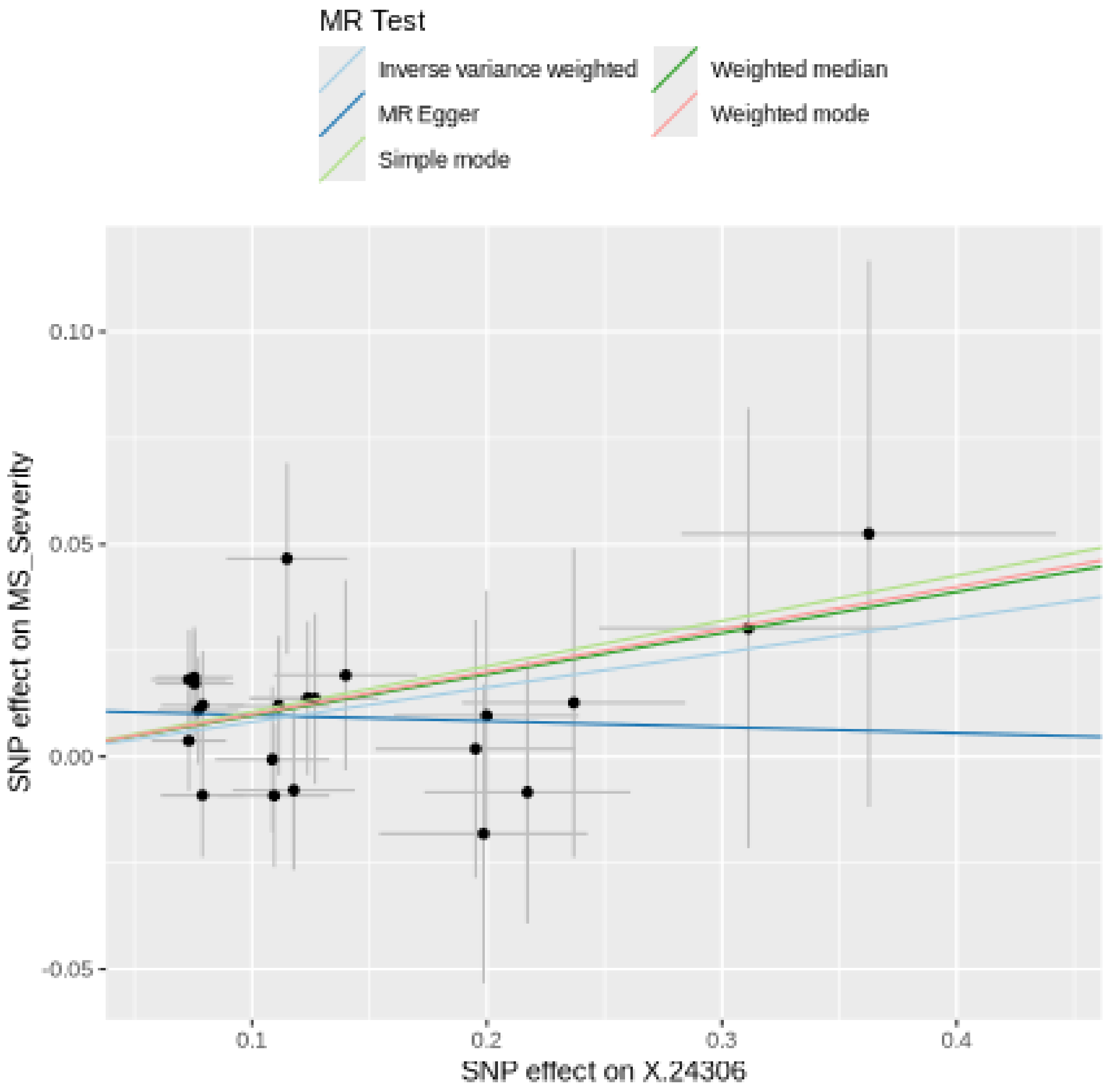

#### Funnel plot

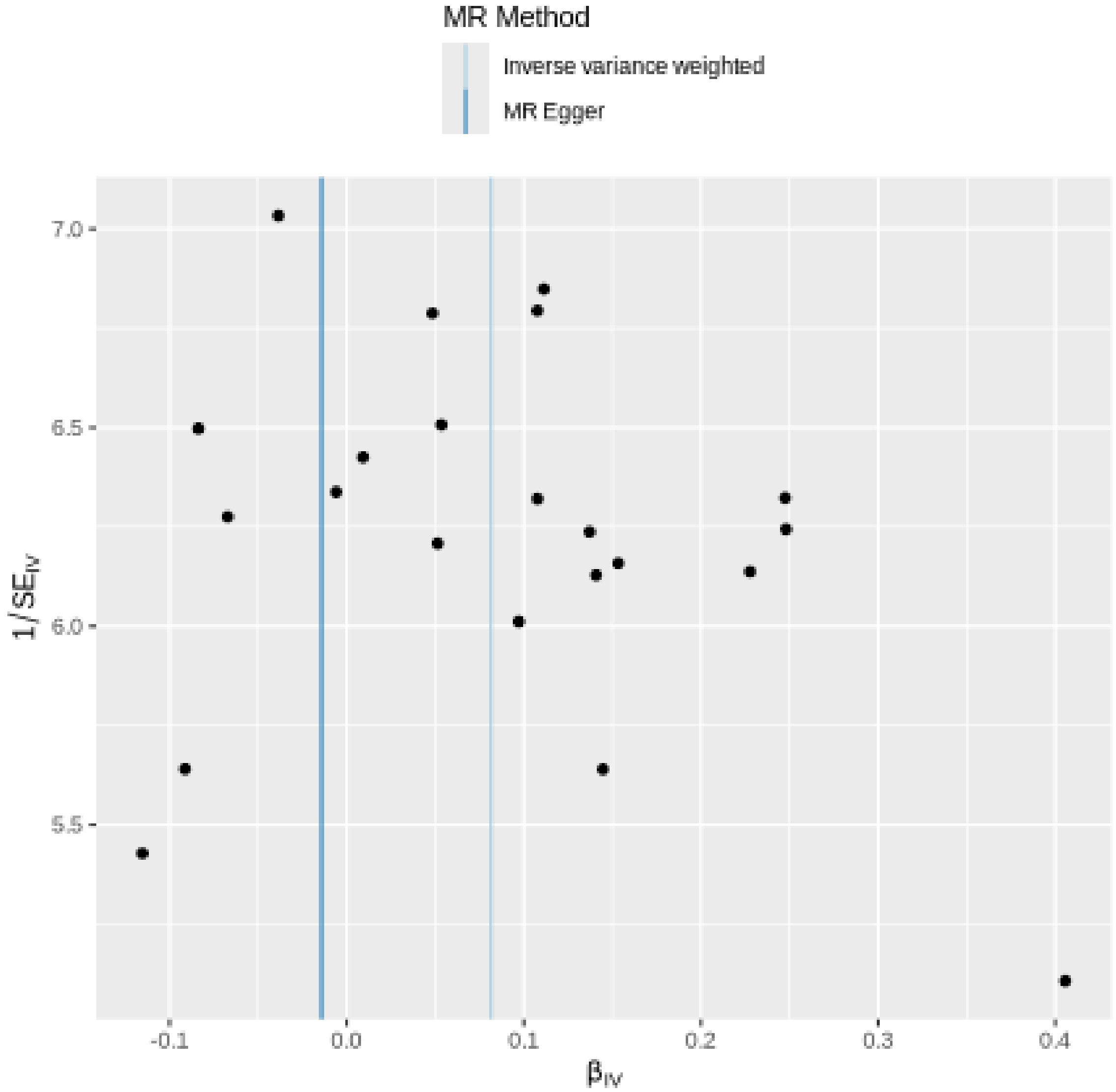

MR Method
Inverse variance weighted
MR Egger
7.0
6.5
6.0
5.5
1/SE_IV
-0.1
0.0
0.1
0.2
0.3
0.4
β_IV

### Leave-one-out sensitivity analysis

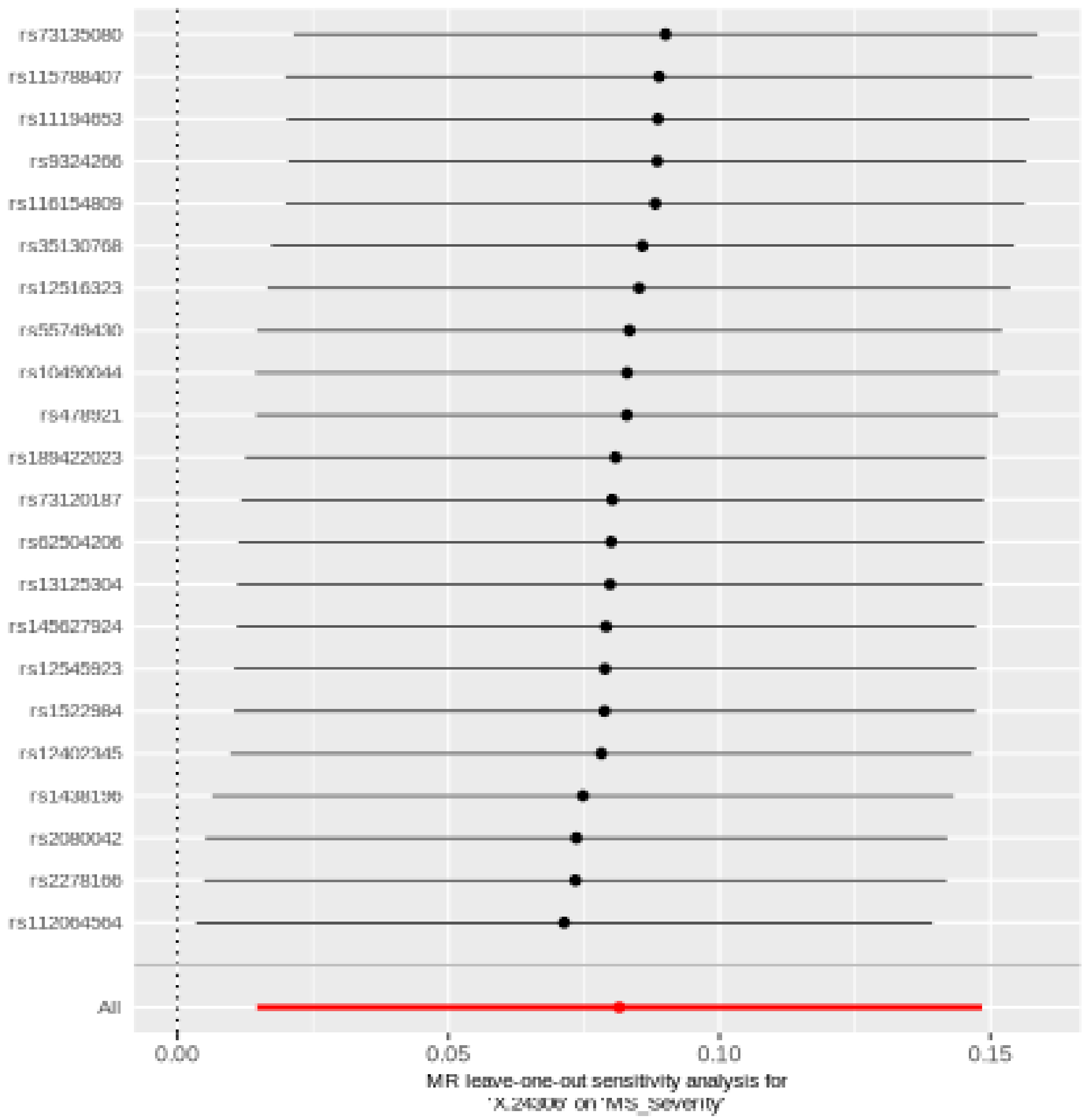

## X.24565

| method | nsnp | b | se | pval |
|---|---|---|---|---|
| MR Egger | 29 | 0.0522923 | 0.0483979 | 0.2894914 |
| Weighted median | 29 | 0.0452917 | 0.0314010 | 0.1492001 |
| Inverse variance weighted | 29 | 0.0517982 | 0.0221990 | 0.0196293 |
| Simple mode | 29 | 0.1010714 | 0.0629810 | 0.1197609 |
| Weighted mode | 29 | 0.1010714 | 0.0616460 | 0.1122888 |

##### Heterogeneity tests

| method | Q | Q_df | Q_pval |
|---|---|---|---|
| MR Egger | 17.63719 | 27 | 0.9142178 |
| Inverse variance weighted | 17.63732 | 28 | 0.9349184 |

##### Test for directional horizontal pleiotropy

| egger_intercept | se | pval |
|---|---|---|
| -9.89e-05 | 0.0086077 | 0.9909177 |

### Forest plot of single SNP MR

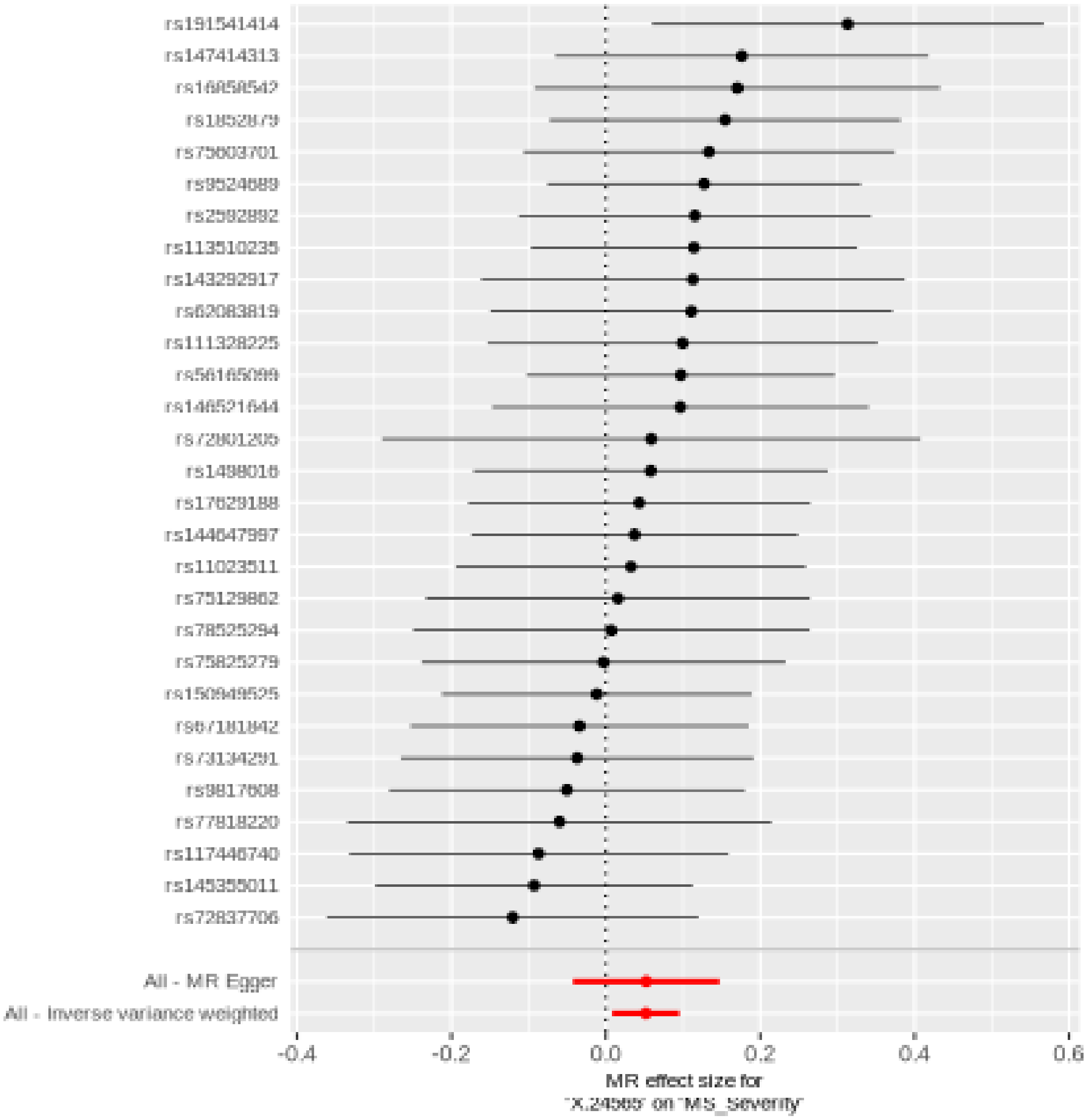

#### Comparison of results using different MR methods

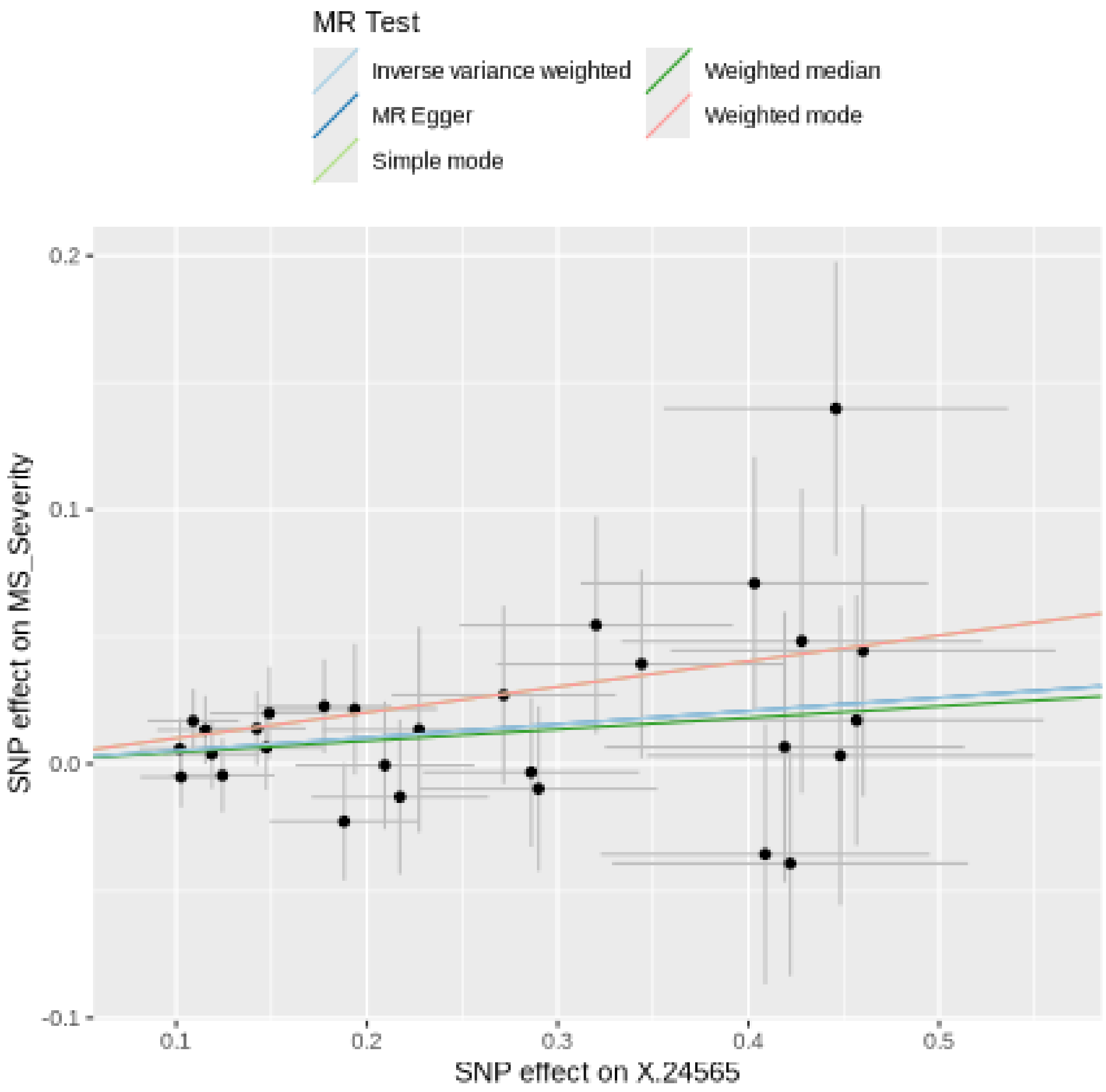
MR Test
Inverse variance weighted
MR Egger
Simple mode
Weighted median
Weighted mode
0.2
0.1
0.0
-0.1
SNP effect on MS_Severity
0.1
0.2
0.3
0.4
0.5
SNP effect on X.24565

#### Funnel plot

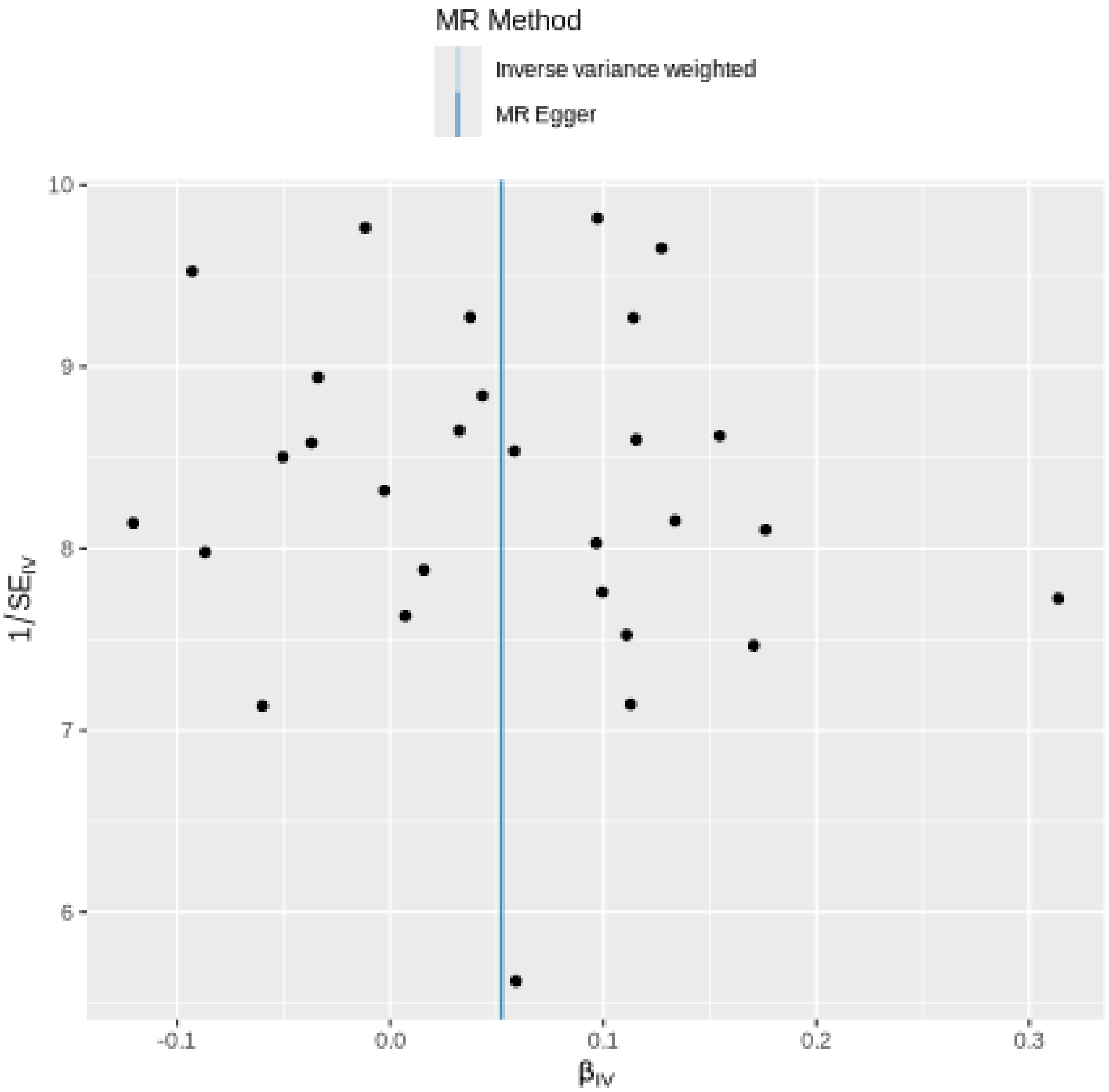
MR Method
Inverse variance weighted
MR Egger
10
9
8
7
6
1/SE_IV
-0.1
0.0
0.1
0.2
0.3
β_IV

#### Leave-one-out sensitivity analysis

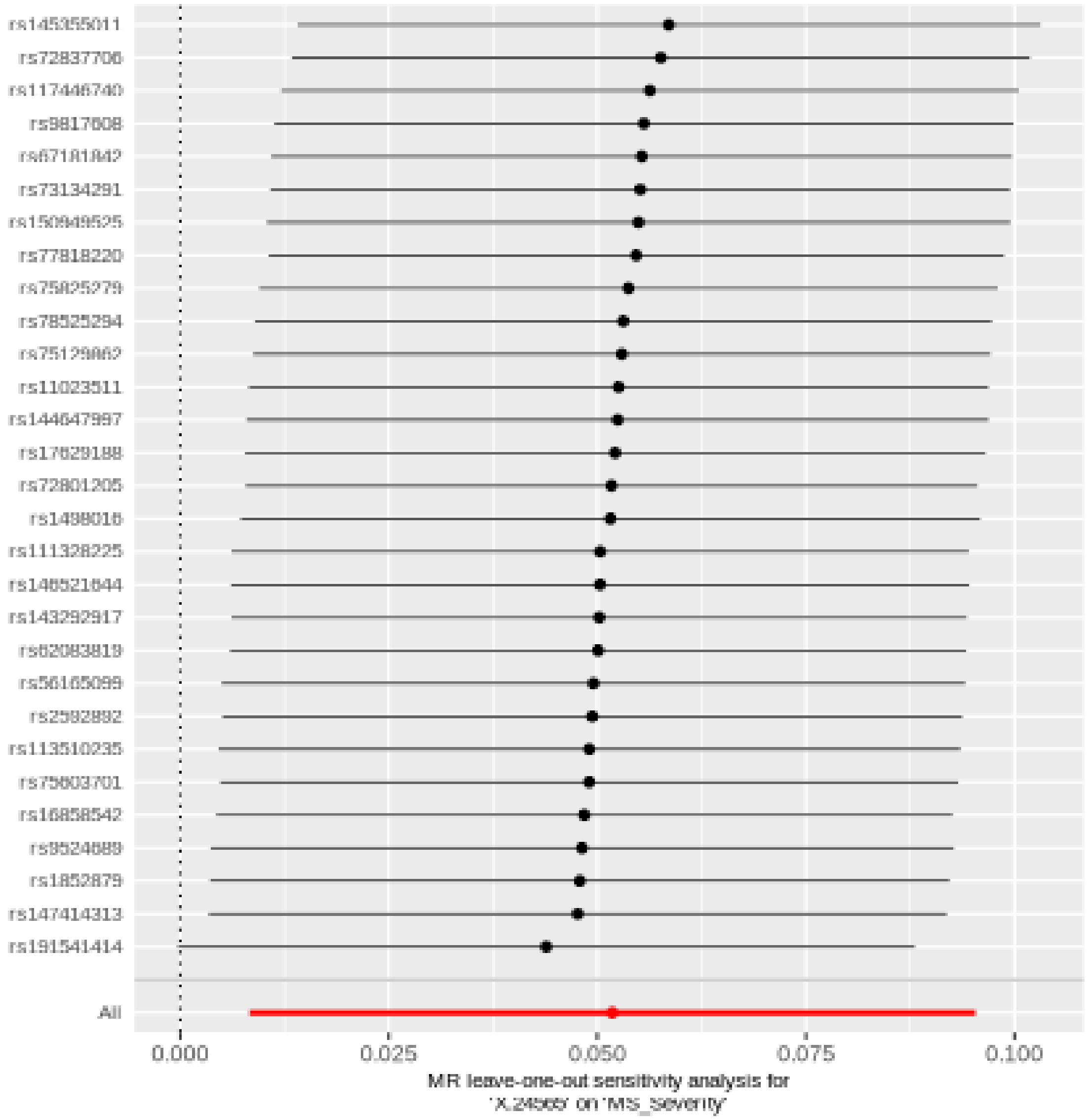

## X.26054

| method | nsnp | b | se | pval |
|---|---|---|---|---|
| MR Egger | 34 | 0.0782465 | 0.0329502 | 0.0237271 |
| Weighted median | 34 | 0.0628843 | 0.0271843 | 0.0207087 |
| Inverse variance weighted | 34 | 0.0450557 | 0.0190534 | 0.0180443 |
| Simple mode | 34 | 0.0183584 | 0.0614200 | 0.7668915 |
| Weighted mode | 34 | 0.0568688 | 0.0276113 | 0.0473952 |

##### Heterogeneity tests

| method | Q | Q_df | Q_pval |
|---|---|---|---|
| MR Egger | 23.84908 | 32 | 0.8498266 |
| Inverse variance weighted | 25.37344 | 33 | 0.8261003 |

##### Test for directional horizontal pleiotropy

| egger_intercept | se | pval |
|---|---|---|
| -0.0061511 | 0.0049821 | 0.2259508 |

### Forest plot of single SNP MR

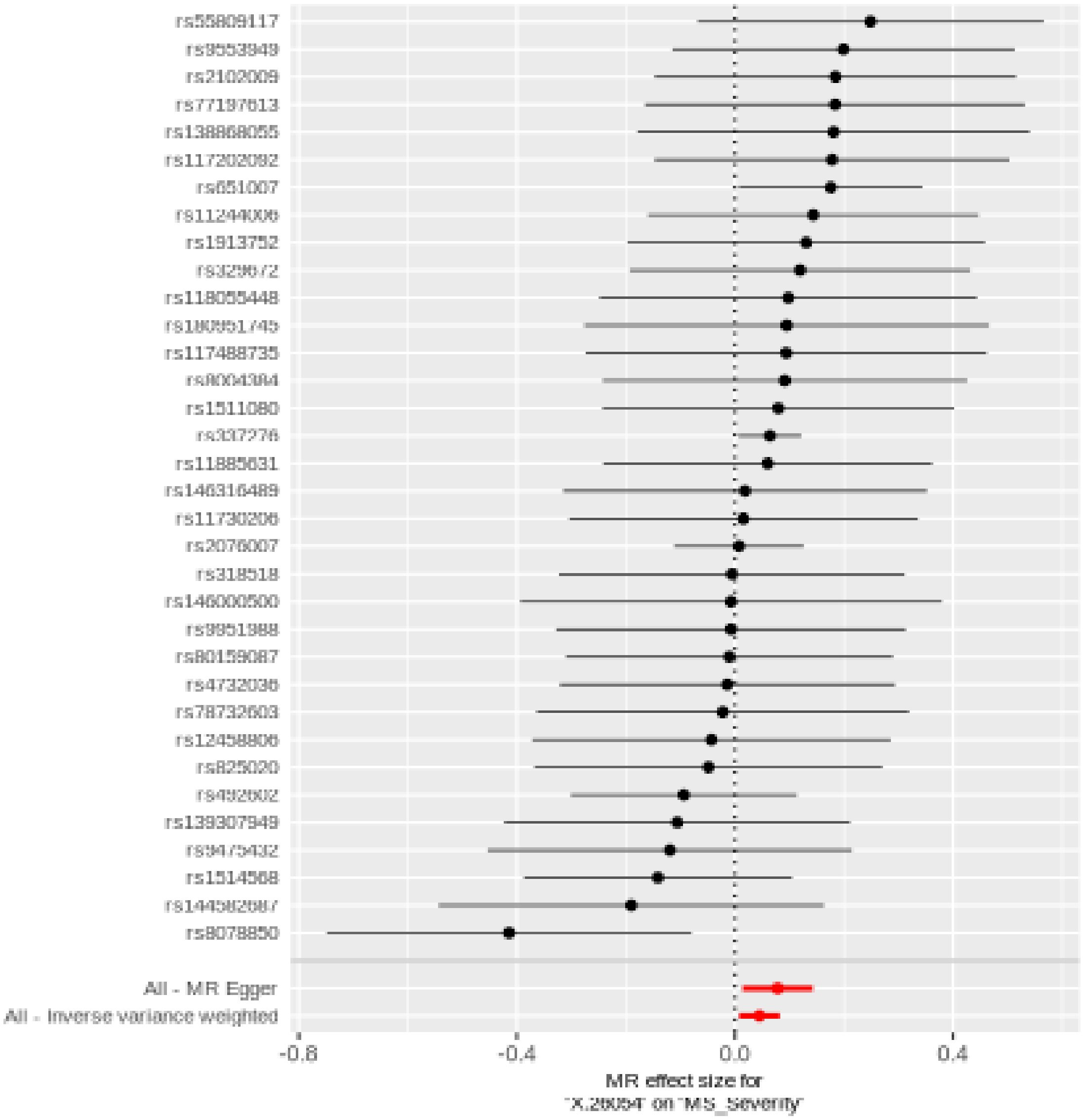

#### Comparison of results using different MR methods

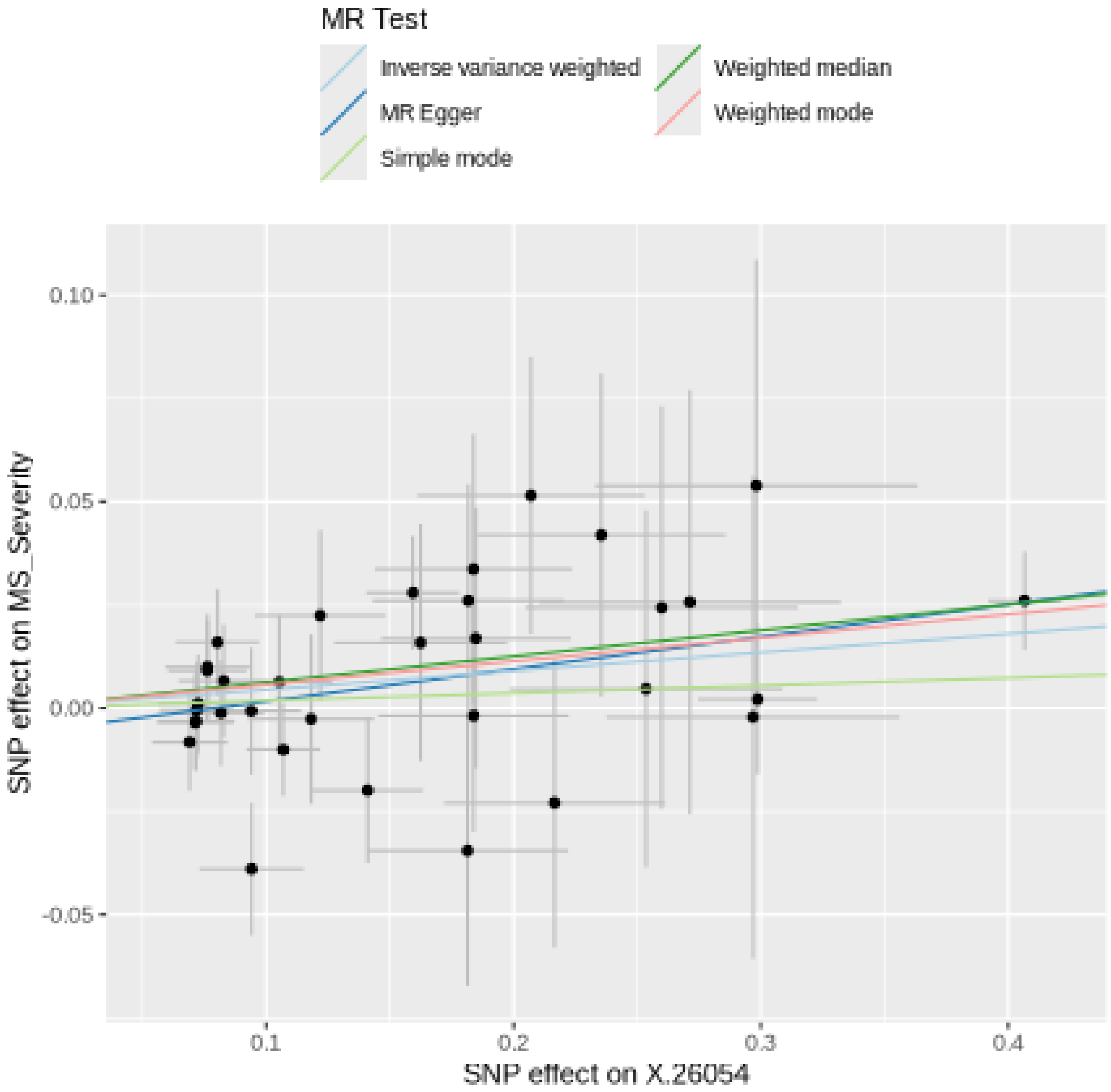

#### Funnel plot

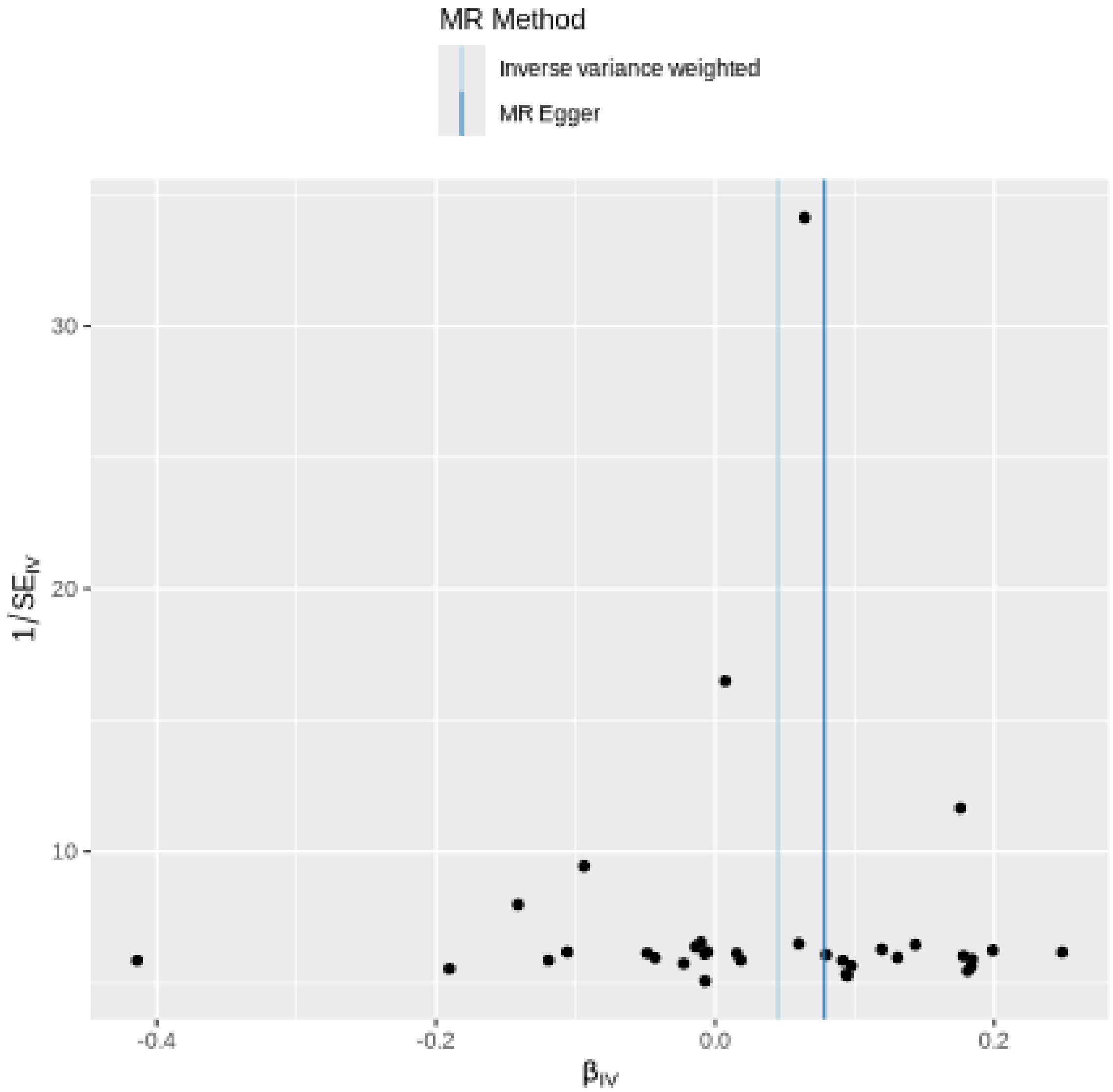

MR Method
Inverse variance weighted
MR Egger
30
20
10
1/SE$_{IV}$
-0.4
-0.2
0.0
0.2
β$_{IV}$

### Leave-one-out sensitivity analysis

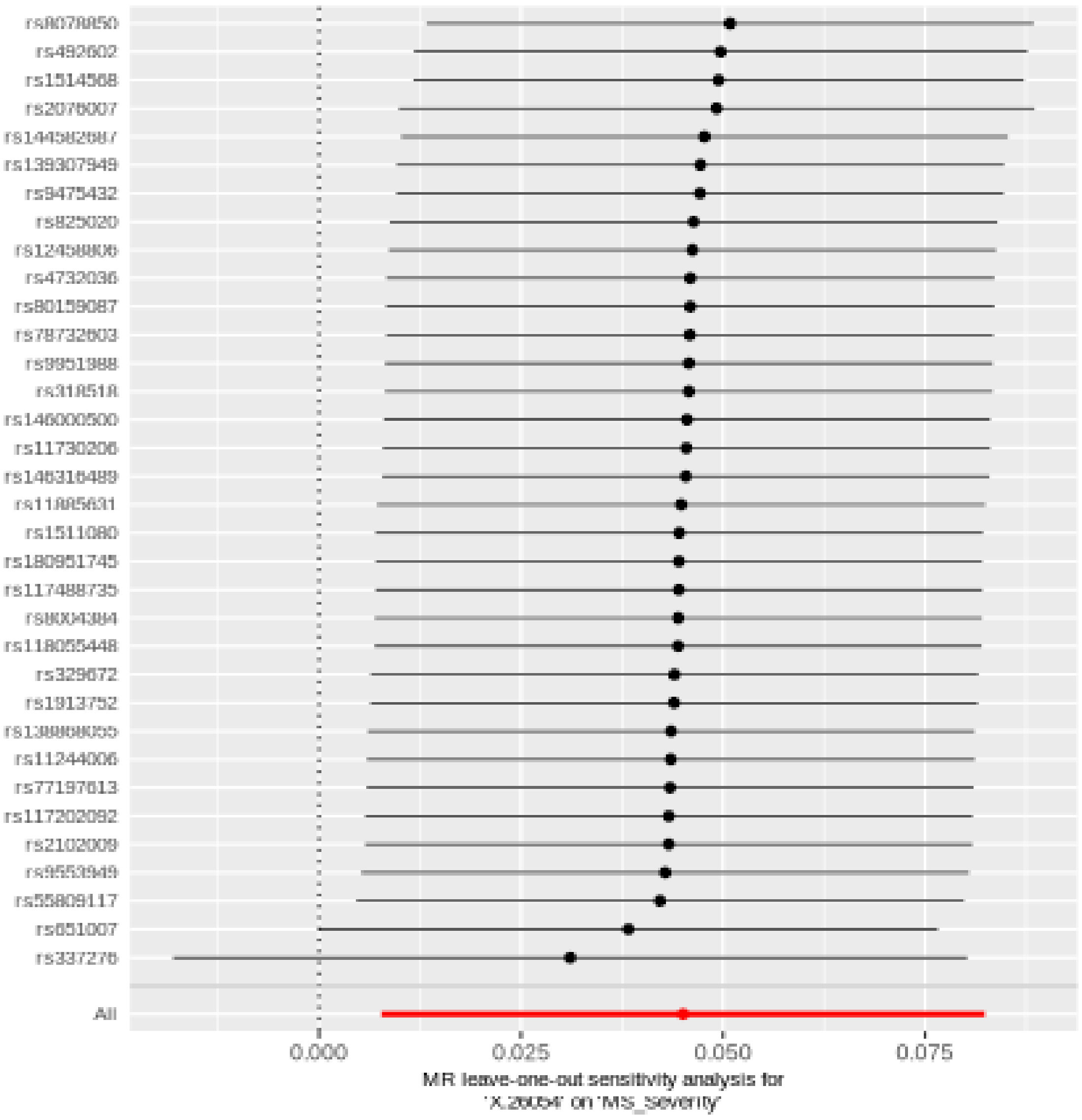
